# European and multi-ancestry genome-wide association meta-analysis of atopic dermatitis highlights importance of systemic immune regulation

**DOI:** 10.1101/2022.10.05.22279072

**Authors:** Ashley Budu-Aggrey, Anna Kilanowski, Maria K Sobczyk, Suyash S Shringarpure, Ruth Mitchell, Kadri Reis, Anu Reigo, Reedik Mägi, Mari Nelis, Nao Tanaka, Ben M Brumpton, Laurent F Thomas, Pol Sole-Navais, Christopher Flatley, Antonio Espuela-Ortiz, Esther Herrera-Luis, Jesus VT Lominchar, Jette Bork-Jensen, Ingo Marenholz, Aleix Arnau-Soler, Ayoung Jeong, Katherine A Fawcett, Hansjorg Baurecht, Elke Rodriguez, Alexassander Couto Alves, Ashish Kumar, Patrick M Sleiman, Xiao Chang, Carolina Medina-Gomez, Chen Hu, Cheng-jian Xu, Cancan Qi, Sarah El-Heis, Philip Titcombe, Elie Antoun, João Fadista, Carol A Wang, Elisabeth Thiering, Shujie Xiao, Sara Kress, Dilini M Kothalawala, Latha Kadalayil, Jiasong Duan, Hongmei Zhang, Thomas Hoffmann, Eric Jorgenson, Hélène Choquet, Neil Risch, Pål Njølstad, Ole A Andreassen, Stefan Johansson, Catarina Almqvist, Tong Gong, Vilhelmina Ullemar, Robert Karlsson, Patrik KE Magnusson, Agnieszka Szwajda, Esteban G Burchard, Jacob P Thyssen, Torben Hansen, Line L Kårhus, Thomas M Dantoft, Alexander C.S.N. Jeanrenaud, Ahla Ghauri, Andreas Arnold, Georg Homuth, Susanne Lau, Markus M Nöthen, Norbert Hübner, Medea Imboden, Alessia Visconti, Mario Falchi, Veronique Bataille, Pirro Hysi, Natalia Ballardini, Dorret I Boomsma, Jouke J Hottenga, Martina Müller-Nurasyid, Tarunveer S Ahluwalia, Jakob Stokholm, Bo Chawes, Ann-Marie M Schoos, Ana Esplugues, Mariona Bustamante, Benjamin Raby, Hasan Arshad, Chris German, 23andMe Research Team, Tõnu Esko, Lili A Milani, Andres Metspalu, Chikashi Terao, Katrina Abuabara, Mari Løset, Kristian Hveem, Bo Jacobsson, Maria Pino-Yanes, David P Strachan, Niels Grarup, Allan Linneberg, Young-Ae Lee, Nicole Probst-Hensch, Stephan Weidinger, Marjo-Riitta Jarvelin, Erik Melén, Hakon Hakonarson, Alan D Irvine, Debbie L Jarvis, Tamar Nijsten, Liesbeth Duijts, Judith M Vonk, Gerard H Koppelmann, Keith M Godfrey, Sheila J Barton, Bjarke Feenstra, Craig E Pennell, Peter D Sly, Patrick G Holt, Keoki L Williams, Hans Bisgaard, Klaus Bønnelykke, John Curtin, Angela Simpson, Clare Murray, Tamara Schikowski, Supinda Bunyavanich, Scott T Weiss, John W Holloway, Josine Min, Sara J Brown, Marie Standl, Lavinia Paternoster

## Abstract

Atopic dermatitis (AD) is a common inflammatory skin condition and prior genome-wide association studies have identified 71 associated loci. In the current study we conducted the largest AD GWAS to date (discovery N=1,086,394, replication N=3,604,027), combining previously reported cohorts with additional available data. We identified 81 loci (29 novel) in the European-only analysis and 15 additional loci in the multi-ancestry analysis (6 novel). All 81 variants replicated in a separate European analysis. Eleven variants from the multi-ancestry analysis replicated in at least one of the populations tested (European, Latino or African). While four variants appeared to be specific to individuals of Japanese ancestry. AD loci showed enrichment for DNAse I hypersensitivity and eQTL signals in blood. At each locus we prioritised candidate genes by integrating multi-omic data. The implicated genes are predominantly in immune pathways of relevance to atopic inflammation and some offer drug repurposing opportunities.

## Introduction

Atopic dermatitis (AD, or eczema) is a common allergic disease, characterised by (often relapsing) skin inflammation affecting up to 20% of children and 10% of adults^1^. Several genome-wide association studies (GWASs) have been performed in recent years, identifying genetic risk loci for AD.

Our most recent GWAS meta-analysis within the EAGLE (EArly Genetics and Lifecourse Epidemiology) consortium, published in 2015 uncovered 31 AD risk loci^2^. Since then, additional GWAS have been published which have confirmed known risk loci^3,4^ and discovered additional signals^5^. Five novel loci were identified in a European meta-analysis^6^, and variants in 3 genes were implicated in a rare variant study in addition to 5 novel loci^7^. Four novel loci were reported in a Japanese population (and another 4 identified in a trans-ethnic meta-analysis in the same study)^8^, giving a total of 71 previously reported AD loci^2–14^ (defined as 1Mb regions) of which 57 have been reported in European ancestry individuals, 18 have been reported in individuals of non-European ancestry and 29 in individuals across multiple ancestry groups (Supplementary Table 1).

The availability of several new large population-based studies has provided an opportunity to perform an updated GWAS of AD, aiming to incorporate data from all cohorts that have contributed to previously published AD GWAS, as well as data from additional cohorts, to present the most comprehensive GWAS of AD to date, including comparison of effects between European, East Asian, Latino and African ancestral groups. We identify novel loci and use multi-omic data to further characterise these associations, prioritising candidate causal genes at individual loci and investigating the genetic architecture of AD in relation to tissues of importance and shared genetic risk with other traits.

## Results

### European GWAS

The discovery European meta-analysis (N=864,982 from 40 cohorts, summarized in Supplementary

Table 2) identified 81 genome-wide significant independent signals (Figure 1a and Supplementary Figure 1). 52 were at previously reported loci (Table 1) and 29 (Table 2) were novel (according to criteria detailed in the methods). All 81 were associated in the European 23andMe replication analysis (*P*<6×10^−4^, N=2,904,664, Table 1). There was little evidence of genomic inflation in the individual studies (lambda < 1.05) and overall (1.06). Conditional analysis determined 44 additional secondary independent signals (*P*<1×10^−5^) across 21 loci (Supplementary Table 3).

**Figure 1.**
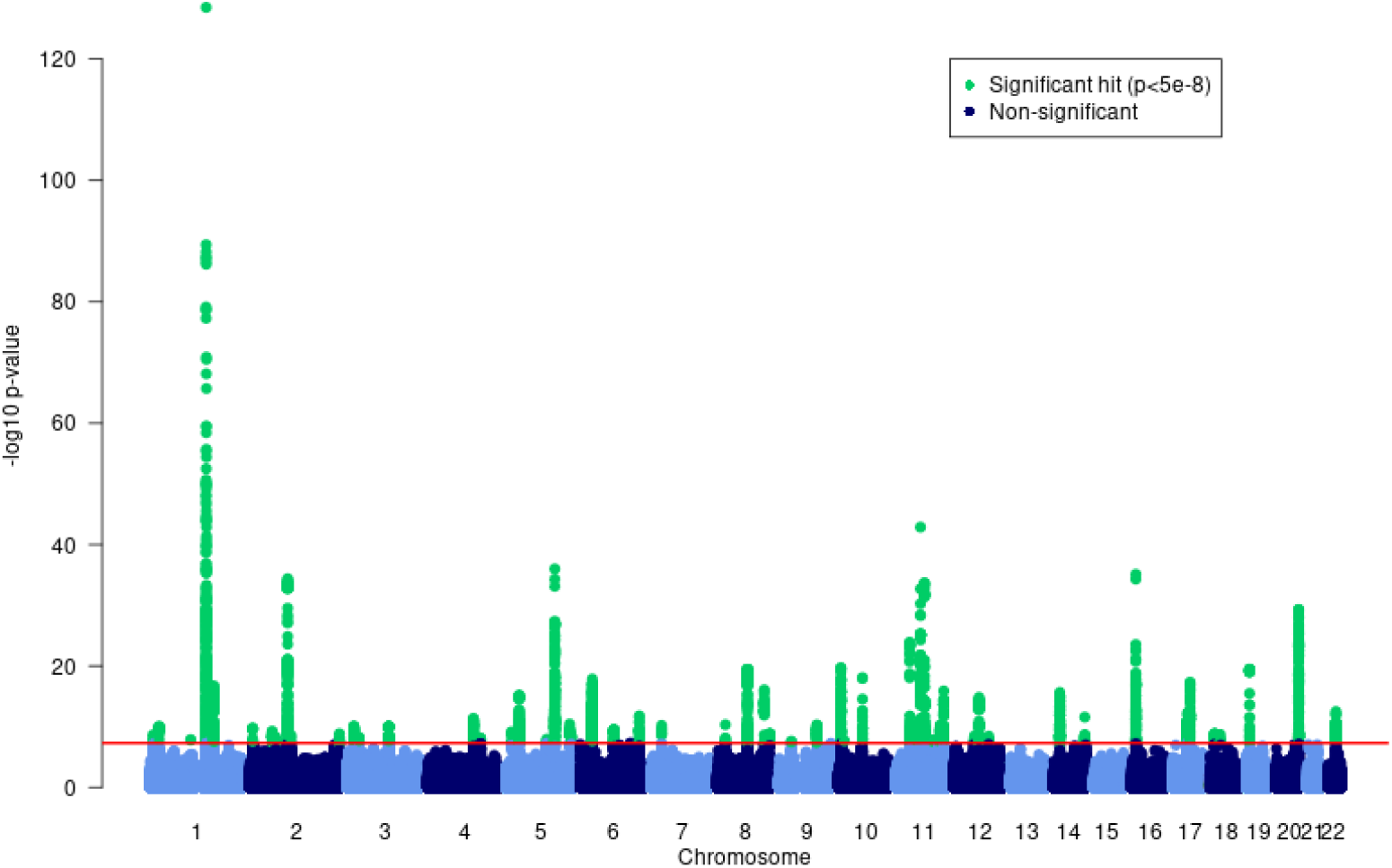
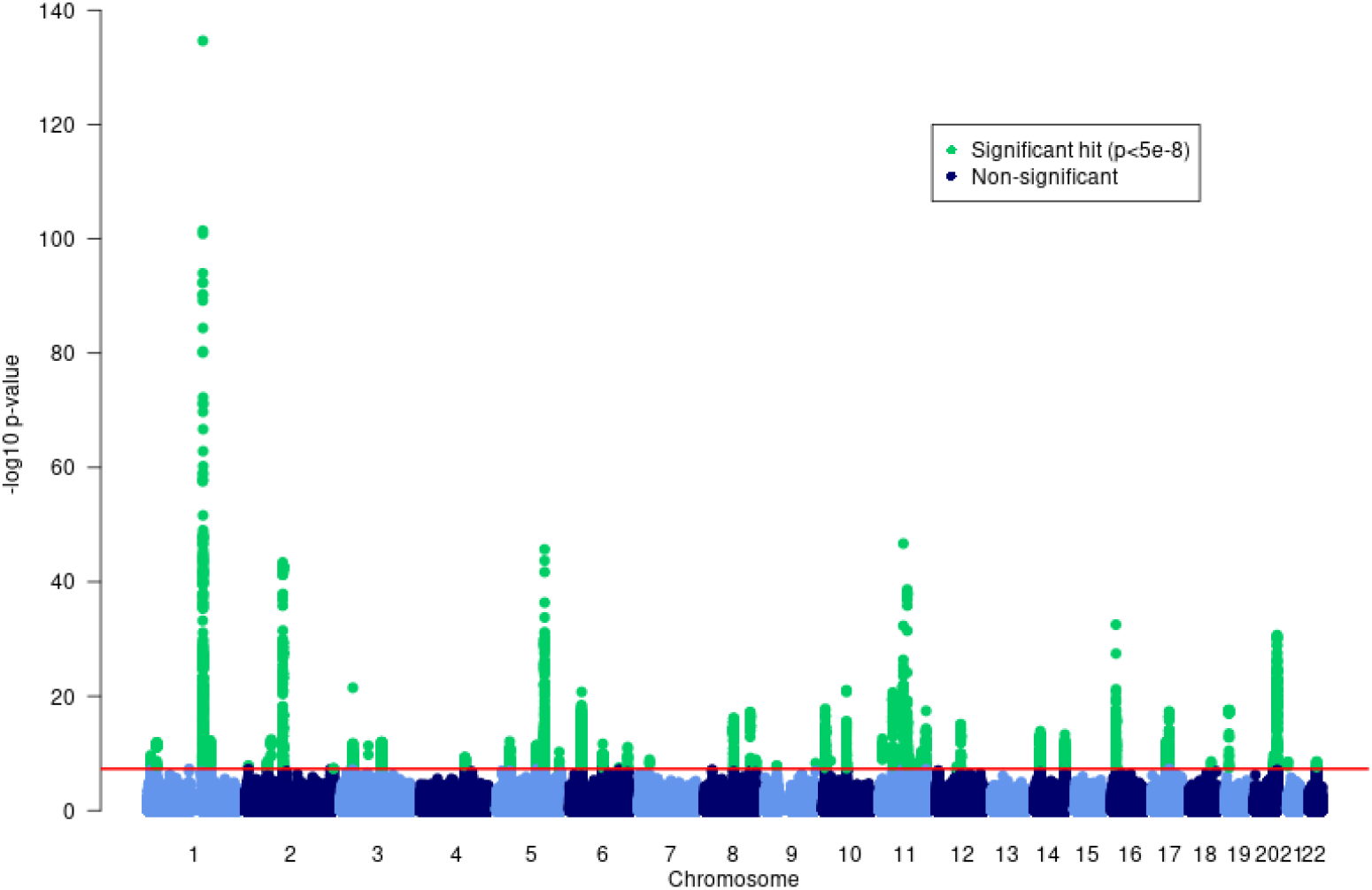
Manhattan plots of atopic dermatitis GWAS for (A) the European-only fixed effects meta-analysis (N= 864,982) and (B) the multi-ancestry MR-MEGA meta-analysis (N=1,086,394). -log_10_(*P*-values) are displayed for all variants in the meta-analysis. Variants that meet the genome-wide significance threshold (5×10^−8^, red line) are shown in green

The SNP-based heritability (h^2^_SNP_) for AD was estimated to be 5.6% in the European discovery meta-analysis. This is low in comparison to heritability estimates for twin studies (∼80%)^15,16^, but comparable with previous h^2^_SNP_estimates for AD in Europeans (5.4%)^6^.

### Multi-ancestry GWAS

In a multi-ancestry analysis including individuals of European, Japanese, Latino and African ancestry (Supplementary Table 2, N=1,086,394), a total of 89 signals were identified as associated with AD (Figure 1b and Supplementary Figure 1). 70 of these had been detected in the European-only analysis and a further 12 showed some evidence for association (*P*<5×10^−4^) in the European analysis, but 7 SNPs were not associated (*P*>0.1) in Europeans (Table 3, Supplementary Table 4).

Of the 19 loci that reached genome-wide significance in the multi-ancestry discovery analysis only (Table 3), 11 replicated in at least one of the replication samples (of European, Latino and/or African ancestry; *P*<2.63×10^−3^). Four SNPs which did not replicate in any of the samples (rs9864845, rs34665982, rs45602133, rs4312054) appeared to have been driven by association in the Japanese RIKEN study only (Supplementary Table 4, Supplementary Figure 2). A further 4 SNPs did not replicate, and on closer examination (Supplementary Figure 2, and MAF in cases <1%), their association in the discovery analysis appeared to be driven by a false positive outlying result in a single European cohort.

Nine of the loci in Table 3 have been previously reported as associated with AD. Two (rs117137535^7^ and rs1059513^8^) were previously only associated in Europeans (and these were variants that were just below the genome-wide significance threshold in our European only analysis). The locus at rs4262739 had been previously reported in a Japanese and European meta-analysis^8^, while the other 6 were previously associated in Japanese studies^8,10^, using the same Japanese data (RIKEN) that we include here. Therefore in our multi-ancestry analysis (and replication) we identify 6 loci that have not previously been reported in a GWAS of AD of any ancestry (rs77869365, rs9247, rs9263868, rs34599047, rs7773987, rs17120177), all of which are associated in two or more populations in our data (Table 3).

In addition, for 5 loci which had previously been associated in individuals of European and/or Japanese ancestry, we now show evidence that these are also associated in individuals of Latino ancestry (Table 3) and one is also associated in individuals of African ancestry (Table 3).

### Comparison of associations between ancestries

Findings from both the European and multi-ancestry results are remarkably similar across populations of European and Latino ancestry (Supplementary Figure 2). Of the 67 variants with results available from Japanese, European and African ancestry individuals, 32 showed consistent effects across all three ancestries; 22 showed consistent effects between Japanese and European individuals, but no association in individuals of African ancestry; and 6 variants showed consistent effects between individuals of European and African ancestry with no association in Japanese (Supplementary Figure 2). In addition to those reported above as specific to the Japanese population, a further 3 appeared to be specific to individuals of European ancestry (Supplementary Figure 2). Our assessment of allele frequencies showed no consistent differences between populations to account for the differences in association, except that a Japanese-specific variant, rs45602133, had a MAF of 17% in the Japanese data, but only 8% in Africans and 1% in Europeans (Supplementary Table 5).

### Established associations

Review of previous work in this field (Supplementary Table 1) shows that a total of 198 unique variants (across a much smaller number of loci) have been reported to be associated with AD. We found evidence for all but 7 variants of these being nominally associated in the current GWAS (81% in the European and 96% in the multi-ancestry analysis).

### Genetic correlation between AD and other traits

LD score regression analyses showed high genetic correlation, as expected, between AD and related allergic traits, e.g. asthma (rg=0.53, p=2×10^−32^), hay fever (rg=0.51, p=7×10^−17^) and eosinophil count (rg=0.27, *P*=1×10^−7^) (Supplementary Figure 3 & Supplementary Table 6). In addition, depression and anxiety showed notable genetic correlation with AD (rg=0.17, *P*=2×10^−7^), a relationship which has been reported previously, but causality has not been established^17^. Furthermore, gastritis also showed substantial genetic correlation (rg=0.31, *P*=1×10^−5^), which may be due to the AD genetic signal including variants with pervasive inflammatory function or the observed correlation could indicate a shared risk locus for inflammation or microbiome alteration in the upper gastrointestinal tract, or it may reflect the use of systemic corticosteroid treatment for atopic disease which in some cases causes gastritis as a side effect.

### Tissue, cell and gene-set enrichment

The tissue enrichment analyses each found blood to be the tissue showing strongest enrichment of GWAS signals. The Garfield test for enrichment of genome-wide signals (with *P*<10^−8^) in DNase I hypersensitive sites (DHS peaks) found evidence of enrichment (*P*<0.00012) in 41 blood tissue analyses, a greater signal than another tissue or cell type (Figure 2a and Supplementary Table 7). The strongest enrichment (OR>5.5 and *P*<1×10^−10^) was seen for T-cell, B-cell and natural killer lymphocytes (CD3+, CD4+, CD56+ and CD19+). As expected for AD, Th2 showed stronger enrichment (OR=4.3, *P*=1×10^−8^) than Th1 (OR=2.3, *P*=2×10^−4^). The strongest enrichment in tissue samples representing skin was seen for foreskin keratinocytes (OR=2.0, *P*=0.008), but this did not meet a Bonferroni-corrected *P*-value threshold (0.05/425=1.2×10^−4^).

**Figure 2.**
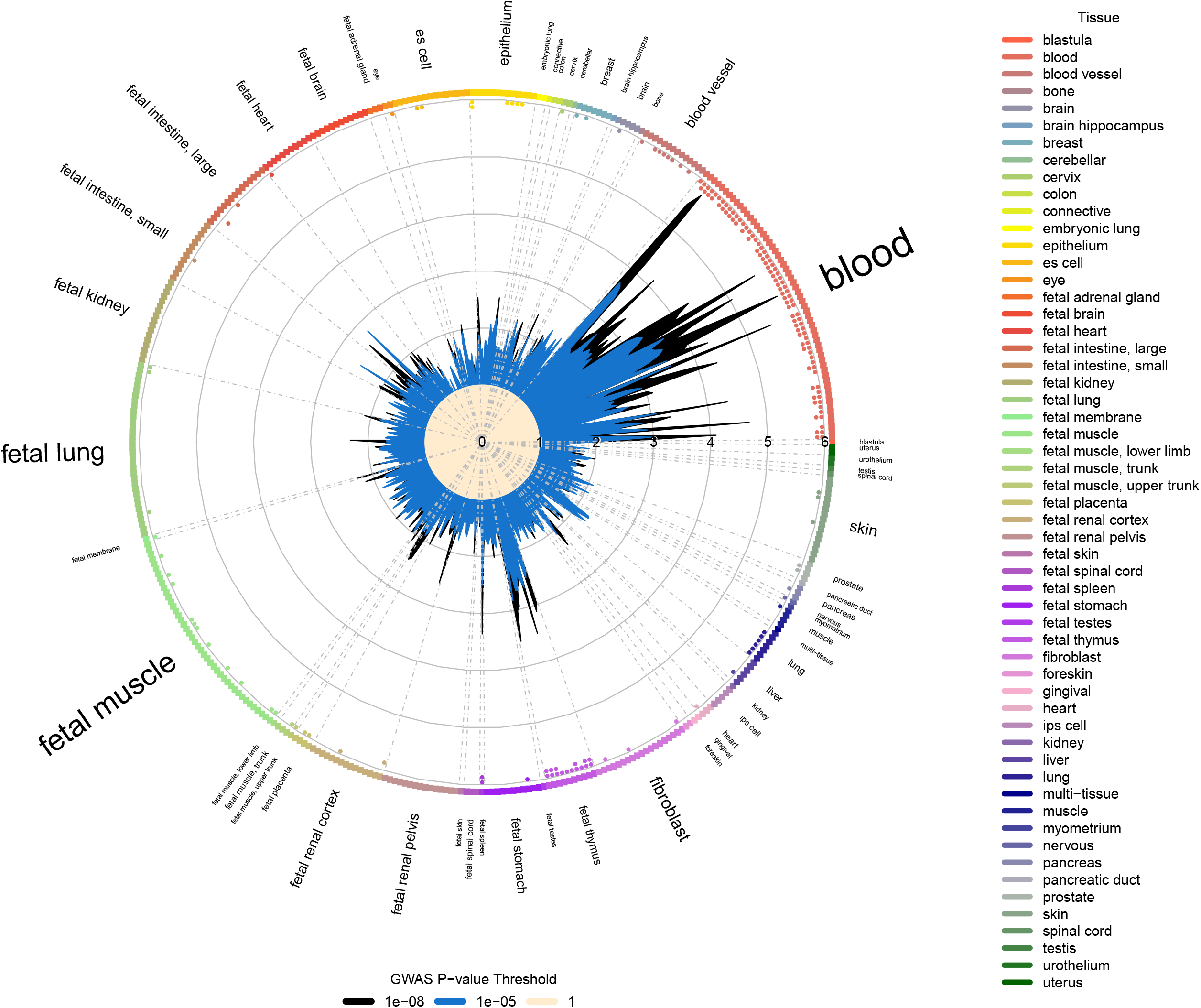
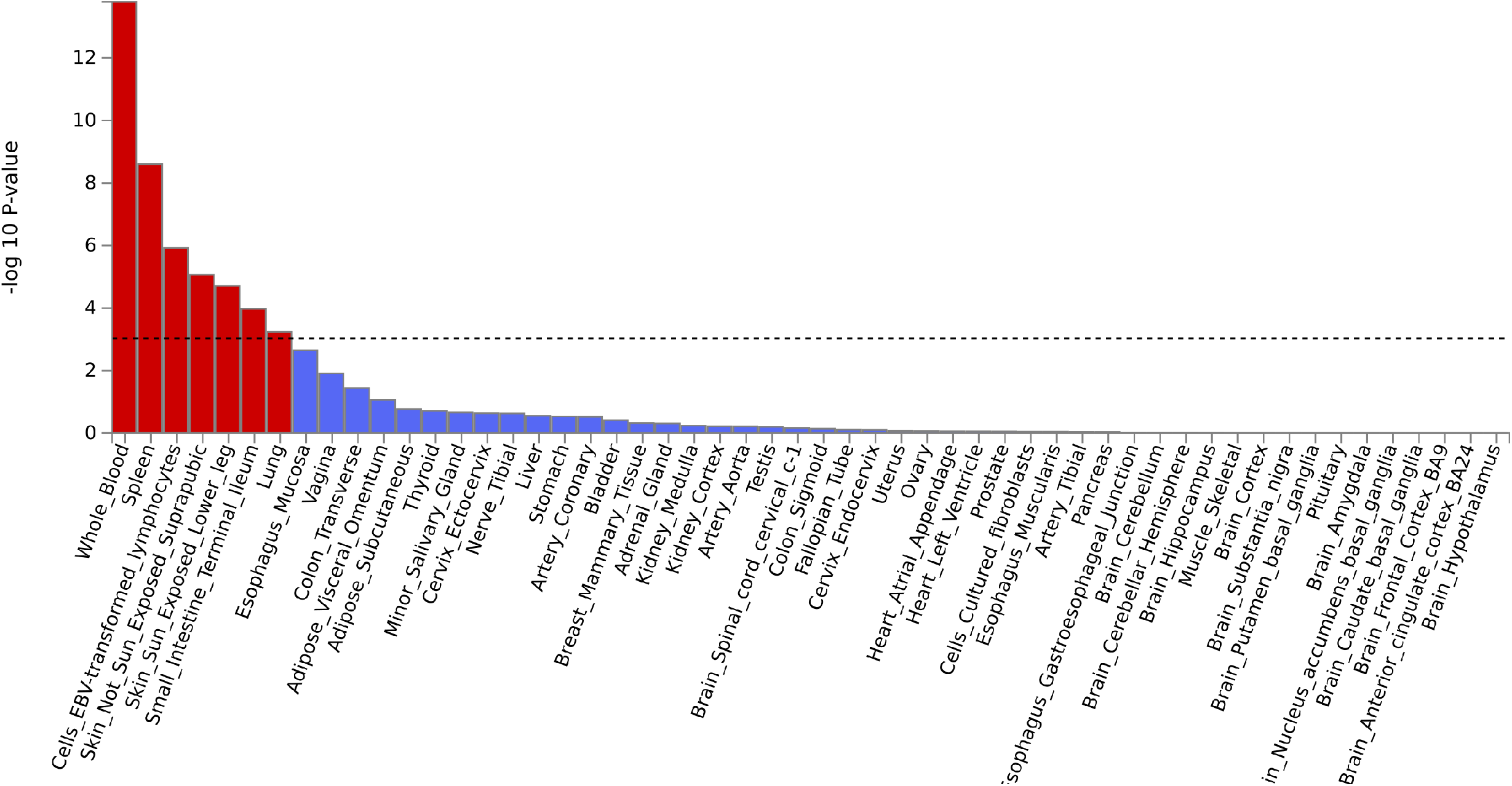
Cell type tissue enrichment analysis. **A. GARFIELD enrichment analysis**. Plot shows enrichment for AD associated variants in DNase I Hypersensitive sites (broad peaks) from ENCODE and Roadmap Epigenomics datasets across cell types. Cell types are sorted and labelled by tissue type. ORs for enrichment are shown for variants at GWAS thresholds of *P*<1×10^−8^ (black) and *P*<1×10^−5^ (blue) after multiple-testing correction for the number of effective annotations. Outer dots represent enrichment thresholds of *P*<1×10^−5^ (one dot) and *P*<1×10^−6^ (two dots). Font size of tissue labels corresponds to the number of cell types from that tissue tested. **B. MAGMA enrichment analysis**. Plot shows p-value for MAGMA enrichment for AD associated variants with gene expression from 54 GTEx ver.8 tissue types. The enrichment –log_10_(*P*-value) for each tissue type is plotted on the y axis. The Bonferroni corrected threshold *P*=0.0009 is shown as a dotted line and the 7 tissue types that meet this threshold are highlighted as red bars.

The most enriched tissue type in MAGMA gene expression enrichment analysis was whole blood (*P*=2×10^−14^). Others that met our Bonferroni-corrected *P*-value (*P*<0.009) were spleen, EBV-transformed lymphocytes, sun-exposed and unexposed skin, small intestine and lung (Figure 2b and Supplementary Table 8).

DEPICT cell-type enrichment analysis identified a similar set of enriched cell-types: blood, leukocytes, lymphocytes and natural killer cells, but with the addition that the strongest enrichment was seen for synovial fluid (*P*=2×10^−7^), which may be due to its immune cell component.

The DEPICT pathway analysis found 420 GO terms with enrichment (FDR<5%) amongst the genes from our GWAS loci (Supplementary Table 9). The pathway with the strongest evidence of enrichment was ‘*hemopoietic or lymphoid organ development*’ (*P*=1×10^−16^). All terms with FDR<5% are represented in Supplementary Figure 4, where the terms are grouped according to similarity and the parent terms labelled illustrating the strong theme of immune system development and signalling.

### Gene prioritisation and biological interpretation *in silico*

The top genes prioritised using our composite score from publicly available data for each of the established European AD loci are shown in Table 1 and Figure 3a (and the evidence that makes up the prioritisation scores is shown in Supplementary Figure 5). The top three prioritised genes at each locus are shown in Supplementary Table 10 and a summary of all evidence for all genes reviewed *in silico* is presented in Supplementary Table 11.

**Figure 3.**
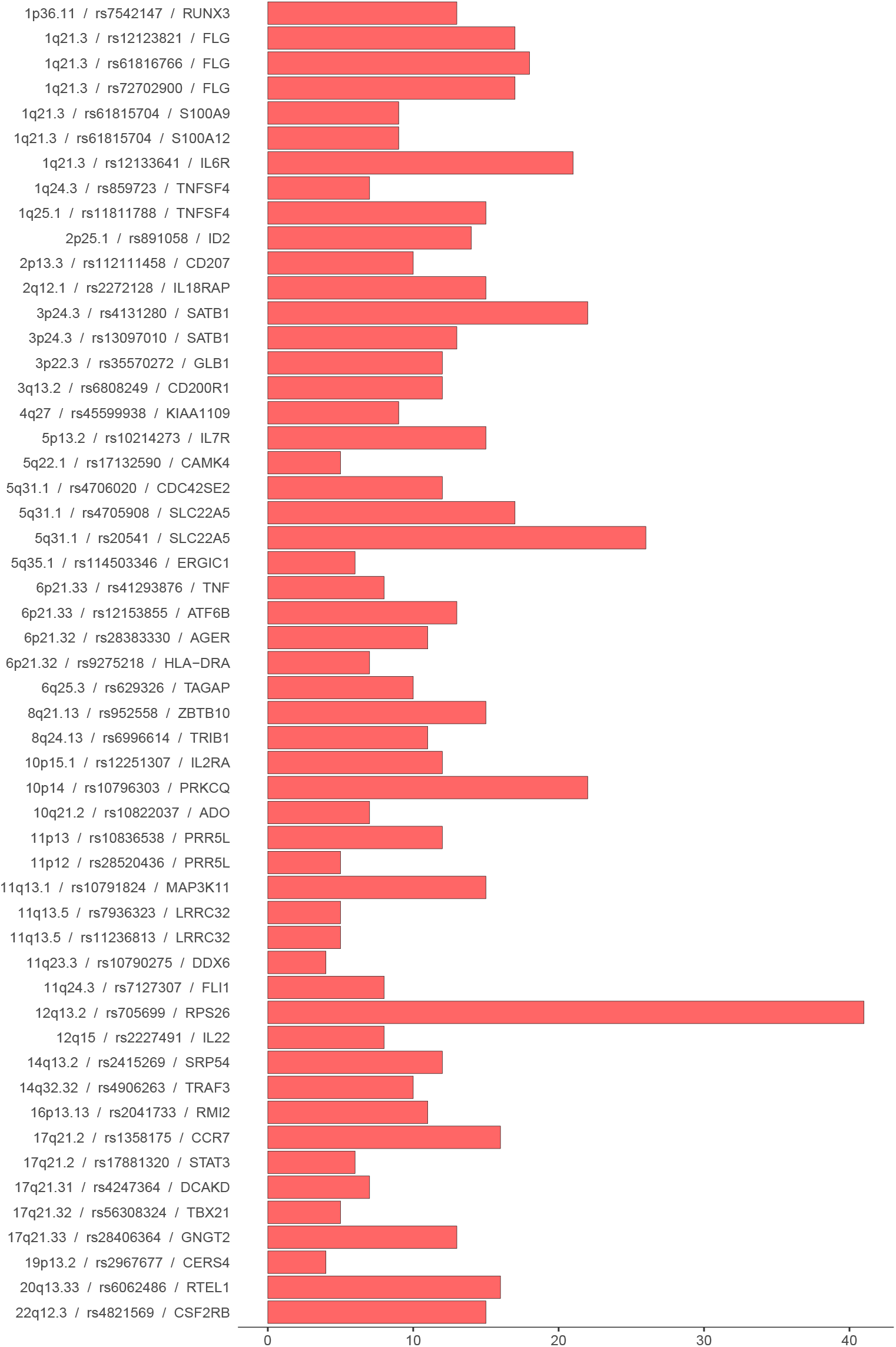
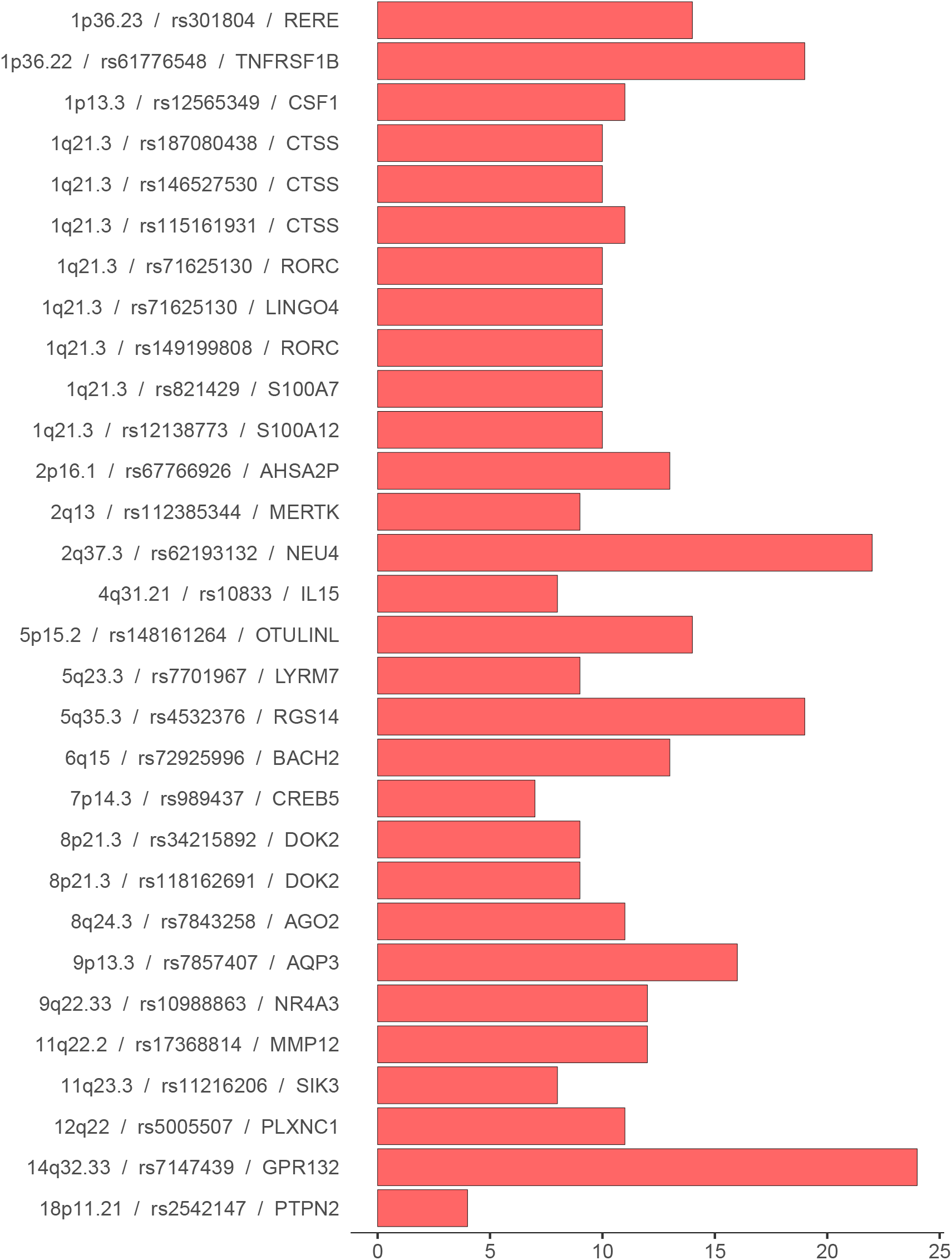
Prioritised genes amongst known (A) and novel (B) loci. For each GWAS locus the top prioritised gene from our bioinformatic analysis is presented along with a bar representing the total evidence score for that gene. A more detailed breakdown of the constituent parts of this evidence score is presented in supplementary figure 5 and the total evidence scores for the top 3 genes at each locus are presented in supplementary table 10.

In most cases the top prioritised gene had been implicated previously or is only superseded marginally by an alternative candidate. One interesting exception is on chromosome 11, where *MAP3K11* (with a role in cytokine signalling – regulating the JNK signalling pathway) is markedly prioritised over the previously implicated *OVOL1*^18^ (involved in hair formation and spermatogenesis), although the prioritisation of *MAP3K11* is predominantly driven by TWAS evidence in multiple cell types rather than colocalisation or other evidence.

There are three instances where multiple signals in the region implicate additional novel genes. Two are genes involved in TLR4 signalling: *S100A9* (prioritised in addition to the established *FLG* and *IL6R* on chromosome 1) and *AGER* (prioritised in addition to *HLA-DRA* on chromosome 6). The third has a likely role in T-cell activation: *CDC42SE2* (prioritised in addition to *SLC22A5* on chromosome 5).

The top prioritised gene at each of the novel European loci are shown in Table 2 and Figure 3b. Many are in pathways already identified by previous findings (e.g. cytokine signalling – especially IL-23, antigen presentation and NF-kappaB proinflammatory response). At one locus, the lead SNP, rs34215892 is a missense mutation within the *DOK2* gene, although this mutation is categorised as tolerated or benign by SIFT and polyphen. The genes with the highest prioritisation score amongst the novel loci were *GPR132* (total evidence score=24), *NEU4* (score=22), *TNFRSF1B* (score=19) and *RGS14* (score=19) and each show biological plausibility as candidates for AD pathogenesis.

GPR132 is a proton-sensing transmembrane receptor, involved in modulating several downstream biological processes, including immune regulation and inflammatory response, as reported previously in an investigation of this protein’s role in inflammatory bowel disease^19^. The index SNP at this locus, rs7147439 (which was associated in Europeans, Latinos, Africans, but not Japanese), is an intronic variant within the *GPR132* gene. The AD GWAS signal at this locus colocalises with the eQTL signal for *GPR132* in several immune cell types (macrophages^20^, neutrophils^21^, several T-cell datasets^22^) as well as in colon, lung and small intestine in GTEx^23^. *GPR132* has also been shown to be upregulated in lesional and nonlesional skin in AD patients, compared to skin from controls^24,25^. OpenTargets and POSTGAP both prioritise *GPR132* for this signal.

The SNP rs62193132 (which showed consistent effects in European, Latino and Japanese, but little evidence for association in African individuals), is in an intergenic region between *NEU4* (∼26kb) and *PDCD1* (∼4kb away) on chromosome 2. *NEU4* was the highest scoring in our gene prioritisation pipeline (score=22). However, *PDCD1* also scores highly (score=18, Supplementary Table 10). NEU4 is an enzyme that removes sialic acid residues from glycoproteins and glycolipids, whereas *PDCD1* is involved in the regulation of T cell function. The AD GWAS signal at this locus colocalises with the eQTL for *NEU4* in several monocyte and macrophage datasets^22,26–28^ as well as in the ileum, colon and skin^23,29^. The eQTL for *PDCD1* also colocalises in monocytes and macrophages^27,28^ as well as T-cells^22^, skin and whole blood^23^. In addition to the eQTL evidence, *PCDC1* is upregulated in lesional and non-lesional skin versus controls^24,25^. OpenTargets and PoPs prioritise *NEU4*, whilst POSTGAP prioritises *PDCD1* at this locus.

TNFRSF1B is part of the TNF receptor, with an established role in cytokine signalling. rs61776548 (which showed consistent associations across all major ancestries tested) is 136kb upstream of *TNFRSF1B*, actually within an intron of *MIIP*. MIIP encodes Migration and Invasion-Inhibitory Protein, which may function as a tumour suppressor. However, *TNFRSF1B* is a stronger candidate gene since the AD GWAS signal at this locus colocalises with the eQTL for *TNFRSF1B* T cells^22,30^, macrophages^20^, fibrobasts^31^ and platelets^29^. Furthermore, *TNFRSF1B* gene expression and the corresponding protein are upregulated in lesional and nonlesional skin compared to controls^24,25,32^ and the PoPs method prioritised this gene at this locus.

RGS14 is a multifunctional cytoplasmic-nuclear shuttling protein which regulates G-protein signalling, but whose role in the immune system is yet to be established. rs4532376 is 10.5kb upstream of *RGS14* and within an intron of *LMAN2*. The AD GWAS signal at this locus colocalises with the eQTL for *RGS14* in macrophages^20^, CD8 T-cells^22^, blood^33^ and colon^23^. *RGS14* has also been shown to be upregulated in lesional skin of AD cases compared to skin from control individuals^25^ and DEPICT prioritises this gene. However, at this locus *LMAN2* is also a reasonably promising candidate (score=15) based on colocalisation and differential expression evidence (Supplementary Table 11). OpenTargets and POSTGAP prioritise this alternative gene at this locus and it is possible that genetic variants at this locus influence AD risk through both genetic mechanisms.

We did not include the 6 novel variants from the multi-ancestry analysis in the comprehensive gene prioritisation pipeline because the available resources used predominantly represent European samples only. We did however investigate these variants using Open Targets Genetics, to identify any evidence implicating specific genes at these loci. rs9247 is a missense variant in *INPP5D*, encoding SHIP1, a protein that functions as a negative regulator of myeloid cell proliferation and survival. The *INPP5D* gene has been implicated in hay fever and/or eczema^5^ and other epithelial barrier disorders including inflammatory bowel disease. rs7773987 and rs17120177 are intronic for *AHI1* and *SIK3*, respectively. *AHI1* (Abelson helper integration site 1) is involved with brain development but it is expressed in a range of tissues throughout the body; single cell analysis in skin shows expression in multiple cell types including specialised immune cells and keratinocytes, but the highest abundance is in endothelial cells (data available from v21.1 proteinatlas.org). *SIK3* encodes a salt-inducible serine/threonine-protein kinase that is predicted to be a positive regulator of mTOR signalling, a pathway of relevance to skin barrier dysfunction as well as inflammation^34^. rs9263868 is in the HLA region, with many plausible candidates in the vicinity. The nearest genes to rs77869365 are *MERTK* (a TAM receptor with an innate immunity role) and *ANAPC1* (part of the anaphase promoting complex in the cell cycle). The closest genes to rs34599047 are *ATG5* (involved in autophagic vesicle formation) and *PRDM1* (which encodes a master regulator of B cells).

### Network analysis

STRING network analysis of the 70 human proteins encoded by genes listed in Tables 1 and 2 showed a protein-protein interaction (PPI) enrichment p-value <1×10^−16^. The five most highly significant (FDR *P*=1.2×10^−9^) Gene Ontology (GO) terms for biological process relate to immune system activation and regulation (Supplementary Table 12). The network described by the highly enriched term ‘Regulation of immune system process’ (GO:0002682) is shown in Figure 4.

**Figure 4.**
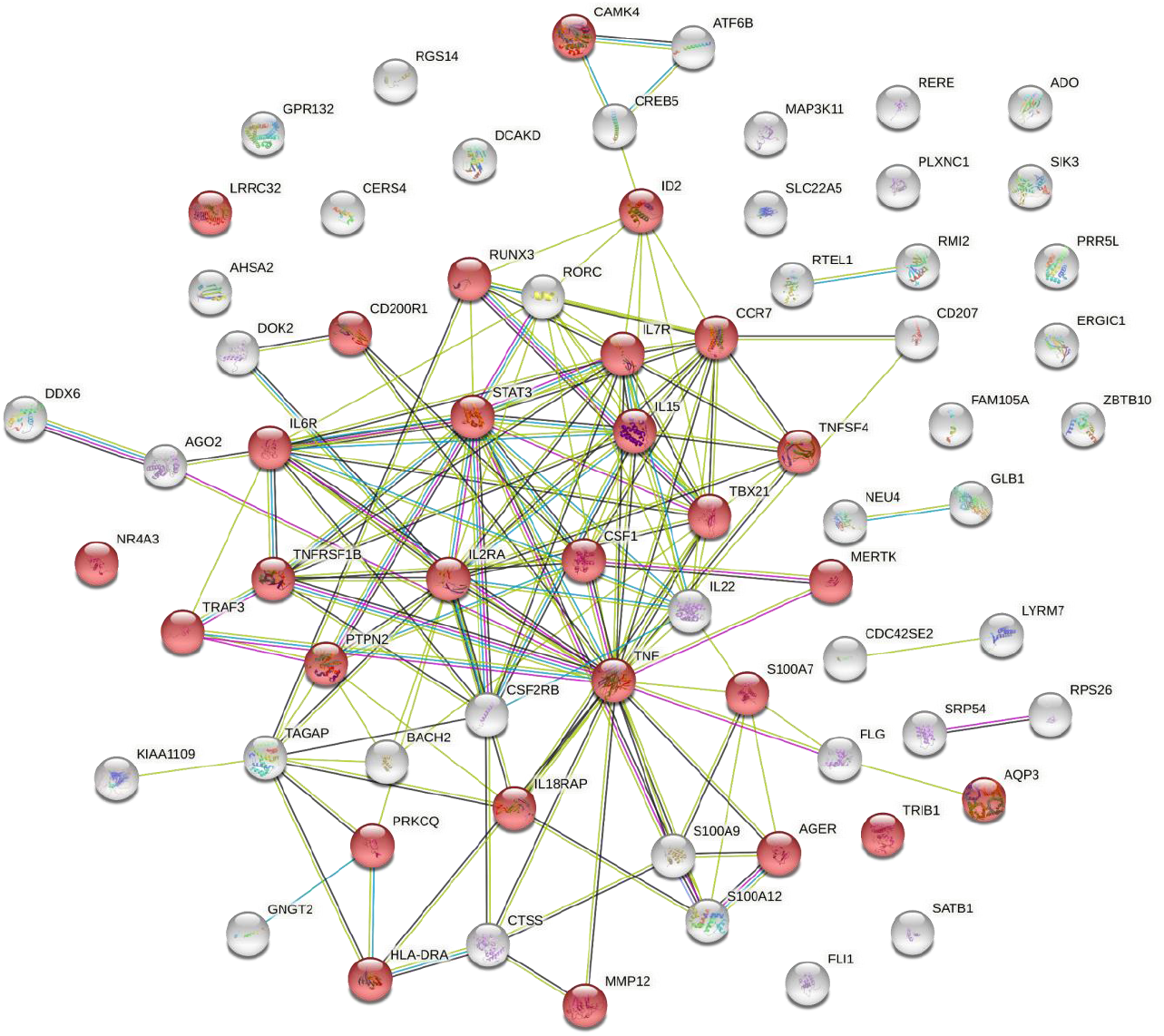
Predicted interaction network of proteins encoded by the top prioritised genes from known and novel European GWAS loci. Protein-protein interaction analysis carried out in STRING v11.5^86^ ; nodes coloured red represent the GO term ‘Regulation of immune system process’ (GO:0002682) for which 28/1514 proteins are included (FDR 1.18× 10^−9^).

Extending the network to include the less well characterised genes/proteins from the multi-ancestry analysis further strengthened this predicted network: The PPI enrichment was again *P*<1×10^−16^ and ‘Regulation of immune system process’ was the most enriched term (FDR *P*=1.6×10^−13^).

## Discussion

We present the results of a comprehensive genome-wide association meta-analysis of AD in which we have identified a total of 96 associated loci. This includes 81 loci identified amongst individuals of European ancestry replicated in a further sample of 2.9 million European individuals (as well as many showing replication in data for other ancestries). Of the additional 15 loci identified in a multi-ancestry analysis, 11 replicated in at least one of the populations tested (European, Latino and African ancestry) and 4 appear to be specific to individuals of East Asian ancestry (but require replication).

The majority of the loci associated with AD are shared between the ancestry groups represented in our data, though there were some notable exceptions. We report 4 previously identified loci (lead SNPs rs9864845, rs34665982, rs45602133, rs4312054) that appear to be specific to the Japanese cohort. Furthermore, we have identified several loci with association in Europeans (many of which also showed association in individuals of Japanese or Latino ancestry) but which showed no evidence of association in individuals of African ancestry. In most cases these differences in associations between ancestries do not seem to be driven by different allele frequencies between the populations, although rs45602133 is an exception where association is seen only in Japanese and the minor allele is at much lower frequency in European and African individuals. The differences in associations between ancestries could be because there are genuine interactions between loci and ancestry (i.e., certain genes only affect AD in specific populations). Alternatively, our results may be an artifact driven by differing linkage disequilibrium between populations, as the lead SNP is unlikely to be the causal variant. Future fine-mapping of these loci in different populations will help to resolve this as well as provide an opportunity to identify causal variants at these loci. However, it is tempting to speculate, using our knowledge of the differing AD phenotypes between European, Asian and African people^35,36^ that the differing genetic architecture may contribute to these clinical observations. rs7773987 within an intron of *AHI1* may, for example, indicate a mechanism contributing to neuronal sensitization leading to the marked lichenification and nodular prurigo-type lesions^37^ that characterise AD in some people of African and European ethnicities^38^. Conversely, the variant rs34665982 for which the nearest gene is *HLA-DRB1* may contribute to the accentuated Th22 and Th17 pathways in patients of Asian ancestry^39^. The use of large scale population cohorts has been useful for identifying association variants. However we do note that the variants identified should be further examined with respect to specific aspects of AD (age of onset, severity and longitudinal classes^40^) in further analysis.

The dominance of blood as the tissue showing most enrichment of our GWAS signals in regions of DNAse hypersensitivity and of eQTLs suggests the importance of systemic inflammation in AD and this is in keeping with knowledge of the multisystem comorbidities associated with AD^41^. The dominance of blood also supports the utility of this easily accessible tissue when characterising genetic risk mechanisms, and for the measurement of biomarkers for many of the implicated loci. However, skin tissue also showed enrichment and there are likely to be some genes for which the effect is only seen in skin. For example, we know that two genes previously implicated in AD, *FLG* and *CD207*^2,18^ are predominantly expressed in the skin and in our gene prioritisation investigations there was no evidence from blood linking *FLG* to the rs61816766 association and only one analysis of monocytes separated from peripheral blood mononuclear cell (PBMC) samples^28^ which implicated *CD207* for the rs112111458 association, amongst an abundance of evidence from skin for both genes playing a role in AD (Supplementary Table 11). So, whilst the enrichment analysis suggests blood as a useful tissue for genome scale studies of AD and a reasonable tissue to include for further investigation at specific loci, it does not preclude skin as the more relevant tissue for a subset of important genes.

At many of the loci identified in this GWAS, our gene prioritisation analysis, as well as the DEPICT pathway analysis, implicated genes from pathways that are already known to have a role in AD pathology. The overwhelming majority of these are in pathways related to immune system function; STRING network analysis highlighted the importance of immune system regulation, in keeping with an increasing awareness of the importance of balance in opposing immune mechanisms that can cause paradoxical atopic or psoriatic skin inflammation^42^. Whilst our *in silico analyses* cannot definitively identify specific <u>causal</u> genes (rather, we present a prioritised list of all genes at each locus along with the corresponding evidence for individual evaluation), it is of note that for many of the previously known loci (Table 1) our approach identifies genes which have been validated in experimental settings, e.g. *FLG*^43^, *TNF*^44^ and *IL22*^45^. The individual components of the gene prioritisation analysis have their limitations, particularly the high probability that findings, whilst demonstrating correlation, do not necessarily provide evidence for a causal relationship. This has been particularly highlighted with respect to colocalisation of GWAS signals with eQTL signals, where high co-regulation can implicate many potentially causal genes^46^. Another limitation is that only cell types (and conditions) that have been studied and made available are included in the *in silico* analysis, and gaps in the data may prove crucial. However, we believe this broad-reaching review of complementary datasets and methods is a useful initial approach to summarise the available evidence, prioritise genes for follow-up and provide information to inform functional experiments. The best evidence is likely to be produced from triangulation of multiple experiments and/or datasets and we have presented our workflow and findings in a way to allow readers to make their own assessments. Another important limitation of our gene prioritisation, is that we only undertook the comprehensive approach for loci associated in European individuals, given that the majority of datasets used come from (and may only be relevant for) European individuals. Expansion of resources that allow for similarly comprehensive follow-up of GWAS loci in individuals of non-European ancestry are urgently needed^47^. However, we do report some evidence that implicates certain genes at loci from our multi-ancestry analysis, whilst noting that these require further investigation in appropriate samples from representative population.

Amongst the genes prioritised at the novel loci identified in this study, four are targets of existing drugs showing the required direction as reported by Open Targets^48^): *CSF1* is targeted by a macrophage colony-stimulating factor 1 inhibiting antibody (in phase II trials as cancer therapy but also for treatment of rheumatoid arthritis and cutaneous lupus); *CTSS* is targeted by a small molecule cathepsin S inhibitor (in phase I-II trials for coeliac disease and Sjogren syndrome); *IL15*, targeted by an anti-IL-15 antibody (in phase II trials for autoimmune conditions including vitiligo and psoriasis); and *MMP12*, targeted by small molecule matrix metalloprotease inhibitors (in phase III studies for breast and lung cancer, plus phase II for cystic fibrosis and COPD). ^49^These may offer valuable drug repurposing opportunities.

We have presented the largest GWAS of AD to date, identifying 96 robustly associated loci, 35 with some evidence of population-specific effects. We have prioritised potentially causal genes at each locus, implicating many immune system genes and pathways.

## Methods

### Phenotype definition

Cases were defined as those who have “ever had atopic dermatitis”, according to the best definition for the cohort, where doctor-diagnosed cases were preferred. Controls were defined as those who had never had AD. Further details on the phenotype definitions for the included studies can be found in the Supplementary Methods and Supplementary Table 2.

### GWAS analysis and quality control of summary-data

We performed genome-wide association analysis (GWAS) for AD case-control status across 40 cohorts including 60,653 AD cases and 804,329 controls of European ancestry. We also included cohorts with individuals of mixed ancestry (Generation R), as well as Japanese (Biobank Japan), African American (SAGE II and SAPPHIRE) and Latino American (GALA II) ancestry, giving a total of 65,107 AD cases and 1,021,287 controls.

Genetic data was imputed separately for each cohort with the majority of European cohorts using haplotype reference consortium (HRC version r1.1) reference panel^49^ (imputed with either the Michigan or Sanger server). 8 European and 2 non-European cohorts instead used the 1000 Genomes Project Phase 1 reference panel for imputation. GWAS was performed separately for each cohort while adjusting for sex and principal components (as appropriate in each cohort). Genetic variants were restricted to a MAF >1% and an imputation quality score > 0.5 unless otherwise specified in the Supplementary Methods. In order to robustly incorporate cohorts with small sample sizes, we applied additional filtering based on the expected minor allele count (EMAC) as previously demonstrated^50^. EMAC combines information on sample size, MAF and imputation quality (2*N*MAF*imputation quality score) and a threshold of >50 EMAC was used to include variants for all cohorts. QQ-plots and Manhattan plots for each cohort were generated and visually inspected as part of the quality control process.

### Meta-analysis

For the discovery phase, meta-analysis of the European cohorts was performed with GWAMA^51^ assuming fixed effects, while the multi-ancestry analysis of all cohorts was conducted in MR-MEGA^52^ (which models the heterogeneity in allelic effects that is correlated with ancestry). *P* < 5×10^−8^ was used to define genome-wide significance. Clumping was performed (in PLINK 1.90^53^) to identify independent loci. We formed clumps of all SNPs which were +/-500kb of each index SNP with a linkage disequilibrium r^2^>0.001. Only the index SNP within each clump is reported.

### Known/Novel assignment

Novel loci are defined as a SNP that had not been reported in a previous GWAS (Supplementary Table 1), or was not correlated (r^2^ < 0.1) with a known SNP from this list. In addition, following assignment of genes to loci (see gene prioritisation) any loci annotated with a gene that has been previously reported were also moved to the ‘known’ list. NB. Therefore, some loci which are reported in Open Targets^54,55^ (but not reported in a published AD GWAS study) have been classed as novel. These loci are marked as such in Table 2.

### Conditional analysis

Conditional analysis was performed to identify any independent secondary association signals in the European meta-analysis. Genome-wide complex trait analysis-conditional and joint analysis (GCTA-COJO^56^) was used to test for independent signals 250kb either side of the index SNPs using UK Biobank HRC imputed data as the reference. COJO-slct was used to determine which SNPs in the region were conditionally independent (*P*<1×10^−5^) and therefore represent independent secondary signals. COJO-cond was then used to condition on the top hit in each region to determine the conditional effect estimates.

### Replication

The genome-wide index SNPs identified from the European and mixed-ancestry discovery meta-analyses were taken forward for replication in 23andMe, Inc. Individuals of European (N=2,904,664), Latino (N=525,348) and African ancestry (N=174,015) were analysed separately. Full details are available in the Supplementary Methods.

### LD score regression

Linkage disequilibrium score (LDSC) regression software (version 1.0.1)^57^ was used to estimate the SNP-based heritability (h^2^_SNP_) for AD. This was performed with the summary statistics of the h^2^_SNP_European discovery meta-analysis. The was estimated on liability scale with a population prevalence of 0.15 and sample prevalence of 0.070.

Genetic correlation with other traits was assessed using all the traits available on CTG-VL58 (accessed on 5^th^ November 2021). We considered phenotypes with p-values below the Bonferroni-corrected alpha threshold (i.e., 0.05/1376=4×10^−5^) to be genetically correlated with AD (a conservative threshold given the likely correlation between many traits tested).

### Bioinformatic analysis

For the following analyses we defined the regions within which the true causal SNP resides to be determined by boundaries containing furthest distanced SNPs with r2 >= 0.2 within +/-500kb of the index SNP (as described previously18). We refer to such regions as locus intervals and we used them as input for FUMA, MendelVar and colocalisation analyses described below.

### Enrichment analysis

Enrichment of tissues and cell types and gene sets for AD GWAS signal was investigated using DEPICT^59^ and GARFIELD (GWAS analysis of regulatory or functional information enrichment with LD correction)^60^ ran with default settings, as well as MAGMA v.1.06^61^ (using GTEx ver. 8^23^ on the FUMA^62^ platform). In addition, we used MendelVar^63^ run with default settings to check for enrichment of any ontology terms assigned to Mendelian disease genes within the locus interval regions.

MAGMA (as default) assigns only SNPs within the gene to each gene. DEPICT maps all genes within a given LD (r^2^>0.5) boundary of the lead SNP to a locus. DEPICT gene set enrichment results for GO terms only were grouped (using the Biological Processes ontology) and displayed using the rrvgo package. The default scatter function was adapted to only plot parent terms^64^.

### Prioritisation of candidate genes

To prioritise candidate genes at each of the loci identified in the European GWAS, we investigated all genes within +/- 500kb of each index SNP (selected to capture an estimated 98% of causal genes)^65^. For each gene we collated evidence from a range of approaches (as described below) to link SNP to gene, resulting in 14 annotation categories (represented as columns in Supplementary Figure 4). We summarised these annotations for each gene into a score in order to prioritise genes at each locus. We present the top prioritised gene in the main tables, but strength of evidence varies and so we encourage readers to use our full evaluation (of all the evidence presented in Supplementary Table 11 for all genes at each locus) for loci of interest.

We tested for colocalisation with molecular QTLs, where full summary statistics were available, using coloc66 method (with betas as input). We used the eQTL Catalogue67 and Open GWAS68 to download a range of eQTL datasets from all skin, whole blood and immune cell types as well as additional tissue types which showed enrichment for our GWAS signals, such as spleen and esophagus mucosa^18^. A complete list of eQTL datasets^20–23,26–31,33,69–73^ is displayed in Supplementary Table 13. pQTL summary statistics for plasma proteins74 were downloaded from Open GWAS. An annotation was included in our gene prioritisation pipeline if there was a posterior probability >95% that the signals from the AD GWAS and the relevant QTL analysis shared the same causal variant.

Additional colocalisation methods were also applied. TWAS (Transcriptome-Wide association Study)-based S-MultiXcan75 and SMR (Summary-based Mendelian Randomization)76 were run on datasets available via the CTG-VL platform (including GTEx tissue types and 2 whole blood pQTL74,77 datasets available for SMR pipeline). For S-MultiXcan and SMR, we report only results with p-values below the alpha threshold established with Bonferroni correction, as well as no evidence of heterogeneity (HEIDI *P*-value > 0.05) in SMR analysis.

Genes were also annotated if they were included in any of the globally enriched ontology/pathway terms from the MendelVar analysis described above or if they were identified in direct look-ups of keywords: “skin”, “kera”, “derma” in their OMIM78 descriptions, or Human Phenotype Ontology79/Disease Ontology80 terms.

We also used machine learning candidate gene prioritization pipelines – DEPICT59, PoPs81, POSTGAP82 and Open Targets Genetics55 Variant 2 Gene mapping tool as well as gene-based MAGMA61 test. We added annotations to genes reported in the top 3 (by each of the pipelines).

We mined the literature for a list of differential expression studies and found 9 RNA-Seq/microarray plus 4 proteomic analyses involving comparisons of AD lesional25,32,83–86 or AD nonlesional24,25,32,84,87–89 skin vs healthy controls. Studies with comparisons of AD lesional acute vs chronic90, blood proteome in AD vs healthy control32 and *FLG* knockdown vs control in living skin-equivalent91 were also included. We annotated each gene (including direction of effect, i.e. upregulated/downregulated) with FDR < 0.05 in any dataset.

Lastly, we annotated genes where the index SNP resided within the coding region according to VEP (Variant Effect Predictor)92 analysis.

For each candidate gene, we established a pragmatic approach to combine all available evidence in order to prioritise which the most plausible candidate gene(s). This prioritisation was carried out as follows:

- The number of annotations (each representing one piece of evidence) were summed across all methods and datasets, to derive a ‘**total evidence score**’, i.e., if coloc evidence was observed for 5 datasets for a particular gene, this would add 5 to the score for that gene.
- Additionally, to assess if evidence was coming from multiple datasets using the same method, or evidence was coming from diverse approaches, we counted ‘**evidence types**’, summing up the methods (as opposed to datasets) with an annotation for each gene tested (up to a maximum of 14), i.e., in the same example of coloc evidence observed in 5 datasest, this would add 1 to this measure for this gene. Evidence types are represented by the columns in Supplementary Figure 4.
- In order to prioritise genes with the most evidence, whilst ensuring there was some evidence of triangulation across methods, at each locus we prioritised the gene with the highest ‘total evidence score’ with a minimum ‘evidence type’ of 3. ‘Evidence type’ was also used to break ties.

### Network analysis

Network analysis of the prioritised genes was carried out using standard settings (minimum interaction score 0.4) in STRING v11.5^93^.

## Data availability

Full summary statistics of the GWAS meta-analyses will be made available on publication.

The variant-level data for the 23andMe replication dataset are fully disclosed in the main tables and supplementary tables. Individual-level data are not publicly available due to participant confidentiality, and in accordance with the IRB-approved protocol under which the study was conducted.

## Data Availability

Full summary statistics of the GWAS meta-analyses will be made available on publication.
The variant-level data for the 23andMe replication dataset are fully disclosed in the main tables and supplementary tables. Individual-level data are not publicly available due to participant confidentiality, and in accordance with the IRB-approved protocol under which the study was conducted.

## Funding & acknowledgements

For this work, AB-A, SJB and LP were funded by the Innovative Medicines Initiative 2 Joint Undertaking (JU) under grant agreement No 821511 (BIOMAP). The JU receives support from the European Union’s Horizon 2020 research and innovation programme and EFPIA.

AB-A, MKS, JM and LP and work in a research unit funded by the UK Medical Research Council (MC_UU_00011/1). LP received funding from the British Skin Foundation (8010 Innovative Project) and the Academy of Medical Sciences Springboard Award, which is supported by the Wellcome Trust, The Government Department for Business, Energy and Industrial Strategy, the Global Challenges Research Fund and the British Heart Foundation [SBF003\1094]. SJB holds a Wellcome Trust Senior Research Fellowship in Clinical Science [220875/Z/20/Z].

Thanks to Sergi Sayols (developer of rrvgo), who provided additional code to alter the scatter plot produced by rrvgo to only display parent terms.

This publication is the work of the authors and LP will serve as guarantor for the contents of this paper.

This work was carried out using the computational facilities of the Advanced Computing Research Centre, University of Bristol - http://www.bristol.ac.uk/acrc/.

Individual cohort acknowledgements are in the Supplementary Methods.

## Conflict of interest

KMG has received reimbursement for speaking at conferences sponsored by companies selling nutritional products, and is part of an academic consortium that has received research funding from Abbott Nutrition, Nestec, BenevolentAI Bio Ltd. and Danone.

CG, SSS and 23andMe Research Team are employed by and hold stock or stock options in 23andMe, Inc.

**Designed and co ordinated the study:** M.S., L.P.

**Performed the meta analysis:** A.B.A., A. Kilanowski

**Performed the bioinformatic analysis:** M.K.S.

**Performed the STRING analysis:** S.J. Brown

**Performed statistical analysis within cohorts:** A.B.A., A. Kilanowski, S.S.S., R. Mitchell, K.R., R. Mägi, M.N., N.T., B.M.B., L.F.T., P.S.N., C.F., A.E.O., E.H.L., J.V.T.L., J.B.J., I.M., A.A.S., A.J., H. Baurecht, E.R., C.M.G., C.H., C.Q., P.T., E.A., J.F., C.A.W., E.T., S.X., S.K., D.M.K., L.K., J.D., H.Z., C.A., V.U., R.K., A. Szwajda, A.C.S.N.J., A.G., M.I., M. Müller-Nurasyid, T.S.A., M.B., C.G., M.P.Y., D.P.S., N.G., Y.A.L., A.D.I., K.L.W., C.M., S.J. Brown

**Data acquisition/supported analysis/interpretation of data:** A.B.A., A. Kilanowski, A.R., P.S.N., A.J., C.j.X., S.E.H., J.F., E.T., S.K., H.Z., T. Hoffmann, E.J., H.C., N.R., P.N., O.A.A., S.J., C.A., T.G., V.U., P.K.E.M., E.G.B., J.P.T., T. Hansen, L.L.K., T.M.D., A.A., G.H., S.L., M.M. Nöthen, N.H., M.I., T.S.A., J.S., B.C., A.M.M.S., A.E., T.E., L.A.M., A.M., C.T., K.A., M.L., K.H., B.J., D.P.S., Y.A.L., N.P.H., S.W., A.D.I., D.L.J., T.N., L.D., J.M.V., G.H.K., K.M.G., B.F., C.E.P., P.D.S., P.G.H., H. Bisgaard, K.B., J.C., A. Simpson, T.S., S.J. Brown, M.S., L.P.

**Wrote the paper:** A.B.A., A. Kilanowski, M.K.S., S.J. Brown, M.S., L.P.

**Approved final version of paper:** A.B.A., A. Kilanowski, M.K.S., S.S.S., R. Mitchell, K.R., A.R., R. Mägi, M.N., N.T., B.M.B., L.F.T., P.S.N., C.F., A.E.O., E.H.L., J.V.T.L., J.B.J., I.M., A.A.S., A.J., K.A.F., H. Baurecht, E.R., A.C.A., A. Kumar, P.M.S., X.C., C.M.G., C.H., C.j.X., C.Q., S.E.H., P.T., E.A., J.F., C.A.W., E.T., S.X., S.K., D.M.K., L.K., J.D., H.Z., T. Hoffmann, E.J., H.C., N.R., P.N., O.A.A., S.J., C.A., T.G., V.U., R.K., P.K.E.M., A. Szwajda, E.G.B., J.P.T., T. Hansen, L.L.K., T.M.D., A.C.S.N.J., A.G., A.A., G.H., S.L., M.M. Nöthen, N.H., M.I., A.V., M.F., V.B., P.H., N.B., D.I.B., J.J.H., M. Müller-Nurasyid, T.S.A., J.S., B.C., A.M.M.S., A.E., M.B., B.R., H.A., C.G., NA, T.E., L.A.M., A.M., C.T., K.A., M.L., K.H., B.J., M.P.Y., D.P.S., N.G., A.L., Y.A.L., N.P.H., S.W., M.R.J., E.M., H.H., A.D.I., D.L.J., T.N., L.D., J.M.V., G.H.K., K.M.G., S.J. Barton, B.F., C.E.P., P.D.S., P.G.H., K.L.W., H. Bisgaard, K.B., J.C., A. Simpson, C.M., T.S., S.B., S.T.W., J.W.H., J.M., S.J. Brown, M.S., L.P.

## Notes

### Author Declarations

All relevant ethical regulations were followed and informed consent was obtained from all participants. Ethical approval for all cohorts were approved by their local ethics committees as detailed in the Supplementary Methods.

### Summary of Updates

This version of the manuscript has been revised to update the funding details.

## References

1. Weidinger, S. et al. Atopic dermatitis. The Lancet 387, 1109–1122 (2016).

2. Paternoster, L. et al. Multi-ancestry genome-wide association study of 21,000 cases and 95,000 controls identifies new risk loci for atopic dermatitis. Nat Genet 47, 1449–1456 (2015).

3. Paternoster, L. et al. Meta-analysis of genome-wide association studies identifies three new risk loci for atopic dermatitis. Nat Genet 44, 187–192 (2012).

4. Weidinger, S. et al. A genome-wide association study of atopic dermatitis identifies loci with overlapping effects on asthma and psoriasis. Hum Mol Genet 22, 4841–56 (2013).

5. Johansson, Å., Rask-Andersen, M., Karlsson, T. & Ek, W. E. Genome-wide association analysis of 350 000 Caucasians from the UK Biobank identifies novel loci for asthma, hay fever and eczema. Association Studies Article 28, 4022–4041 (2019).

6. Sliz, E. et al. Uniting biobank resources reveals novel genetic pathways modulating susceptibility for atopic dermatitis. Journal of Allergy and Clinical Immunology 0, (2021).

7. Grosche, S. et al. Rare variant analysis in eczema identifies exonic variants in DUSP1, NOTCH4 and SLC9A4. Nature Communications 2021 12:1 12, 1–11 (2021).

8. Tanaka, N. et al. Eight novel susceptibility loci and putative causal variants in atopic dermatitis. Journal of Allergy and Clinical Immunology 148, 1293–1306 (2021).

9. Schaarschmidt, H. et al. A genome-wide association study reveals 2 new susceptibility loci for atopic dermatitis. Journal of Allergy and Clinical Immunology 136, 802–806 (2015).

10. Hirota, T. et al. Genome-wide association study identifies eight new susceptibility loci for atopic dermatitis in the Japanese population. Nat Genet 44, 1222–1226 (2012).

11. Kim, K. W. et al. Genome-wide association study of recalcitrant atopic dermatitis in Korean children. J Allergy Clin Immunol 136, 678-684.e4 (2015).

12. Sun, L.-D. et al. Genome-wide association study identifies two new susceptibility loci for atopic dermatitis in the Chinese Han population. Nat Genet 43, 690–694 (2011).

13. Esparza-Gordillo, J. et al. A functional IL-6 receptor (IL6R) variant is a risk factor for persistent atopic dermatitis. Journal of Allergy and Clinical Immunology 132, 371–377 (2013).

14. Ellinghaus, D. et al. High-density genotyping study identifies four new susceptibility loci for atopic dermatitis. Nat Genet 45, 808–12 (2013).

15. Larsen, F. S., Holm, N. v & Henningsen, K. Atopic dermatitis. A genetic-epidemiologic study in a population-based twin sample. J Am Acad Dermatol 15, 487–94 (1986).

16. Schultz Larsen, F. Atopic dermatitis: a genetic-epidemiologic study in a population-based twin sample. J Am Acad Dermatol 28, 719–23 (1993).

17. Budu-Aggrey, A. et al. Investigating the causal relationship between allergic disease and mental health. Clin Exp Allergy 51, 1449–1458 (2021).

18. Sobczyk, M. K. et al. Triangulating Molecular Evidence to Prioritize Candidate Causal Genes at Established Atopic Dermatitis Loci. Journal of Investigative Dermatology 141, 2620–2629 (2021).

19. Zeng, Z. et al. Roles of G protein-coupled receptors in inflammatory bowel disease. http://www.wjgnet.com/ 26, 1242–1261 (2020).

20. Alasoo, K. et al. Shared genetic effects on chromatin and gene expression indicate a role for enhancer priming in immune response. Nat Genet 50, 424 (2018).

21. Chen, L. et al. Genetic Drivers of Epigenetic and Transcriptional Variation in Human Immune Cells. Cell 167, 1398-1414.e24 (2016).

22. Schmiedel, B. J. et al. Impact of Genetic Polymorphisms on Human Immune Cell Gene Expression. Cell 175, 1701-1715.e16 (2018).

23. GTEx Consortium. The GTEx Consortium atlas of genetic regulatory effects across human tissues. Science 369, 1318–1330 (2020).

24. Winge, M. C. G. et al. Filaggrin genotype determines functional and molecular alterations in skin of patients with atopic dermatitis and ichthyosis vulgaris. PLoS One 6, e28254 (2011).

25. He, H. et al. Tape strips detect distinct immune and barrier profiles in atopic dermatitis and psoriasis. J Allergy Clin Immunol 147, 199–212 (2021).

26. Fairfax, B. P. et al. Innate immune activity conditions the effect of regulatory variants upon monocyte gene expression. Science 343, 1246949 (2014).

27. Nédélec, Y. et al. Genetic Ancestry and Natural Selection Drive Population Differences in Immune Responses to Pathogens. Cell 167, 657-669.e21 (2016).

28. Quach, H. et al. Genetic Adaptation and Neandertal Admixture Shaped the Immune System of Human Populations. Cell 167, 643-656.e17 (2016).

29. Momozawa, Y. et al. IBD risk loci are enriched in multigenic regulatory modules encompassing putative causative genes. Nat Commun 9, 2427 (2018).

30. Kasela, S. et al. Pathogenic implications for autoimmune mechanisms derived by comparative eQTL analysis of CD4+ versus CD8+ T cells. PLoS Genet 13, e1006643 (2017).

31. Gutierrez-Arcelus, M. et al. Passive and active DNA methylation and the interplay with genetic variation in gene regulation. Elife 2013, (2013).

32. Pavel, A. B. et al. The proteomic skin profile of moderate-to-severe atopic dermatitis patients shows an inflammatory signature. J Am Acad Dermatol 82, 690–699 (2020).

33. Buil, A. et al. Gene-gene and gene-environment interactions detected by transcriptome sequence analysis in twins. Nat Genet 47, 88 (2015).

34. Naeem, A. S. et al. A mechanistic target of rapamycin complex 1/2 (mTORC1)/V-Akt murine thymoma viral oncogene homolog 1 (AKT1)/cathepsin H axis controls filaggrin expression and processing in skin, a novel mechanism for skin barrier disruption in patients with atopic dermatitis. J Allergy Clin Immunol 139, 1228–1241 (2017).

35. Nomura, T., Wu, J., Kabashima, K. & Guttman-Yassky, E. Endophenotypic Variations of Atopic Dermatitis by Age, Race, and Ethnicity. J Allergy Clin Immunol Pract 8, 1840–1852 (2020).

36. Yew, Y. W., Thyssen, J. P. & Silverberg, J. I. A systematic review and meta-analysis of the regional and age-related differences in atopic dermatitis clinical characteristics. J Am Acad Dermatol 80, 390–401 (2019).

37. Ständer, H. F., Elmariah, S., Zeidler, C., Spellman, M. & Ständer, S. Diagnostic and treatment algorithm for chronic nodular prurigo. J Am Acad Dermatol 82, 460–468 (2020).

38. Sangha, A. M. Dermatological Conditions in SKIN OF COLOR-: Managing Atopic Dermatitis. J Clin Aesthet Dermatol 14, S20–S22 (2021).

39. Tokura, Y. & Hayano, S. Subtypes of atopic dermatitis: From phenotype to endotype. Allergol Int 71, 14–24 (2022).

40. Paternoster, L. et al. Identification of atopic dermatitis subgroups in children from 2 longitudinal birth cohorts. Journal of Allergy and Clinical Immunology 141, 964–971 (2018).

41. Langan, S. M., Irvine, A. D. & Weidinger, S. Atopic dermatitis. The Lancet 396, 345–360 (2020).

42. Al-Janabi, A., Foulkes, A. C., Griffiths, C. E. M. & Warren, R. B. Paradoxical eczema in patients with psoriasis receiving biologics: a case series. Clin Exp Dermatol 47, 1174–1178 (2022).

43. McAleer, M. A. & Irvine, A. D. The multifunctional role of filaggrin in allergic skin disease. J Allergy Clin Immunol 131, 280–91 (2013).

44. Danso, M. O. et al. TNF-α and Th2 Cytokines Induce Atopic Dermatitis–Like Features on Epidermal Differentiation Proteins and Stratum Corneum Lipids in Human Skin Equivalents. Journal of Investigative Dermatology 134, 1941–1950 (2014).

45. Gutowska-Owsiak, D., Schaupp, A. L., Salimi, M., Taylor, S. & Ogg, G. S. Interleukin-22 downregulates filaggrin expression and affects expression of profilaggrin processing enzymes. British Journal of Dermatology 165, 492–498 (2011).

46. Wainberg, M. et al. Opportunities and challenges for transcriptome-wide association studies. Nat Genet 51, 592 (2019).

47. Hindorff, L. A. et al. Prioritizing diversity in human genomics research. Nat Rev Genet 19, 175– 185 (2018).

48. Ochoa, D. et al. Open Targets Platform: supporting systematic drug–target identification and prioritisation. Nucleic Acids Res 49, D1302–D1310 (2021).

49. McCarthy, S. et al. A reference panel of 64,976 haplotypes for genotype imputation. Nat Genet 48, 1279–1283 (2016).

50. Bustamante, M. et al. A genome-wide association meta-analysis of diarrhoeal disease in young children identifies FUT2 locus and provides plausible biological pathways. Hum Mol Genet 25, 4127–4142 (2016).

51. Mägi, R. & Morris, A. P. GWAMA: Software for genome-wide association meta-analysis. BMC Bioinformatics (2010) doi:10.1186/1471-2105-11-288.

52. Mägi, R. et al. Trans-ethnic meta-regression of genome-wide association studies accounting for ancestry increases power for discovery and improves fine-mapping resolution. Hum Mol Genet 26, 3639–3650 (2017).

53. Purcell, S. et al. PLINK: a tool set for whole-genome association and population-based linkage analyses. Am J Hum Genet 81, 559–75 (2007).

54. Ghoussaini, M. et al. Open Targets Genetics: systematic identification of trait-associated genes using large-scale genetics and functional genomics. Nucleic Acids Res 49, D1311–D1320 (2021).

55. Mountjoy, E. et al. An open approach to systematically prioritize causal variants and genes at all published human GWAS trait-associated loci. Nat Genet 53, 1527–1533 (2021).

56. Yang, J. et al. Conditional and joint multiple-SNP analysis of GWAS summary statistics identifies additional variants influencing complex traits. Nat Genet 44, 369–75, S1-3 (2012).

57. Zheng, J. et al. LD Hub: A centralized database and web interface to perform LD score regression that maximizes the potential of summary level GWAS data for SNP heritability and genetic correlation analysis. Bioinformatics 33, 272–279 (2017).

58. Cuéllar-Partida, G. et al. Complex-Traits Genetics Virtual Lab: A community-driven web platform for post-GWAS analyses. bioRxiv (2019) doi:10.1101/518027.

59. Pers, T. H. et al. Biological interpretation of genome-wide association studies using predicted gene functions Genetic Investigation of ANthropometric Traits (GIANT) Consortium. Nat Commun (2015) doi:10.1038/ncomms6890.

60. Iotchkova, V. et al. GARFIELD classifies disease-relevant genomic features through integration of functional annotations with association signals. Nat Genet 51, 343–353 (2019).

61. de Leeuw, C. A., Mooij, J. M., Heskes, T. & Posthuma, D. MAGMA: Generalized Gene-Set Analysis of GWAS Data. PLoS Comput Biol 11, 1004219 (2015).

62. Watanabe, K., Taskesen, E., van Bochoven, A. & Posthuma, D. Functional mapping and annotation of genetic associations with FUMA. Nat Commun 8, 1–11 (2017).

63. Sobczyk, M. K., Gaunt, T. R. & Paternoster, L. MendelVar: gene prioritization at GWAS loci using phenotypic enrichment of Mendelian disease genes. Bioinformatics 37, 1–8 (2021).

64. Sayols, S. rrvgo: a Bioconductor package to reduce and visualize Gene Ontology terms. https://ssayols.github.io/rrvgo (2020).

65. Stacey, D. et al. ProGeM: a framework for the prioritization of candidate causal genes at molecular quantitative trait loci. Nucleic Acids Res 47, e3–e3 (2019).

66. Giambartolomei, C. et al. Bayesian test for colocalisation between pairs of genetic association studies using summary statistics. PLoS Genet 10, e1004383 (2014).

67. Kerimov, N. et al. A compendium of uniformly processed human gene expression and splicing quantitative trait loci. Nat Genet 53, 1290–1299 (2021).

68. Elsworth, B. et al. The MRC IEU OpenGWAS data infrastructure. bioRxiv (2020) doi:10.1101/2020.08.10.244293.

69. Võsa, U. et al. Large-scale cis- and trans-eQTL analyses identify thousands of genetic loci and polygenic scores that regulate blood gene expression. Nat Genet 53, 1300–1310 (2021).

70. Fairfax, B. P. et al. Genetics of gene expression in primary immune cells identifies cell type-specific master regulators and roles of HLA alleles. Nat Genet 44, 502–10 (2012).

71. Lappalainen, T. et al. Transcriptome and genome sequencing uncovers functional variation in humans. Nature 501, 506–11 (2013).

72. Lepik, K. et al. C-reactive protein upregulates the whole blood expression of CD59 - an integrative analysis. PLoS Comput Biol 13, e1005766 (2017).

73. Naranbhai, V. et al. Genomic modulators of gene expression in human neutrophils. Nat Commun 6, 7545 (2015).

74. Sun, B. B. et al. Genomic atlas of the human plasma proteome. Nature 558, 73–79 (2018).

75. Barbeira, A. N. et al. Integrating predicted transcriptome from multiple tissues improves association detection. PLoS Genet 15, e1007889 (2019).

76. Zhu, Z. et al. Integration of summary data from GWAS and eQTL studies predicts complex trait gene targets. Nat Genet 48, 481–7 (2016).

77. Suhre, K. et al. Connecting genetic risk to disease end points through the human blood plasma proteome. Nat Commun 8, 14357 (2017).

78. Amberger, J. S., Bocchini, C. A., Scott, A. F. & Hamosh, A. OMIM.org: leveraging knowledge across phenotype-gene relationships. Nucleic Acids Res 47, D1038–D1043 (2019).

79. Köhler, S. et al. Expansion of the Human Phenotype Ontology (HPO) knowledge base and resources. Nucleic Acids Res 47, D1018–D1027 (2019).

80. Schriml, L. M. et al. Human Disease Ontology 2018 update: classification, content and workflow expansion. Nucleic Acids Res 47, D955–D962 (2019).

81. Weeks, E. M. et al. Leveraging polygenic enrichments of gene features to predict genes underlying complex traits and diseases. medRxiv (2020) doi:10.1101/2020.09.08.20190561.

82. Peat, G. et al. The open targets post-GWAS analysis pipeline. Bioinformatics 36, 2936–2937 (2020).

83. Rojahn, T. B. et al. Single-cell transcriptomics combined with interstitial fluid proteomics defines cell type-specific immune regulation in atopic dermatitis. J Allergy Clin Immunol 146, 1056–1069 (2020).

84. Pavel, A. B. et al. Tape strips from early-onset pediatric atopic dermatitis highlight disease abnormalities in nonlesional skin. Allergy 76, 314–325 (2021).

85. Dyjack, N. et al. Minimally invasive skin tape strip RNA sequencing identifies novel characteristics of the type 2-high atopic dermatitis disease endotype. J Allergy Clin Immunol 141, 1298–1309 (2018).

86. Molin, S. et al. The hand eczema proteome: imbalance of epidermal barrier proteins. Br J Dermatol 172, 994–1001 (2015).

87. Cole, C. et al. Filaggrin-stratified transcriptomic analysis of pediatric skin identifies mechanistic pathways in patients with atopic dermatitis. Journal of Allergy and Clinical Immunology 134, 82–91 (2014).

88. Ewald, D. A. et al. Meta-analysis derived atopic dermatitis (MADAD) transcriptome defines a robust AD signature highlighting the involvement of atherosclerosis and lipid metabolism pathways. BMC Med Genomics 8, 60 (2015).

89. Morelli, P. et al. Proteomic analysis from skin swabs reveals a new set of proteins identifying skin impairment in atopic dermatitis. Exp Dermatol 30, 811–819 (2021).

90. Tsoi, L. C. et al. Progression of acute-to-chronic atopic dermatitis is associated with quantitative rather than qualitative changes in cytokine responses. J Allergy Clin Immunol 145, 1406–1415 (2020).

91. Elias, M. S. et al. Proteomic analysis of filaggrin deficiency identifies molecular signatures characteristic of atopic eczema. J Allergy Clin Immunol 140, 1299–1309 (2017).

92. McLaren, W. et al. The Ensembl Variant Effect Predictor. Genome Biol 17, 122 (2016).

93. Szklarczyk, D. et al. STRING v11: protein–protein association networks with increased coverage, supporting functional discovery in genome-wide experimental datasets. Nucleic Acids Res 47, D607–D613 (2019).

94. Kichaev, G. et al. Leveraging Polygenic Functional Enrichment to Improve GWAS Power. Am J Hum Genet 104, 65–75 (2019).

95. http://www.nealelab.is/uk-biobank/.

