## Supplementary tables for "European and multi-ancestry genome-wide association meta-analysis of atopic dermatitis highlights importance of systemic immune regulation"

**Supplementary table legends**

**Supplementary Table 1. Previously associated variants.** Association estimates are derived from the European discovery meta-analysis and the multi-ancestry meta-analysis of the current study. References are listed in the main text. The ancestries within which the associations were originally identified are reported.
Chr, chromosome; E/O, effect allele/ other allele; SE, standard error; *P*, *P*-value.

**Supplementary Table 2. Cohort details.** Characteristics of the European discovery cohorts, multi-ancestry cohorts, and replication cohorts.

**Supplementary Table 3. Conditional analyses.** Crude association is the association of the secondary SNP unadjusted for the primary SNP. Conditional association is the association of the secondary SNP adjusted for the primary SNP.
SNP, single nucleotide polymorphism; Chr, chromosome; EA, effect allele; EAF, effect allele frequency; OR, odds ratio; CI, confidence interval; *P*, *P*-value.

**Supplementary Table 4. Multi-ancestry results.** 19 loci meeting genome-wide significance in the multi-ancestry meta-analysis. Replication estimates are derived from the RIKEN cohort for Japanese individuals, and 23andMe for individuals of Latino, African and European ancestry.
Chr, chromosome; EAF, effect allele frequency; *P*, *P*-value

**Supplementary Table 5. Allele frequency comparison.** Allele frequencies for the 81 variants from the European discovery meta-analysis, and the 19 variants from the multi-ancestry meta-analysis. Allele frequencies derived from the European discovery meta-analysis, RIKEN cohort and 23andMe for individuals of European, Japanese, Latino and African ancestry respectively.
Chr, chromosome; EA, effect allele; OA, other allele

**Supplementary Table 6. Genetic correlations.** Results from LD score regression analysis against all phenotypes available on CTG-VL (5th Nov 2021). Genetic correlations (rg) with the corresponding p-values (p) are displayed for each tested trait with atopic dermatitis.
rg, genetic correlation; se, standard error of rg; z, z-score; p, p-value for rg; h2_obs and h2_obs_se, observed scale heritability for trait 2 and standard error; h2_int and h2_int_se, single-trait LD Score regression intercept for trait 2 and standard error; gcov_int and gcov_int_se, cross-trait LD Score regression intercept and standard error; N, number of individuals; NCases, number of cases; NControls, number of controls

**Supplementary Table 7. Garfield DHS peak enrichment.** Enrichment was tested at two GWAS *P*-value threshold ($1x10^{-8}$ and $1x10^{-5}$, PThresh). The OR and *P*-value for enrichment are given in the corresponding columns for each cell type tested (only DHS 'peaks' are reported in the main text).

**Supplementary Table 8. MAGMA GTEx enrichment.** Beta, SE and P for enrichment are reported for each tissue type tested in the GTEx ver.8 expression dataset.

**Supplementary Table 9. DEPICT pathway analysis.** The nominal P-value for enrichment is reported for each gene set term tested. Gene sets significant at 5% FDR are highlighted.

**Supplementary Table 10. Post-GWAS gene prioritisation – top 3 genes per locus.** For each locus, the top 3 genes (along with their Total Evidence Scores) are presented.

**Supplementary Table 11. Post-GWAS gene prioritisation – all genes tested.** For each locus, the available evidence implicating each gene tested is summarised. Columns H to U summarises the evidence from specific methods. The final 2 columns display the 2 summary scores generated and used to prioritise genes at each locus.

**Supplementary Table 12. STRING network analysis.** Top prioritised genes from the 81 European loci are used as input.

**Supplementary Table 13. eQTL datasets used in post-GWAS prioritisation.** Reference numbers correspond to references in the main text.

### Supplementary Figure legends

**Supplementary Figure 1. Locus zoom plots.** LocusZoom plots were created with the downloadable standalone version of the programme (V1.4). SNPs genome-wide significant in both meta-analyses are displayed next to each other (European-only meta-analysis on the left, multi-ancestry meta-analysis on the right). For SNPs only genome-wide significant in one of the meta-analyses, only one LocusZoom plots is shown.

**Supplementary Figure 2. Forest plots.** Plots of all genome-wide significant SNPs from both, European-only and Multi-ancestry, meta-analyses. For each lead SNP the summary statistics of all cohorts are presented. Meta-analysis results are also presented for the European cohorts with $I^2$ as a measure of heterogeneity. *Marked SNPs showed a batch effect in 23andme Latinos but are still included in the dataset.

**Supplementary Figure 3. Genetic correlations.** Plot of all genetic correlations (with atopic dermatitis) as generated from LD score regression analysis. Strength of correlation evidence ($-\log_{10}$ *P*-value) is plotted against correlation coefficient (rg), with increasing negative correlations presented towards the left and positive correlations towards the right of the plot. Traits with good evidence for correlation ($P<=4x10^{-5}$) are labelled.

**Supplementary Figure 4. Pathway enrichment analysis.** DEPICT analysis of GO terms. Terms with significant (<5% FDR) enrichment in our GWAS were grouped (by similarity) using rrvgo, here the first 2 components of a principal coordinate analysis of the similarity matrix for parent GO terms are plotted. Size of the point represents the group size (i.e. the number of enriched child GO terms represented by the parent term). Distances between points represent the similarity of terms.

**Supplementary Figure 5. Gene prioritisation.** Summary of evidence from GWAS gene prioritisation methods. Counts within the heatmap represent the total of individual contributing results for these methods, combining evidence from across different tissues and studies. Size of black circle represents the rank given to a gene by a particular method (only genes ranked 1-3 in each method are shown), while blue diamonds represent presence of a given annotation. The evidence is summarised in a 'total evidence score' and an 'evidence types score', as described in the methods.
