## Supplementary Figure 1 for "European and multi-ancestry genome-wide association meta-analysis of atopic dermatitis highlights importance of systemic immune regulation"

### Discovered in European ancestry analysis

## 1:8476441 - rs301804

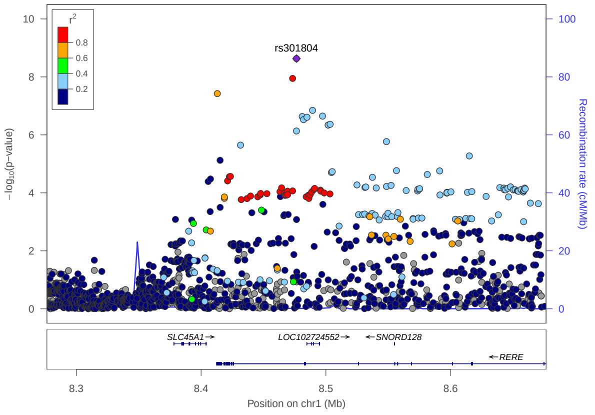


## 1:12091024 - rs61776548

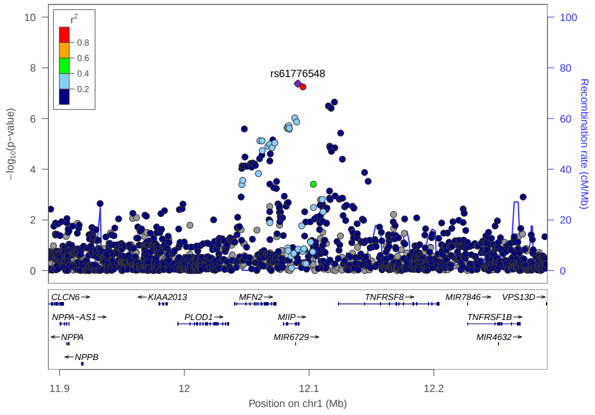


## 1:25294618 - rs7542147

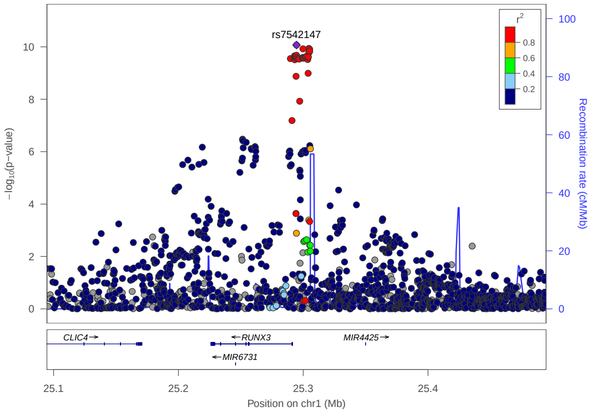


## 1:110371629 - rs12565349

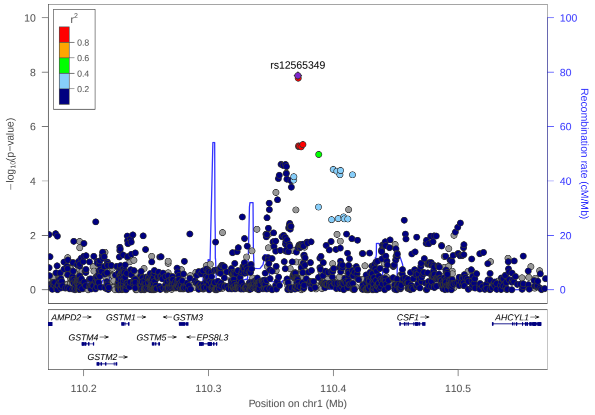


## 1:150374354 - rs187080438

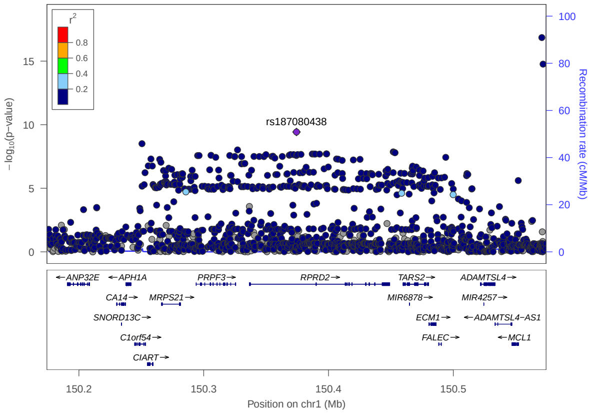


## 1:151059196 - rs146527530

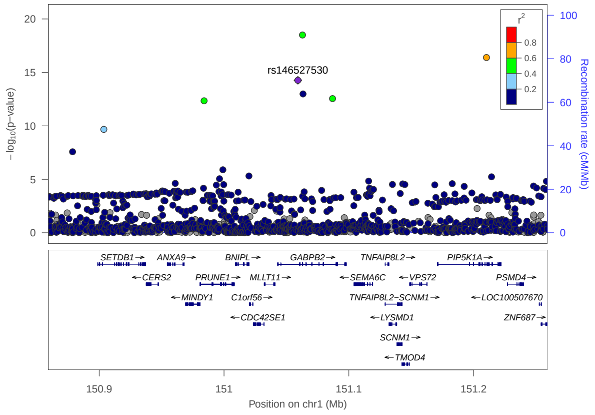

1:151063299 - rs115161931

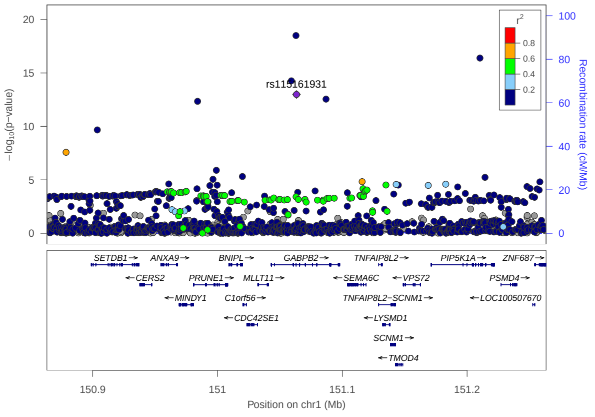


1:151625094 - rs71625130

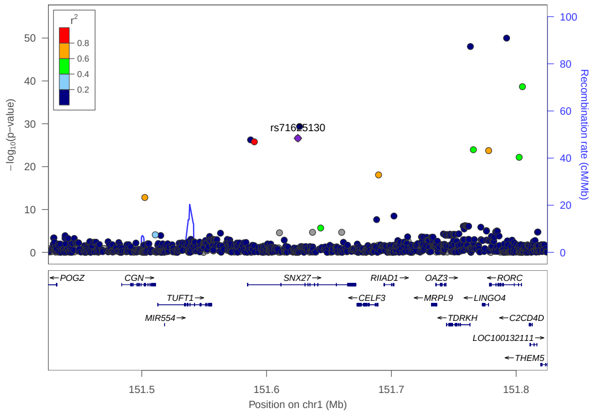


1:151626396 - rs149199808

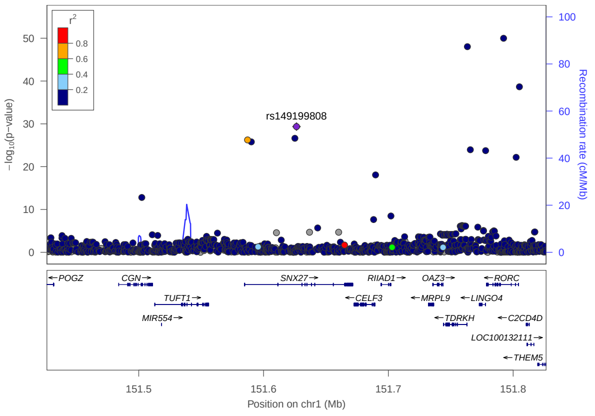


1:152179152 - rs12123821

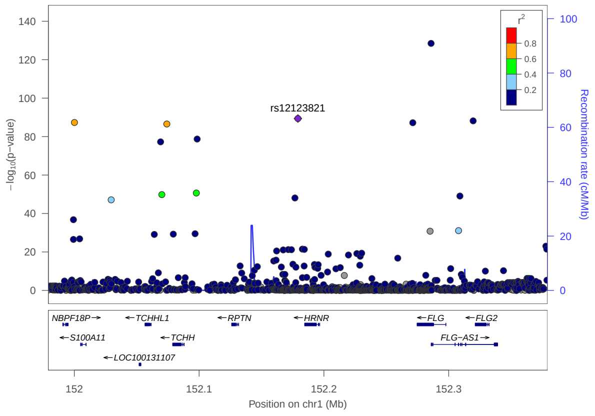


1:152319572 - rs61816766

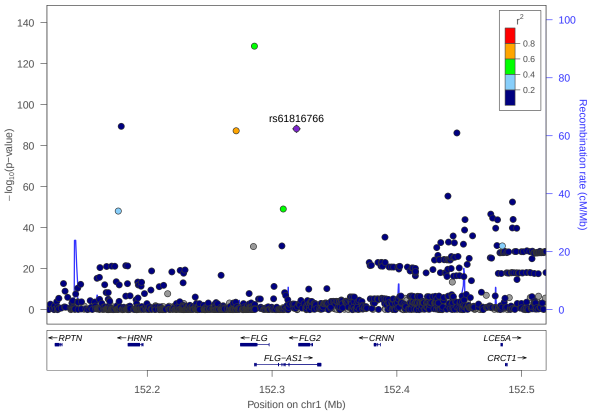


1:152771963 - rs72702900

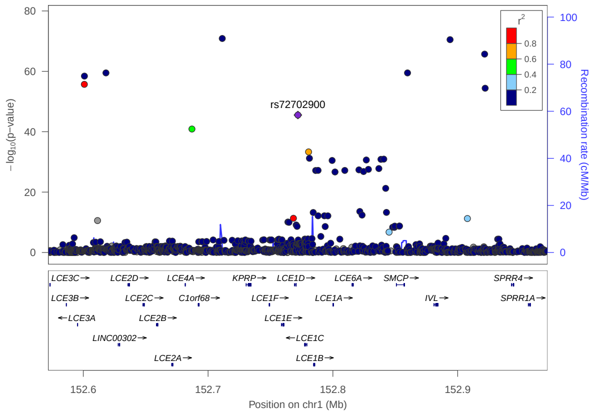

# 1:152893891 - rs61815704

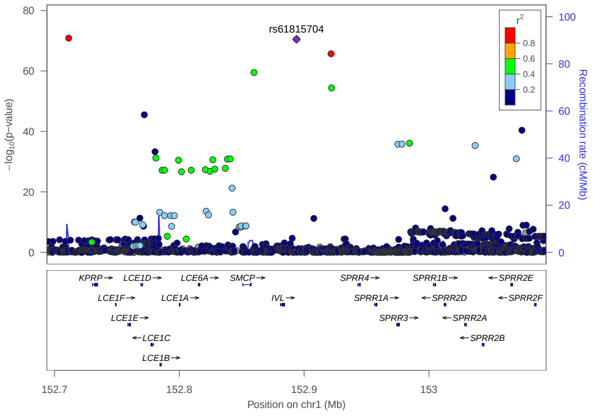


# 1:153275443 - rs821429

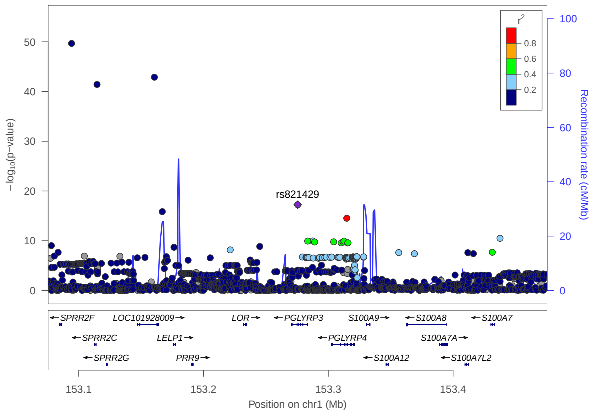


# 1:153843489 - rs12138773

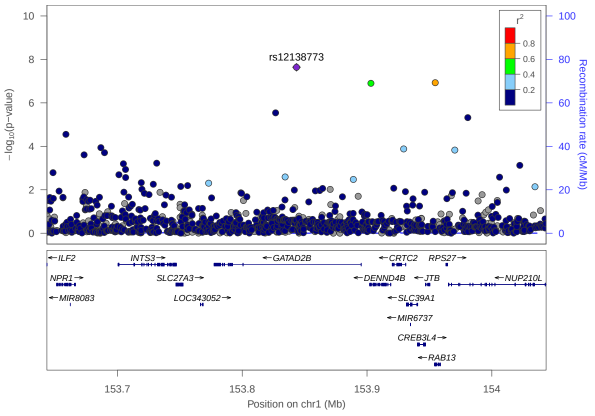


# 1:154428283 - rs12133641

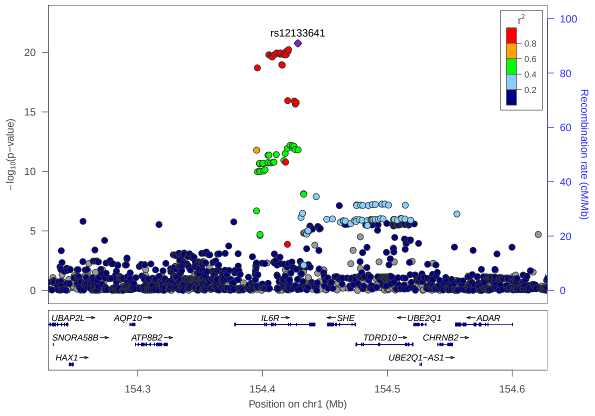


# 1:172744543 - rs859723

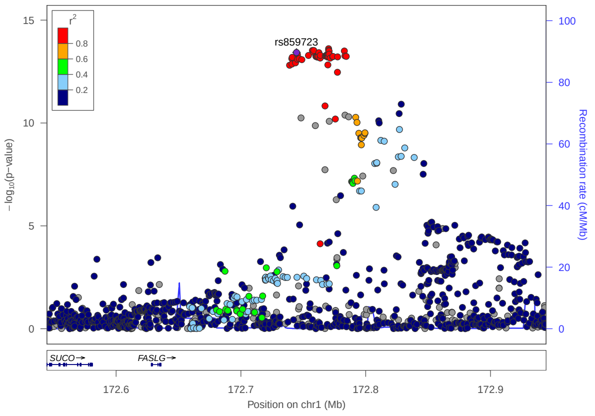


# 1:173150727 - rs11811788

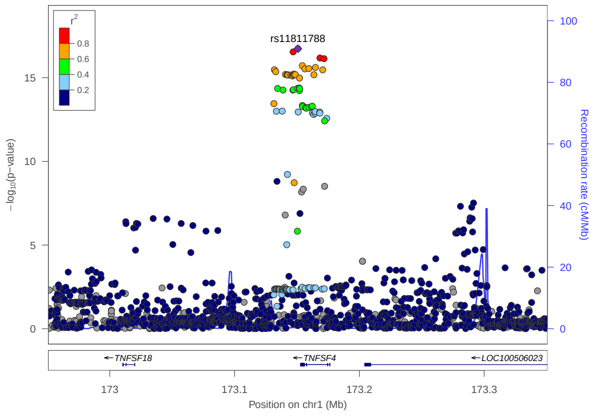

## 2:8442547 - rs891058

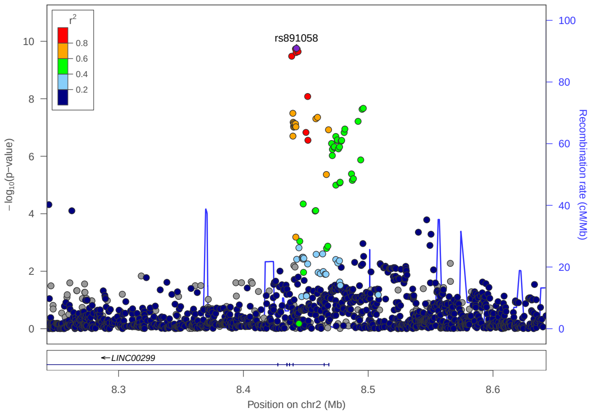


## 2:61163581 - rs67766926

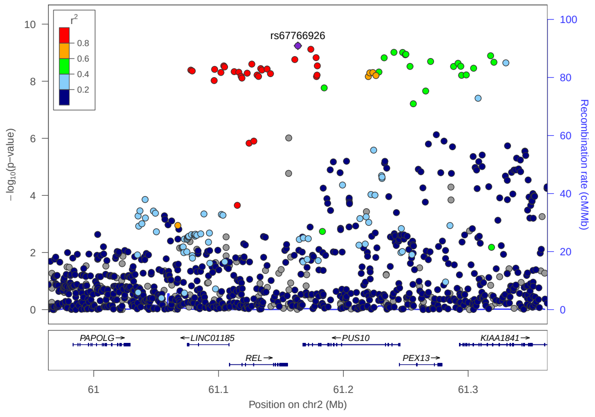


## 2:71100105 - rs112111458

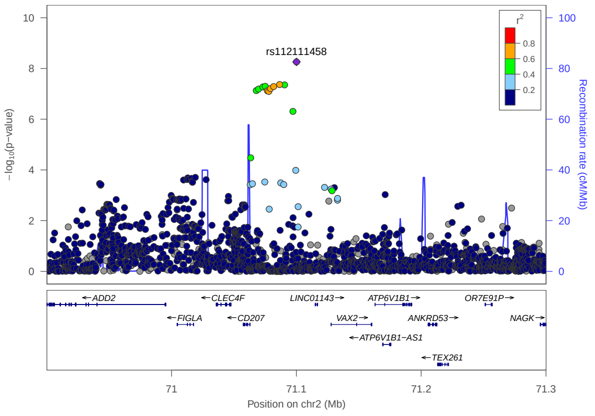


## 2:103039929 - rs2272128

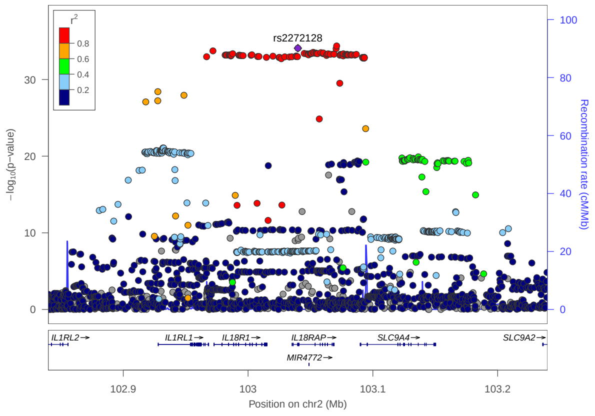


## 2:112275538 - rs112385344

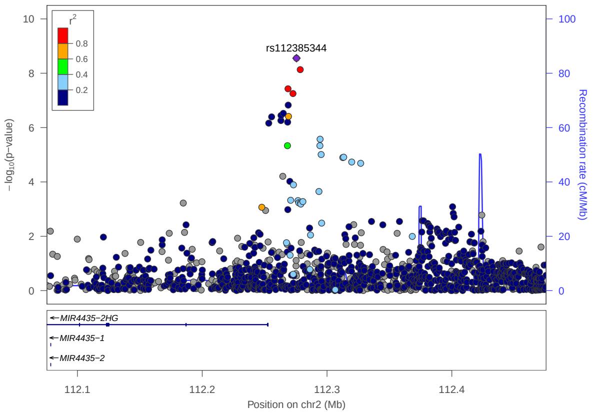


## 2:242788256 - rs62193132

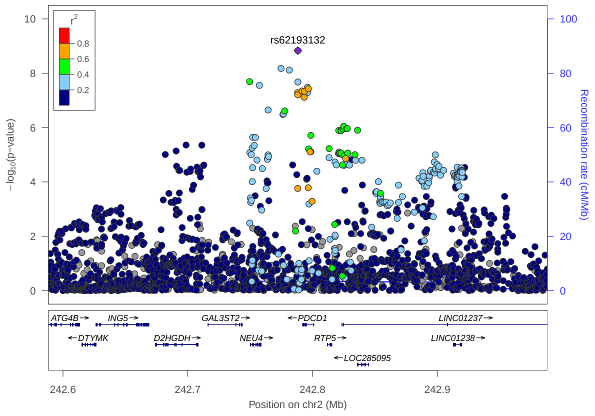

3:18414570 - rs4131280

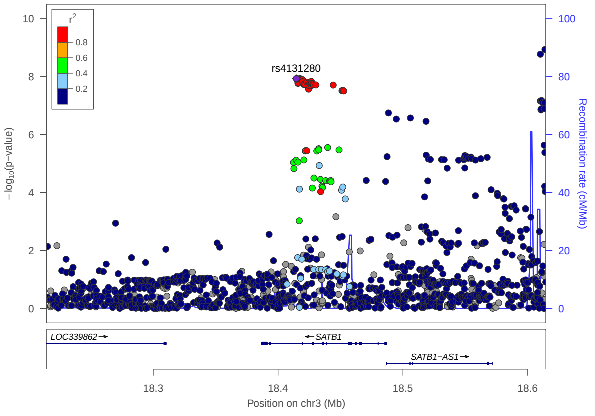


3:18673161 - rs13097010

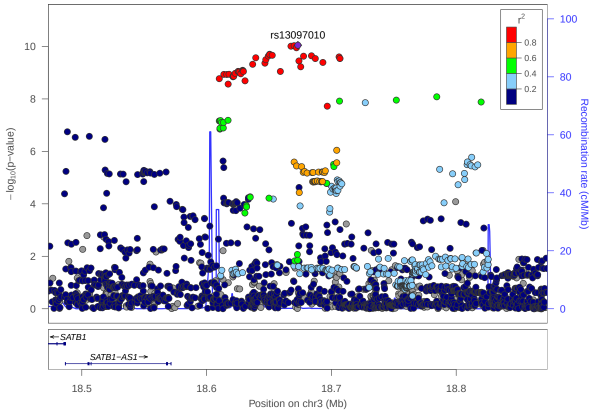


3:33047662 - rs35570272

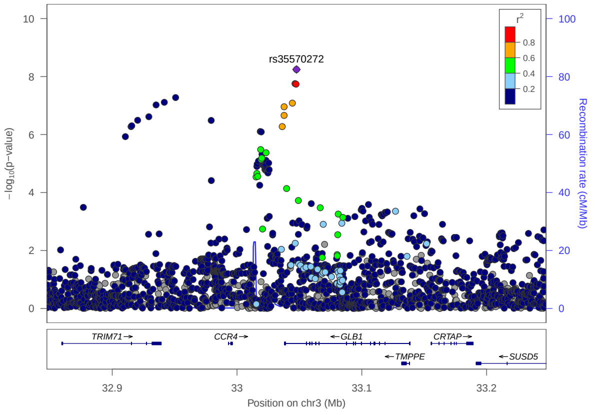


3:112648985 - rs6808249

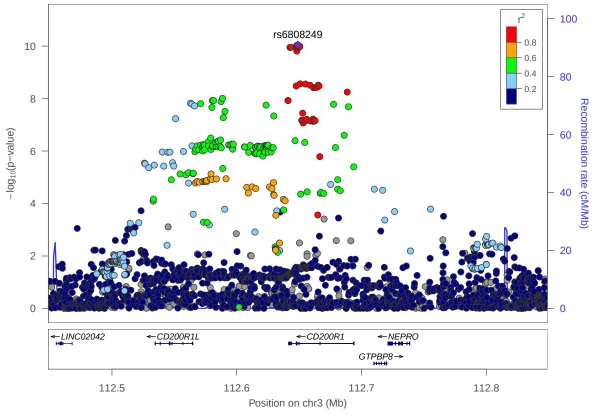


4:123386720 - rs45599938

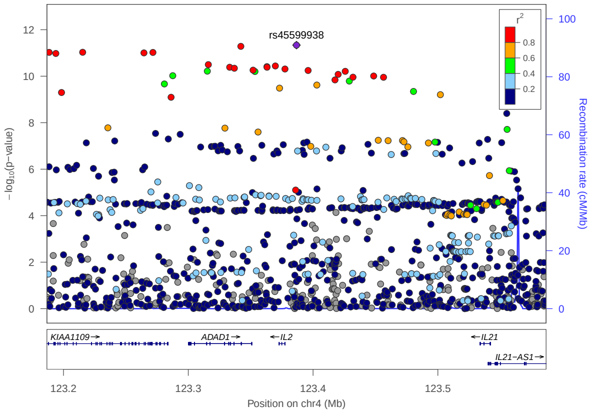


4:142654547 - rs10833

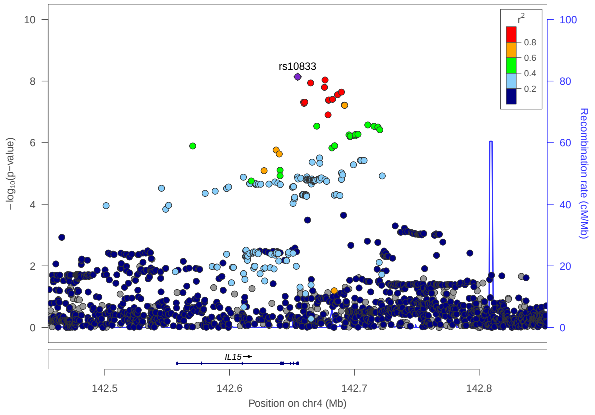

## 5:14604521 - rs148161264

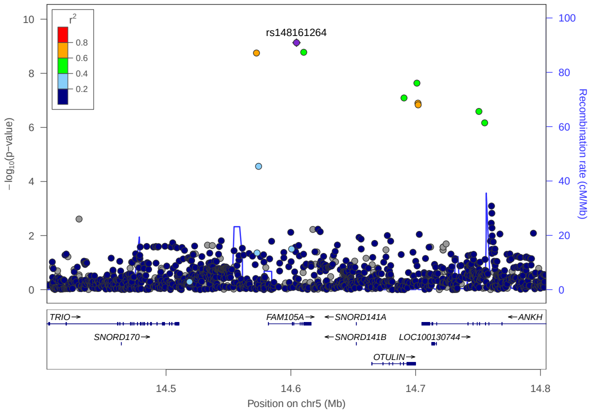


## 5:35883986 - rs10214273

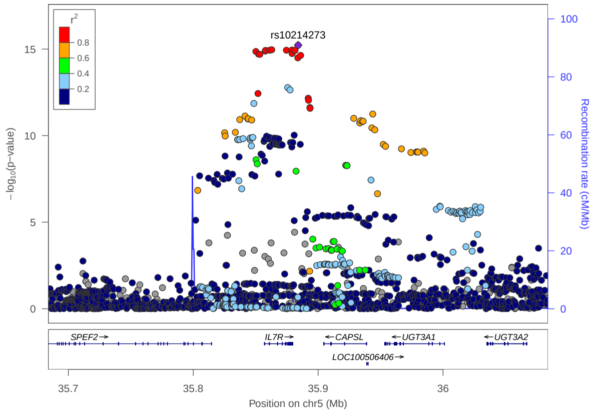


## 5:110331899 - rs17132590

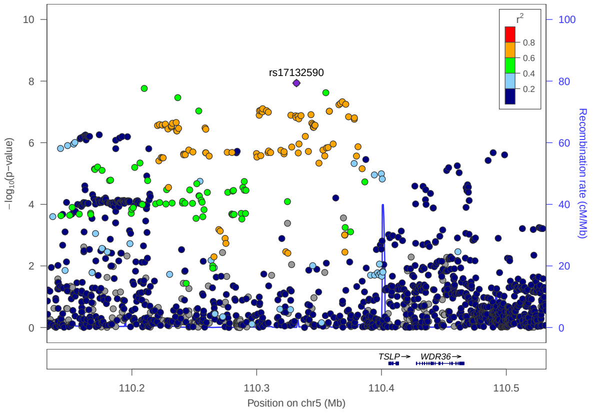


## 5:130059750 - rs7701967

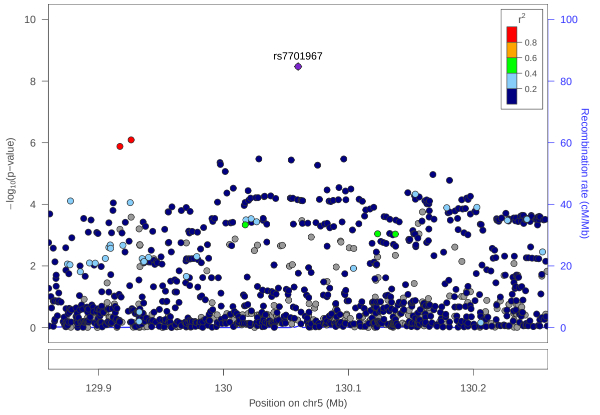


## 5:130674076 - rs4706020

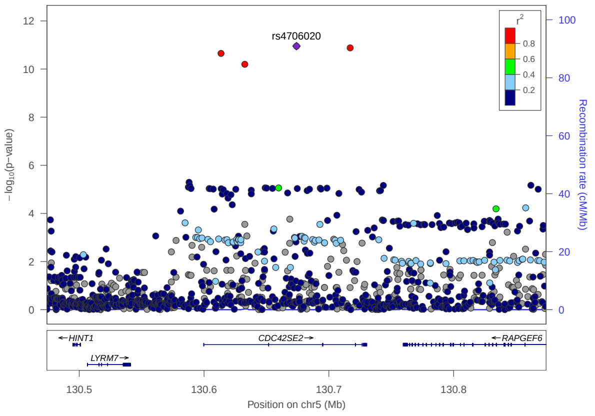


## 5:131347520 - rs4705908

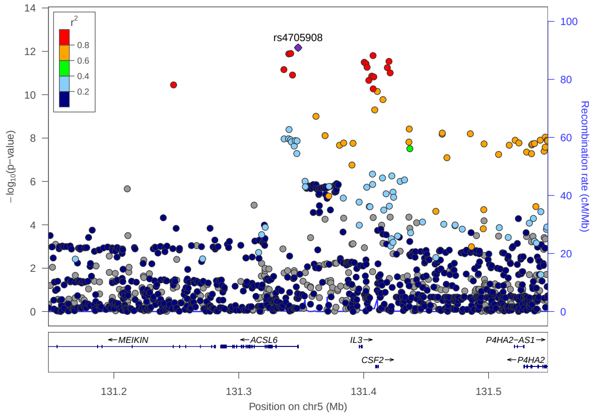

5:131995964 - rs20541

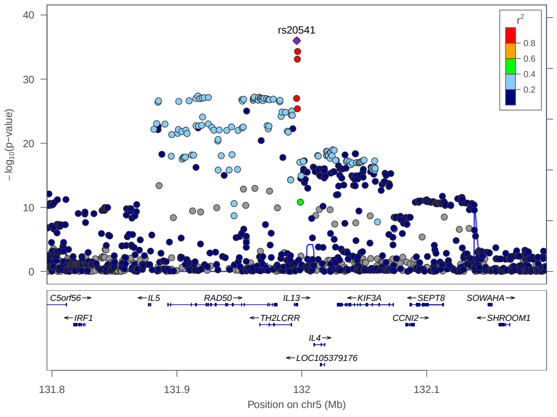


5:172192350 - rs114503346

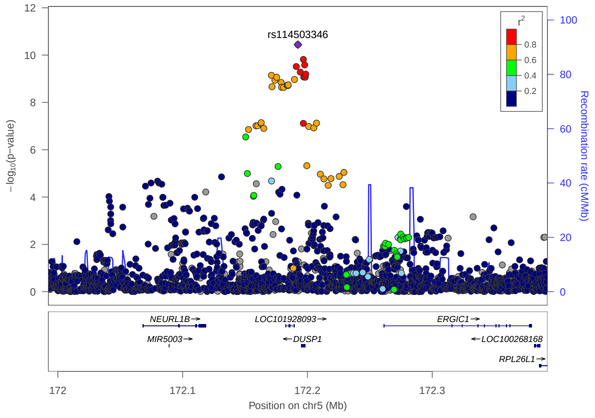


5:176774403 - rs4532376

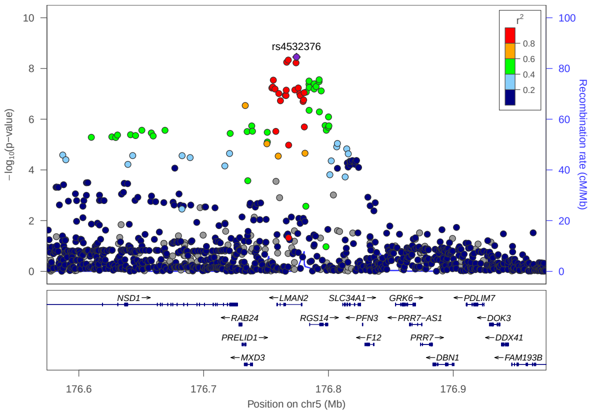


6:31466536 - rs41293876

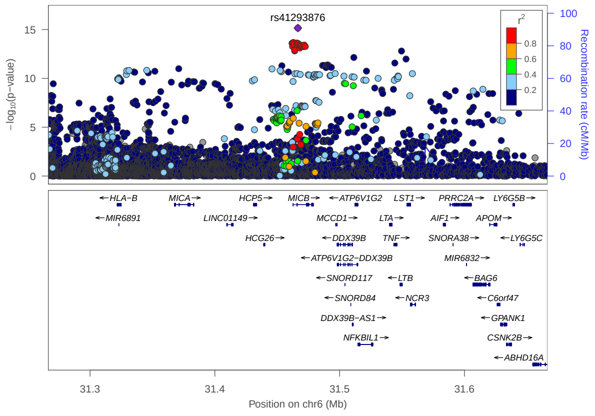


6:32074804 - rs12153855

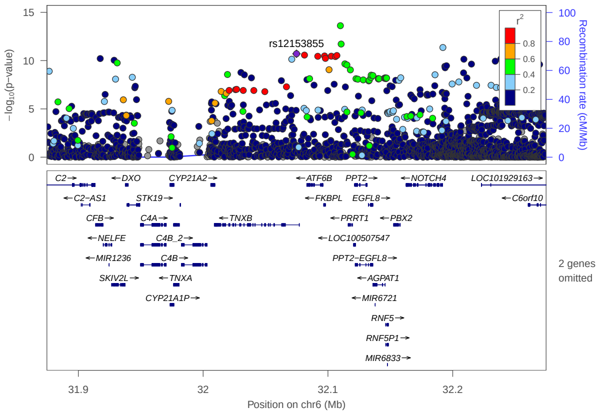


6:32600340 - rs28383330

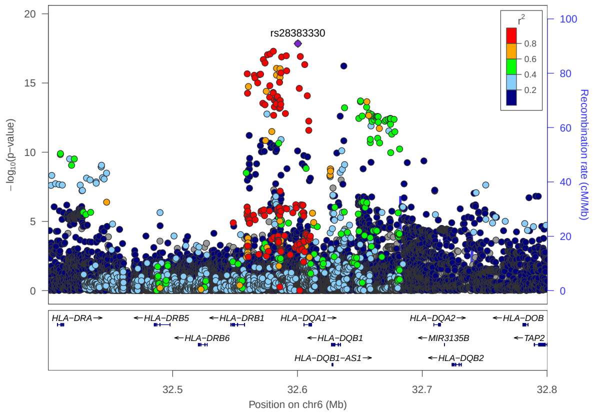

## 6:32658933 - rs9275218

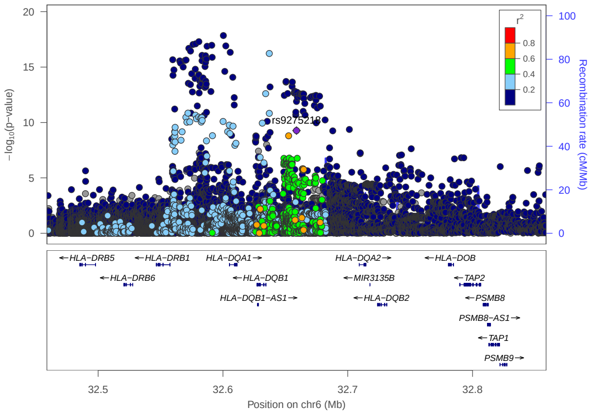


## 6:90930513 - rs72925996

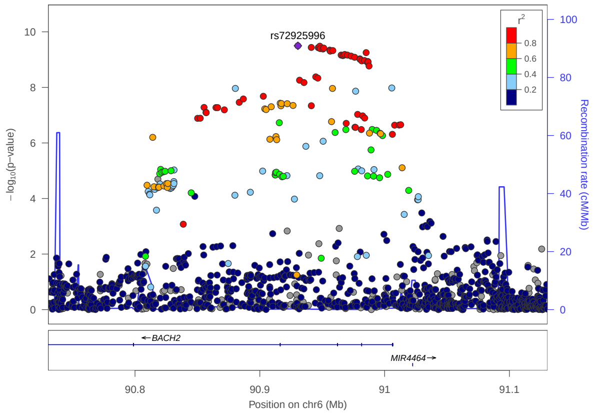


## 6:159496713 - rs629326

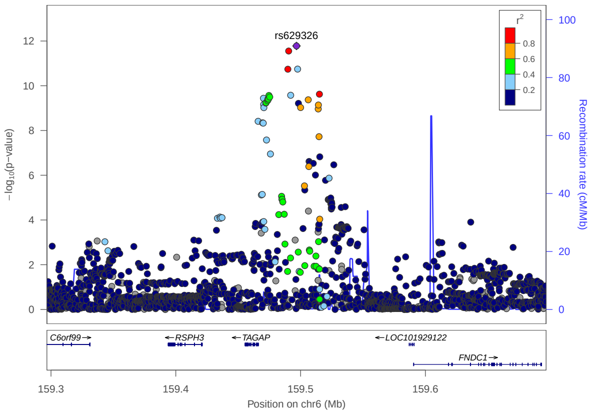


## 7:28830498 - rs989437

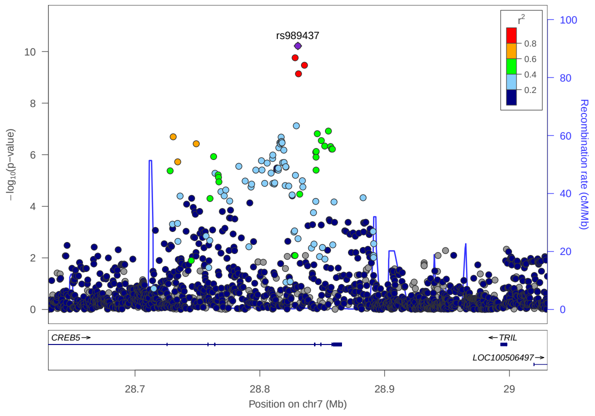


## 8:21767240 - rs34215892

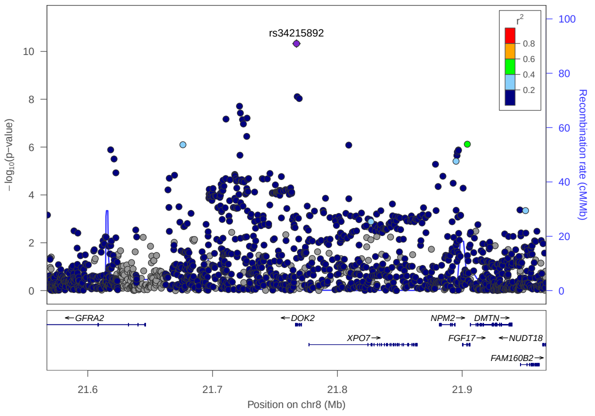


## 8:21767809 - rs118162691

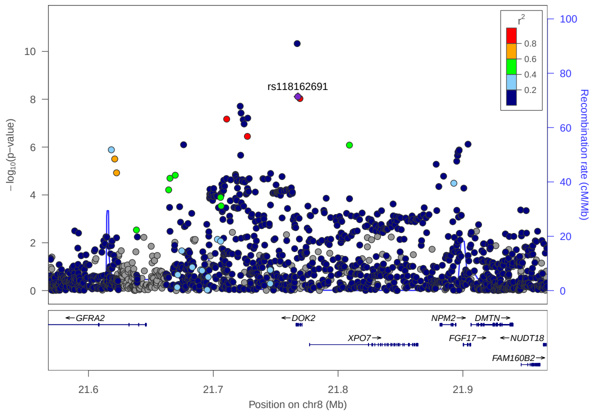

## 8:81288734 - rs952558

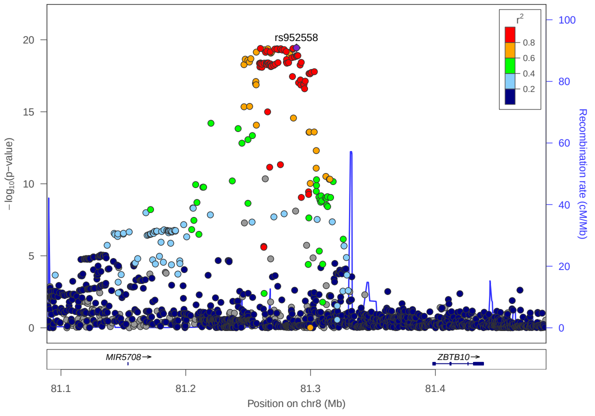


## 8:126609868 - rs6996614

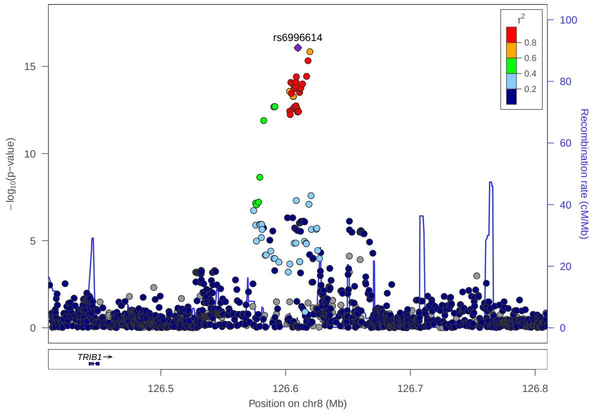


## 8:141601542 - rs7843258

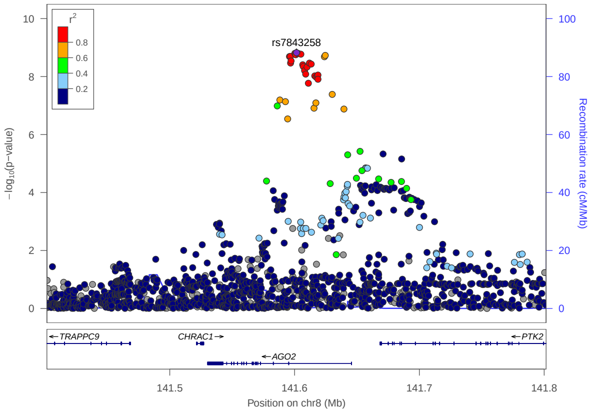


## 9:33430707 - rs7857407

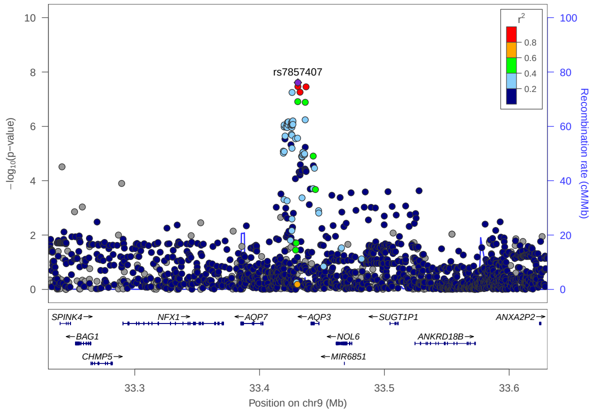


## 9:102331281 - rs10988863

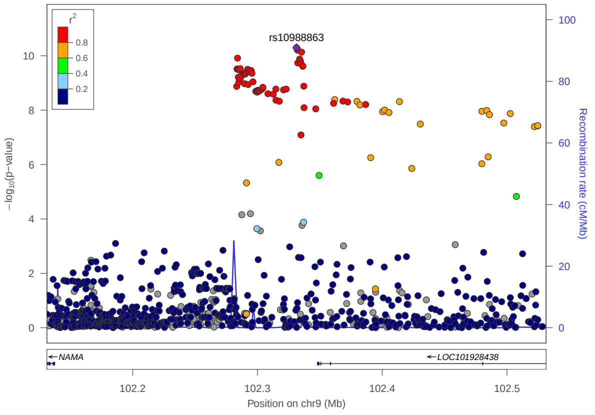


## 10:6123495 - rs12251307

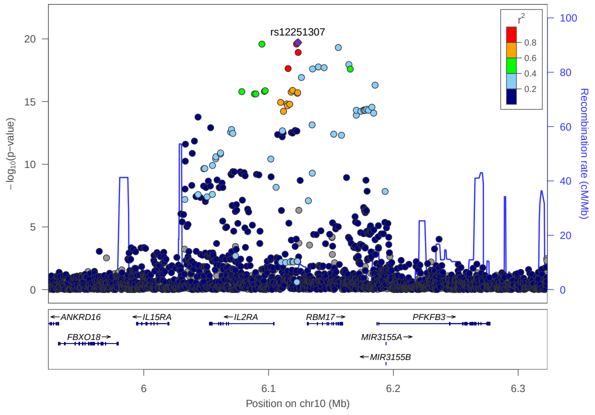

## 10:6627700 - rs10796303

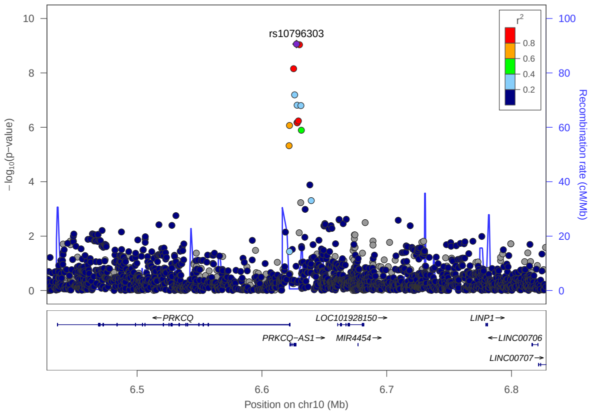


## 10:64376558 - rs10822037

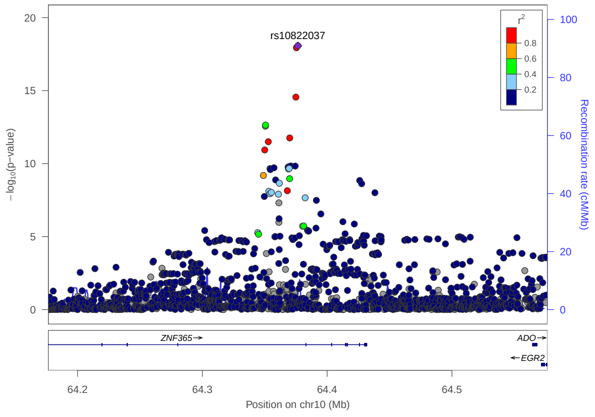


## 11:36365253 - rs10836538

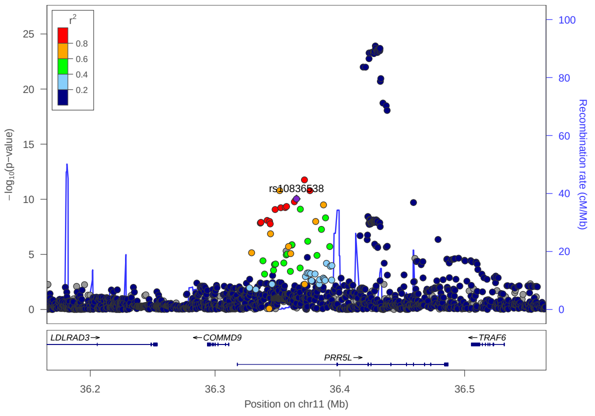


## 11:36428447 - rs28520436

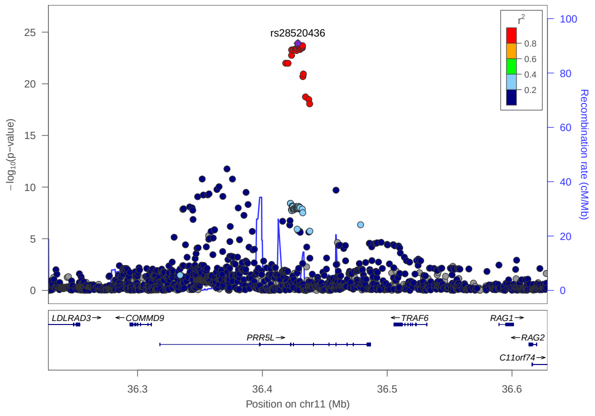


## 11:65559266 - rs10791824

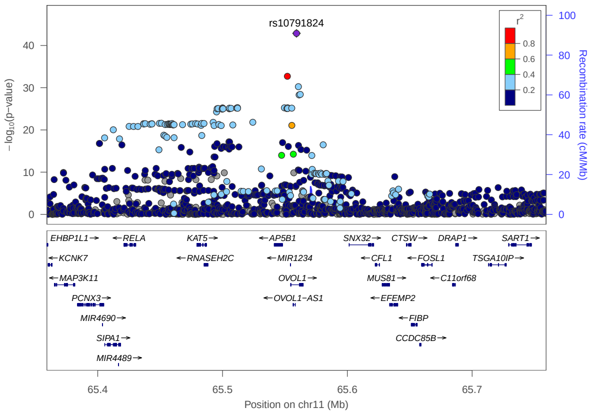


## 11:76293758 - rs7936323

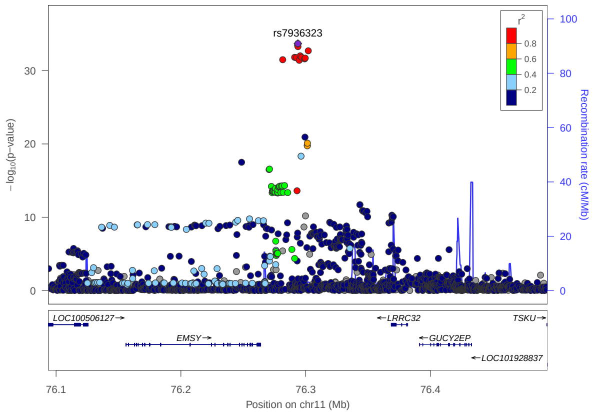

## 11:76343427 - rs11236813

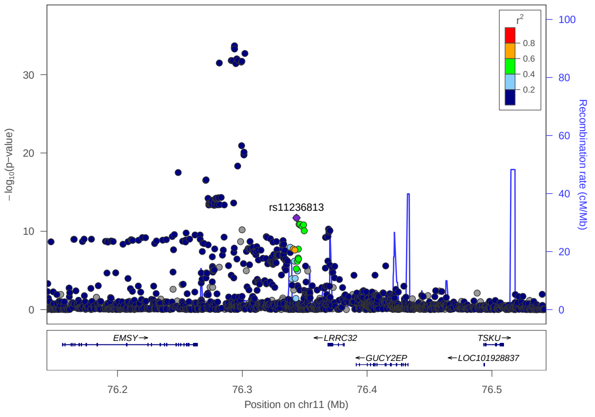


## 11:102748695 - rs17368814

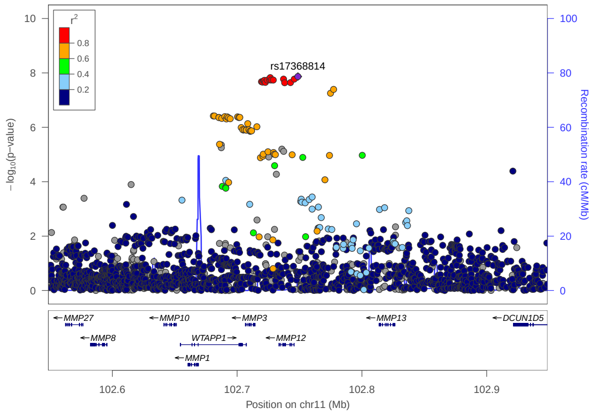


## 11:116843425 - rs11216206

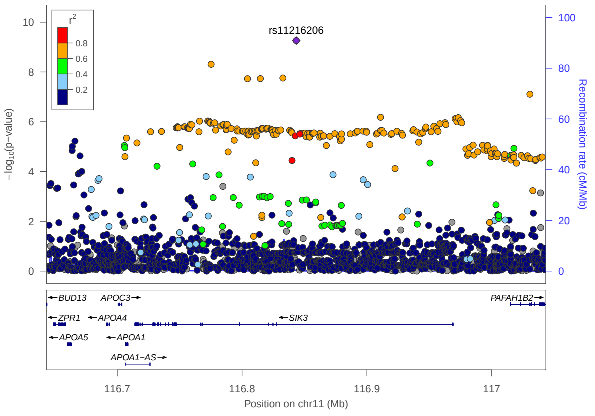


## 11:118745884 - rs10790275

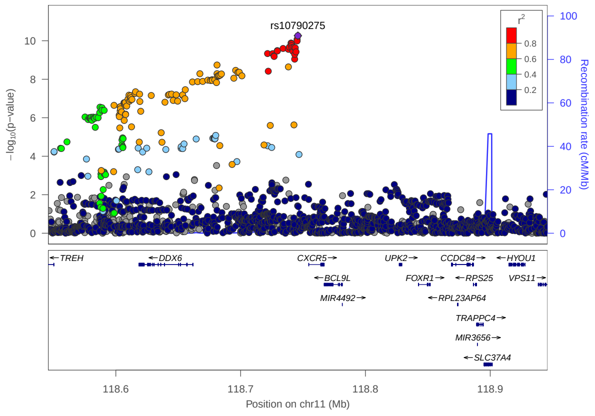


## 11:128187383 - rs7127307

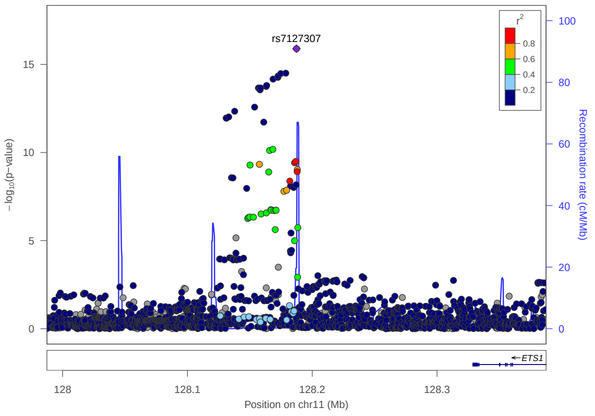


## 12:56384804 - rs705699

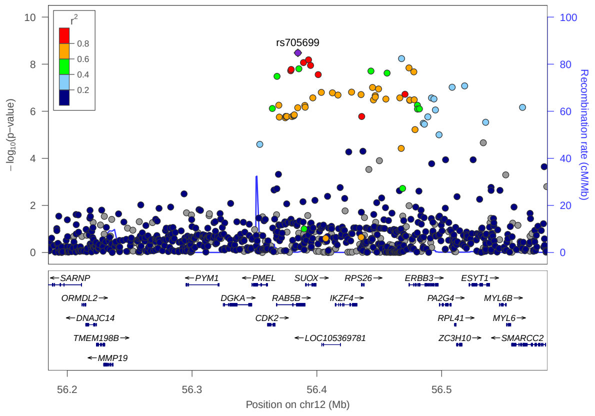

## 12:68646521 - rs2227491

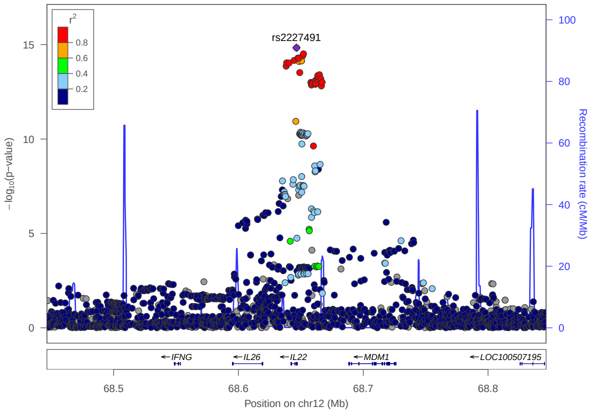


## 12:94611908 - rs5005507

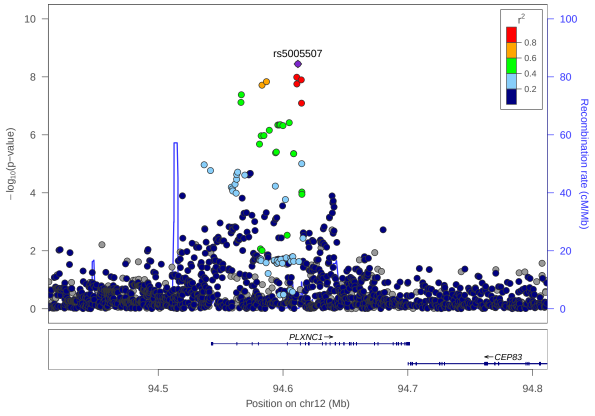


## 14:35638937 - rs2415269

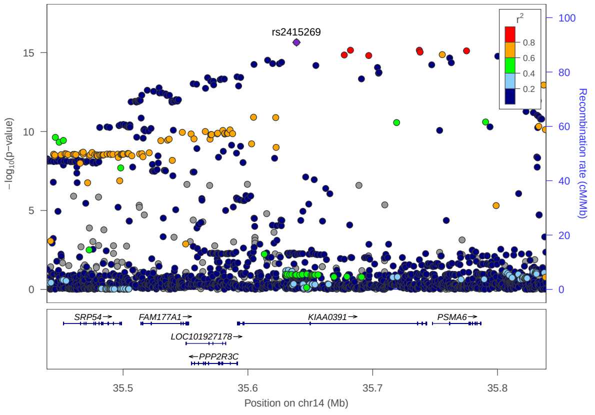


## 14:103249127 - rs4906263

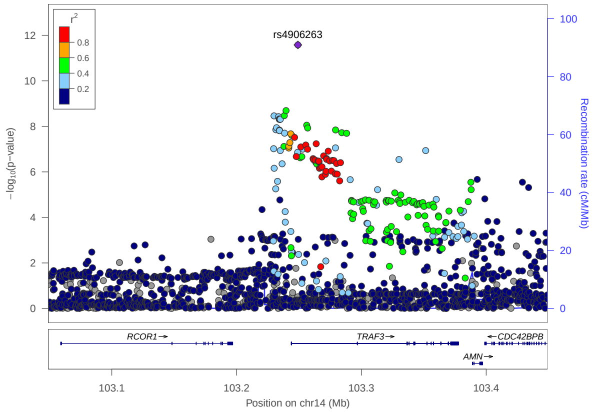


## 14:105523663 - rs7147439

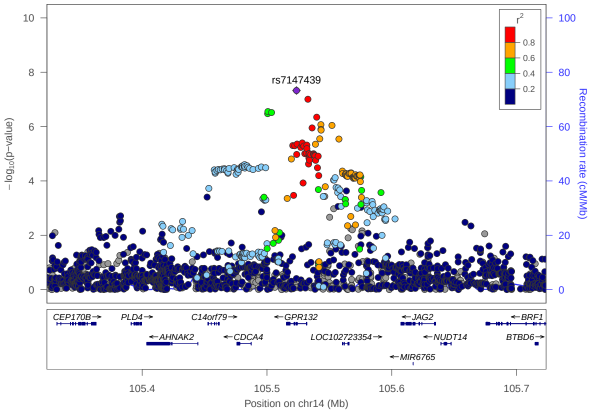


## 16:11229589 - rs2041733

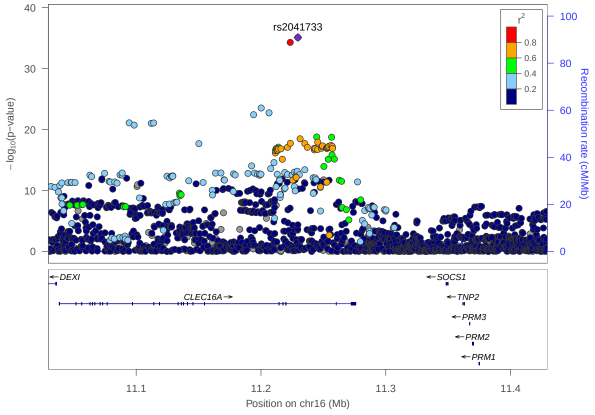

## 17:38757789 - rs1358175

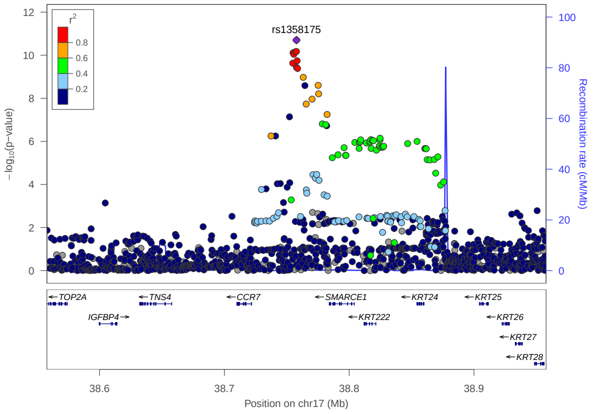


## 17:40485239 - rs17881320

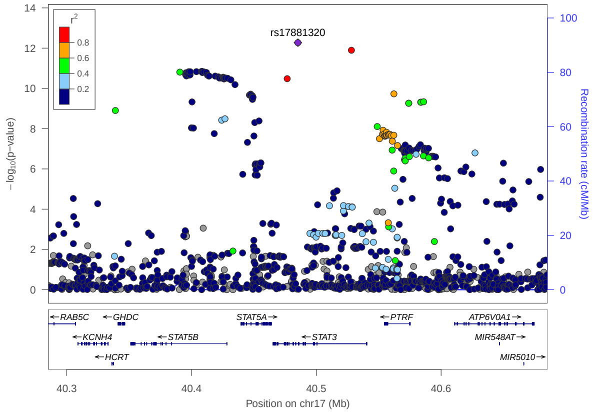


## 17:43336687 - rs4247364

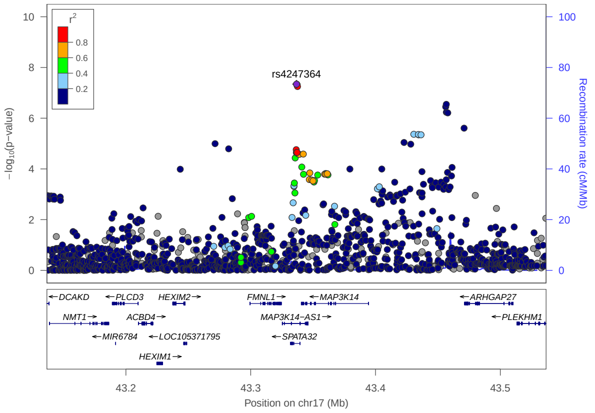


## 17:45819206 - rs56308324

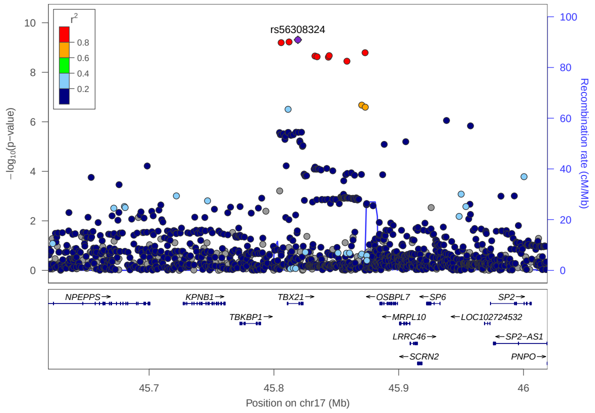


## 17:47454507 - rs28406364

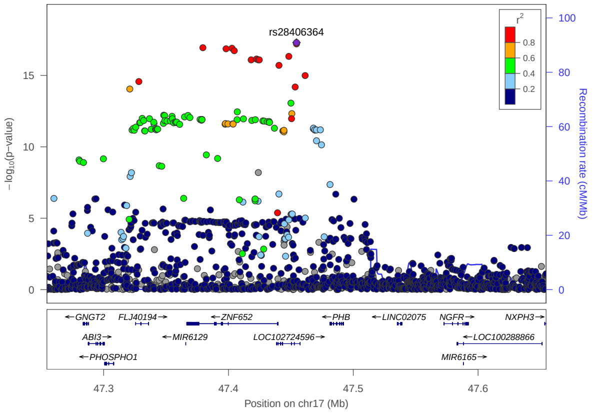


## 18:12775851 - rs2542147

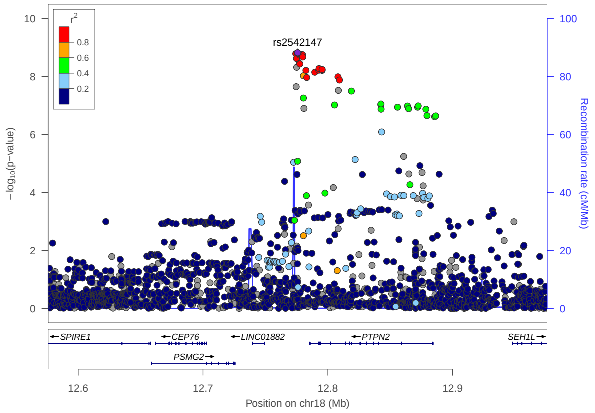

19:8789721 - rs2967677

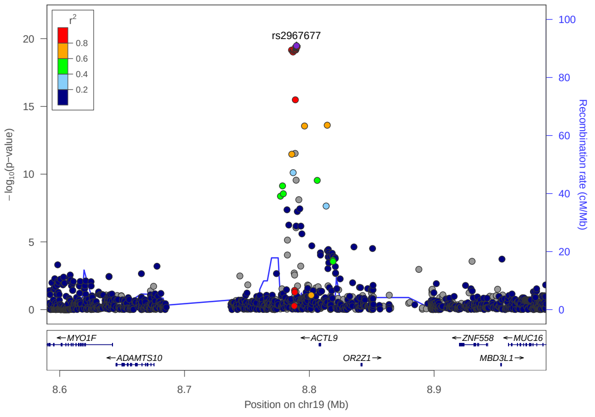


20:62302539 - rs6062486

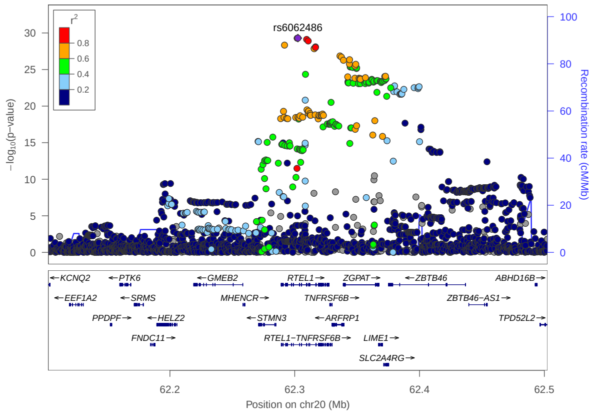


22:37316873 - rs4821569

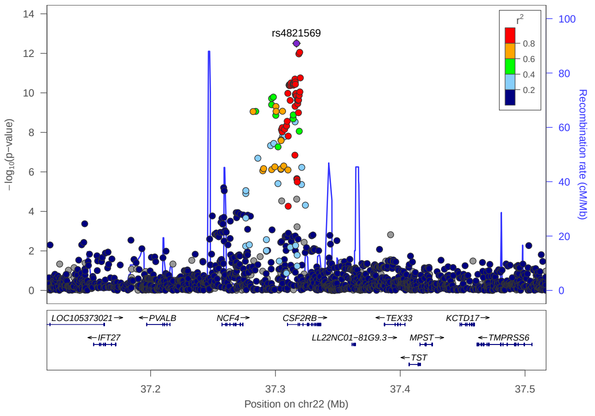

### Additional SNPs from Multi-ancestry analysis

## 1:19804918 - rs114059822

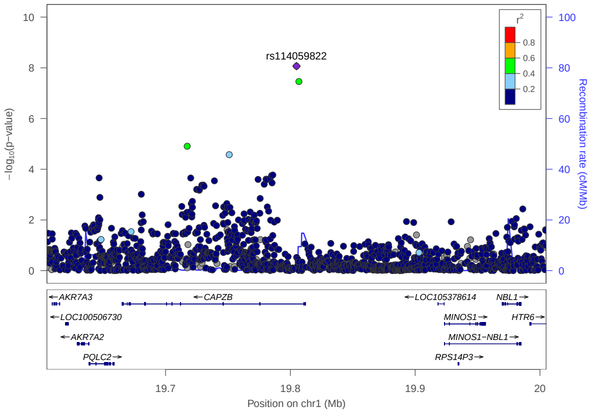


## 2:112294446 - rs77869365

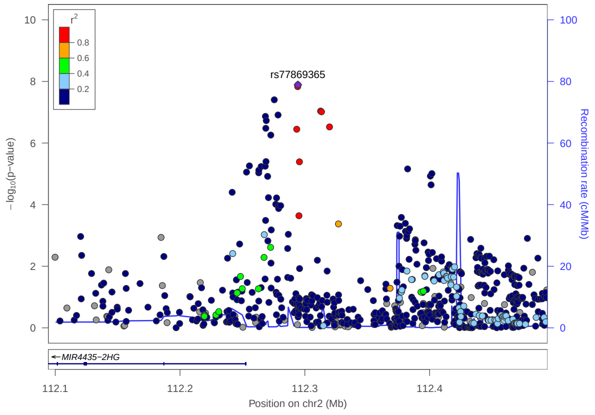


## 2:234113301 - rs9247

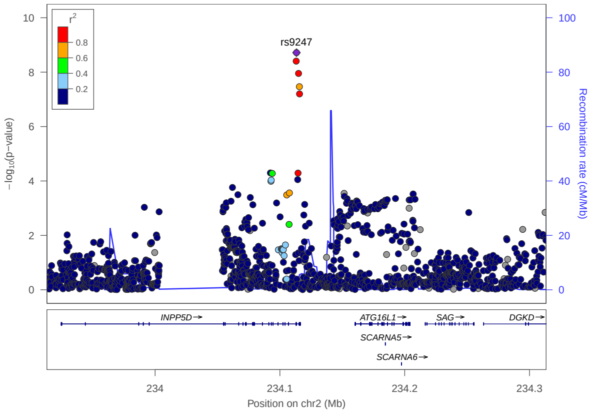


## 3:112383847 - rs9864845

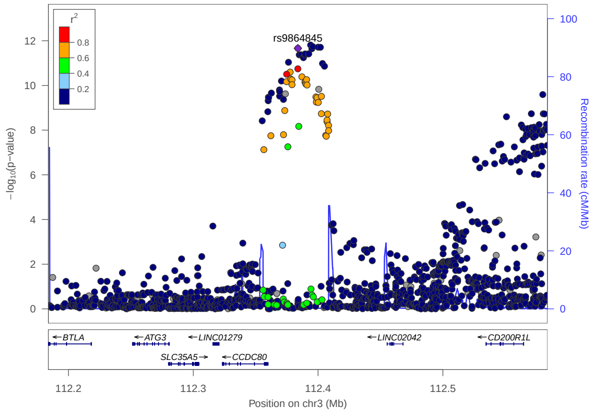


## 6:31168397 - rs9263868

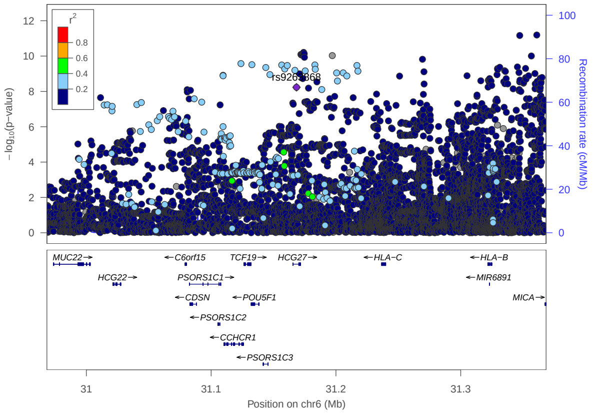


## 6:32560306 - rs34665982

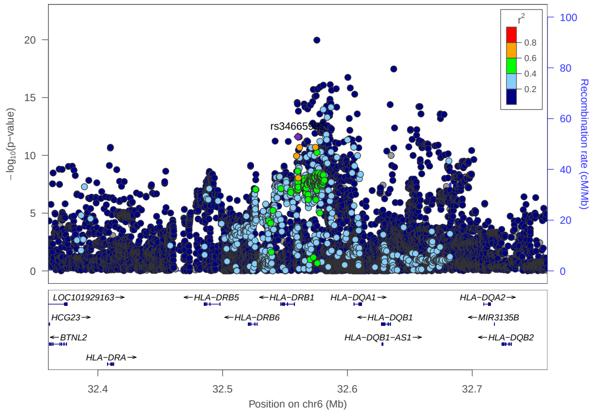

## 6:106629690 - rs34599047

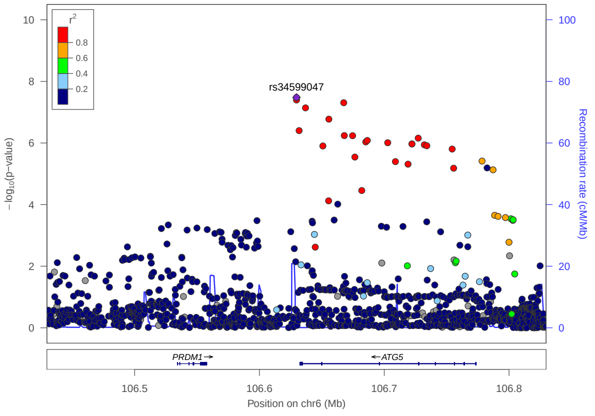


## 6:135707486 - rs7773987

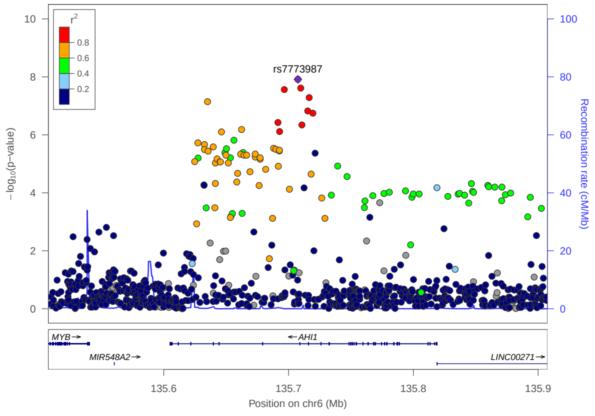


## 9:1894613 - rs118029610

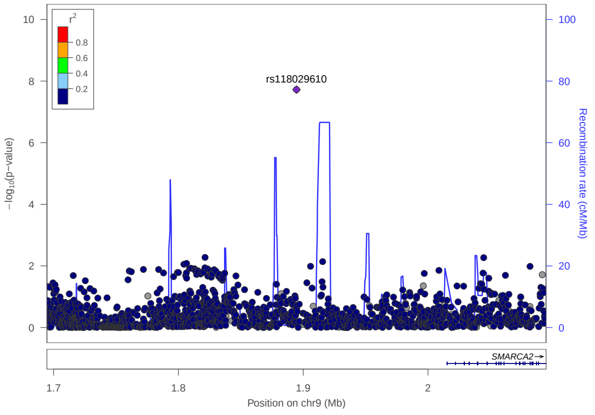


## 9:140500443 - rs117137535

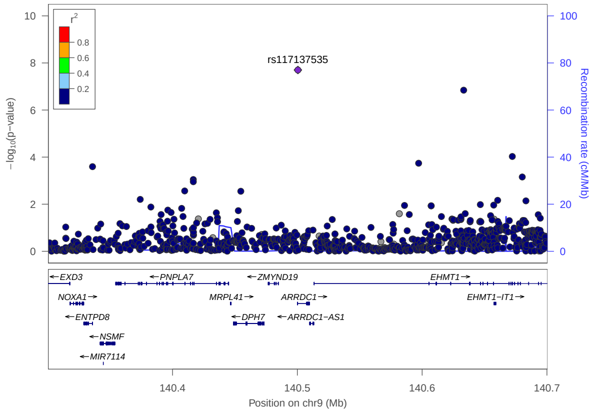


## 10:64573312 - rs45602133

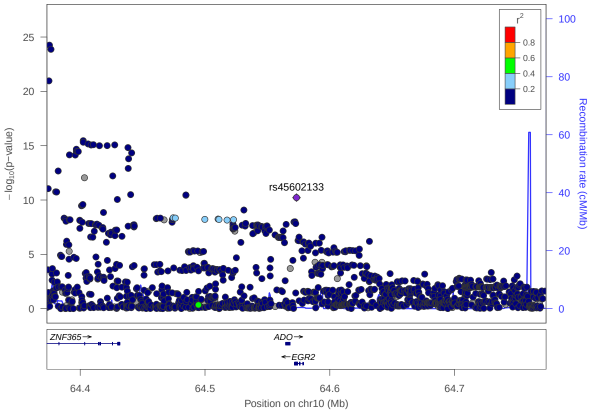


## 11:7977161 - rs4312054

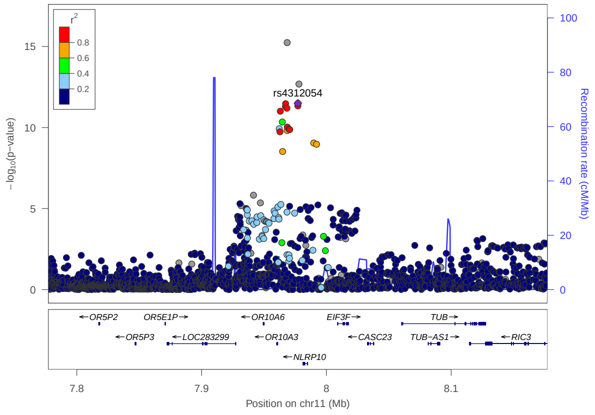

## 11:83439186 - rs150113720

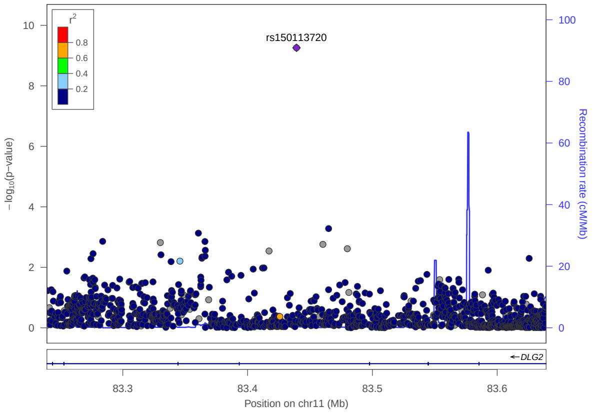


## 11:101361300 - rs115148078

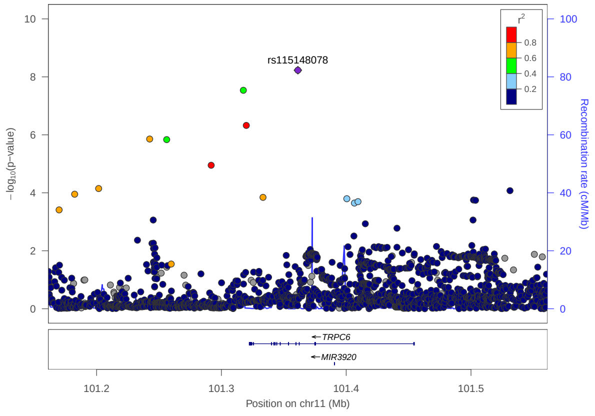


## 11:116827369 - rs17120177

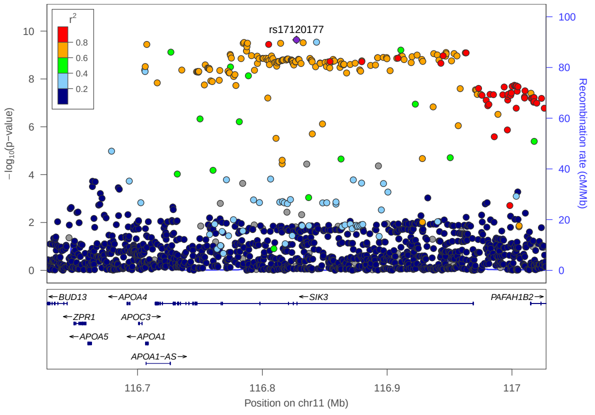


## 11:128421175 - rs4262739

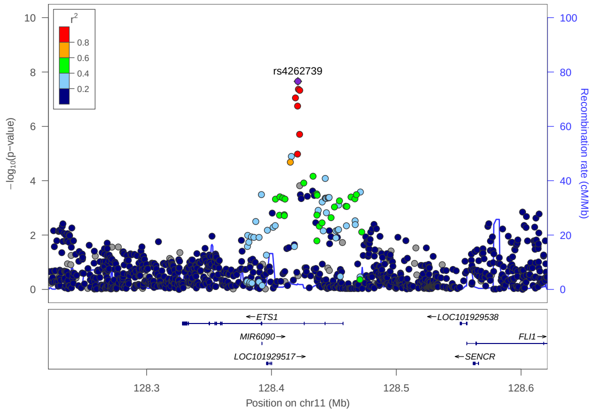


## 12:57489709 - rs1059513

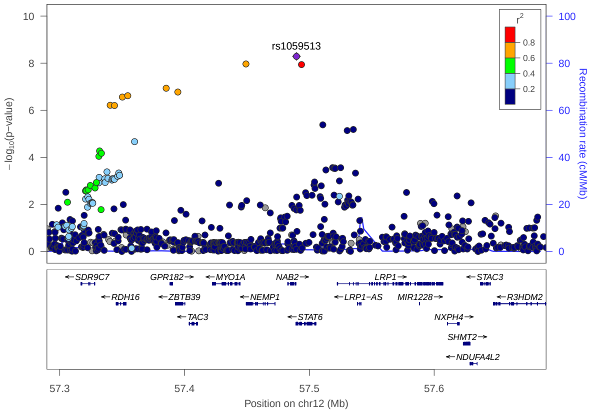


## 18:60009814 - rs4574025

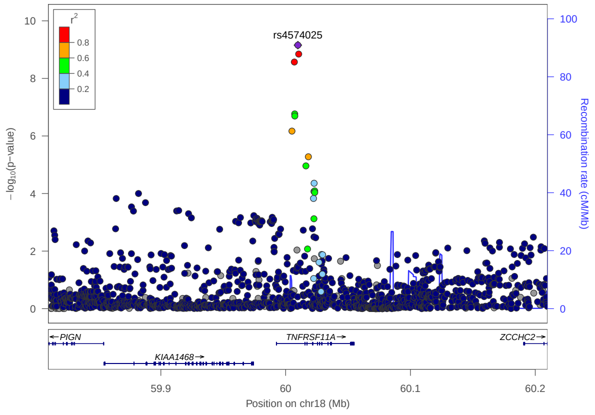

# 20:52797237 - rs6023002

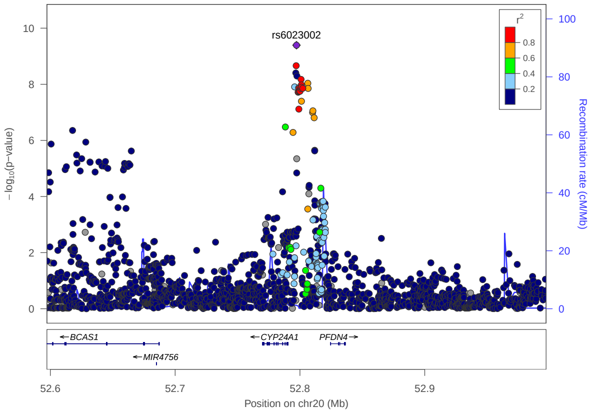
