## Supplementary Figure 2 for "European and multi-ancestry genome-wide association meta-analysis of atopic dermatitis highlights importance of systemic immune regulation"

### Discovered in European ancestry analysis

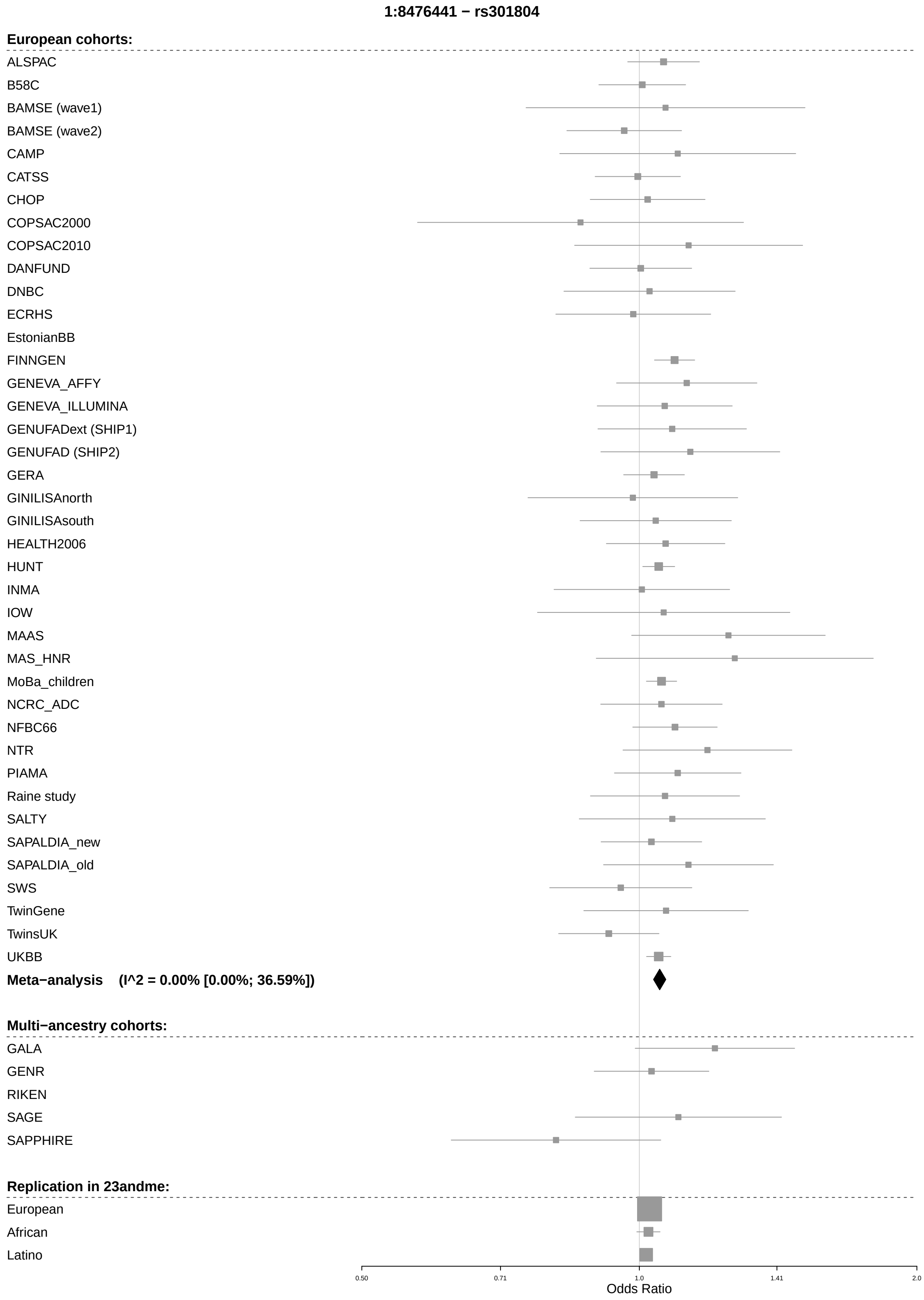

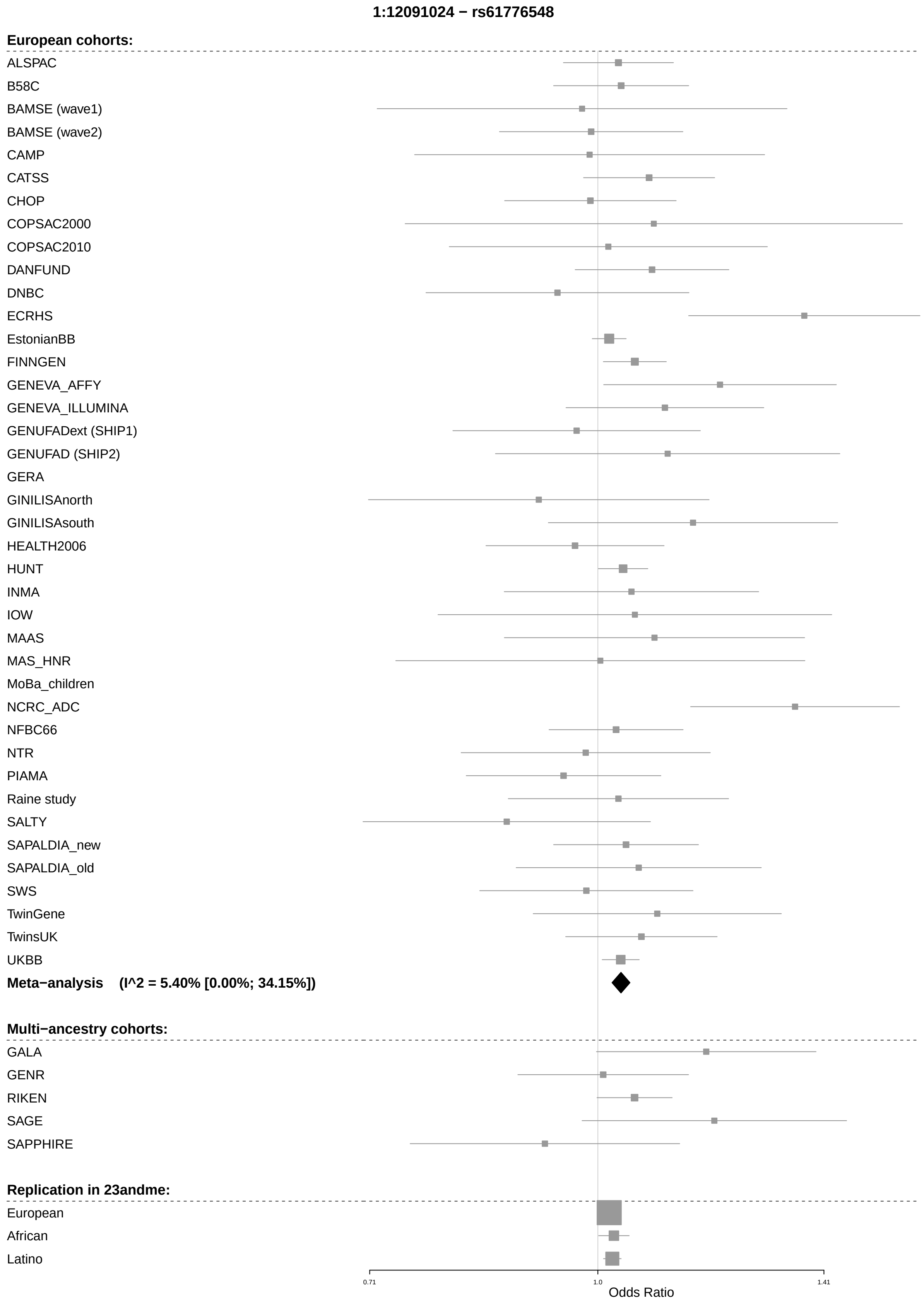

1:12091024 − rs61776548
European cohorts:
ALSPAC
B58C
BAMSE (wave1)
BAMSE (wave2)
CAMP
CATSS
CHOP
COPSAC2000
COPSAC2010
DANFUND
DNBC
ECRHS
EstonianBB
FINNGEN
GENEVA_AFFY
GENEVA_ILLUMINA
GENUFADext (SHIP1)
GENUFAD (SHIP2)
GERA
GINILISAnorth
GINILISAsouth
HEALTH2006
HUNT
INMA
IOW
MAAS
MAS_HNR
MoBa_children
NCRC_ADC
NFBC66
NTR
PIAMA
Raine study
SALTY
SAPALDIA_new
SAPALDIA_old
SWS
TwinGene
TwinsUK
UKBB
Meta−analysis (I^2 = 5.40% [0.00%; 34.15%])
Multi−ancestry cohorts:
GALA
GENR
RIKEN
SAGE
SAPPHIRE
Replication in 23andme:
European
African
Latino
0.71
1.0
1.41
Odds Ratio

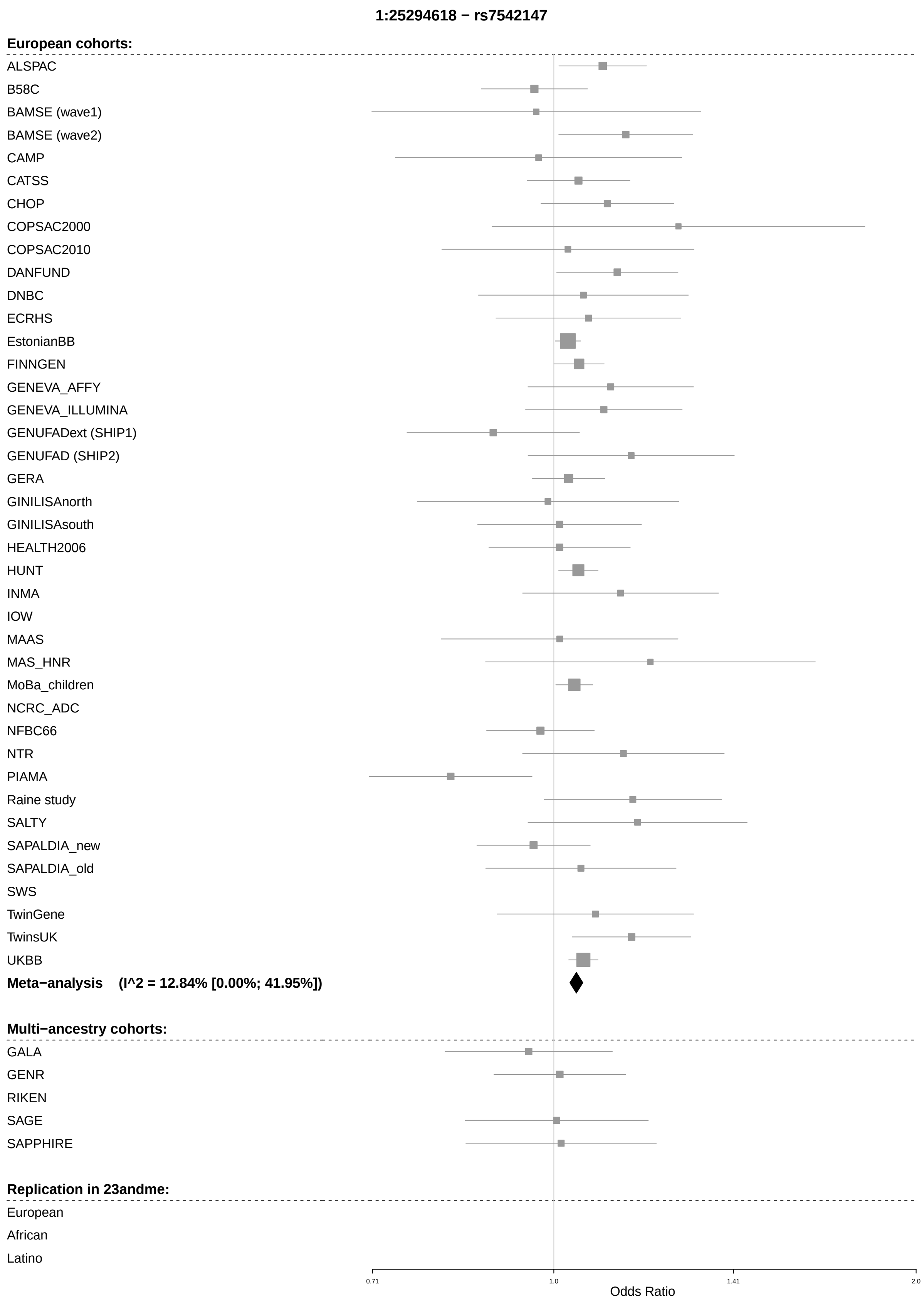
1:25294618 − rs7542147
European cohorts:
ALSPAC
B58C
BAMSE (wave1)
BAMSE (wave2)
CAMP
CATSS
CHOP
COPSAC2000
COPSAC2010
DANFUND
DNBC
ECRHS
EstonianBB
FINNGEN
GENEVA_AFFY
GENEVA_ILLUMINA
GENUFADext (SHIP1)
GENUFAD (SHIP2)
GERA
GINILISAnorth
GINILISAsouth
HEALTH2006
HUNT
INMA
IOW
MAAS
MAS_HNR
MoBa_children
NCRC_ADC
NFBC66
NTR
PIAMA
Raine study
SALTY
SAPALDIA_new
SAPALDIA_old
SWS
TwinGene
TwinsUK
UKBB
Meta−analysis (I^2 = 12.84% [0.00%; 41.95%])
Multi−ancestry cohorts:
GALA
GENR
RIKEN
SAGE
SAPPHIRE
Replication in 23andme:
European
African
Latino
0.71
1.0
1.41
2.0
Odds Ratio

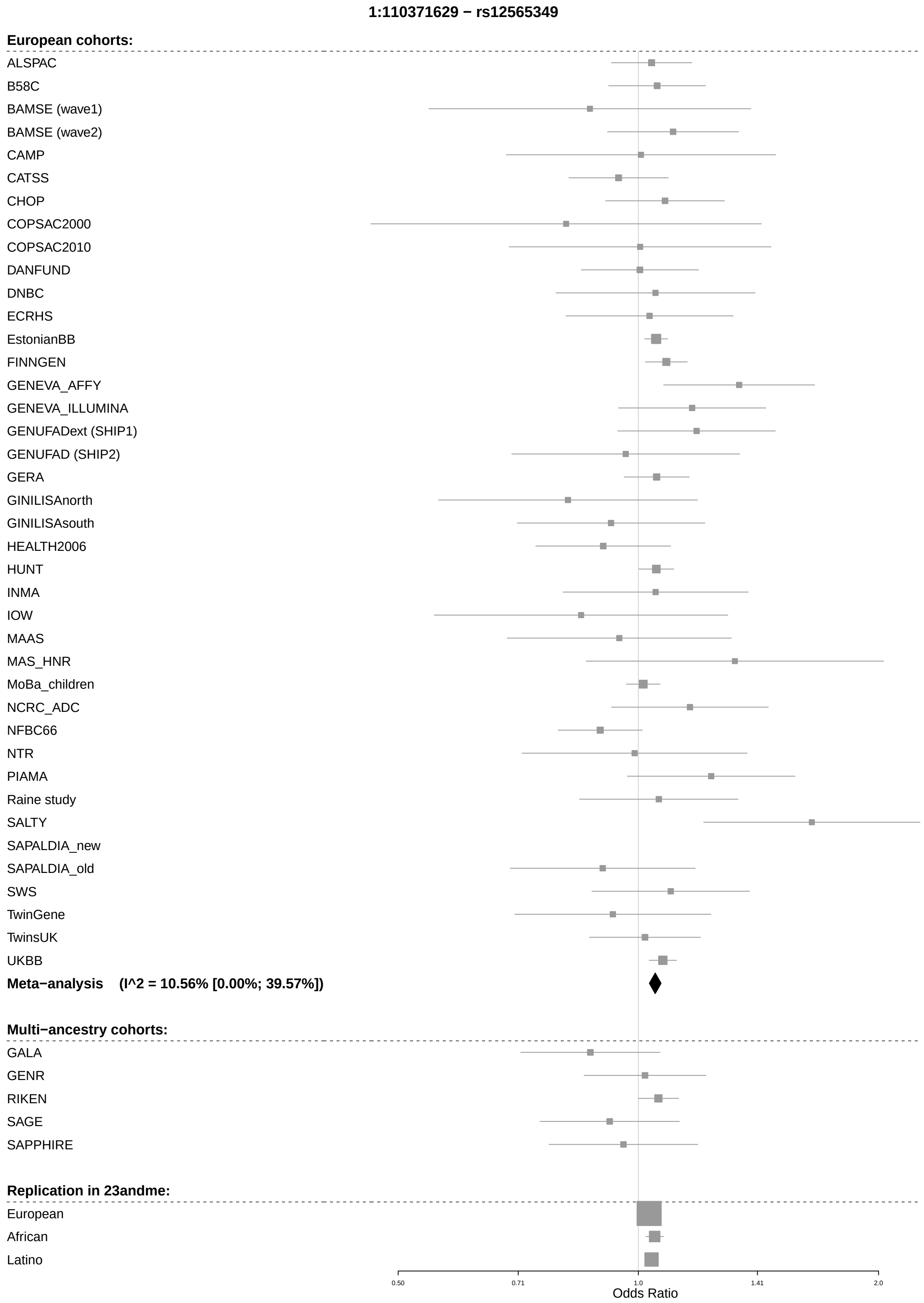

1:110371629 − rs12565349
European cohorts:
ALSPAC
B58C
BAMSE (wave1)
BAMSE (wave2)
CAMP
CATSS
CHOP
COPSAC2000
COPSAC2010
DANFUND
DNBC
ECRHS
EstonianBB
FINNGEN
GENEVA_AFFY
GENEVA_ILLUMINA
GENUFADext (SHIP1)
GENUFAD (SHIP2)
GERA
GINILISAnorth
GINILISAsouth
HEALTH2006
HUNT
INMA
IOW
MAAS
MAS_HNR
MoBa_children
NCRC_ADC
NFBC66
NTR
PIAMA
Raine study
SALTY
SAPALDIA_new
SAPALDIA_old
SWS
TwinGene
TwinsUK
UKBB
Meta−analysis (I^2 = 10.56% [0.00%; 39.57%])
Multi−ancestry cohorts:
GALA
GENR
RIKEN
SAGE
SAPPHIRE
Replication in 23andme:
European
African
Latino
0.50
0.71
1.0
1.41
2.0
Odds Ratio

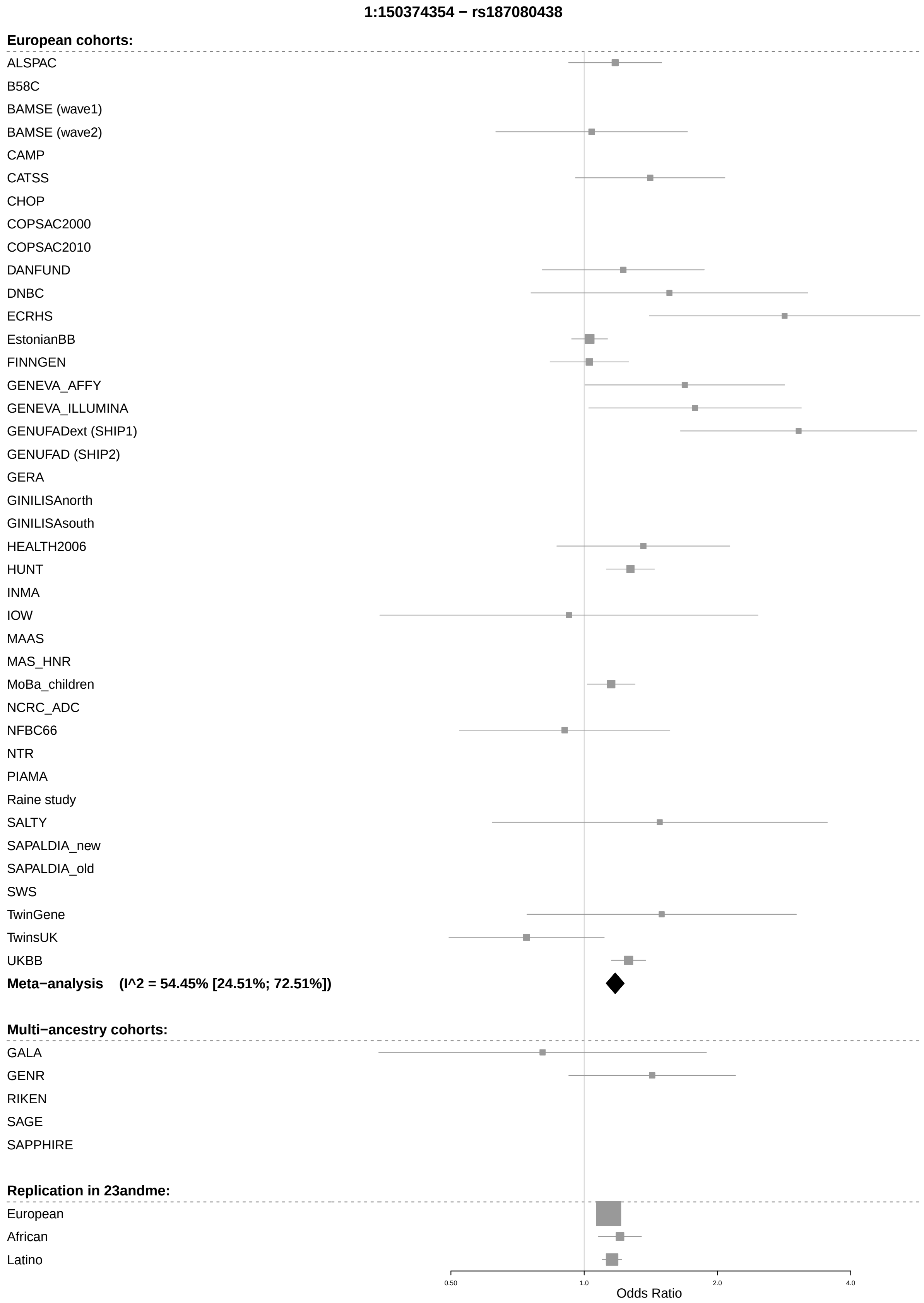
1:150374354 − rs187080438
European cohorts:
ALSPAC
B58C
BAMSE (wave1)
BAMSE (wave2)
CAMP
CATSS
CHOP
COPSAC2000
COPSAC2010
DANFUND
DNBC
ECRHS
EstonianBB
FINNGEN
GENEVA_AFFY
GENEVA_ILLUMINA
GENUFADext (SHIP1)
GENUFAD (SHIP2)
GERA
GINILISAnorth
GINILISAsouth
HEALTH2006
HUNT
INMA
IOW
MAAS
MAS_HNR
MoBa_children
NCRC_ADC
NFBC66
NTR
PIAMA
Raine study
SALTY
SAPALDIA_new
SAPALDIA_old
SWS
TwinGene
TwinsUK
UKBB
Meta−analysis (I^2 = 54.45% [24.51%; 72.51%])
Multi−ancestry cohorts:
GALA
GENR
RIKEN
SAGE
SAPPHIRE
Replication in 23andme:
European
African
Latino
0.50
1.0
2.0
4.0
Odds Ratio

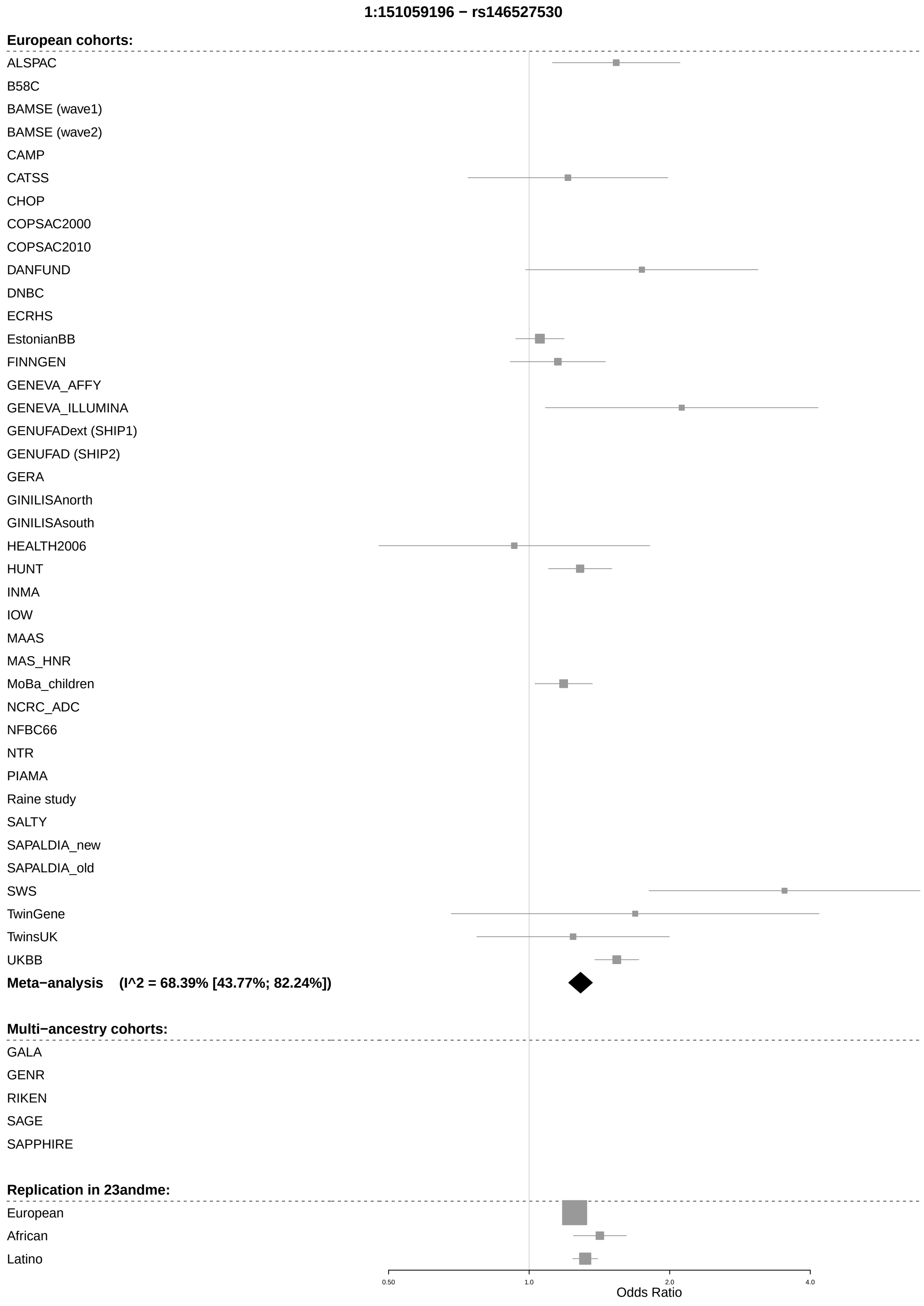
1:151059196 − rs146527530
European cohorts:
ALSPAC
B58C
BAMSE (wave1)
BAMSE (wave2)
CAMP
CATSS
CHOP
COPSAC2000
COPSAC2010
DANFUND
DNBC
ECRHS
EstonianBB
FINNGEN
GENEVA_AFFY
GENEVA_ILLUMINA
GENUFADext (SHIP1)
GENUFAD (SHIP2)
GERA
GINILISAnorth
GINILISAsouth
HEALTH2006
HUNT
INMA
IOW
MAAS
MAS_HNR
MoBa_children
NCRC_ADC
NFBC66
NTR
PIAMA
Raine study
SALTY
SAPALDIA_new
SAPALDIA_old
SWS
TwinGene
TwinsUK
UKBB
Meta−analysis (I^2 = 68.39% [43.77%; 82.24%])
Multi−ancestry cohorts:
GALA
GENR
RIKEN
SAGE
SAPPHIRE
Replication in 23andme:
European
African
Latino
0.50
1.0
2.0
4.0
Odds Ratio

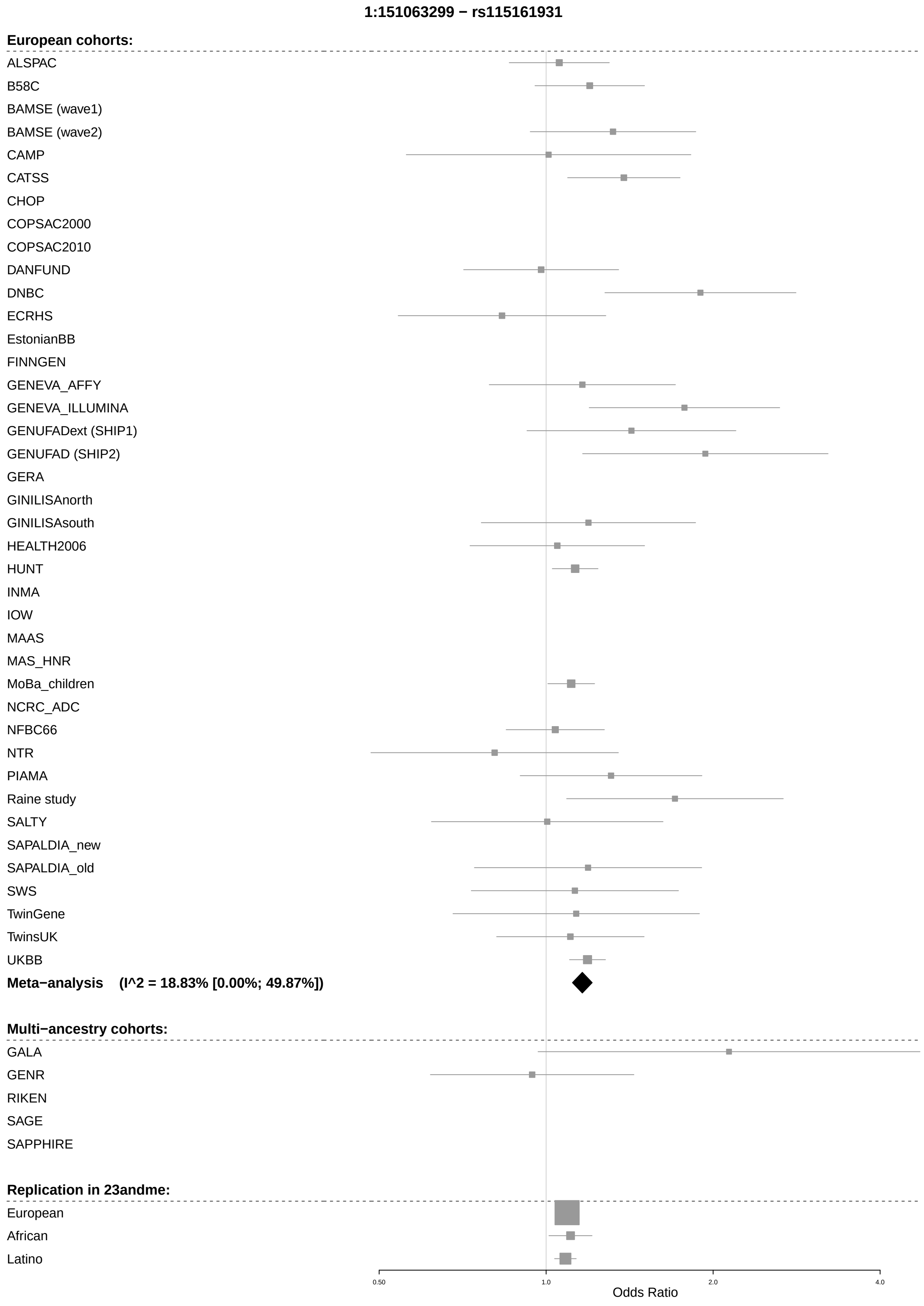
1:151063299 − rs115161931
European cohorts:
ALSPAC
B58C
BAMSE (wave1)
BAMSE (wave2)
CAMP
CATSS
CHOP
COPSAC2000
COPSAC2010
DANFUND
DNBC
ECRHS
EstonianBB
FINNGEN
GENEVA_AFFY
GENEVA_ILLUMINA
GENUFADext (SHIP1)
GENUFAD (SHIP2)
GERA
GINILISAnorth
GINILISAsouth
HEALTH2006
HUNT
INMA
IOW
MAAS
MAS_HNR
MoBa_children
NCRC_ADC
NFBC66
NTR
PIAMA
Raine study
SALTY
SAPALDIA_new
SAPALDIA_old
SWS
TwinGene
TwinsUK
UKBB
Meta−analysis (I^2 = 18.83% [0.00%; 49.87%])
Multi−ancestry cohorts:
GALA
GENR
RIKEN
SAGE
SAPPHIRE
Replication in 23andme:
European
African
Latino
0.50
1.0
2.0
4.0
Odds Ratio

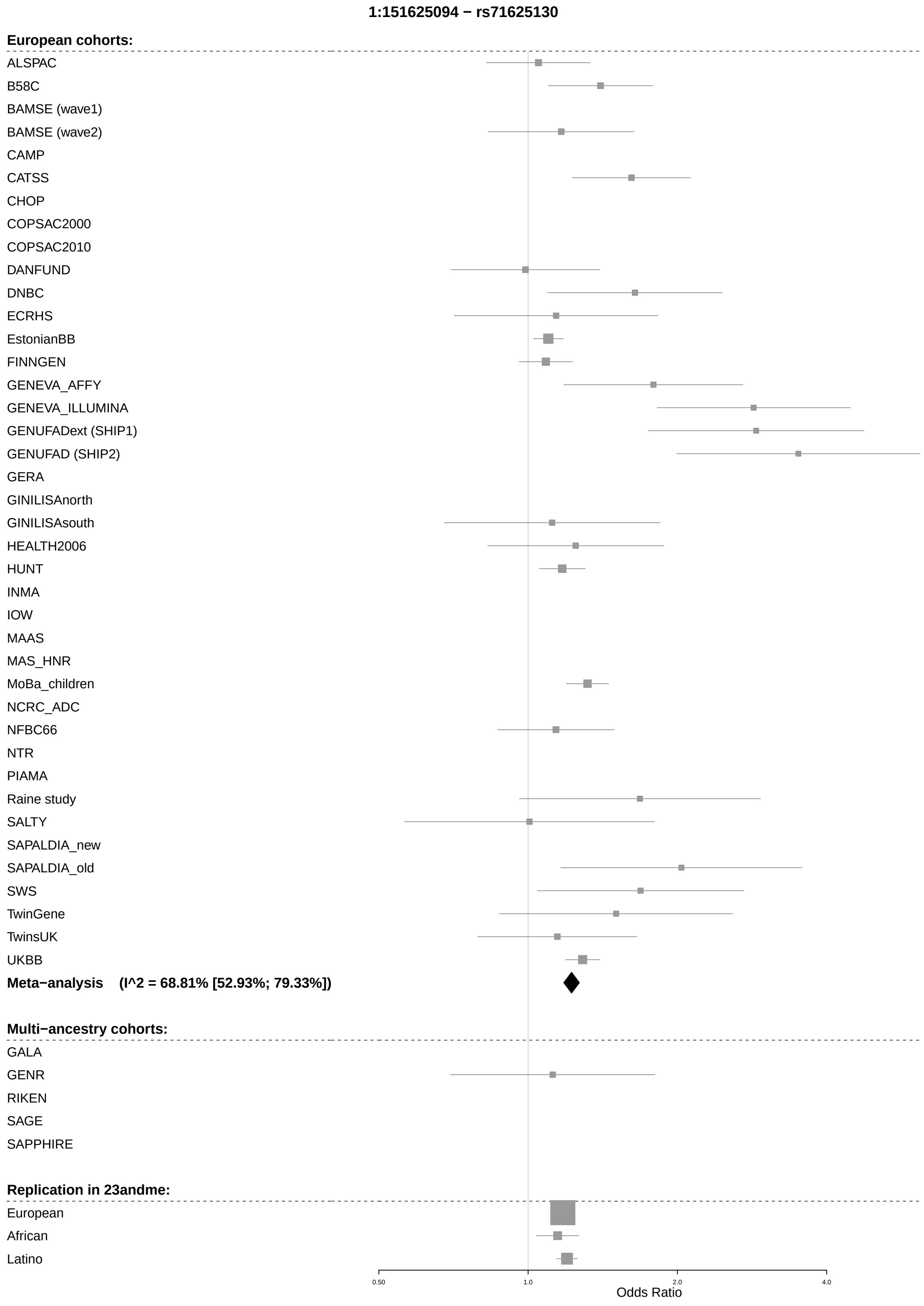
1:151625094 − rs71625130
European cohorts:
ALSPAC
B58C
BAMSE (wave1)
BAMSE (wave2)
CAMP
CATSS
CHOP
COPSAC2000
COPSAC2010
DANFUND
DNBC
ECRHS
EstonianBB
FINNGEN
GENEVA_AFFY
GENEVA_ILLUMINA
GENUFADext (SHIP1)
GENUFAD (SHIP2)
GERA
GINILISAnorth
GINILISAsouth
HEALTH2006
HUNT
INMA
IOW
MAAS
MAS_HNR
MoBa_children
NCRC_ADC
NFBC66
NTR
PIAMA
Raine study
SALTY
SAPALDIA_new
SAPALDIA_old
SWS
TwinGene
TwinsUK
UKBB
Meta−analysis (I^2 = 68.81% [52.93%; 79.33%])
Multi−ancestry cohorts:
GALA
GENR
RIKEN
SAGE
SAPPHIRE
Replication in 23andme:
European
African
Latino
0.50
1.0
2.0
4.0
Odds Ratio

1:151626396 − rs149199808

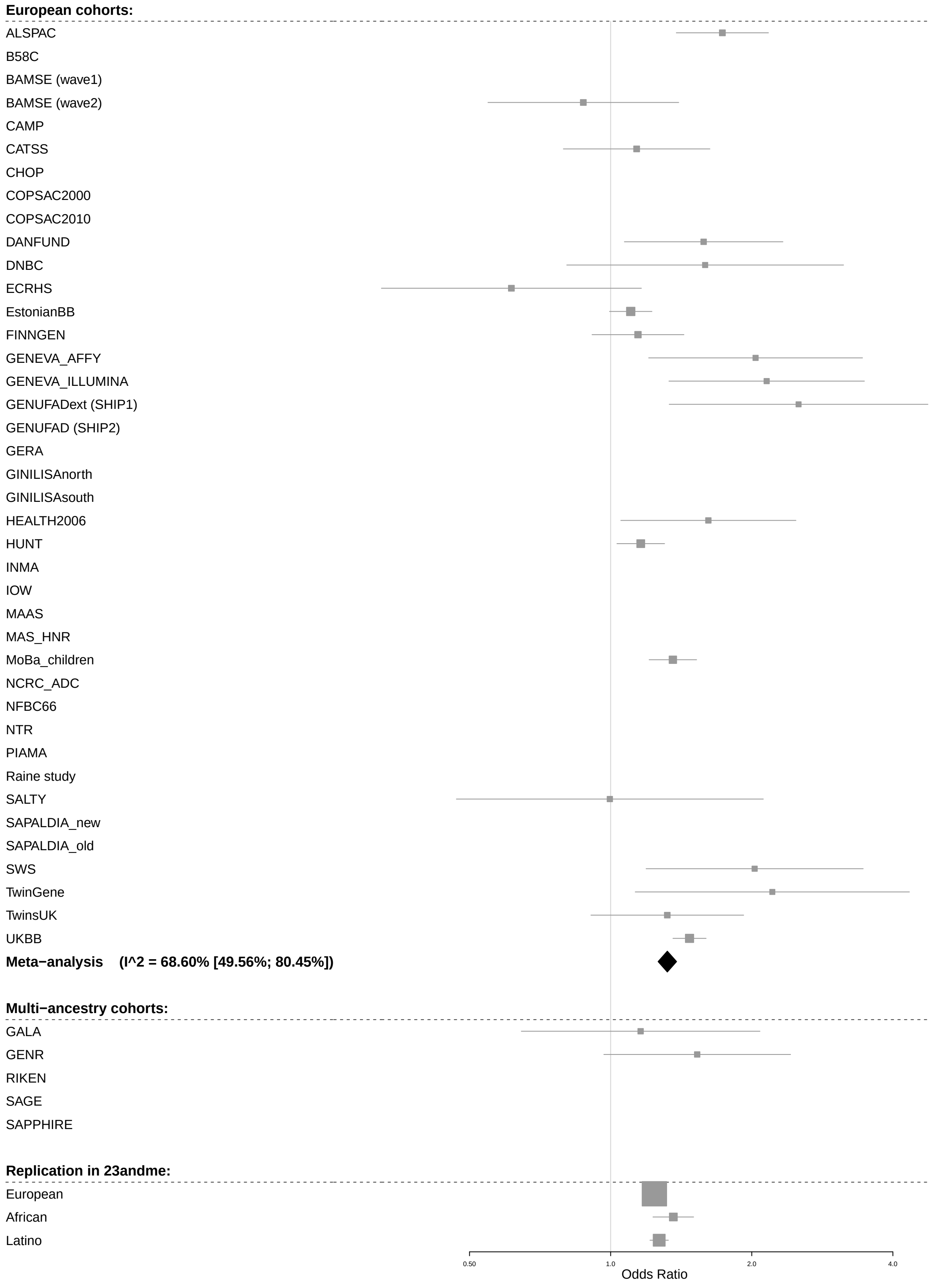

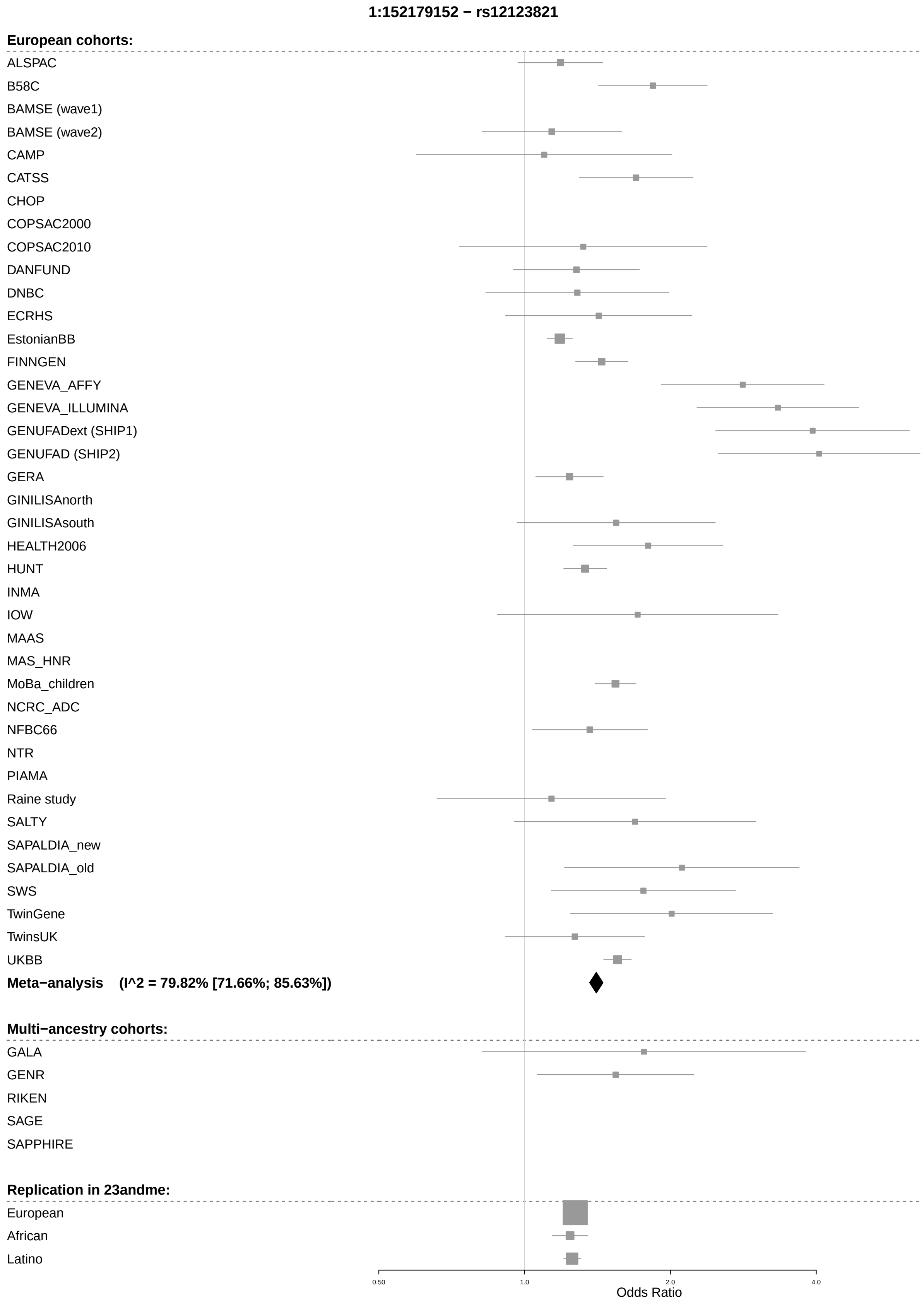
1:152179152 − rs12123821
European cohorts:
ALSPAC
B58C
BAMSE (wave1)
BAMSE (wave2)
CAMP
CATSS
CHOP
COPSAC2000
COPSAC2010
DANFUND
DNBC
ECRHS
EstonianBB
FINNGEN
GENEVA_AFFY
GENEVA_ILLUMINA
GENUFADext (SHIP1)
GENUFAD (SHIP2)
GERA
GINILISAnorth
GINILISAsouth
HEALTH2006
HUNT
INMA
IOW
MAAS
MAS_HNR
MoBa_children
NCRC_ADC
NFBC66
NTR
PIAMA
Raine study
SALTY
SAPALDIA_new
SAPALDIA_old
SWS
TwinGene
TwinsUK
UKBB
Meta−analysis (I^2 = 79.82% [71.66%; 85.63%])
Multi−ancestry cohorts:
GALA
GENR
RIKEN
SAGE
SAPPHIRE
Replication in 23andme:
European
African
Latino
0.50
1.0
2.0
4.0
Odds Ratio

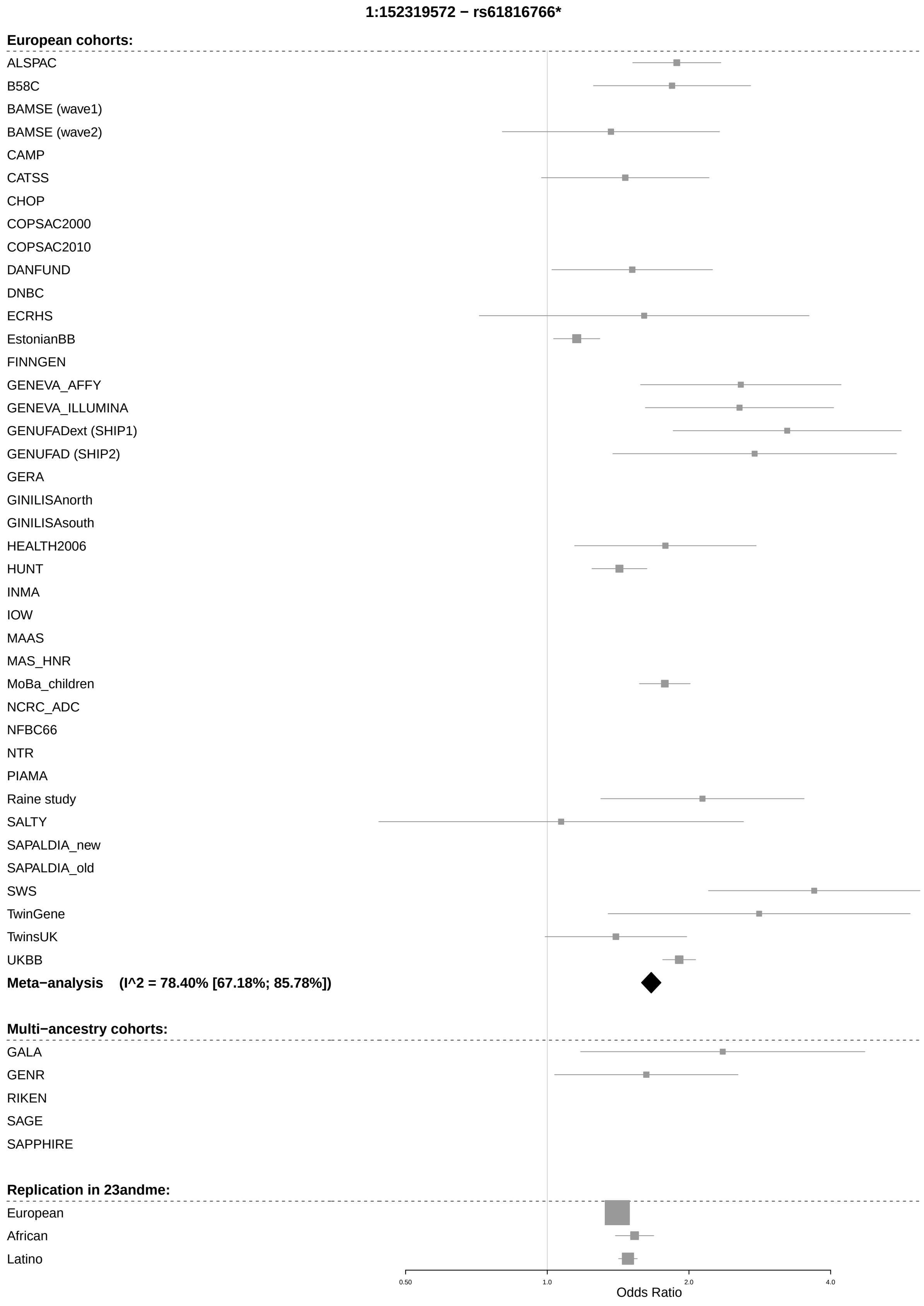
1:152319572 − rs61816766*
European cohorts:
ALSPAC
B58C
BAMSE (wave1)
BAMSE (wave2)
CAMP
CATSS
CHOP
COPSAC2000
COPSAC2010
DANFUND
DNBC
ECRHS
EstonianBB
FINNGEN
GENEVA_AFFY
GENEVA_ILLUMINA
GENUFADext (SHIP1)
GENUFAD (SHIP2)
GERA
GINILISAnorth
GINILISAsouth
HEALTH2006
HUNT
INMA
IOW
MAAS
MAS_HNR
MoBa_children
NCRC_ADC
NFBC66
NTR
PIAMA
Raine study
SALTY
SAPALDIA_new
SAPALDIA_old
SWS
TwinGene
TwinsUK
UKBB
Meta−analysis (I^2 = 78.40% [67.18%; 85.78%])
Multi−ancestry cohorts:
GALA
GENR
RIKEN
SAGE
SAPPHIRE
Replication in 23andme:
European
African
Latino
0.50
1.0
2.0
4.0
Odds Ratio

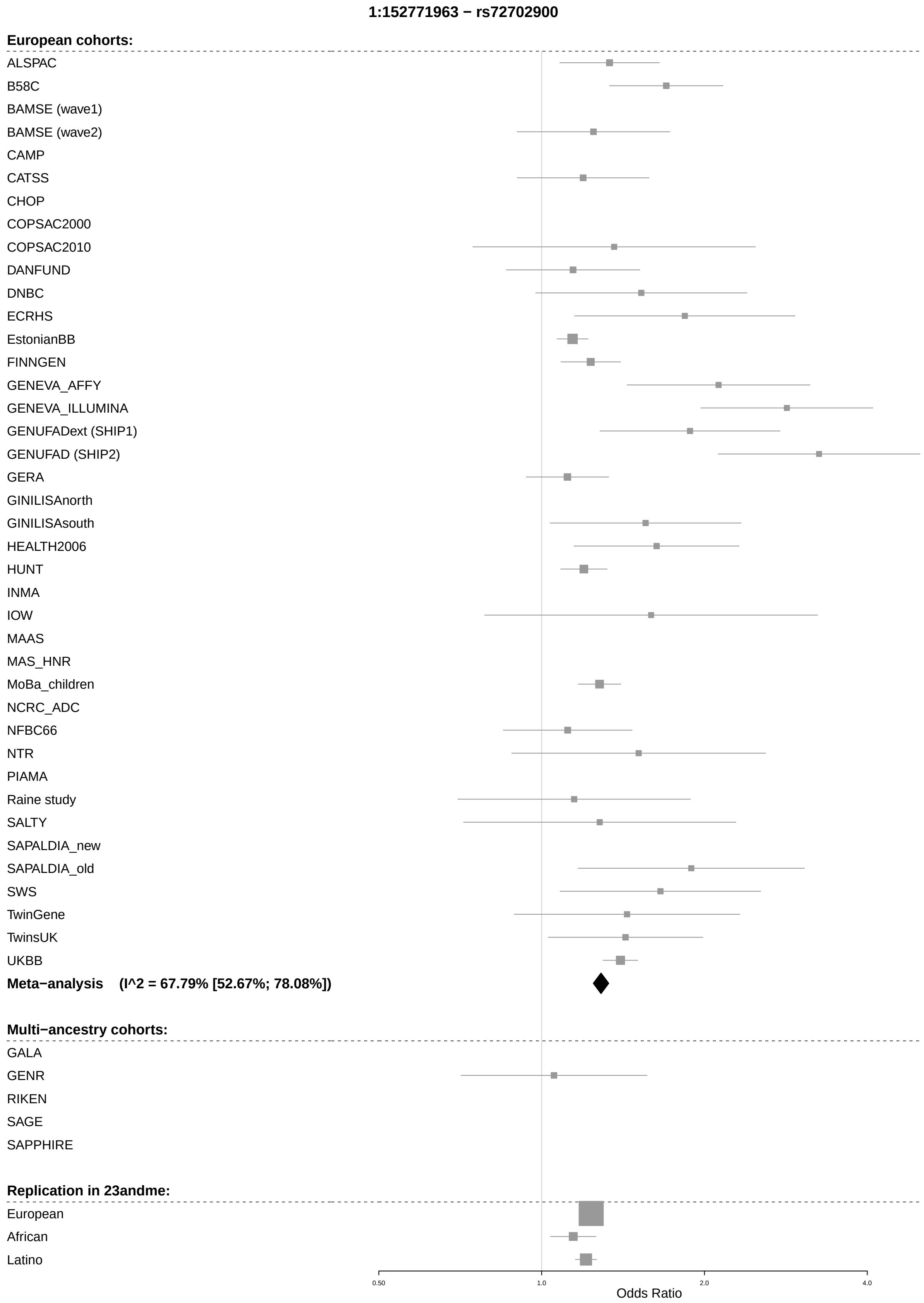
1:152771963 − rs72702900
European cohorts:
ALSPAC
B58C
BAMSE (wave1)
BAMSE (wave2)
CAMP
CATSS
CHOP
COPSAC2000
COPSAC2010
DANFUND
DNBC
ECRHS
EstonianBB
FINNGEN
GENEVA_AFFY
GENEVA_ILLUMINA
GENUFADext (SHIP1)
GENUFAD (SHIP2)
GERA
GINILISAnorth
GINILISAsouth
HEALTH2006
HUNT
INMA
IOW
MAAS
MAS_HNR
MoBa_children
NCRC_ADC
NFBC66
NTR
PIAMA
Raine study
SALTY
SAPALDIA_new
SAPALDIA_old
SWS
TwinGene
TwinsUK
UKBB
Meta−analysis (I^2 = 67.79% [52.67%; 78.08%])
Multi−ancestry cohorts:
GALA
GENR
RIKEN
SAGE
SAPPHIRE
Replication in 23andme:
European
African
Latino
0.50
1.0
2.0
4.0
Odds Ratio

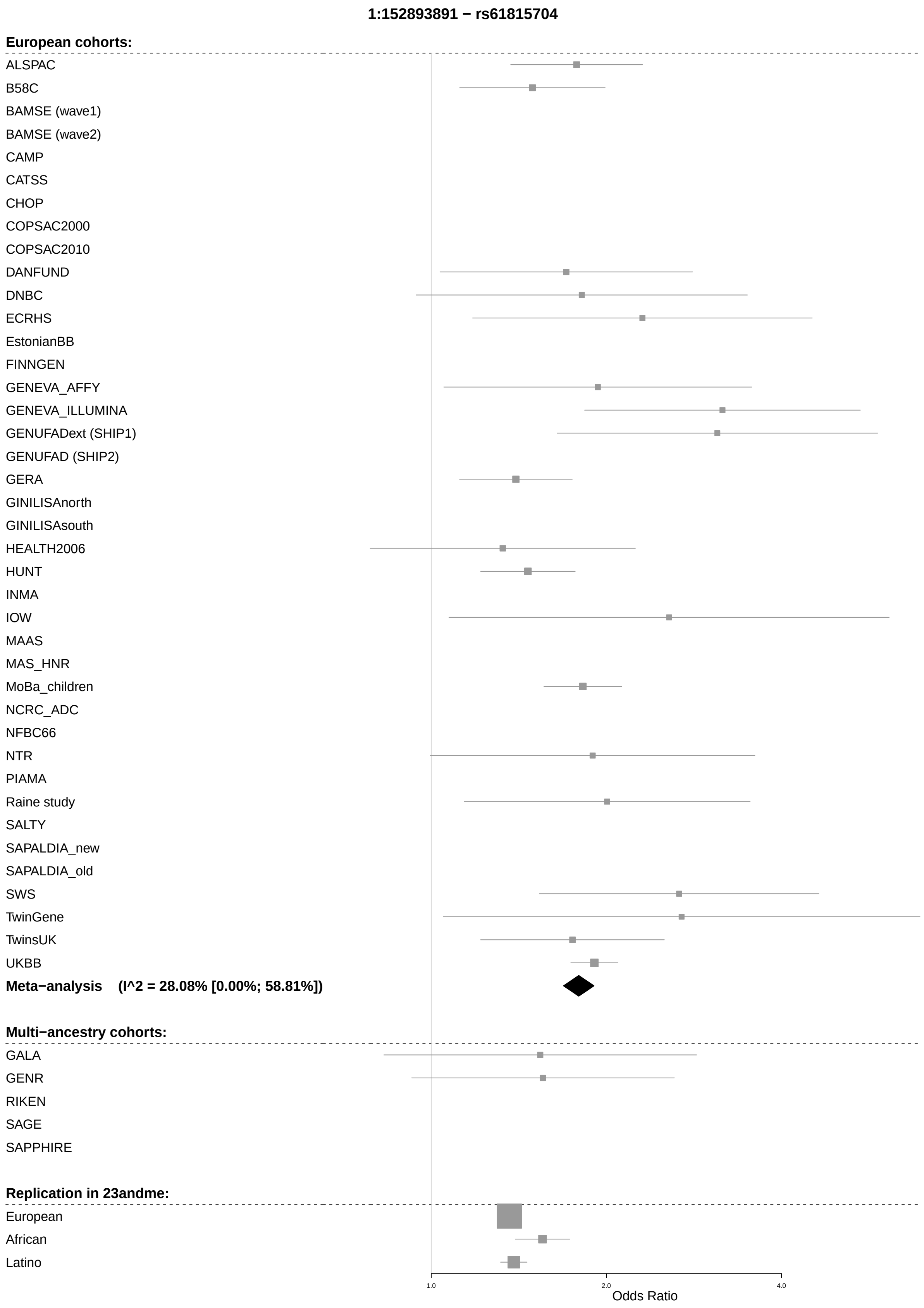
1:152893891 − rs61815704
European cohorts:
ALSPAC
B58C
BAMSE (wave1)
BAMSE (wave2)
CAMP
CATSS
CHOP
COPSAC2000
COPSAC2010
DANFUND
DNBC
ECRHS
EstonianBB
FINNGEN
GENEVA_AFFY
GENEVA_ILLUMINA
GENUFADext (SHIP1)
GENUFAD (SHIP2)
GERA
GINILISAnorth
GINILISAsouth
HEALTH2006
HUNT
INMA
IOW
MAAS
MAS_HNR
MoBa_children
NCRC_ADC
NFBC66
NTR
PIAMA
Raine study
SALTY
SAPALDIA_new
SAPALDIA_old
SWS
TwinGene
TwinsUK
UKBB
Meta−analysis (I^2 = 28.08% [0.00%; 58.81%])
Multi−ancestry cohorts:
GALA
GENR
RIKEN
SAGE
SAPPHIRE
Replication in 23andme:
European
African
Latino
1.0
2.0
4.0
Odds Ratio

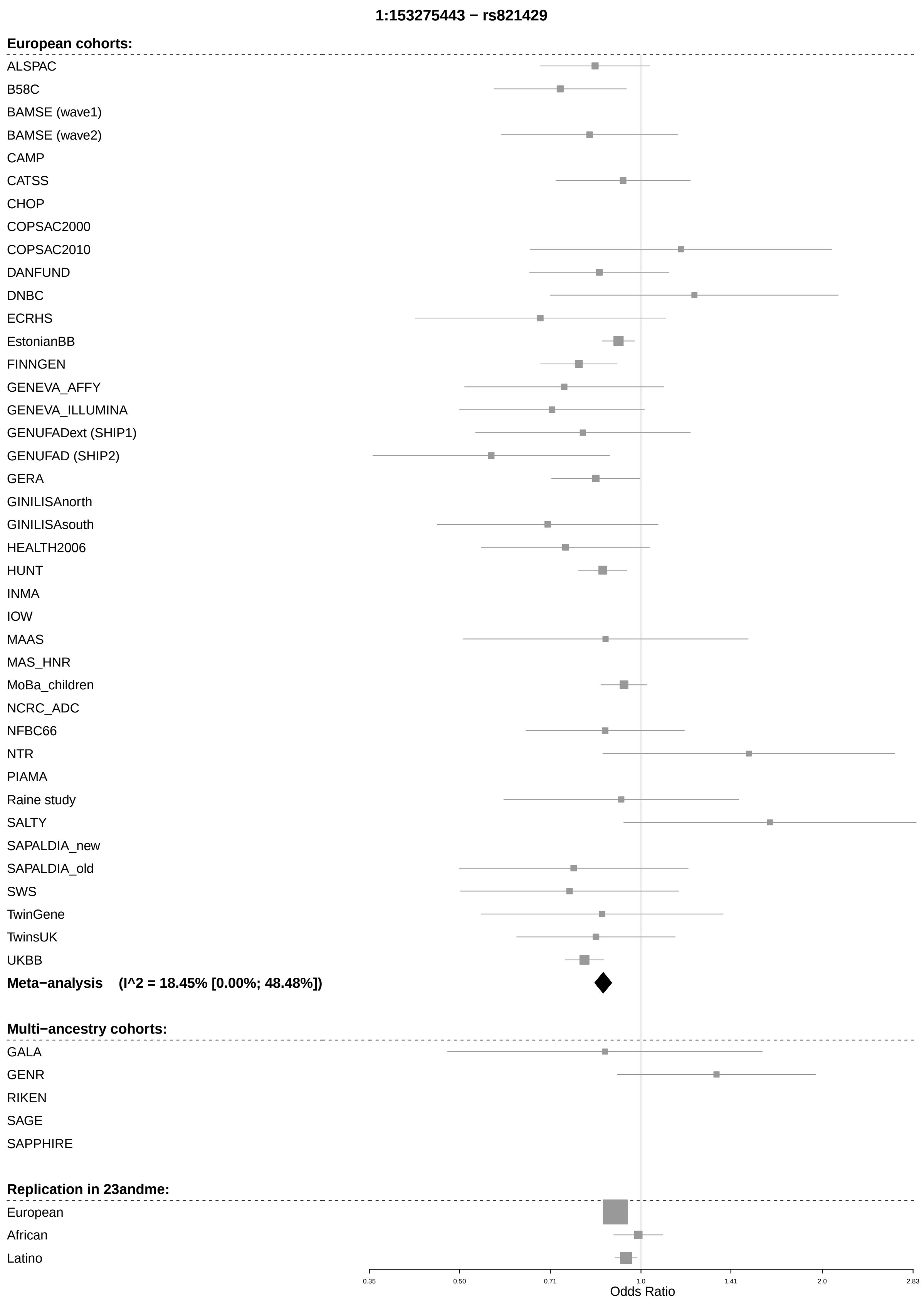

1:153275443 − rs821429
European cohorts:
ALSPAC
B58C
BAMSE (wave1)
BAMSE (wave2)
CAMP
CATSS
CHOP
COPSAC2000
COPSAC2010
DANFUND
DNBC
ECRHS
EstonianBB
FINNGEN
GENEVA_AFFY
GENEVA_ILLUMINA
GENUFADext (SHIP1)
GENUFAD (SHIP2)
GERA
GINILISAnorth
GINILISAsouth
HEALTH2006
HUNT
INMA
IOW
MAAS
MAS_HNR
MoBa_children
NCRC_ADC
NFBC66
NTR
PIAMA
Raine study
SALTY
SAPALDIA_new
SAPALDIA_old
SWS
TwinGene
TwinsUK
UKBB
Meta−analysis (I^2 = 18.45% [0.00%; 48.48%])
Multi−ancestry cohorts:
GALA
GENR
RIKEN
SAGE
SAPPHIRE
Replication in 23andme:
European
African
Latino
0.35
0.50
0.71
1.0
1.41
2.0
2.83
Odds Ratio

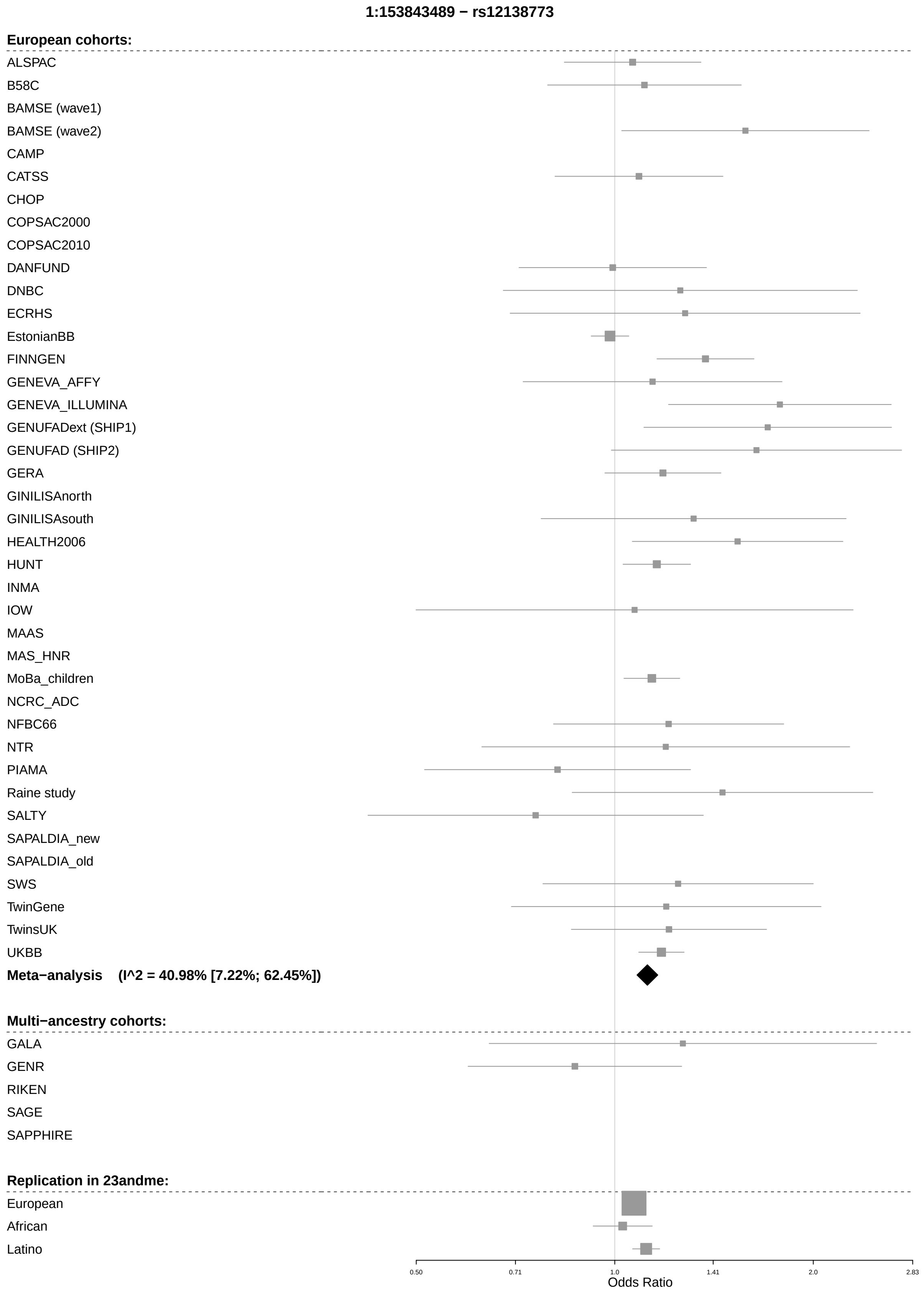
1:153843489 − rs12138773
European cohorts:
ALSPAC
B58C
BAMSE (wave1)
BAMSE (wave2)
CAMP
CATSS
CHOP
COPSAC2000
COPSAC2010
DANFUND
DNBC
ECRHS
EstonianBB
FINNGEN
GENEVA_AFFY
GENEVA_ILLUMINA
GENUFADext (SHIP1)
GENUFAD (SHIP2)
GERA
GINILISAnorth
GINILISAsouth
HEALTH2006
HUNT
INMA
IOW
MAAS
MAS_HNR
MoBa_children
NCRC_ADC
NFBC66
NTR
PIAMA
Raine study
SALTY
SAPALDIA_new
SAPALDIA_old
SWS
TwinGene
TwinsUK
UKBB
Meta−analysis (I^2 = 40.98% [7.22%; 62.45%])
Multi−ancestry cohorts:
GALA
GENR
RIKEN
SAGE
SAPPHIRE
Replication in 23andme:
European
African
Latino
0.50
0.71
1.0
1.41
2.0
2.83
Odds Ratio

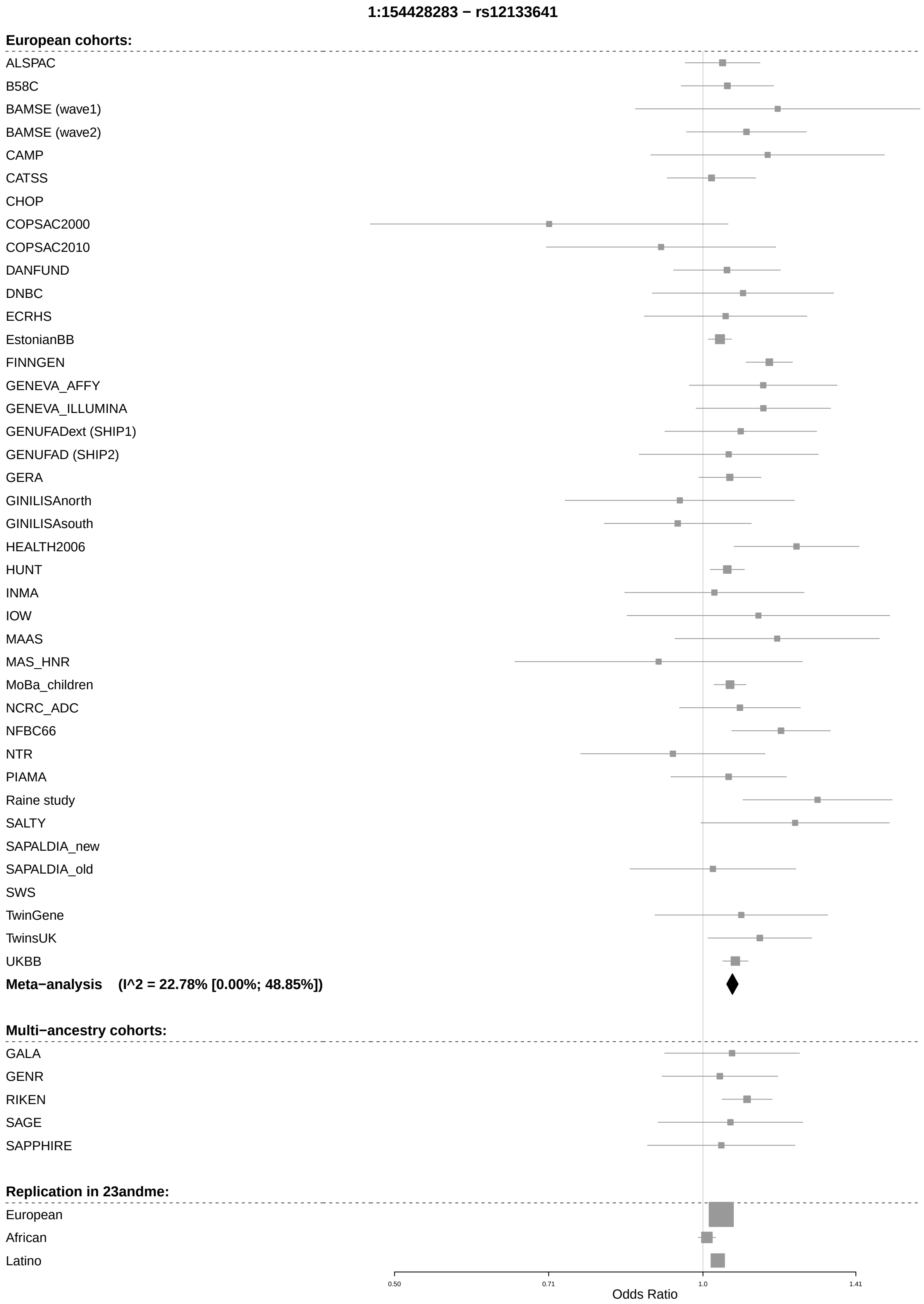
1:154428283 − rs12133641
European cohorts:
ALSPAC
B58C
BAMSE (wave1)
BAMSE (wave2)
CAMP
CATSS
CHOP
COPSAC2000
COPSAC2010
DANFUND
DNBC
ECRHS
EstonianBB
FINNGEN
GENEVA_AFFY
GENEVA_ILLUMINA
GENUFADext (SHIP1)
GENUFAD (SHIP2)
GERA
GINILISAnorth
GINILISAsouth
HEALTH2006
HUNT
INMA
IOW
MAAS
MAS_HNR
MoBa_children
NCRC_ADC
NFBC66
NTR
PIAMA
Raine study
SALTY
SAPALDIA_new
SAPALDIA_old
SWS
TwinGene
TwinsUK
UKBB
Meta−analysis (I^2 = 22.78% [0.00%; 48.85%])
Multi−ancestry cohorts:
GALA
GENR
RIKEN
SAGE
SAPPHIRE
Replication in 23andme:
European
African
Latino
0.50
0.71
1.0
1.41
Odds Ratio

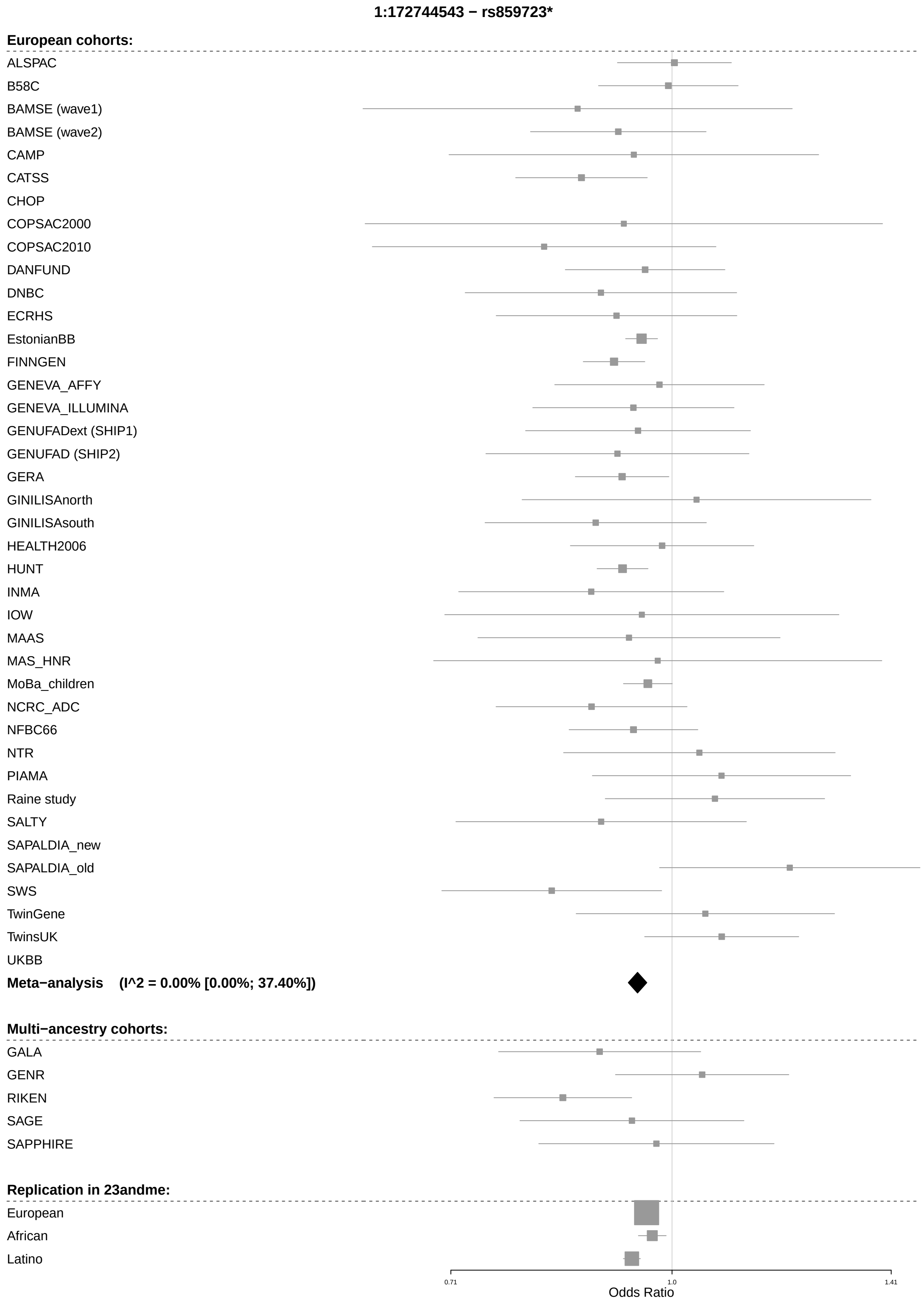
1:172744543 − rs859723*
European cohorts:
ALSPAC
B58C
BAMSE (wave1)
BAMSE (wave2)
CAMP
CATSS
CHOP
COPSAC2000
COPSAC2010
DANFUND
DNBC
ECRHS
EstonianBB
FINNGEN
GENEVA_AFFY
GENEVA_ILLUMINA
GENUFADext (SHIP1)
GENUFAD (SHIP2)
GERA
GINILISAnorth
GINILISAsouth
HEALTH2006
HUNT
INMA
IOW
MAAS
MAS_HNR
MoBa_children
NCRC_ADC
NFBC66
NTR
PIAMA
Raine study
SALTY
SAPALDIA_new
SAPALDIA_old
SWS
TwinGene
TwinsUK
UKBB
Meta−analysis (I^2 = 0.00% [0.00%; 37.40%])
Multi−ancestry cohorts:
GALA
GENR
RIKEN
SAGE
SAPPHIRE
Replication in 23andme:
European
African
Latino
0.71
1.0
1.41
Odds Ratio

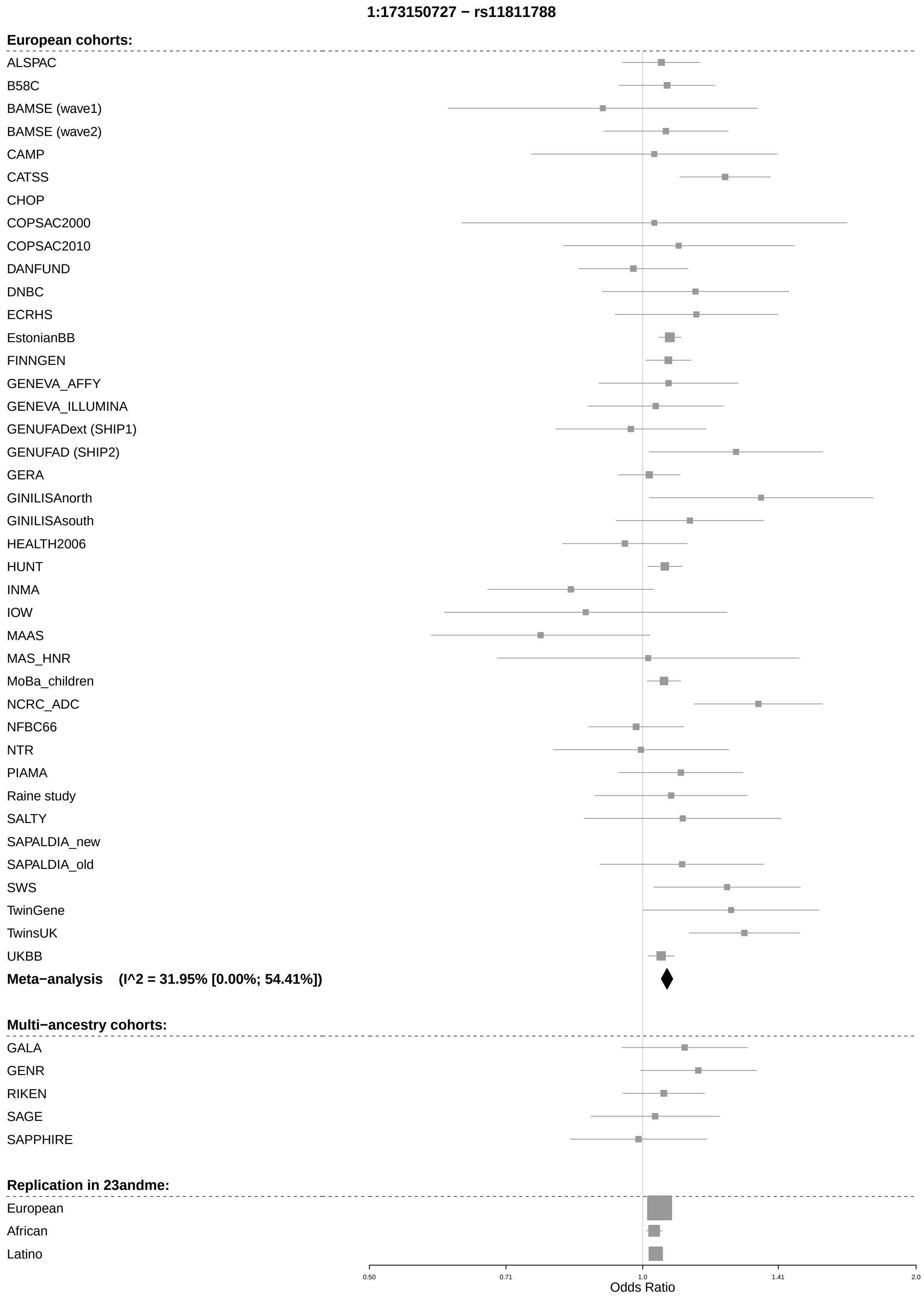
1:173150727 − rs11811788
European cohorts:
ALSPAC
B58C
BAMSE (wave1)
BAMSE (wave2)
CAMP
CATSS
CHOP
COPSAC2000
COPSAC2010
DANFUND
DNBC
ECRHS
EstonianBB
FINNGEN
GENEVA_AFFY
GENEVA_ILLUMINA
GENUFADext (SHIP1)
GENUFAD (SHIP2)
GERA
GINILISAnorth
GINILISAsouth
HEALTH2006
HUNT
INMA
IOW
MAAS
MAS_HNR
MoBa_children
NCRC_ADC
NFBC66
NTR
PIAMA
Raine study
SALTY
SAPALDIA_new
SAPALDIA_old
SWS
TwinGene
TwinsUK
UKBB
Meta−analysis (I^2 = 31.95% [0.00%; 54.41%])
Multi−ancestry cohorts:
GALA
GENR
RIKEN
SAGE
SAPPHIRE
Replication in 23andme:
European
African
Latino
0.50
0.71
1.0
1.41
2.0
Odds Ratio

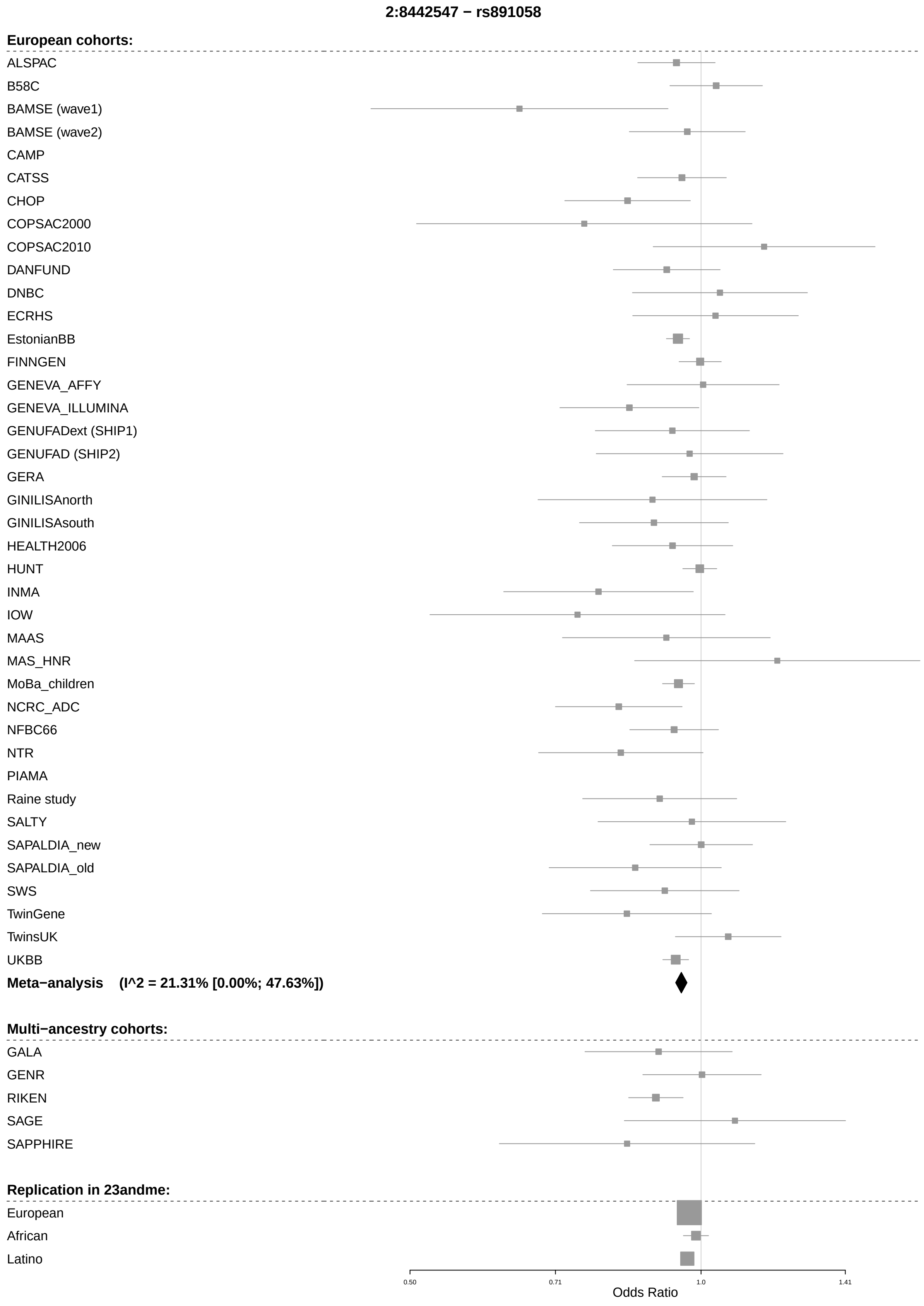
2:8442547 − rs891058
European cohorts:
ALSPAC
B58C
BAMSE (wave1)
BAMSE (wave2)
CAMP
CATSS
CHOP
COPSAC2000
COPSAC2010
DANFUND
DNBC
ECRHS
EstonianBB
FINNGEN
GENEVA_AFFY
GENEVA_ILLUMINA
GENUFADext (SHIP1)
GENUFAD (SHIP2)
GERA
GINILISAnorth
GINILISAsouth
HEALTH2006
HUNT
INMA
IOW
MAAS
MAS_HNR
MoBa_children
NCRC_ADC
NFBC66
NTR
PIAMA
Raine study
SALTY
SAPALDIA_new
SAPALDIA_old
SWS
TwinGene
TwinsUK
UKBB
Meta−analysis (I^2 = 21.31% [0.00%; 47.63%])
Multi−ancestry cohorts:
GALA
GENR
RIKEN
SAGE
SAPPHIRE
Replication in 23andme:
European
African
Latino
0.50
0.71
1.0
1.41
Odds Ratio

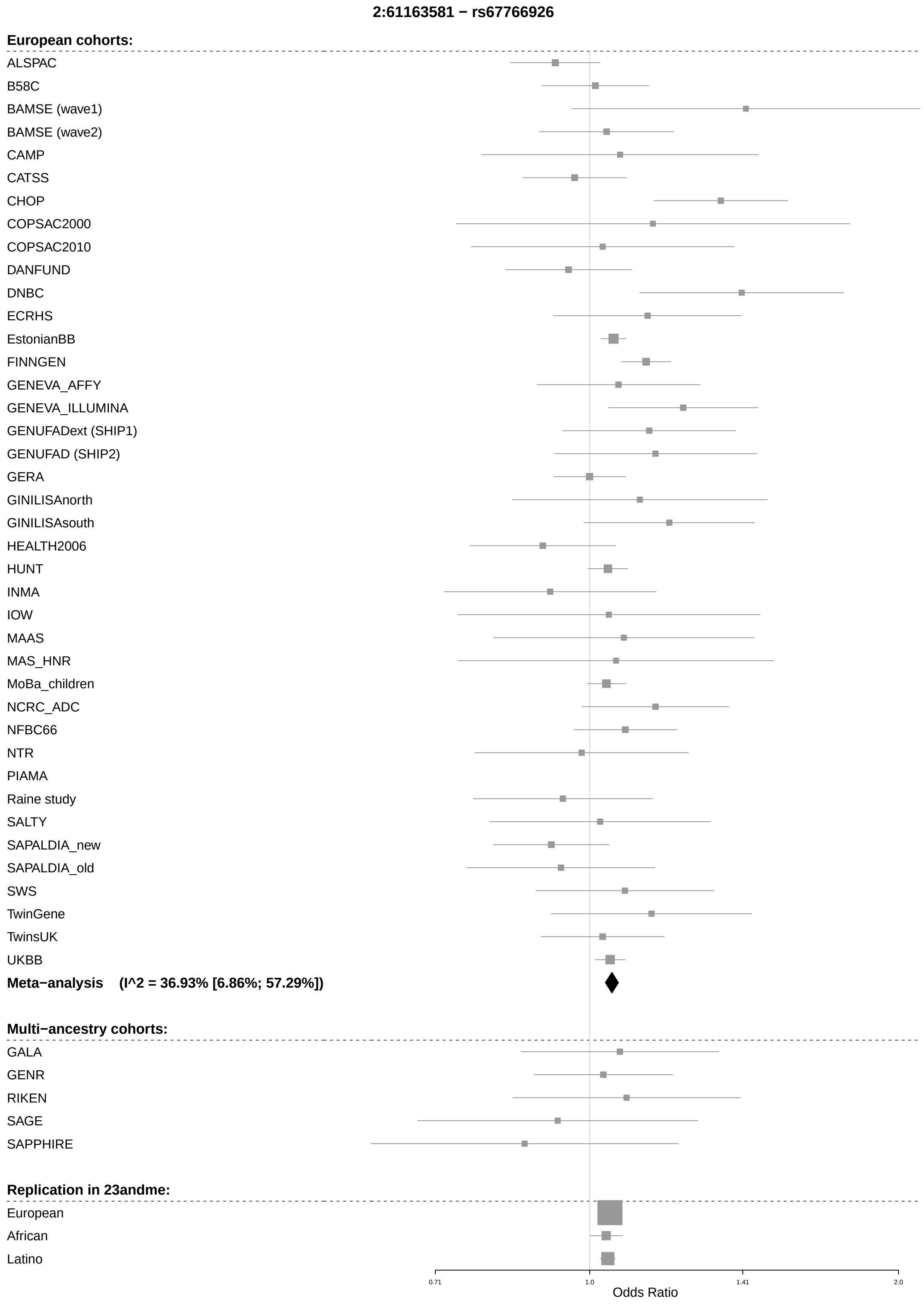
2:61163581 − rs67766926
European cohorts:
ALSPAC
B58C
BAMSE (wave1)
BAMSE (wave2)
CAMP
CATSS
CHOP
COPSAC2000
COPSAC2010
DANFUND
DNBC
ECRHS
EstonianBB
FINNGEN
GENEVA_AFFY
GENEVA_ILLUMINA
GENUFADext (SHIP1)
GENUFAD (SHIP2)
GERA
GINILISAnorth
GINILISAsouth
HEALTH2006
HUNT
INMA
IOW
MAAS
MAS_HNR
MoBa_children
NCRC_ADC
NFBC66
NTR
PIAMA
Raine study
SALTY
SAPALDIA_new
SAPALDIA_old
SWS
TwinGene
TwinsUK
UKBB
Meta−analysis (I^2 = 36.93% [6.86%; 57.29%])
Multi−ancestry cohorts:
GALA
GENR
RIKEN
SAGE
SAPPHIRE
Replication in 23andme:
European
African
Latino
0.71
1.0
1.41
2.0
Odds Ratio

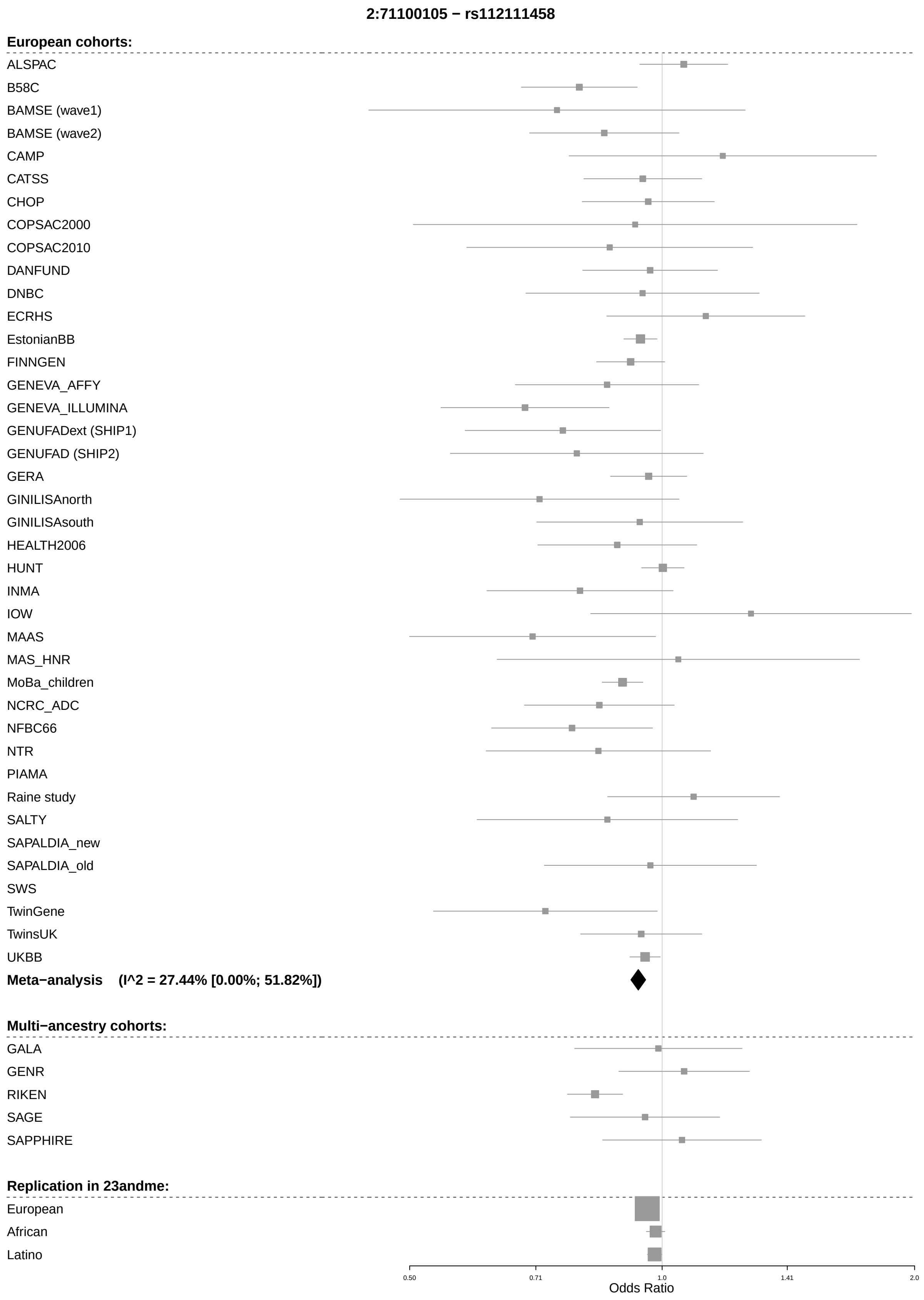
2:71100105 − rs112111458
European cohorts:
ALSPAC
B58C
BAMSE (wave1)
BAMSE (wave2)
CAMP
CATSS
CHOP
COPSAC2000
COPSAC2010
DANFUND
DNBC
ECRHS
EstonianBB
FINNGEN
GENEVA_AFFY
GENEVA_ILLUMINA
GENUFADext (SHIP1)
GENUFAD (SHIP2)
GERA
GINILISAnorth
GINILISAsouth
HEALTH2006
HUNT
INMA
IOW
MAAS
MAS_HNR
MoBa_children
NCRC_ADC
NFBC66
NTR
PIAMA
Raine study
SALTY
SAPALDIA_new
SAPALDIA_old
SWS
TwinGene
TwinsUK
UKBB
Meta−analysis (I^2 = 27.44% [0.00%; 51.82%])
Multi−ancestry cohorts:
GALA
GENR
RIKEN
SAGE
SAPPHIRE
Replication in 23andme:
European
African
Latino
0.50
0.71
1.0
1.41
2.0
Odds Ratio

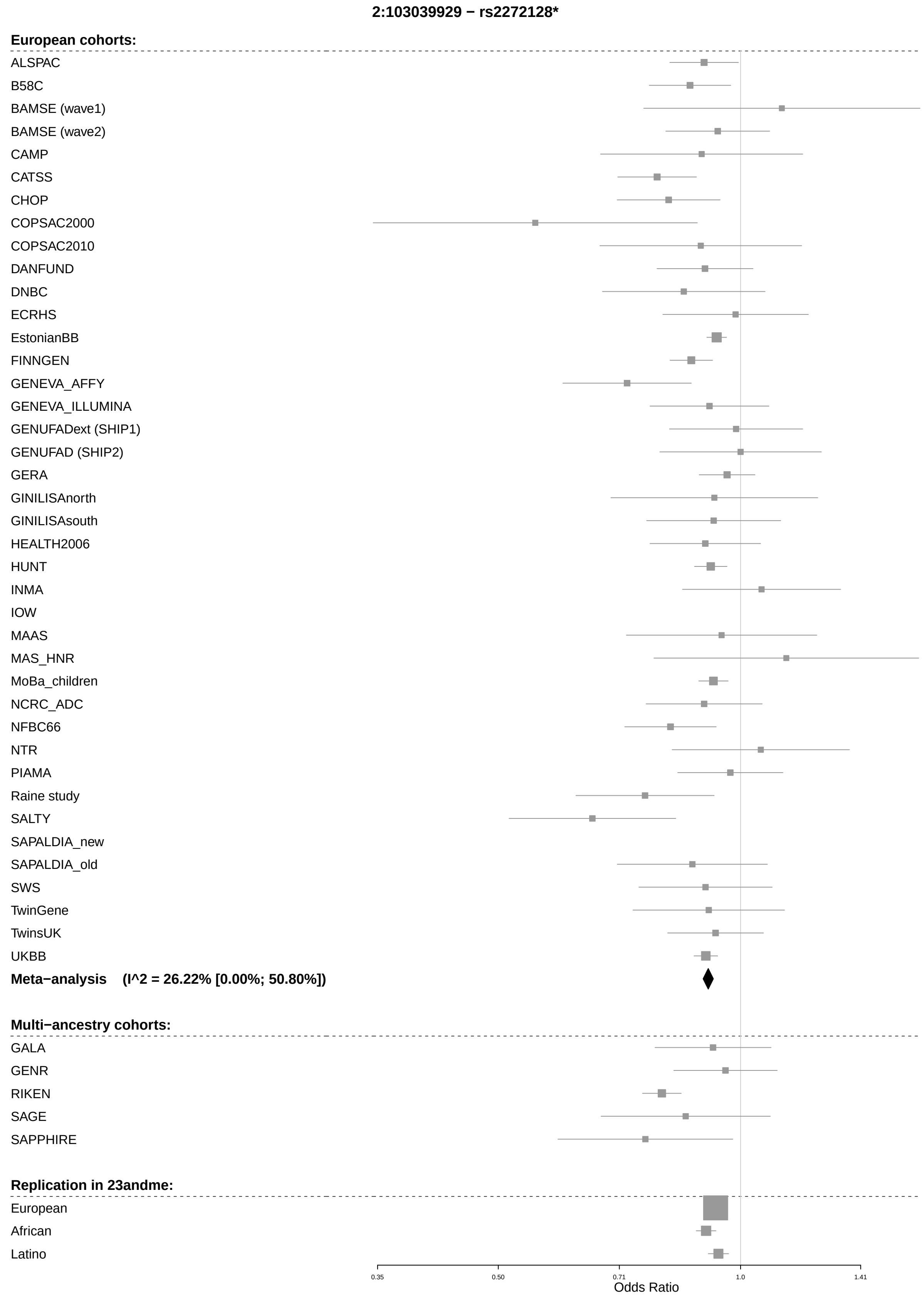
2:103039929 − rs2272128*
European cohorts:
ALSPAC
B58C
BAMSE (wave1)
BAMSE (wave2)
CAMP
CATSS
CHOP
COPSAC2000
COPSAC2010
DANFUND
DNBC
ECRHS
EstonianBB
FINNGEN
GENEVA_AFFY
GENEVA_ILLUMINA
GENUFADext (SHIP1)
GENUFAD (SHIP2)
GERA
GINILISAnorth
GINILISAsouth
HEALTH2006
HUNT
INMA
IOW
MAAS
MAS_HNR
MoBa_children
NCRC_ADC
NFBC66
NTR
PIAMA
Raine study
SALTY
SAPALDIA_new
SAPALDIA_old
SWS
TwinGene
TwinsUK
UKBB
Meta−analysis (I^2 = 26.22% [0.00%; 50.80%])
Multi−ancestry cohorts:
GALA
GENR
RIKEN
SAGE
SAPPHIRE
Replication in 23andme:
European
African
Latino
0.35
0.50
0.71
1.0
1.41
Odds Ratio

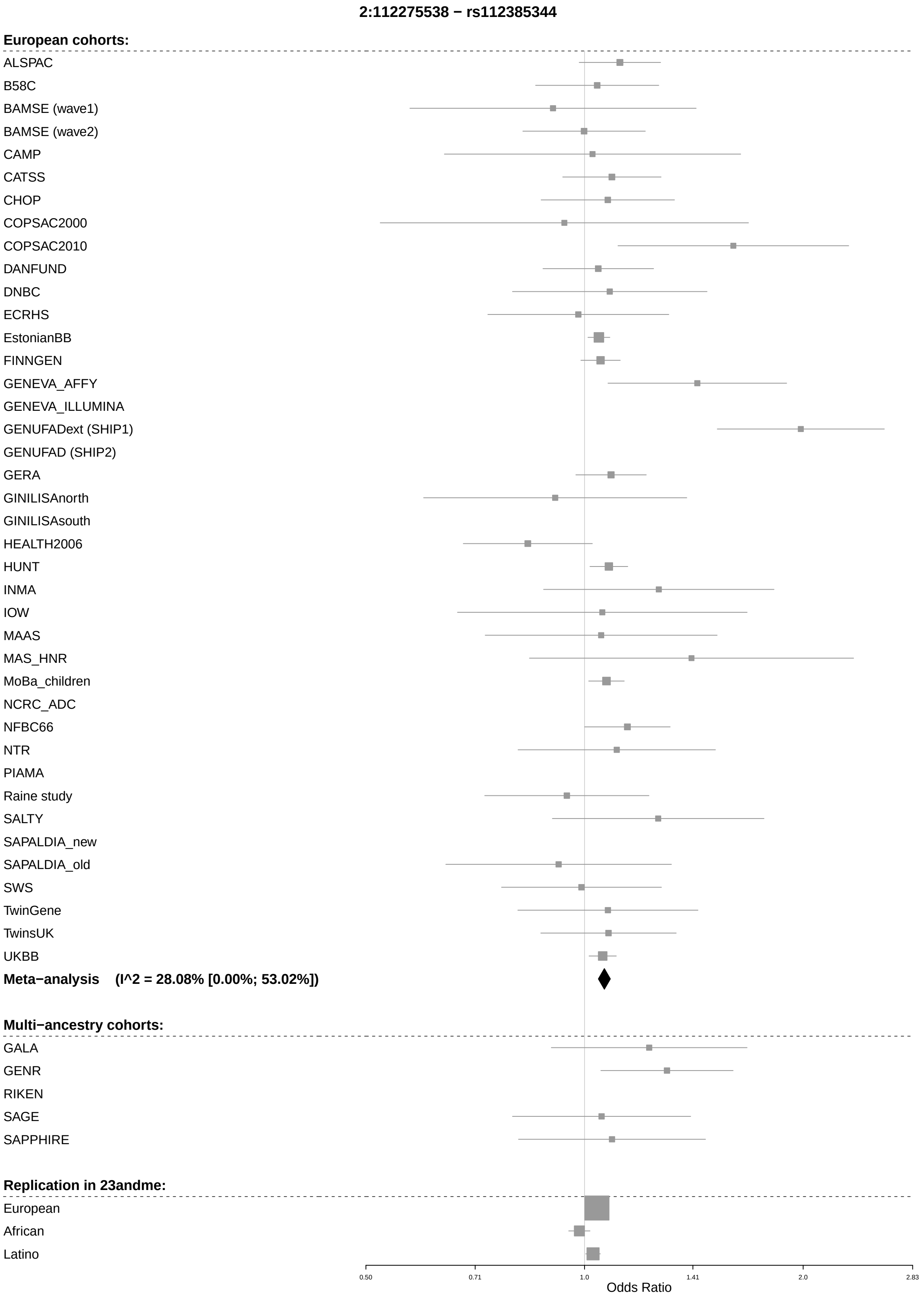

2:112275538 − rs112385344
European cohorts:
ALSPAC
B58C
BAMSE (wave1)
BAMSE (wave2)
CAMP
CATSS
CHOP
COPSAC2000
COPSAC2010
DANFUND
DNBC
ECRHS
EstonianBB
FINNGEN
GENEVA_AFFY
GENEVA_ILLUMINA
GENUFADext (SHIP1)
GENUFAD (SHIP2)
GERA
GINILISAnorth
GINILISAsouth
HEALTH2006
HUNT
INMA
IOW
MAAS
MAS_HNR
MoBa_children
NCRC_ADC
NFBC66
NTR
PIAMA
Raine study
SALTY
SAPALDIA_new
SAPALDIA_old
SWS
TwinGene
TwinsUK
UKBB
Meta−analysis (I^2 = 28.08% [0.00%; 53.02%])
Multi−ancestry cohorts:
GALA
GENR
RIKEN
SAGE
SAPPHIRE
Replication in 23andme:
European
African
Latino
0.50
0.71
1.0
1.41
2.0
2.83
Odds Ratio

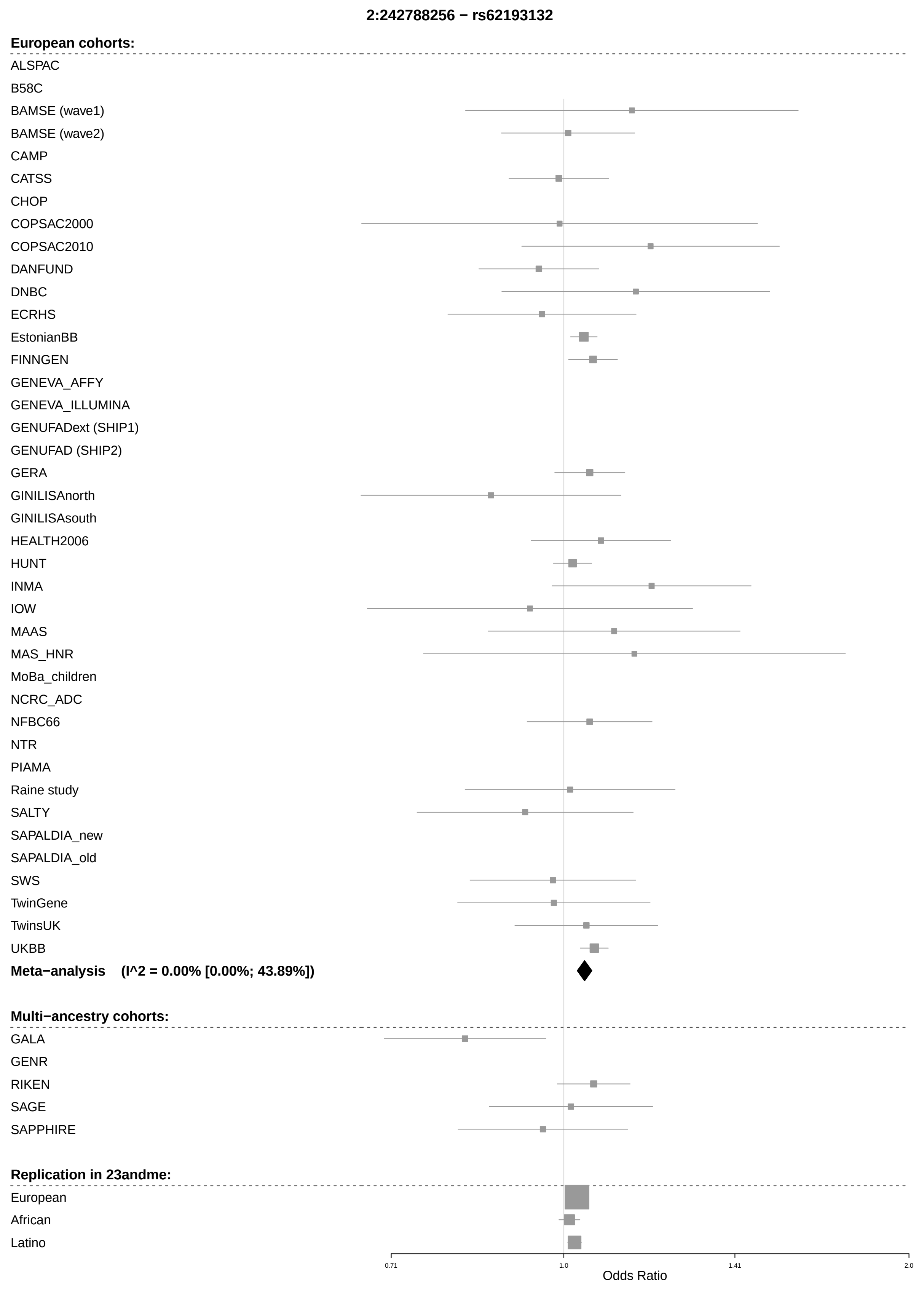
2:242788256 − rs62193132
European cohorts:
ALSPAC
B58C
BAMSE (wave1)
BAMSE (wave2)
CAMP
CATSS
CHOP
COPSAC2000
COPSAC2010
DANFUND
DNBC
ECRHS
EstonianBB
FINNGEN
GENEVA_AFFY
GENEVA_ILLUMINA
GENUFADext (SHIP1)
GENUFAD (SHIP2)
GERA
GINILISAnorth
GINILISAsouth
HEALTH2006
HUNT
INMA
IOW
MAAS
MAS_HNR
MoBa_children
NCRC_ADC
NFBC66
NTR
PIAMA
Raine study
SALTY
SAPALDIA_new
SAPALDIA_old
SWS
TwinGene
TwinsUK
UKBB
Meta−analysis (I^2 = 0.00% [0.00%; 43.89%])
Multi−ancestry cohorts:
GALA
GENR
RIKEN
SAGE
SAPPHIRE
Replication in 23andme:
European
African
Latino
0.71
1.0
1.41
2.0
Odds Ratio

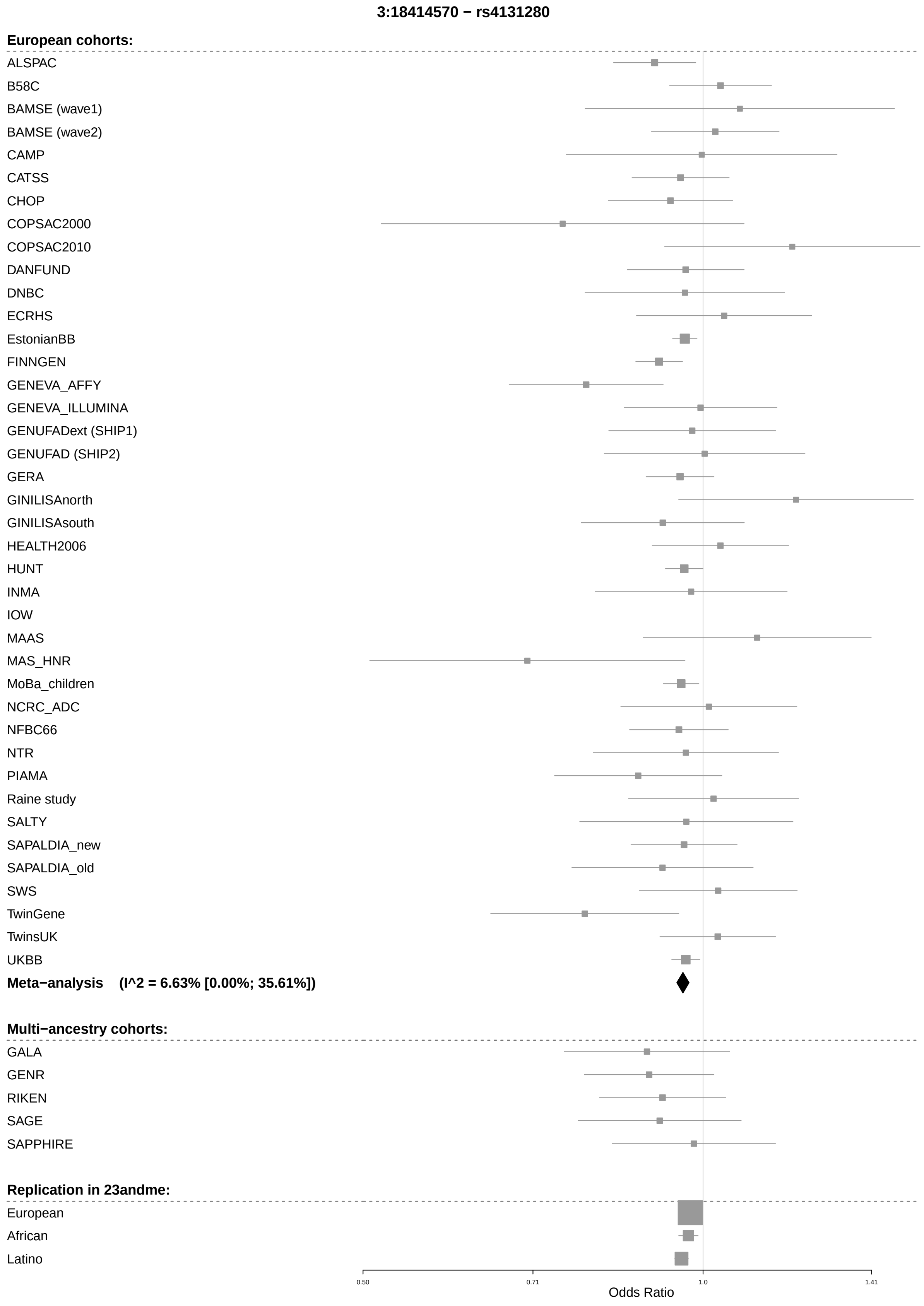
3:18414570 − rs4131280
European cohorts:
ALSPAC
B58C
BAMSE (wave1)
BAMSE (wave2)
CAMP
CATSS
CHOP
COPSAC2000
COPSAC2010
DANFUND
DNBC
ECRHS
EstonianBB
FINNGEN
GENEVA_AFFY
GENEVA_ILLUMINA
GENUFADext (SHIP1)
GENUFAD (SHIP2)
GERA
GINILISAnorth
GINILISAsouth
HEALTH2006
HUNT
INMA
IOW
MAAS
MAS_HNR
MoBa_children
NCRC_ADC
NFBC66
NTR
PIAMA
Raine study
SALTY
SAPALDIA_new
SAPALDIA_old
SWS
TwinGene
TwinsUK
UKBB
Meta−analysis (I^2 = 6.63% [0.00%; 35.61%])
Multi−ancestry cohorts:
GALA
GENR
RIKEN
SAGE
SAPPHIRE
Replication in 23andme:
European
African
Latino
0.50
0.71
1.0
1.41
Odds Ratio

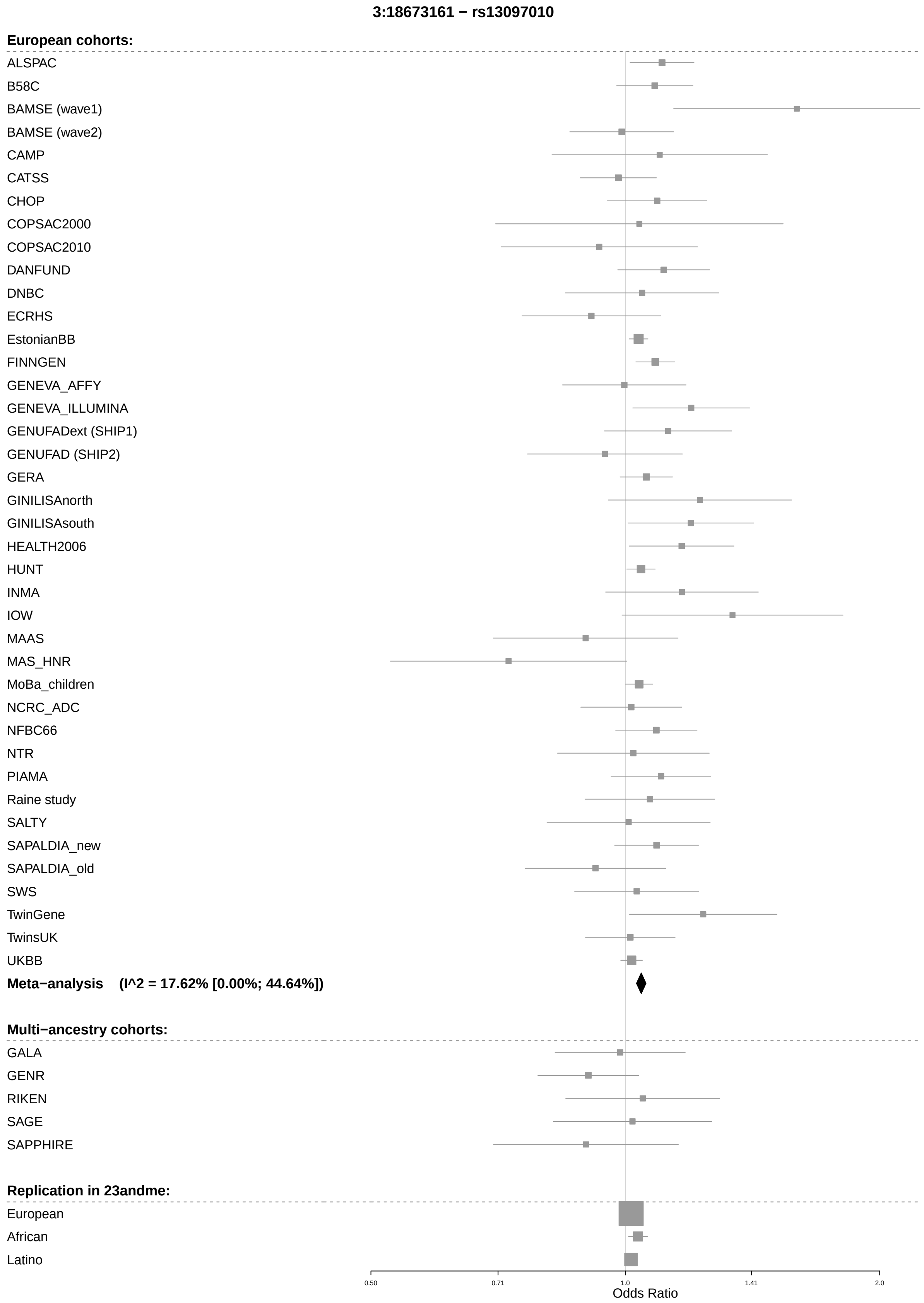
3:18673161 − rs13097010
European cohorts:
ALSPAC
B58C
BAMSE (wave1)
BAMSE (wave2)
CAMP
CATSS
CHOP
COPSAC2000
COPSAC2010
DANFUND
DNBC
ECRHS
EstonianBB
FINNGEN
GENEVA_AFFY
GENEVA_ILLUMINA
GENUFADext (SHIP1)
GENUFAD (SHIP2)
GERA
GINILISAnorth
GINILISAsouth
HEALTH2006
HUNT
INMA
IOW
MAAS
MAS_HNR
MoBa_children
NCRC_ADC
NFBC66
NTR
PIAMA
Raine study
SALTY
SAPALDIA_new
SAPALDIA_old
SWS
TwinGene
TwinsUK
UKBB
Meta−analysis (I^2 = 17.62% [0.00%; 44.64%])
Multi−ancestry cohorts:
GALA
GENR
RIKEN
SAGE
SAPPHIRE
Replication in 23andme:
European
African
Latino
0.50
0.71
1.0
1.41
2.0
Odds Ratio

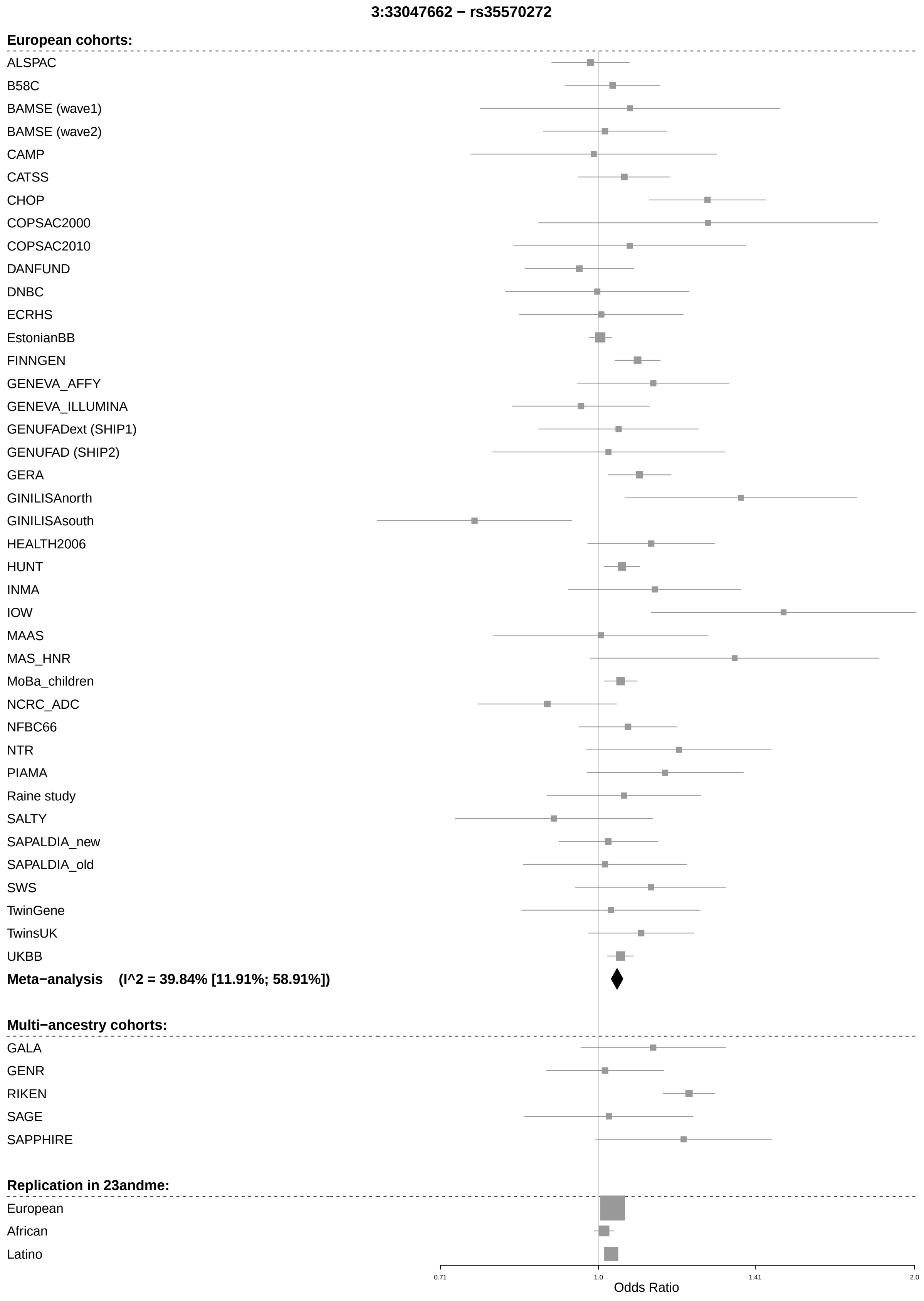
3:33047662 − rs35570272
European cohorts:
ALSPAC
B58C
BAMSE (wave1)
BAMSE (wave2)
CAMP
CATSS
CHOP
COPSAC2000
COPSAC2010
DANFUND
DNBC
ECRHS
EstonianBB
FINNGEN
GENEVA_AFFY
GENEVA_ILLUMINA
GENUFADext (SHIP1)
GENUFAD (SHIP2)
GERA
GINILISAnorth
GINILISAsouth
HEALTH2006
HUNT
INMA
IOW
MAAS
MAS_HNR
MoBa_children
NCRC_ADC
NFBC66
NTR
PIAMA
Raine study
SALTY
SAPALDIA_new
SAPALDIA_old
SWS
TwinGene
TwinsUK
UKBB
Meta−analysis (I^2 = 39.84% [11.91%; 58.91%])
Multi−ancestry cohorts:
GALA
GENR
RIKEN
SAGE
SAPPHIRE
Replication in 23andme:
European
African
Latino
0.71
1.0
1.41
2.0
Odds Ratio

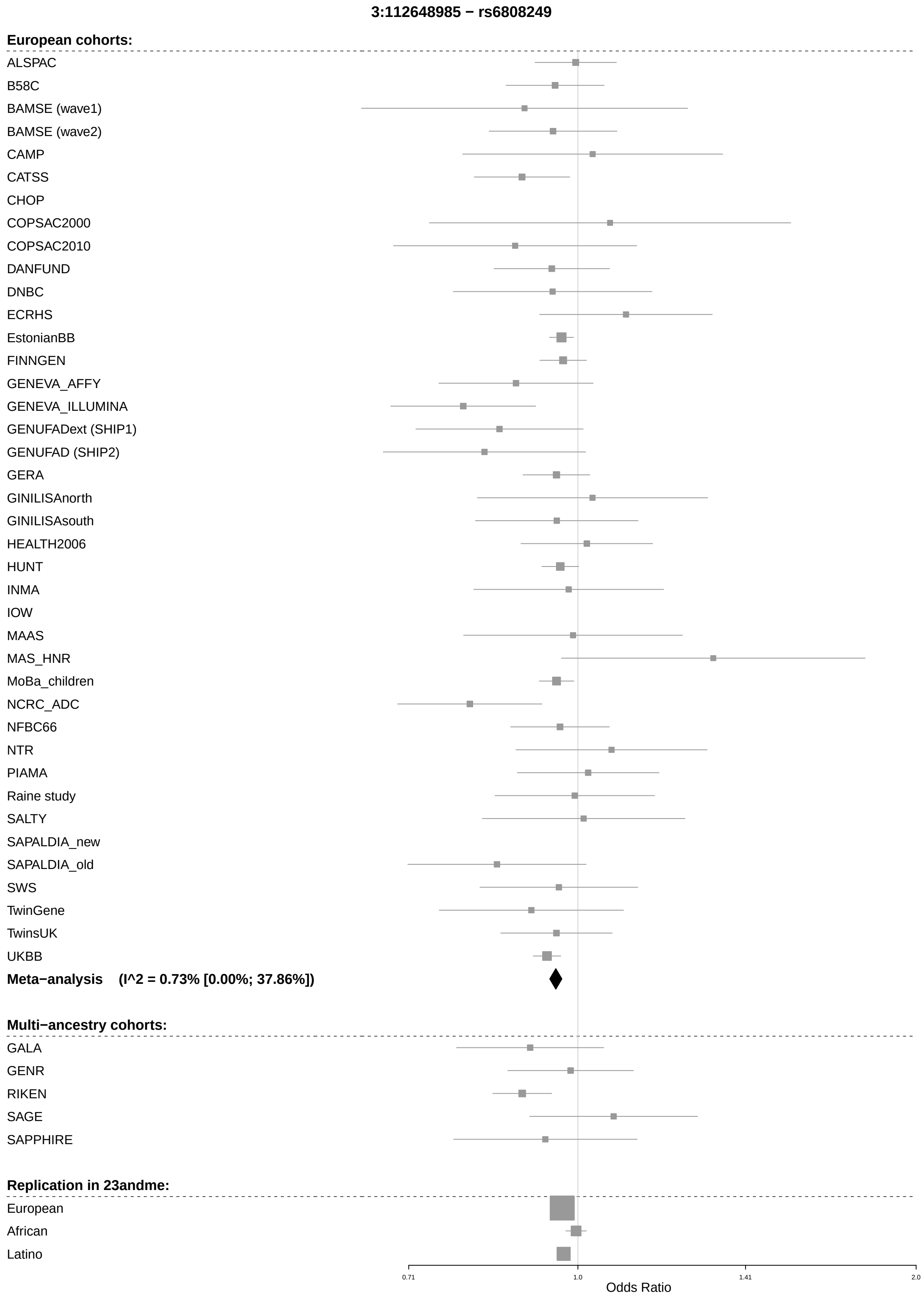

3:112648985 − rs6808249
European cohorts:
ALSPAC
B58C
BAMSE (wave1)
BAMSE (wave2)
CAMP
CATSS
CHOP
COPSAC2000
COPSAC2010
DANFUND
DNBC
ECRHS
EstonianBB
FINNGEN
GENEVA_AFFY
GENEVA_ILLUMINA
GENUFADext (SHIP1)
GENUFAD (SHIP2)
GERA
GINILISAnorth
GINILISAsouth
HEALTH2006
HUNT
INMA
IOW
MAAS
MAS_HNR
MoBa_children
NCRC_ADC
NFBC66
NTR
PIAMA
Raine study
SALTY
SAPALDIA_new
SAPALDIA_old
SWS
TwinGene
TwinsUK
UKBB
Meta−analysis (I^2 = 0.73% [0.00%; 37.86%])
Multi−ancestry cohorts:
GALA
GENR
RIKEN
SAGE
SAPPHIRE
Replication in 23andme:
European
African
Latino
0.71
1.0
1.41
2.0
Odds Ratio

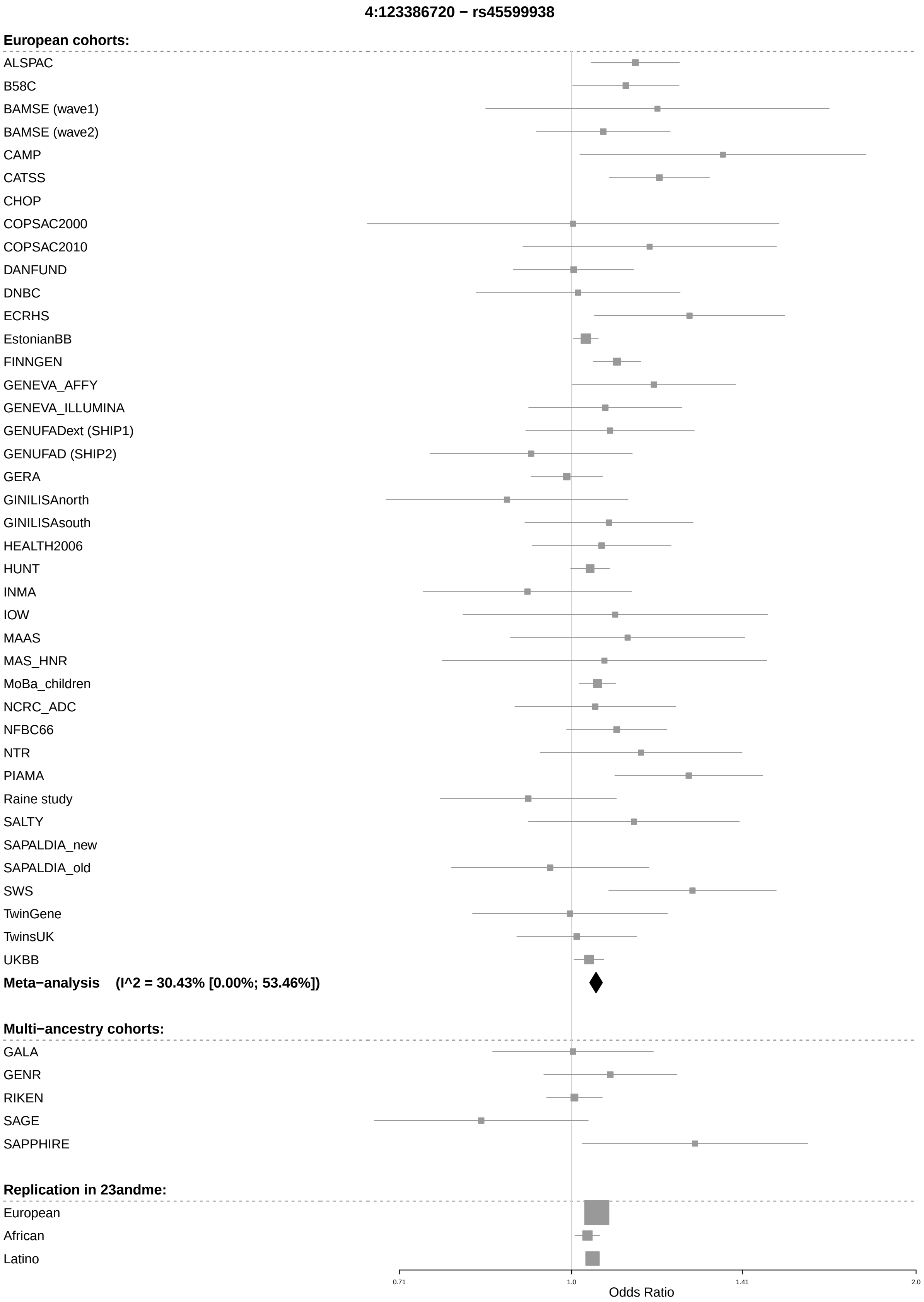

4:123386720 − rs45599938
European cohorts:
ALSPAC
B58C
BAMSE (wave1)
BAMSE (wave2)
CAMP
CATSS
CHOP
COPSAC2000
COPSAC2010
DANFUND
DNBC
ECRHS
EstonianBB
FINNGEN
GENEVA_AFFY
GENEVA_ILLUMINA
GENUFADext (SHIP1)
GENUFAD (SHIP2)
GERA
GINILISAnorth
GINILISAsouth
HEALTH2006
HUNT
INMA
IOW
MAAS
MAS_HNR
MoBa_children
NCRC_ADC
NFBC66
NTR
PIAMA
Raine study
SALTY
SAPALDIA_new
SAPALDIA_old
SWS
TwinGene
TwinsUK
UKBB
Meta−analysis (I^2 = 30.43% [0.00%; 53.46%])
Multi−ancestry cohorts:
GALA
GENR
RIKEN
SAGE
SAPPHIRE
Replication in 23andme:
European
African
Latino
0.71
1.0
1.41
2.0
Odds Ratio

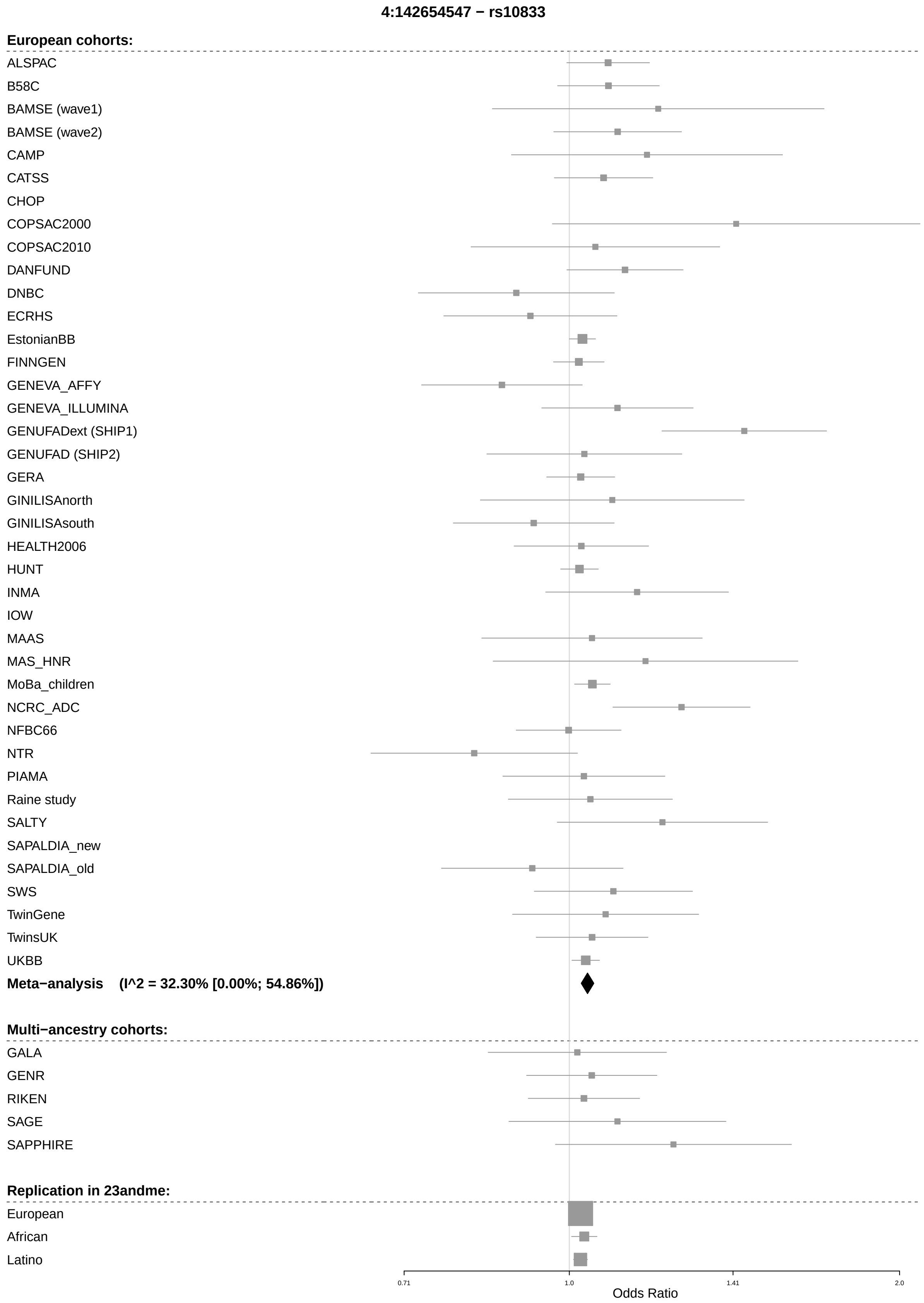
4:142654547 − rs10833
European cohorts:
ALSPAC
B58C
BAMSE (wave1)
BAMSE (wave2)
CAMP
CATSS
CHOP
COPSAC2000
COPSAC2010
DANFUND
DNBC
ECRHS
EstonianBB
FINNGEN
GENEVA_AFFY
GENEVA_ILLUMINA
GENUFADext (SHIP1)
GENUFAD (SHIP2)
GERA
GINILISAnorth
GINILISAsouth
HEALTH2006
HUNT
INMA
IOW
MAAS
MAS_HNR
MoBa_children
NCRC_ADC
NFBC66
NTR
PIAMA
Raine study
SALTY
SAPALDIA_new
SAPALDIA_old
SWS
TwinGene
TwinsUK
UKBB
Meta−analysis (I^2 = 32.30% [0.00%; 54.86%])
Multi−ancestry cohorts:
GALA
GENR
RIKEN
SAGE
SAPPHIRE
Replication in 23andme:
European
African
Latino
0.71
1.0
1.41
2.0
Odds Ratio

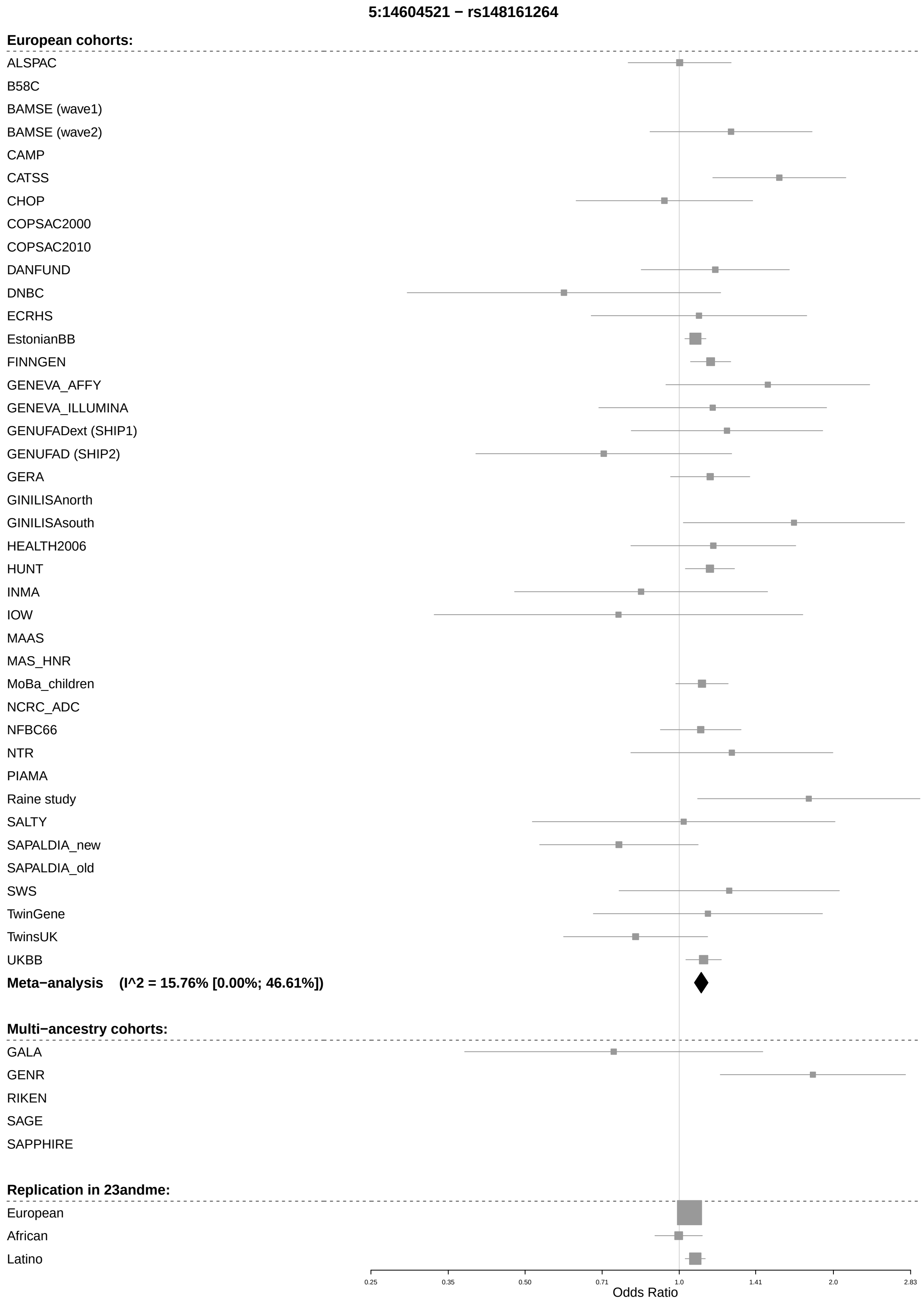
5:14604521 − rs148161264
European cohorts:
ALSPAC
B58C
BAMSE (wave1)
BAMSE (wave2)
CAMP
CATSS
CHOP
COPSAC2000
COPSAC2010
DANFUND
DNBC
ECRHS
EstonianBB
FINNGEN
GENEVA_AFFY
GENEVA_ILLUMINA
GENUFADext (SHIP1)
GENUFAD (SHIP2)
GERA
GINILISAnorth
GINILISAsouth
HEALTH2006
HUNT
INMA
IOW
MAAS
MAS_HNR
MoBa_children
NCRC_ADC
NFBC66
NTR
PIAMA
Raine study
SALTY
SAPALDIA_new
SAPALDIA_old
SWS
TwinGene
TwinsUK
UKBB
Meta−analysis (I^2 = 15.76% [0.00%; 46.61%])
Multi−ancestry cohorts:
GALA
GENR
RIKEN
SAGE
SAPPHIRE
Replication in 23andme:
European
African
Latino
0.25
0.35
0.50
0.71
1.0
1.41
2.0
2.83
Odds Ratio

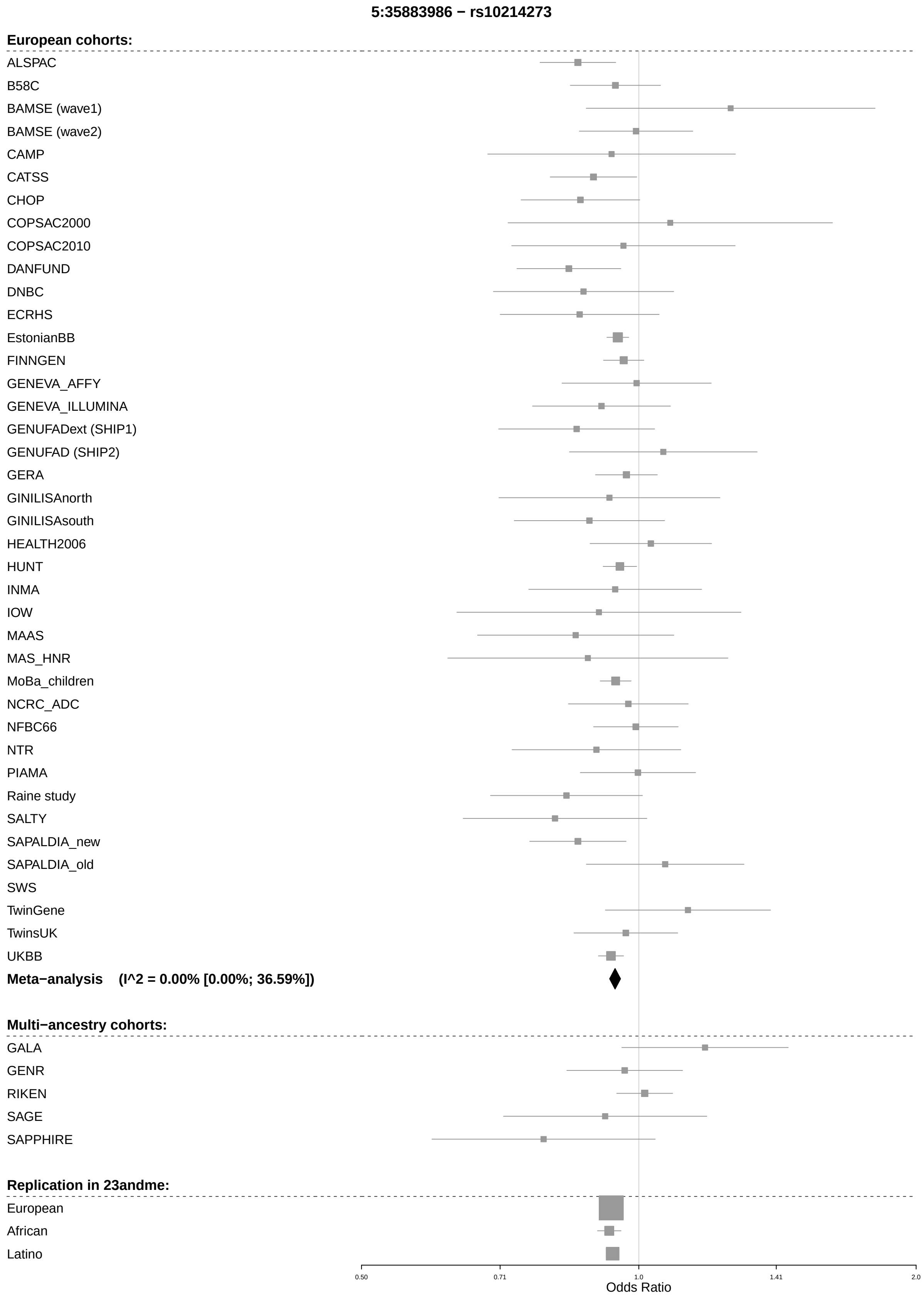
5:35883986 − rs10214273
European cohorts:
ALSPAC
B58C
BAMSE (wave1)
BAMSE (wave2)
CAMP
CATSS
CHOP
COPSAC2000
COPSAC2010
DANFUND
DNBC
ECRHS
EstonianBB
FINNGEN
GENEVA_AFFY
GENEVA_ILLUMINA
GENUFADext (SHIP1)
GENUFAD (SHIP2)
GERA
GINILISAnorth
GINILISAsouth
HEALTH2006
HUNT
INMA
IOW
MAAS
MAS_HNR
MoBa_children
NCRC_ADC
NFBC66
NTR
PIAMA
Raine study
SALTY
SAPALDIA_new
SAPALDIA_old
SWS
TwinGene
TwinsUK
UKBB
Meta−analysis (I^2 = 0.00% [0.00%; 36.59%])
Multi−ancestry cohorts:
GALA
GENR
RIKEN
SAGE
SAPPHIRE
Replication in 23andme:
European
African
Latino
0.50
0.71
1.0
1.41
2.0
Odds Ratio

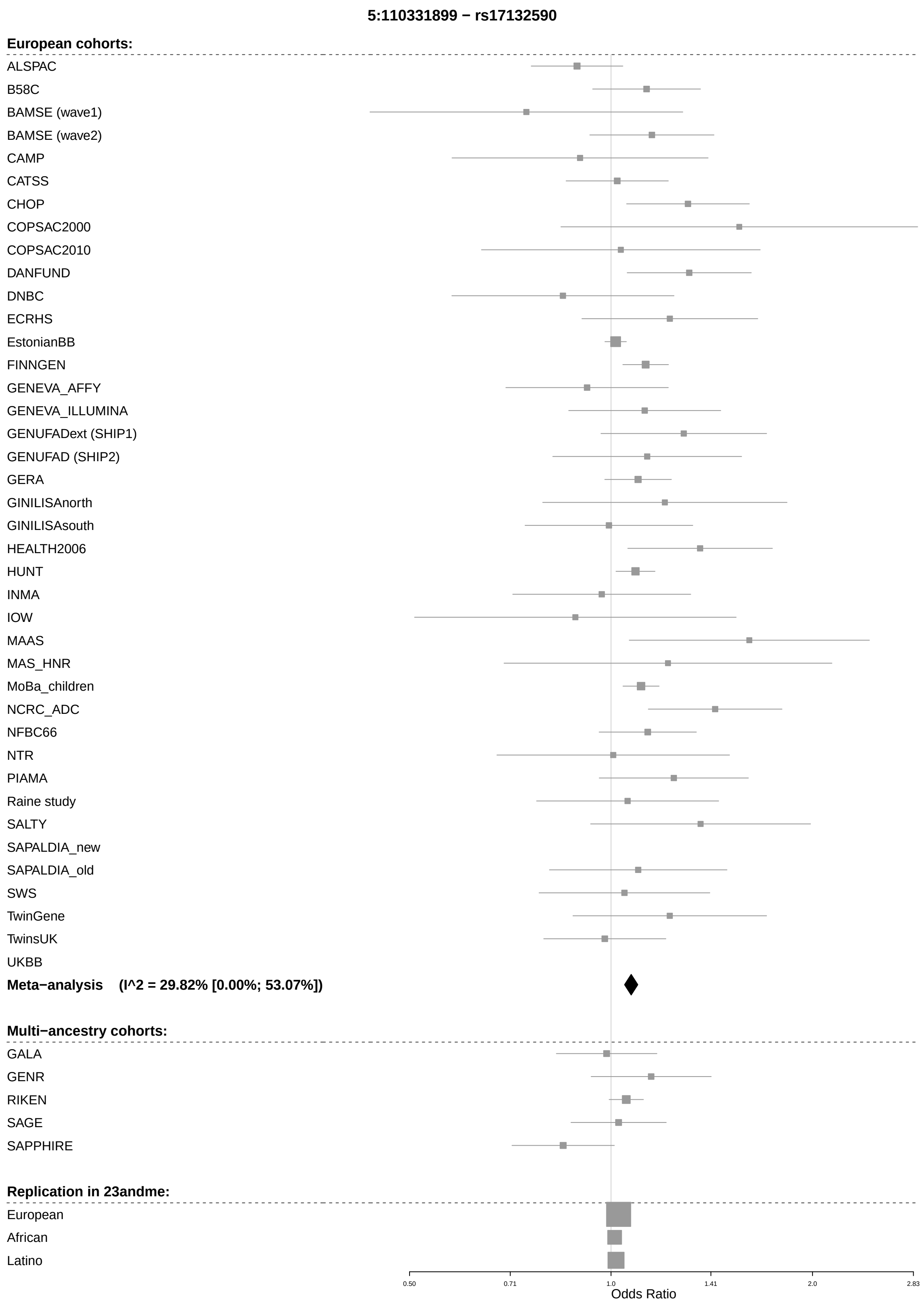
5:110331899 − rs17132590
European cohorts:
ALSPAC
B58C
BAMSE (wave1)
BAMSE (wave2)
CAMP
CATSS
CHOP
COPSAC2000
COPSAC2010
DANFUND
DNBC
ECRHS
EstonianBB
FINNGEN
GENEVA_AFFY
GENEVA_ILLUMINA
GENUFADext (SHIP1)
GENUFAD (SHIP2)
GERA
GINILISAnorth
GINILISAsouth
HEALTH2006
HUNT
INMA
IOW
MAAS
MAS_HNR
MoBa_children
NCRC_ADC
NFBC66
NTR
PIAMA
Raine study
SALTY
SAPALDIA_new
SAPALDIA_old
SWS
TwinGene
TwinsUK
UKBB
Meta−analysis (I^2 = 29.82% [0.00%; 53.07%])
Multi−ancestry cohorts:
GALA
GENR
RIKEN
SAGE
SAPPHIRE
Replication in 23andme:
European
African
Latino
0.50
0.71
1.0
1.41
2.0
2.83
Odds Ratio

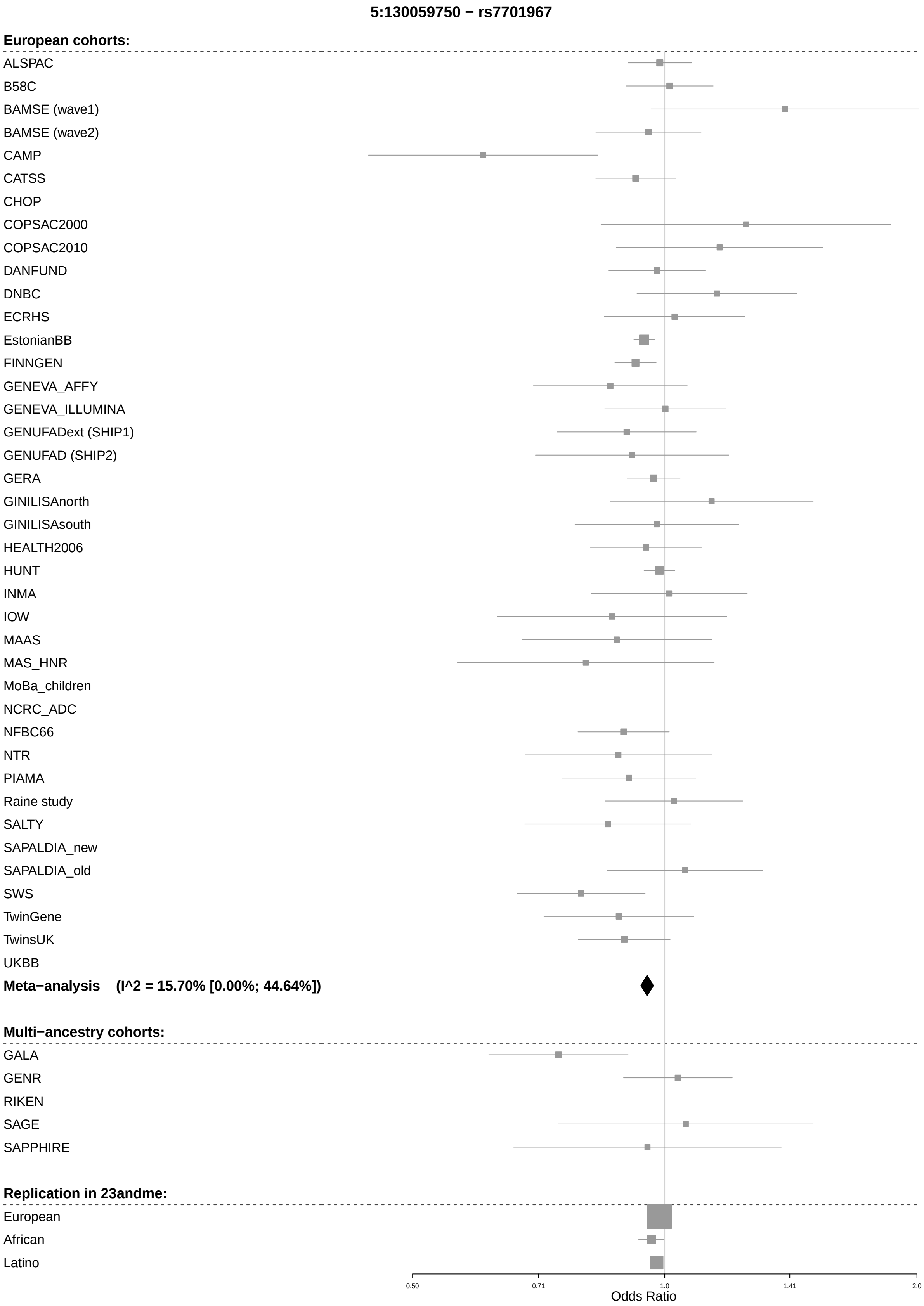
5:130059750 − rs7701967
European cohorts:
ALSPAC
B58C
BAMSE (wave1)
BAMSE (wave2)
CAMP
CATSS
CHOP
COPSAC2000
COPSAC2010
DANFUND
DNBC
ECRHS
EstonianBB
FINNGEN
GENEVA_AFFY
GENEVA_ILLUMINA
GENUFADext (SHIP1)
GENUFAD (SHIP2)
GERA
GINILISAnorth
GINILISAsouth
HEALTH2006
HUNT
INMA
IOW
MAAS
MAS_HNR
MoBa_children
NCRC_ADC
NFBC66
NTR
PIAMA
Raine study
SALTY
SAPALDIA_new
SAPALDIA_old
SWS
TwinGene
TwinsUK
UKBB
Meta−analysis (I^2 = 15.70% [0.00%; 44.64%])
Multi−ancestry cohorts:
GALA
GENR
RIKEN
SAGE
SAPPHIRE
Replication in 23andme:
European
African
Latino
0.50
0.71
1.0
1.41
2.0
Odds Ratio

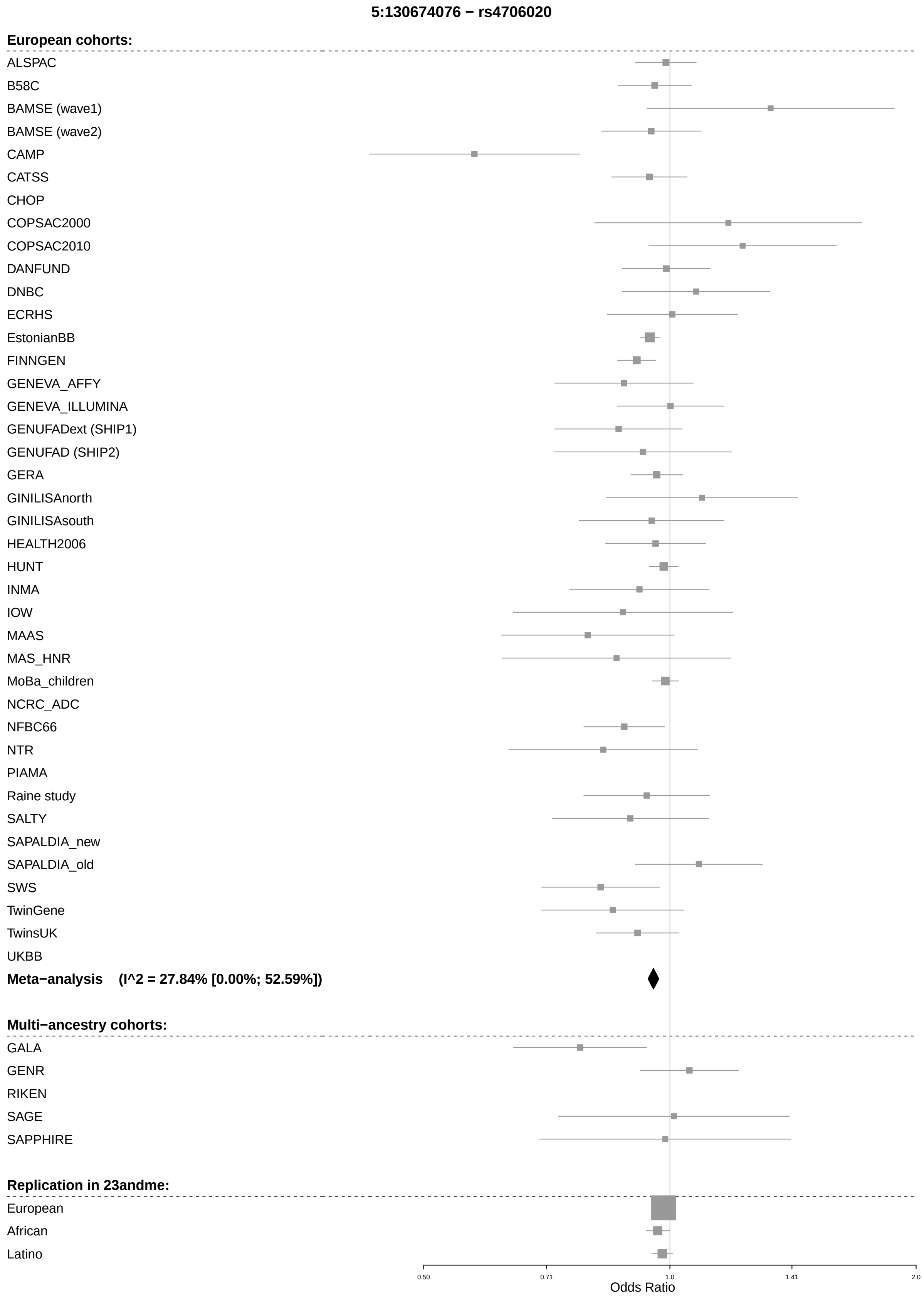
5:130674076 − rs4706020
European cohorts:
ALSPAC
B58C
BAMSE (wave1)
BAMSE (wave2)
CAMP
CATSS
CHOP
COPSAC2000
COPSAC2010
DANFUND
DNBC
ECRHS
EstonianBB
FINNGEN
GENEVA_AFFY
GENEVA_ILLUMINA
GENUFADext (SHIP1)
GENUFAD (SHIP2)
GERA
GINILISAnorth
GINILISAsouth
HEALTH2006
HUNT
INMA
IOW
MAAS
MAS_HNR
MoBa_children
NCRC_ADC
NFBC66
NTR
PIAMA
Raine study
SALTY
SAPALDIA_new
SAPALDIA_old
SWS
TwinGene
TwinsUK
UKBB
Meta−analysis (I^2 = 27.84% [0.00%; 52.59%])
Multi−ancestry cohorts:
GALA
GENR
RIKEN
SAGE
SAPPHIRE
Replication in 23andme:
European
African
Latino
0.50
0.71
1.0
1.41
2.0
Odds Ratio

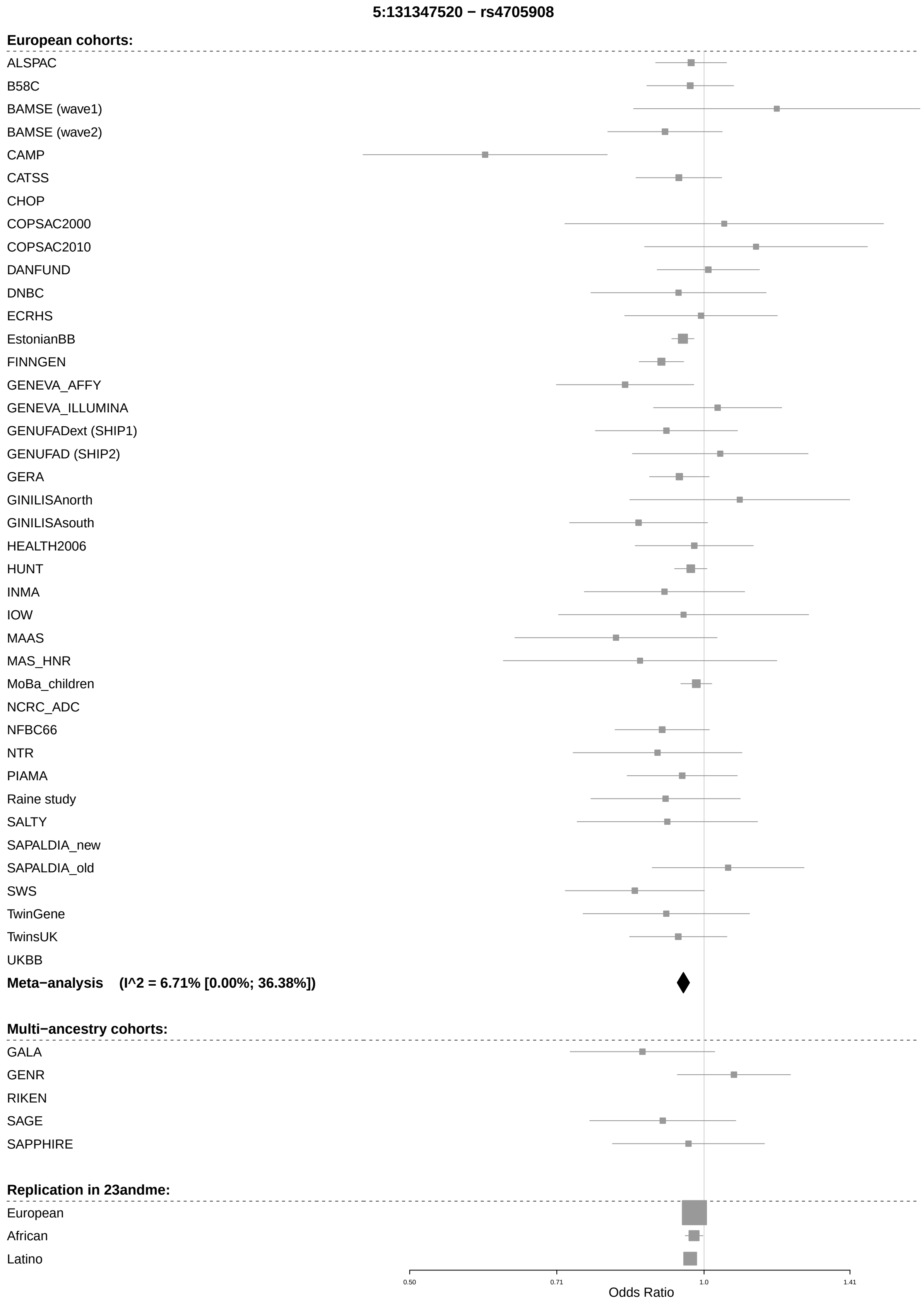
5:131347520 − rs4705908
European cohorts:
ALSPAC
B58C
BAMSE (wave1)
BAMSE (wave2)
CAMP
CATSS
CHOP
COPSAC2000
COPSAC2010
DANFUND
DNBC
ECRHS
EstonianBB
FINNGEN
GENEVA_AFFY
GENEVA_ILLUMINA
GENUFADext (SHIP1)
GENUFAD (SHIP2)
GERA
GINILISAnorth
GINILISAsouth
HEALTH2006
HUNT
INMA
IOW
MAAS
MAS_HNR
MoBa_children
NCRC_ADC
NFBC66
NTR
PIAMA
Raine study
SALTY
SAPALDIA_new
SAPALDIA_old
SWS
TwinGene
TwinsUK
UKBB
Meta−analysis (I^2 = 6.71% [0.00%; 36.38%])
Multi−ancestry cohorts:
GALA
GENR
RIKEN
SAGE
SAPPHIRE
Replication in 23andme:
European
African
Latino
0.50
0.71
1.0
1.41
Odds Ratio

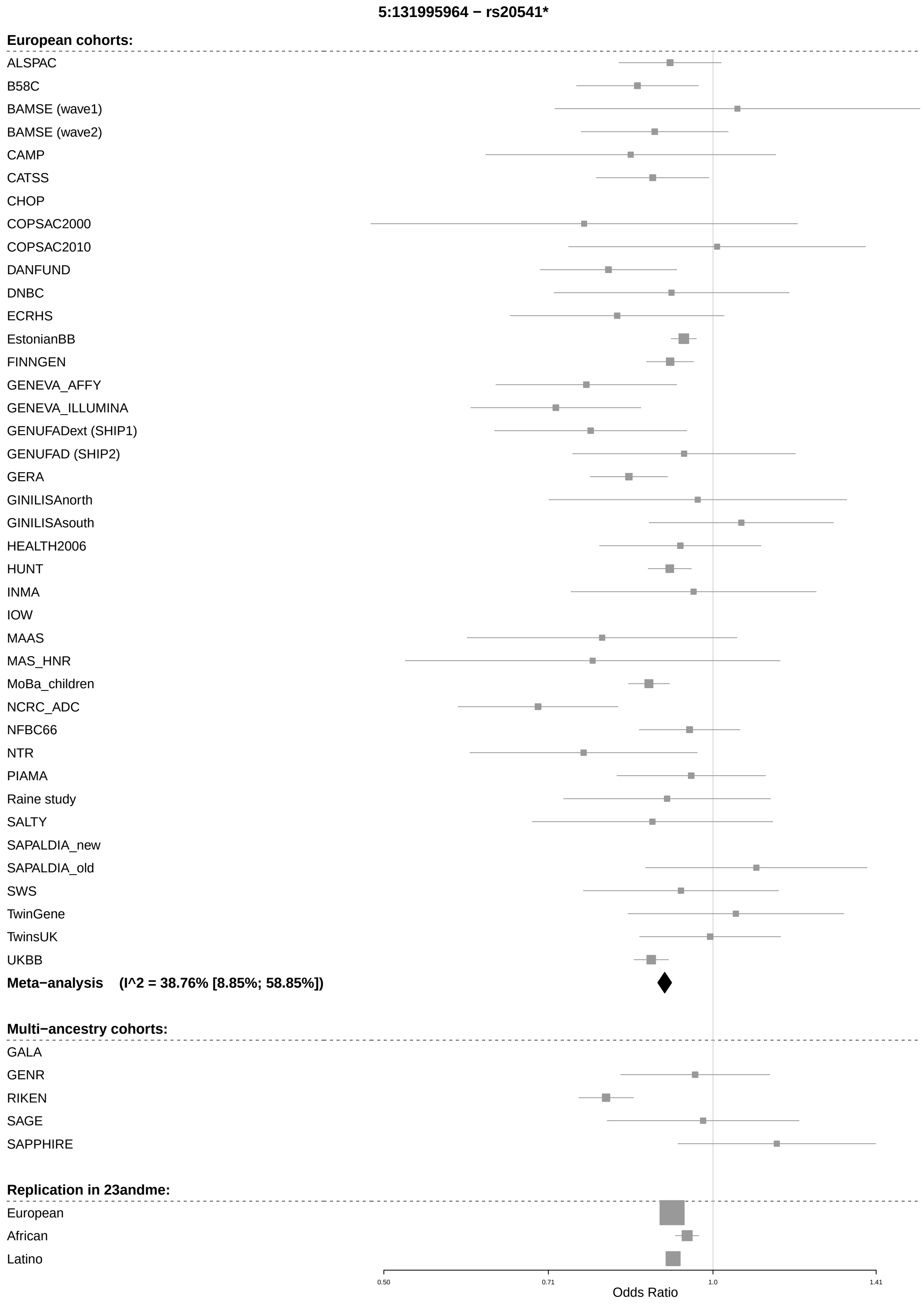
5:131995964 − rs20541*
European cohorts:
ALSPAC
B58C
BAMSE (wave1)
BAMSE (wave2)
CAMP
CATSS
CHOP
COPSAC2000
COPSAC2010
DANFUND
DNBC
ECRHS
EstonianBB
FINNGEN
GENEVA_AFFY
GENEVA_ILLUMINA
GENUFADext (SHIP1)
GENUFAD (SHIP2)
GERA
GINILISAnorth
GINILISAsouth
HEALTH2006
HUNT
INMA
IOW
MAAS
MAS_HNR
MoBa_children
NCRC_ADC
NFBC66
NTR
PIAMA
Raine study
SALTY
SAPALDIA_new
SAPALDIA_old
SWS
TwinGene
TwinsUK
UKBB
Meta−analysis (I^2 = 38.76% [8.85%; 58.85%])
Multi−ancestry cohorts:
GALA
GENR
RIKEN
SAGE
SAPPHIRE
Replication in 23andme:
European
African
Latino
0.50
0.71
1.0
1.41
Odds Ratio

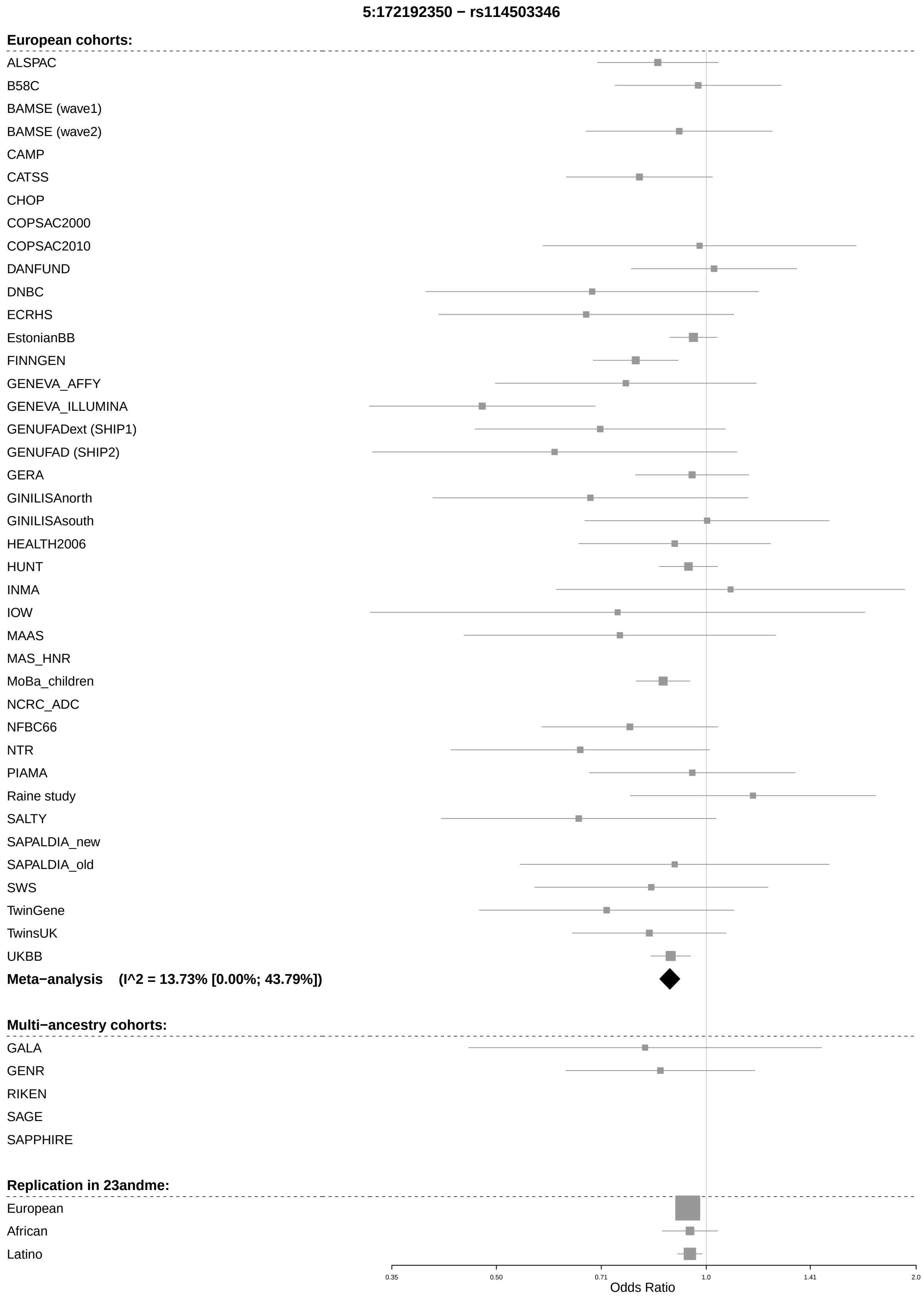
5:172192350 − rs114503346
European cohorts:
ALSPAC
B58C
BAMSE (wave1)
BAMSE (wave2)
CAMP
CATSS
CHOP
COPSAC2000
COPSAC2010
DANFUND
DNBC
ECRHS
EstonianBB
FINNGEN
GENEVA_AFFY
GENEVA_ILLUMINA
GENUFADext (SHIP1)
GENUFAD (SHIP2)
GERA
GINILISAnorth
GINILISAsouth
HEALTH2006
HUNT
INMA
IOW
MAAS
MAS_HNR
MoBa_children
NCRC_ADC
NFBC66
NTR
PIAMA
Raine study
SALTY
SAPALDIA_new
SAPALDIA_old
SWS
TwinGene
TwinsUK
UKBB
Meta−analysis (I^2 = 13.73% [0.00%; 43.79%])
Multi−ancestry cohorts:
GALA
GENR
RIKEN
SAGE
SAPPHIRE
Replication in 23andme:
European
African
Latino
0.35
0.50
0.71
1.0
1.41
2.0
Odds Ratio

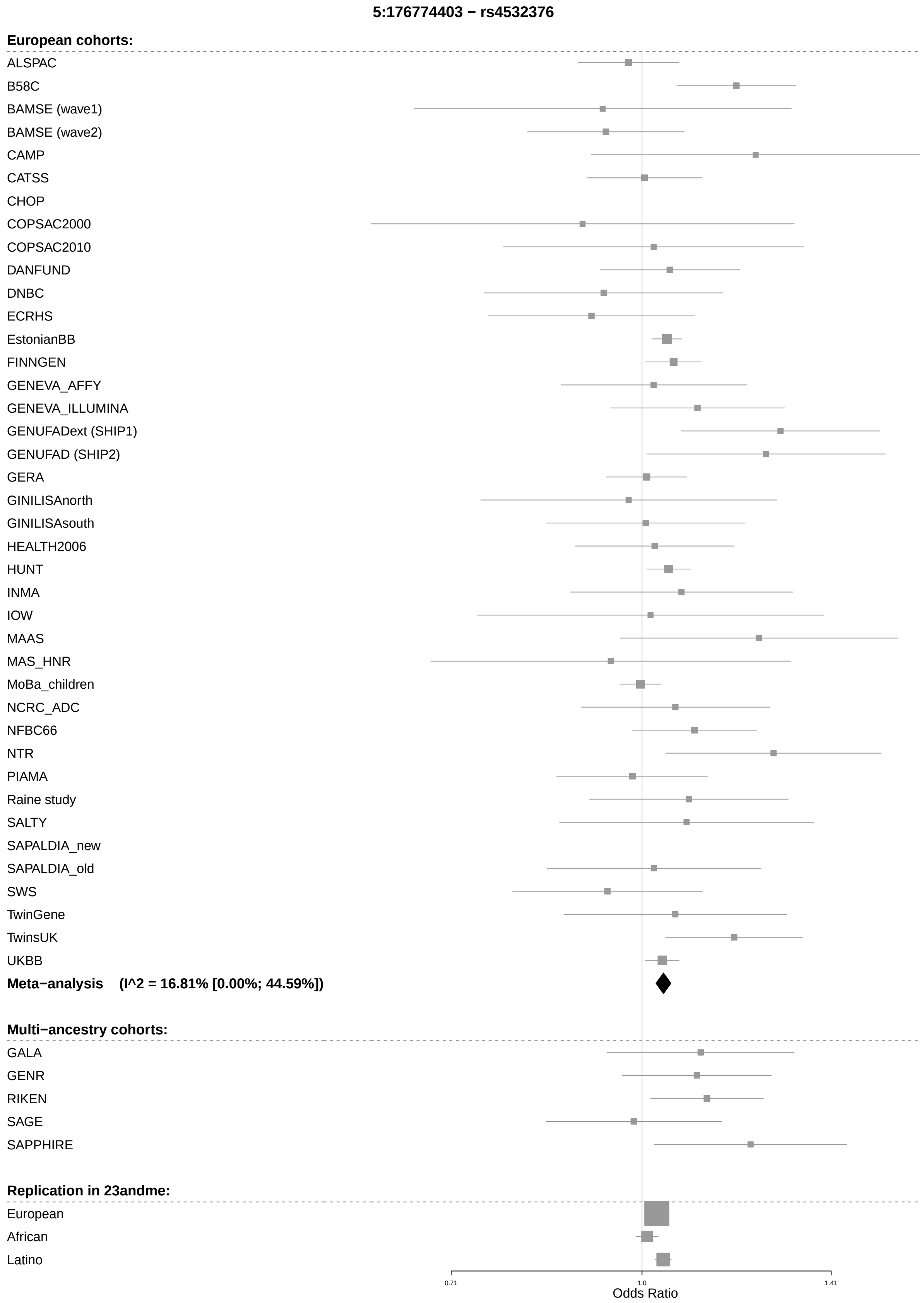
5:176774403 − rs4532376
European cohorts:
ALSPAC
B58C
BAMSE (wave1)
BAMSE (wave2)
CAMP
CATSS
CHOP
COPSAC2000
COPSAC2010
DANFUND
DNBC
ECRHS
EstonianBB
FINNGEN
GENEVA_AFFY
GENEVA_ILLUMINA
GENUFADext (SHIP1)
GENUFAD (SHIP2)
GERA
GINILISAnorth
GINILISAsouth
HEALTH2006
HUNT
INMA
IOW
MAAS
MAS_HNR
MoBa_children
NCRC_ADC
NFBC66
NTR
PIAMA
Raine study
SALTY
SAPALDIA_new
SAPALDIA_old
SWS
TwinGene
TwinsUK
UKBB
Meta−analysis (I^2 = 16.81% [0.00%; 44.59%])
Multi−ancestry cohorts:
GALA
GENR
RIKEN
SAGE
SAPPHIRE
Replication in 23andme:
European
African
Latino
0.71
1.0
1.41
Odds Ratio

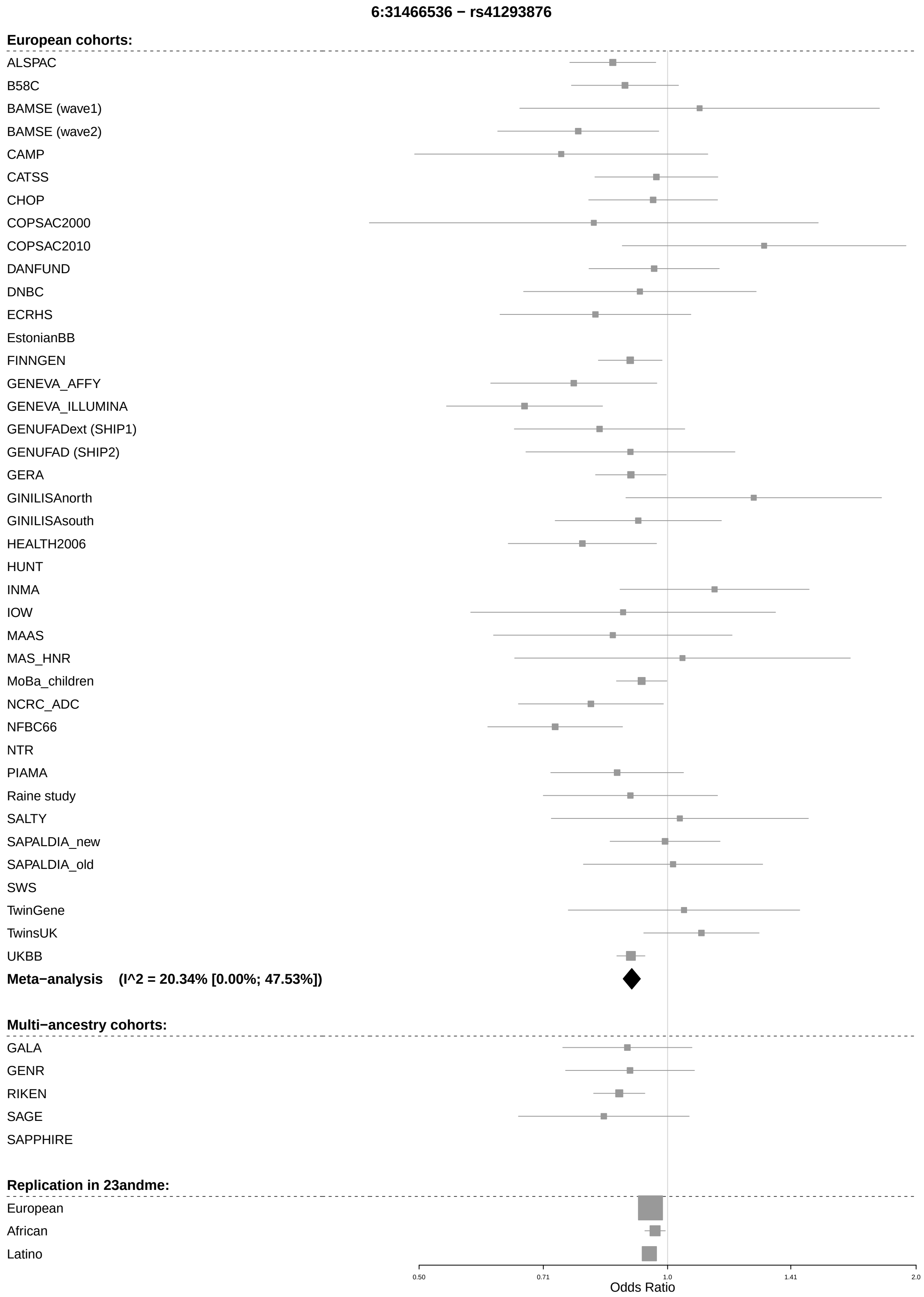

6:31466536 − rs41293876
European cohorts:
ALSPAC
B58C
BAMSE (wave1)
BAMSE (wave2)
CAMP
CATSS
CHOP
COPSAC2000
COPSAC2010
DANFUND
DNBC
ECRHS
EstonianBB
FINNGEN
GENEVA_AFFY
GENEVA_ILLUMINA
GENUFADext (SHIP1)
GENUFAD (SHIP2)
GERA
GINILISAnorth
GINILISAsouth
HEALTH2006
HUNT
INMA
IOW
MAAS
MAS_HNR
MoBa_children
NCRC_ADC
NFBC66
NTR
PIAMA
Raine study
SALTY
SAPALDIA_new
SAPALDIA_old
SWS
TwinGene
TwinsUK
UKBB
Meta−analysis (I^2 = 20.34% [0.00%; 47.53%])
Multi−ancestry cohorts:
GALA
GENR
RIKEN
SAGE
SAPPHIRE
Replication in 23andme:
European
African
Latino
0.50
0.71
1.0
1.41
2.0
Odds Ratio

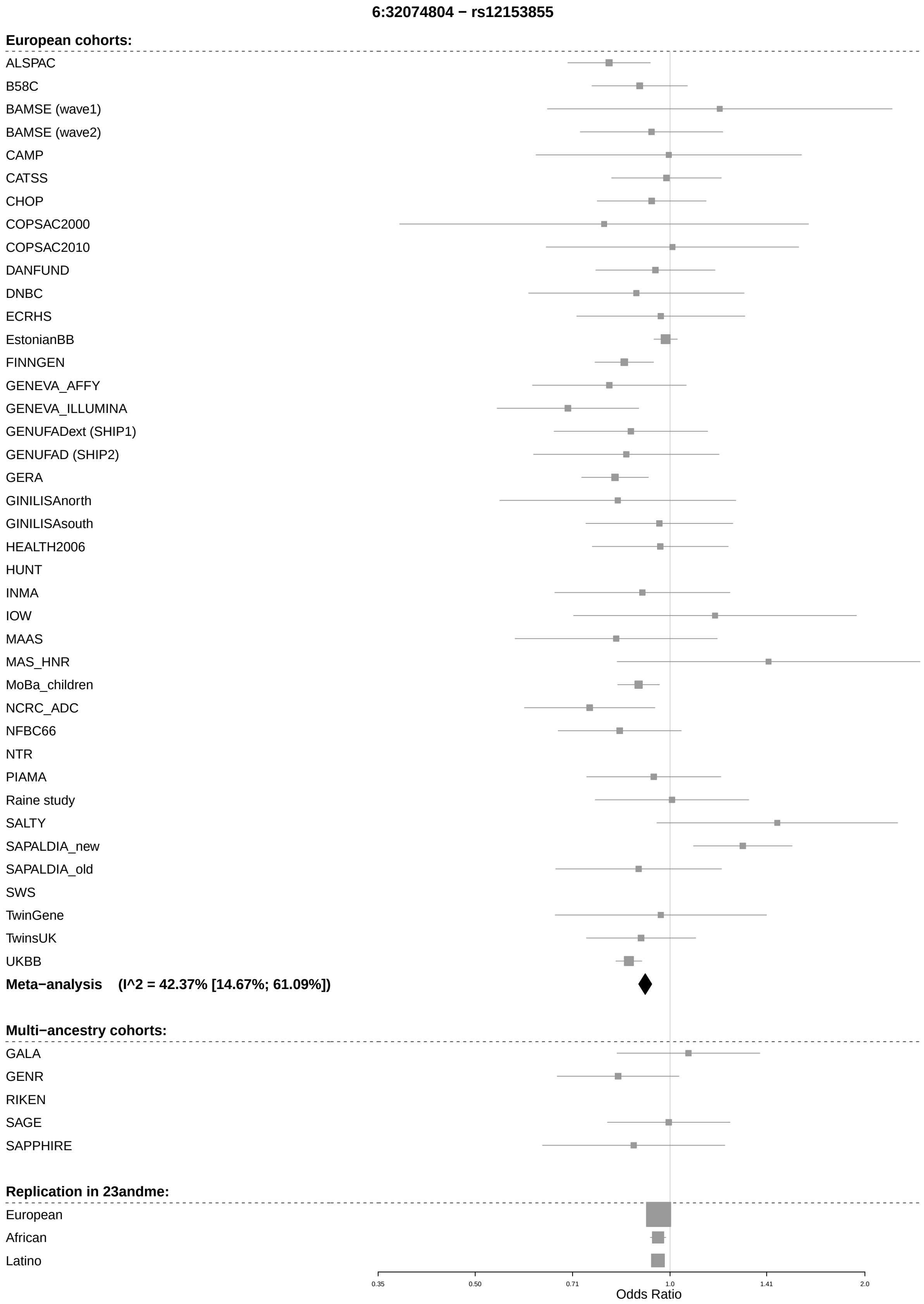
6:32074804 − rs12153855
European cohorts:
ALSPAC
B58C
BAMSE (wave1)
BAMSE (wave2)
CAMP
CATSS
CHOP
COPSAC2000
COPSAC2010
DANFUND
DNBC
ECRHS
EstonianBB
FINNGEN
GENEVA_AFFY
GENEVA_ILLUMINA
GENUFADext (SHIP1)
GENUFAD (SHIP2)
GERA
GINILISAnorth
GINILISAsouth
HEALTH2006
HUNT
INMA
IOW
MAAS
MAS_HNR
MoBa_children
NCRC_ADC
NFBC66
NTR
PIAMA
Raine study
SALTY
SAPALDIA_new
SAPALDIA_old
SWS
TwinGene
TwinsUK
UKBB
Meta−analysis (I^2 = 42.37% [14.67%; 61.09%])
Multi−ancestry cohorts:
GALA
GENR
RIKEN
SAGE
SAPPHIRE
Replication in 23andme:
European
African
Latino
0.35
0.50
0.71
1.0
1.41
2.0
Odds Ratio

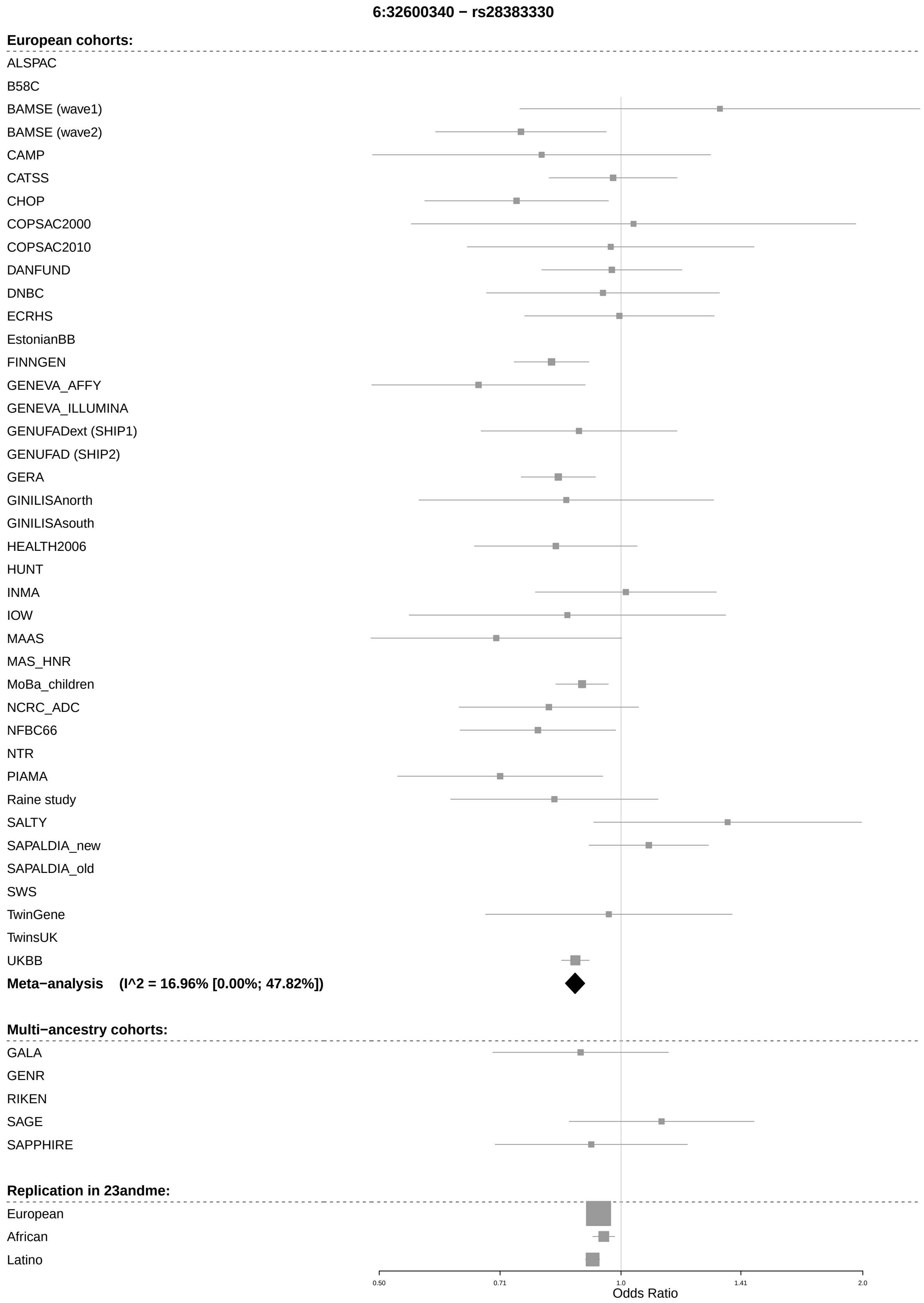
6:32600340 − rs28383330
European cohorts:
ALSPAC
B58C
BAMSE (wave1)
BAMSE (wave2)
CAMP
CATSS
CHOP
COPSAC2000
COPSAC2010
DANFUND
DNBC
ECRHS
EstonianBB
FINNGEN
GENEVA_AFFY
GENEVA_ILLUMINA
GENUFADext (SHIP1)
GENUFAD (SHIP2)
GERA
GINILISAnorth
GINILISAsouth
HEALTH2006
HUNT
INMA
IOW
MAAS
MAS_HNR
MoBa_children
NCRC_ADC
NFBC66
NTR
PIAMA
Raine study
SALTY
SAPALDIA_new
SAPALDIA_old
SWS
TwinGene
TwinsUK
UKBB
Meta−analysis (I^2 = 16.96% [0.00%; 47.82%])
Multi−ancestry cohorts:
GALA
GENR
RIKEN
SAGE
SAPPHIRE
Replication in 23andme:
European
African
Latino
0.50
0.71
1.0
1.41
2.0
Odds Ratio

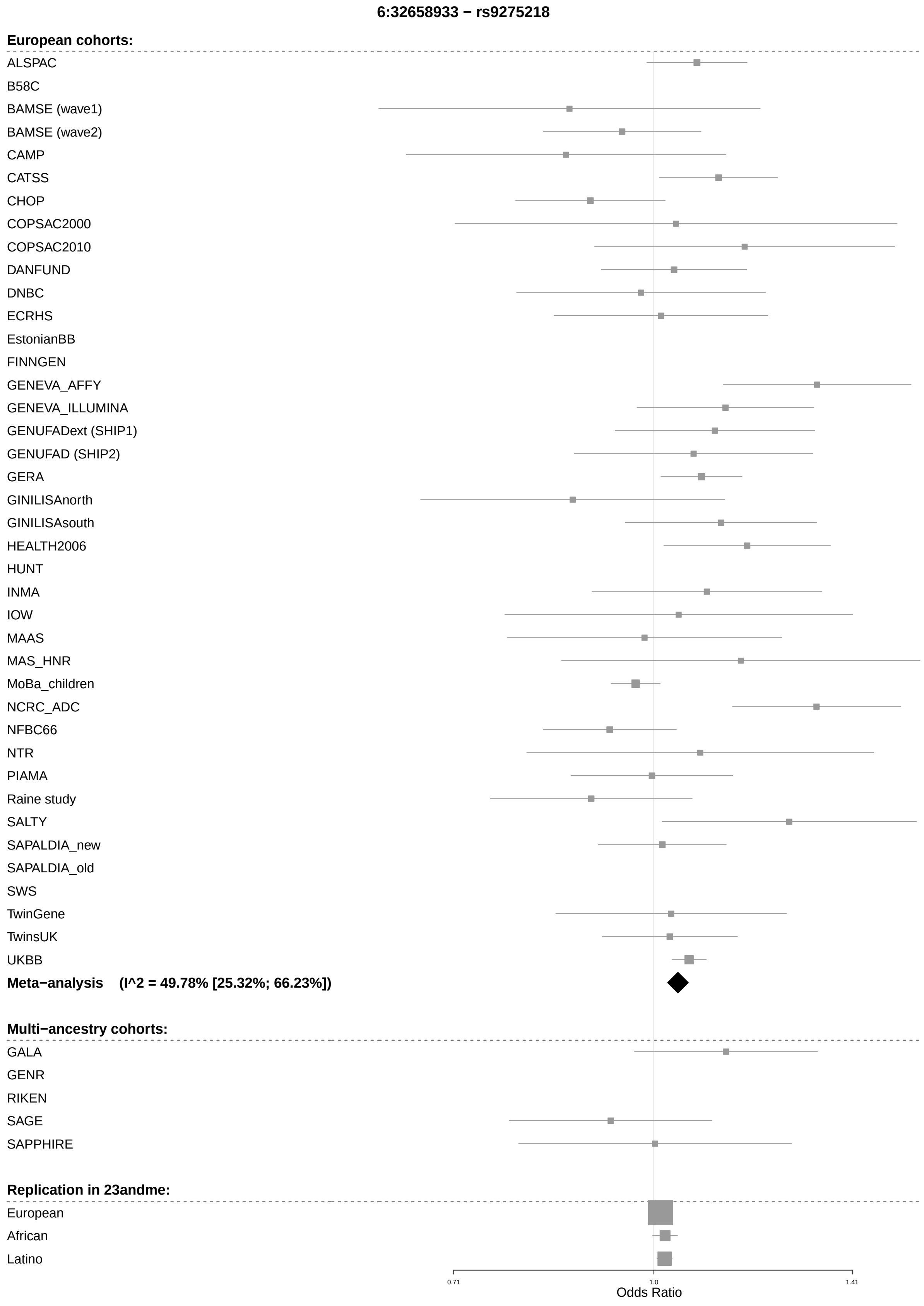
6:32658933 − rs9275218
European cohorts:
ALSPAC
B58C
BAMSE (wave1)
BAMSE (wave2)
CAMP
CATSS
CHOP
COPSAC2000
COPSAC2010
DANFUND
DNBC
ECRHS
EstonianBB
FINNGEN
GENEVA_AFFY
GENEVA_ILLUMINA
GENUFADext (SHIP1)
GENUFAD (SHIP2)
GERA
GINILISAnorth
GINILISAsouth
HEALTH2006
HUNT
INMA
IOW
MAAS
MAS_HNR
MoBa_children
NCRC_ADC
NFBC66
NTR
PIAMA
Raine study
SALTY
SAPALDIA_new
SAPALDIA_old
SWS
TwinGene
TwinsUK
UKBB
Meta−analysis (I^2 = 49.78% [25.32%; 66.23%])
Multi−ancestry cohorts:
GALA
GENR
RIKEN
SAGE
SAPPHIRE
Replication in 23andme:
European
African
Latino
0.71
1.0
1.41
Odds Ratio

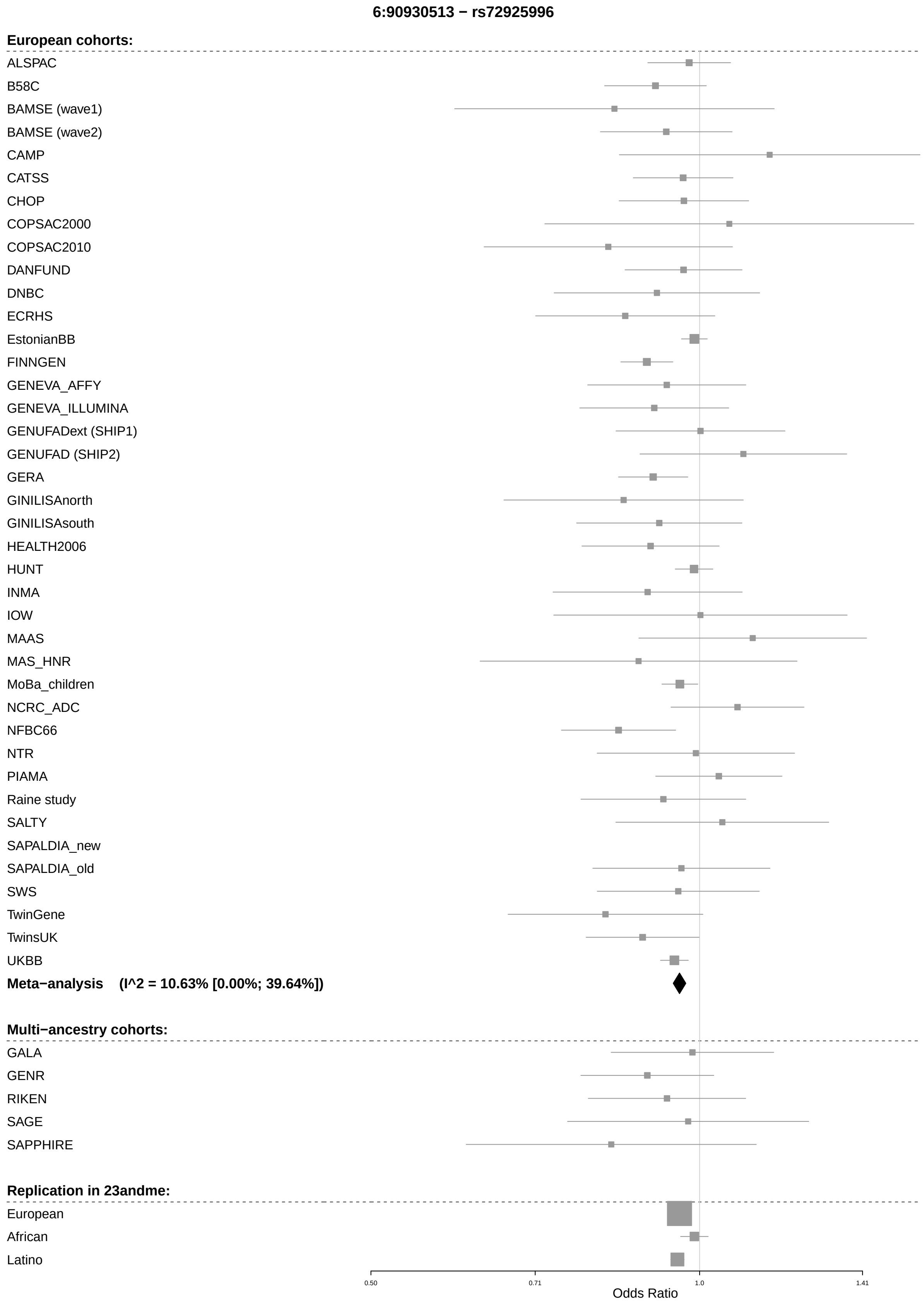
6:90930513 − rs72925996
European cohorts:
ALSPAC
B58C
BAMSE (wave1)
BAMSE (wave2)
CAMP
CATSS
CHOP
COPSAC2000
COPSAC2010
DANFUND
DNBC
ECRHS
EstonianBB
FINNGEN
GENEVA_AFFY
GENEVA_ILLUMINA
GENUFADext (SHIP1)
GENUFAD (SHIP2)
GERA
GINILISAnorth
GINILISAsouth
HEALTH2006
HUNT
INMA
IOW
MAAS
MAS_HNR
MoBa_children
NCRC_ADC
NFBC66
NTR
PIAMA
Raine study
SALTY
SAPALDIA_new
SAPALDIA_old
SWS
TwinGene
TwinsUK
UKBB
Meta−analysis (I^2 = 10.63% [0.00%; 39.64%])
Multi−ancestry cohorts:
GALA
GENR
RIKEN
SAGE
SAPPHIRE
Replication in 23andme:
European
African
Latino
0.50
0.71
1.0
1.41
Odds Ratio

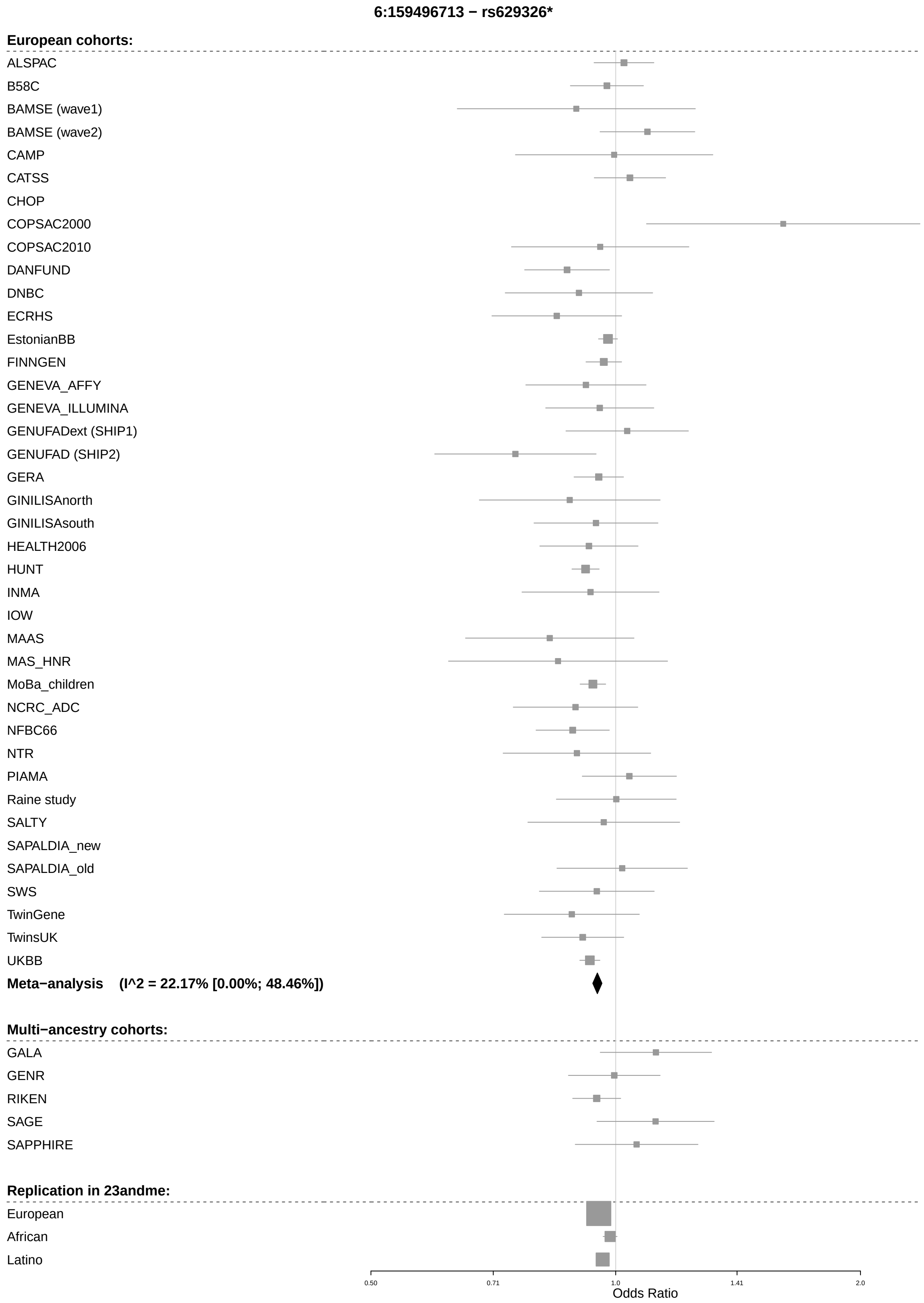
6:159496713 − rs629326*
European cohorts:
ALSPAC
B58C
BAMSE (wave1)
BAMSE (wave2)
CAMP
CATSS
CHOP
COPSAC2000
COPSAC2010
DANFUND
DNBC
ECRHS
EstonianBB
FINNGEN
GENEVA_AFFY
GENEVA_ILLUMINA
GENUFADext (SHIP1)
GENUFAD (SHIP2)
GERA
GINILISAnorth
GINILISAsouth
HEALTH2006
HUNT
INMA
IOW
MAAS
MAS_HNR
MoBa_children
NCRC_ADC
NFBC66
NTR
PIAMA
Raine study
SALTY
SAPALDIA_new
SAPALDIA_old
SWS
TwinGene
TwinsUK
UKBB
Meta−analysis (I^2 = 22.17% [0.00%; 48.46%])
Multi−ancestry cohorts:
GALA
GENR
RIKEN
SAGE
SAPPHIRE
Replication in 23andme:
European
African
Latino
0.50
0.71
1.0
1.41
2.0
Odds Ratio

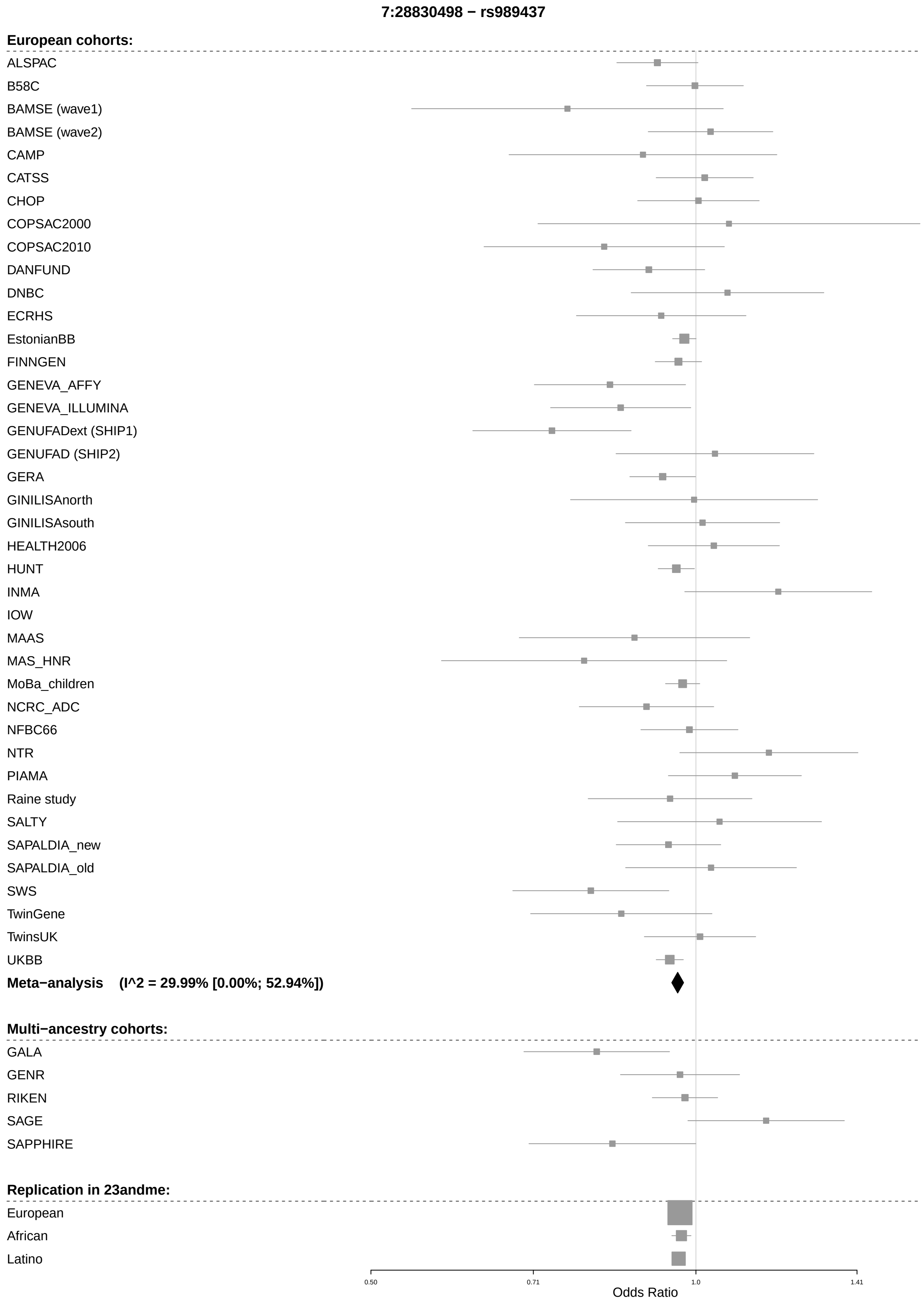

7:28830498 − rs989437
European cohorts:
ALSPAC
B58C
BAMSE (wave1)
BAMSE (wave2)
CAMP
CATSS
CHOP
COPSAC2000
COPSAC2010
DANFUND
DNBC
ECRHS
EstonianBB
FINNGEN
GENEVA_AFFY
GENEVA_ILLUMINA
GENUFADext (SHIP1)
GENUFAD (SHIP2)
GERA
GINILISAnorth
GINILISAsouth
HEALTH2006
HUNT
INMA
IOW
MAAS
MAS_HNR
MoBa_children
NCRC_ADC
NFBC66
NTR
PIAMA
Raine study
SALTY
SAPALDIA_new
SAPALDIA_old
SWS
TwinGene
TwinsUK
UKBB
Meta−analysis (I^2 = 29.99% [0.00%; 52.94%])
Multi−ancestry cohorts:
GALA
GENR
RIKEN
SAGE
SAPPHIRE
Replication in 23andme:
European
African
Latino
0.50
0.71
1.0
1.41
Odds Ratio

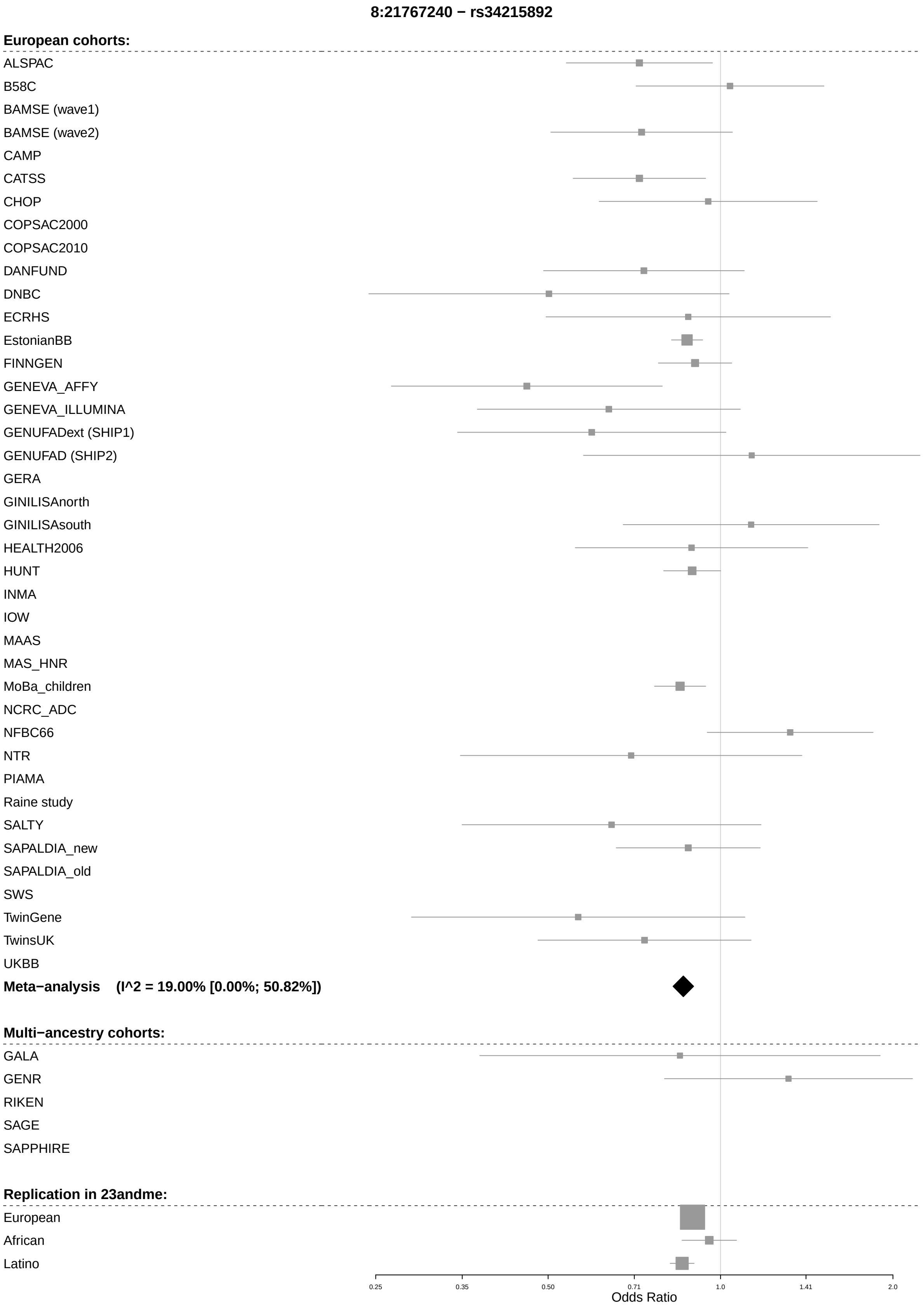

8:21767240 − rs34215892
European cohorts:
ALSPAC
B58C
BAMSE (wave1)
BAMSE (wave2)
CAMP
CATSS
CHOP
COPSAC2000
COPSAC2010
DANFUND
DNBC
ECRHS
EstonianBB
FINNGEN
GENEVA_AFFY
GENEVA_ILLUMINA
GENUFADext (SHIP1)
GENUFAD (SHIP2)
GERA
GINILISAnorth
GINILISAsouth
HEALTH2006
HUNT
INMA
IOW
MAAS
MAS_HNR
MoBa_children
NCRC_ADC
NFBC66
NTR
PIAMA
Raine study
SALTY
SAPALDIA_new
SAPALDIA_old
SWS
TwinGene
TwinsUK
UKBB
Meta−analysis (I^2 = 19.00% [0.00%; 50.82%])
Multi−ancestry cohorts:
GALA
GENR
RIKEN
SAGE
SAPPHIRE
Replication in 23andme:
European
African
Latino
0.25
0.35
0.50
0.71
1.0
1.41
2.0
Odds Ratio

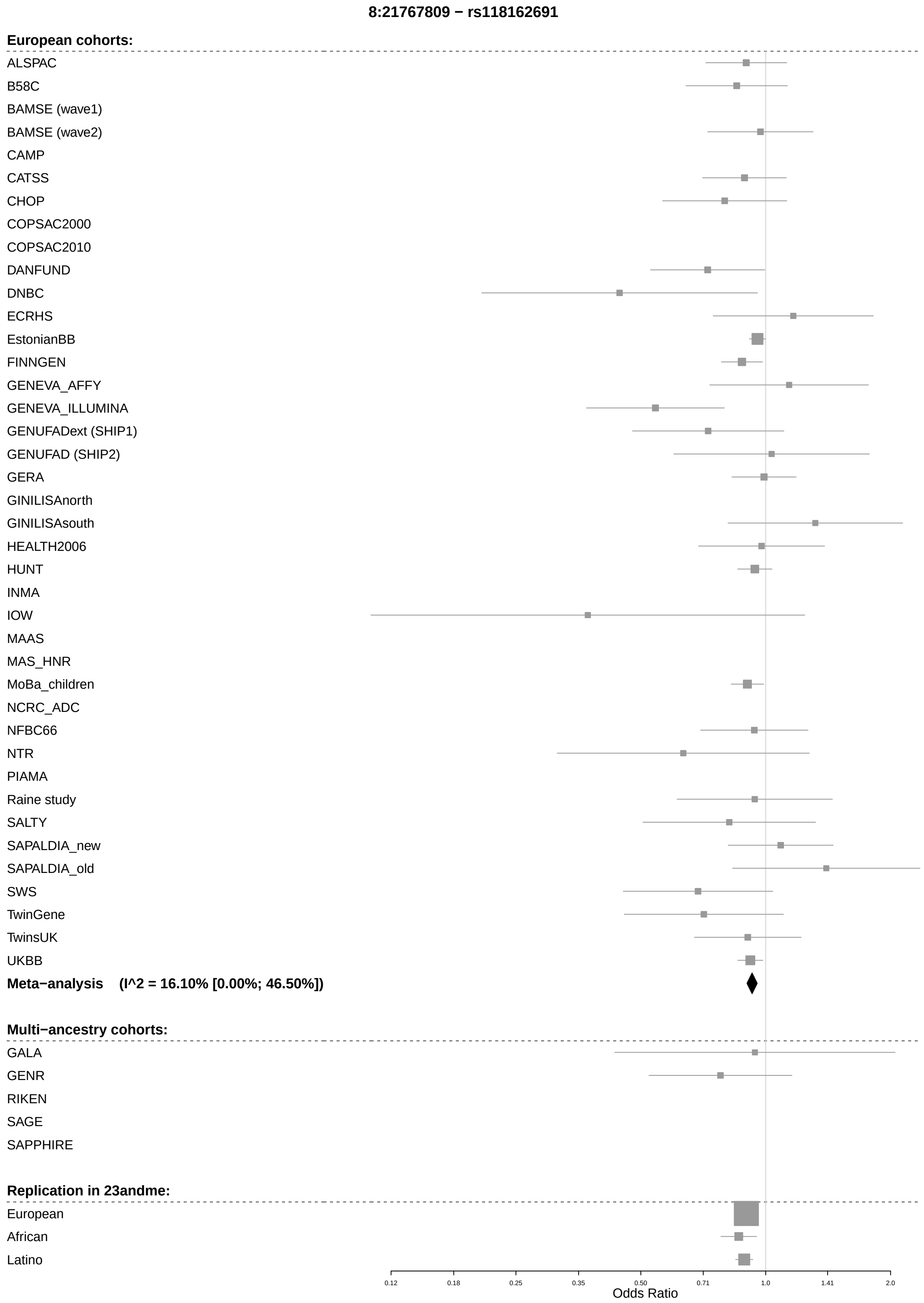
8:21767809 − rs118162691
European cohorts:
ALSPAC
B58C
BAMSE (wave1)
BAMSE (wave2)
CAMP
CATSS
CHOP
COPSAC2000
COPSAC2010
DANFUND
DNBC
ECRHS
EstonianBB
FINNGEN
GENEVA_AFFY
GENEVA_ILLUMINA
GENUFADext (SHIP1)
GENUFAD (SHIP2)
GERA
GINILISAnorth
GINILISAsouth
HEALTH2006
HUNT
INMA
IOW
MAAS
MAS_HNR
MoBa_children
NCRC_ADC
NFBC66
NTR
PIAMA
Raine study
SALTY
SAPALDIA_new
SAPALDIA_old
SWS
TwinGene
TwinsUK
UKBB
Meta−analysis (I^2 = 16.10% [0.00%; 46.50%])
Multi−ancestry cohorts:
GALA
GENR
RIKEN
SAGE
SAPPHIRE
Replication in 23andme:
European
African
Latino
0.12
0.18
0.25
0.35
0.50
0.71
1.0
1.41
2.0
Odds Ratio

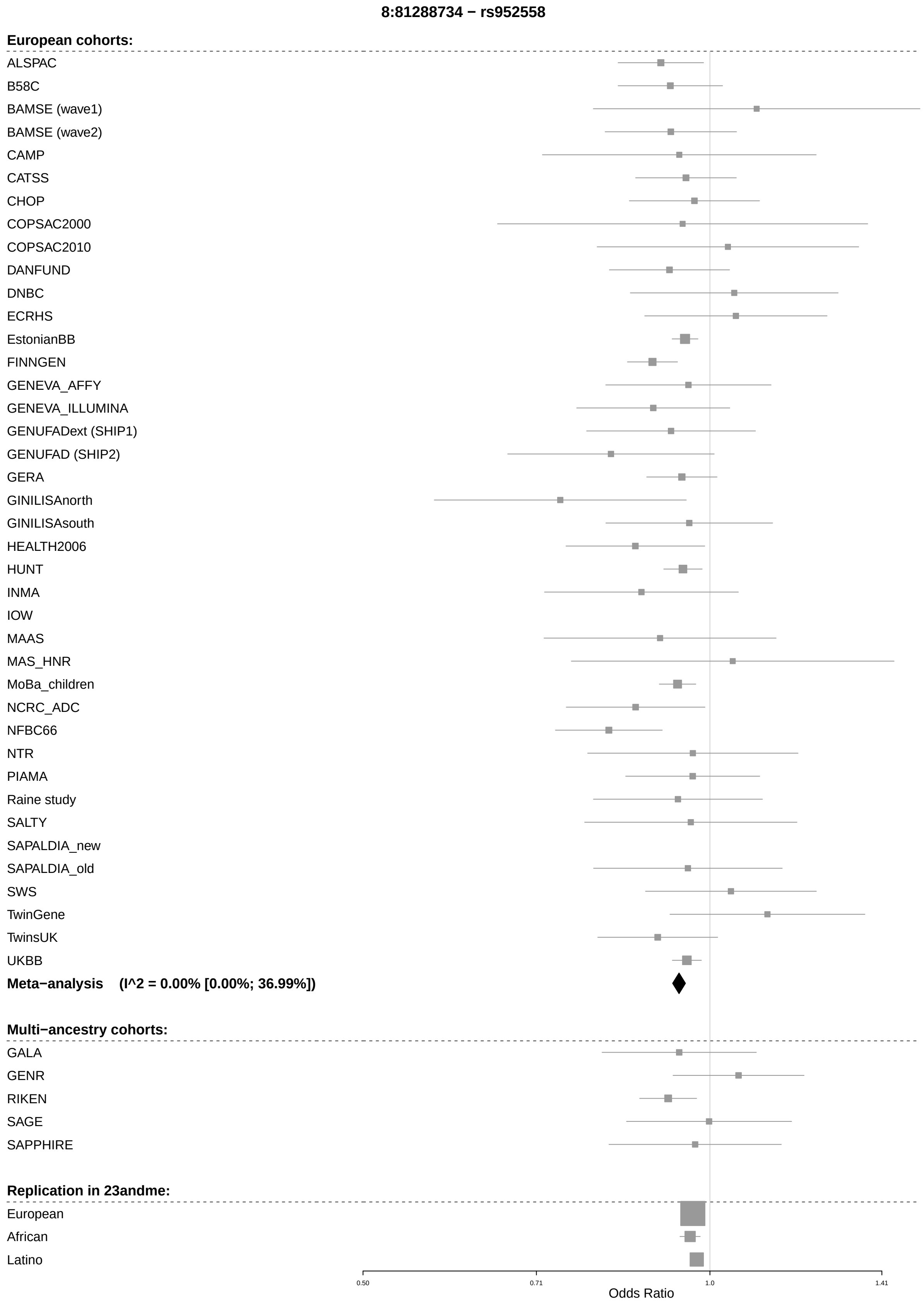
8:81288734 − rs952558
European cohorts:
ALSPAC
B58C
BAMSE (wave1)
BAMSE (wave2)
CAMP
CATSS
CHOP
COPSAC2000
COPSAC2010
DANFUND
DNBC
ECRHS
EstonianBB
FINNGEN
GENEVA_AFFY
GENEVA_ILLUMINA
GENUFADext (SHIP1)
GENUFAD (SHIP2)
GERA
GINILISAnorth
GINILISAsouth
HEALTH2006
HUNT
INMA
IOW
MAAS
MAS_HNR
MoBa_children
NCRC_ADC
NFBC66
NTR
PIAMA
Raine study
SALTY
SAPALDIA_new
SAPALDIA_old
SWS
TwinGene
TwinsUK
UKBB
Meta−analysis (I^2 = 0.00% [0.00%; 36.99%])
Multi−ancestry cohorts:
GALA
GENR
RIKEN
SAGE
SAPPHIRE
Replication in 23andme:
European
African
Latino
0.50
0.71
1.0
1.41
Odds Ratio

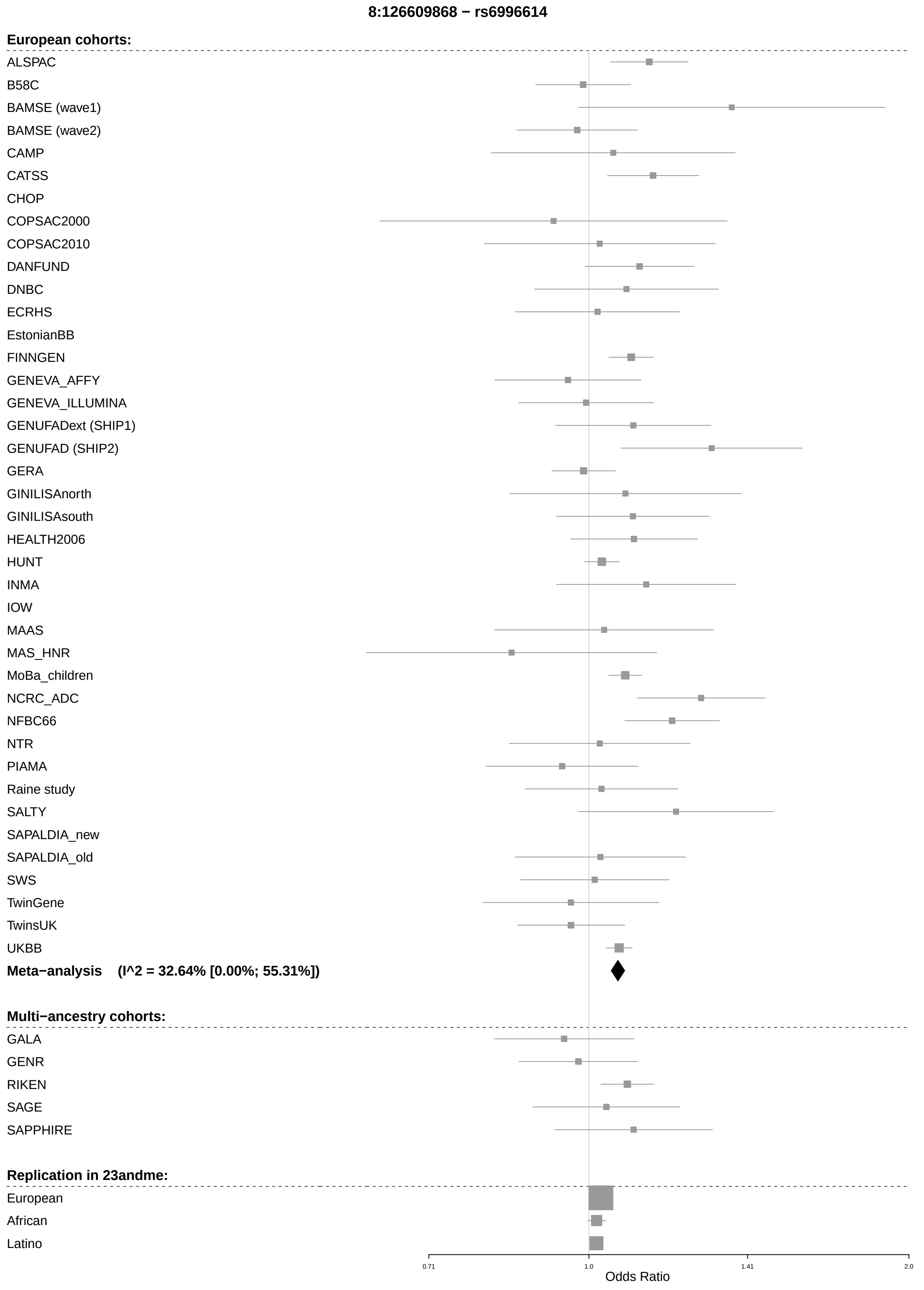
8:126609868 − rs6996614
European cohorts:
ALSPAC
B58C
BAMSE (wave1)
BAMSE (wave2)
CAMP
CATSS
CHOP
COPSAC2000
COPSAC2010
DANFUND
DNBC
ECRHS
EstonianBB
FINNGEN
GENEVA_AFFY
GENEVA_ILLUMINA
GENUFADext (SHIP1)
GENUFAD (SHIP2)
GERA
GINILISAnorth
GINILISAsouth
HEALTH2006
HUNT
INMA
IOW
MAAS
MAS_HNR
MoBa_children
NCRC_ADC
NFBC66
NTR
PIAMA
Raine study
SALTY
SAPALDIA_new
SAPALDIA_old
SWS
TwinGene
TwinsUK
UKBB
Meta−analysis (I^2 = 32.64% [0.00%; 55.31%])
Multi−ancestry cohorts:
GALA
GENR
RIKEN
SAGE
SAPPHIRE
Replication in 23andme:
European
African
Latino
0.71
1.0
1.41
2.0
Odds Ratio

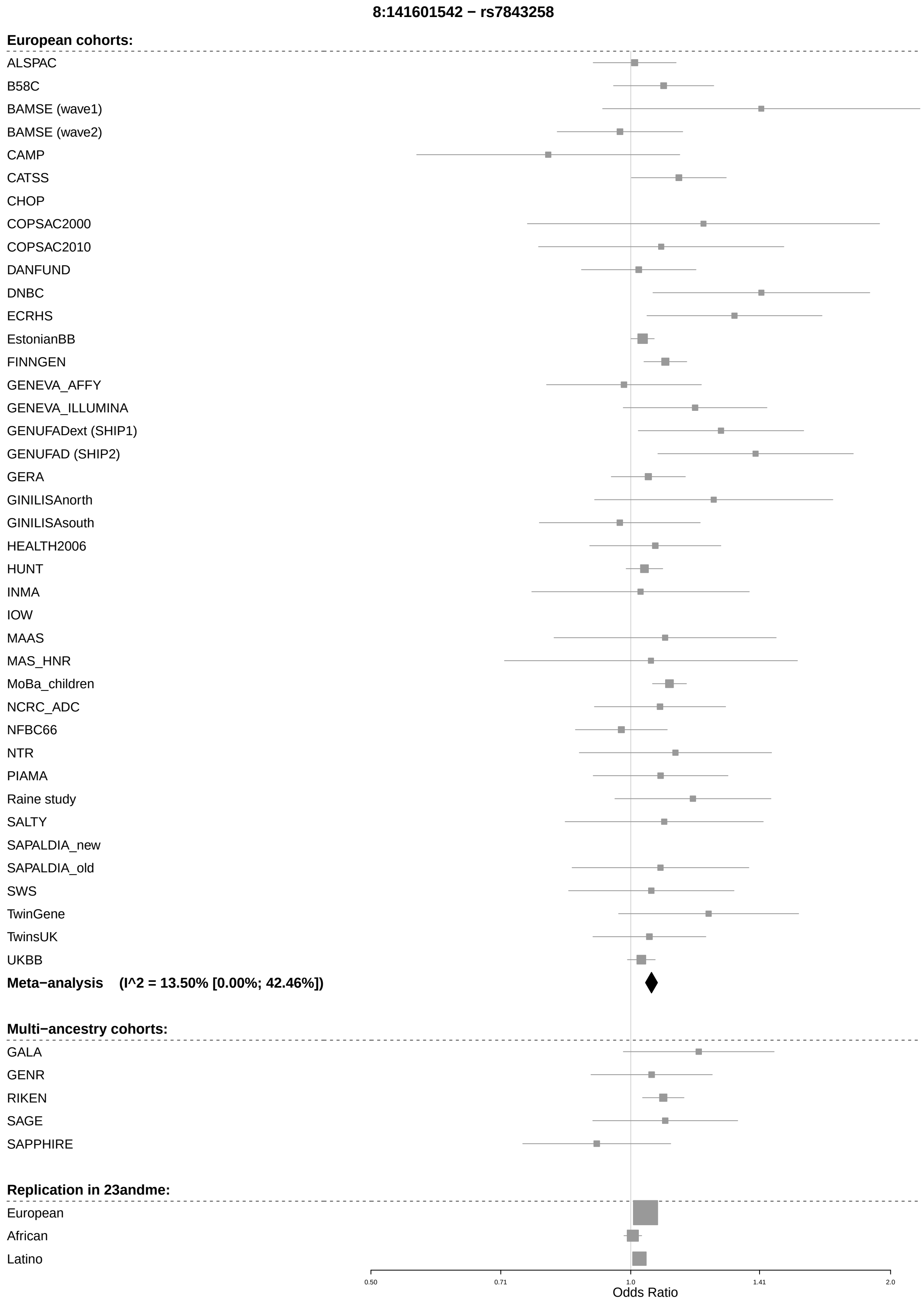
8:141601542 − rs7843258
European cohorts:
ALSPAC
B58C
BAMSE (wave1)
BAMSE (wave2)
CAMP
CATSS
CHOP
COPSAC2000
COPSAC2010
DANFUND
DNBC
ECRHS
EstonianBB
FINNGEN
GENEVA_AFFY
GENEVA_ILLUMINA
GENUFADext (SHIP1)
GENUFAD (SHIP2)
GERA
GINILISAnorth
GINILISAsouth
HEALTH2006
HUNT
INMA
IOW
MAAS
MAS_HNR
MoBa_children
NCRC_ADC
NFBC66
NTR
PIAMA
Raine study
SALTY
SAPALDIA_new
SAPALDIA_old
SWS
TwinGene
TwinsUK
UKBB
Meta−analysis (I^2 = 13.50% [0.00%; 42.46%])
Multi−ancestry cohorts:
GALA
GENR
RIKEN
SAGE
SAPPHIRE
Replication in 23andme:
European
African
Latino
0.50
0.71
1.0
1.41
2.0
Odds Ratio

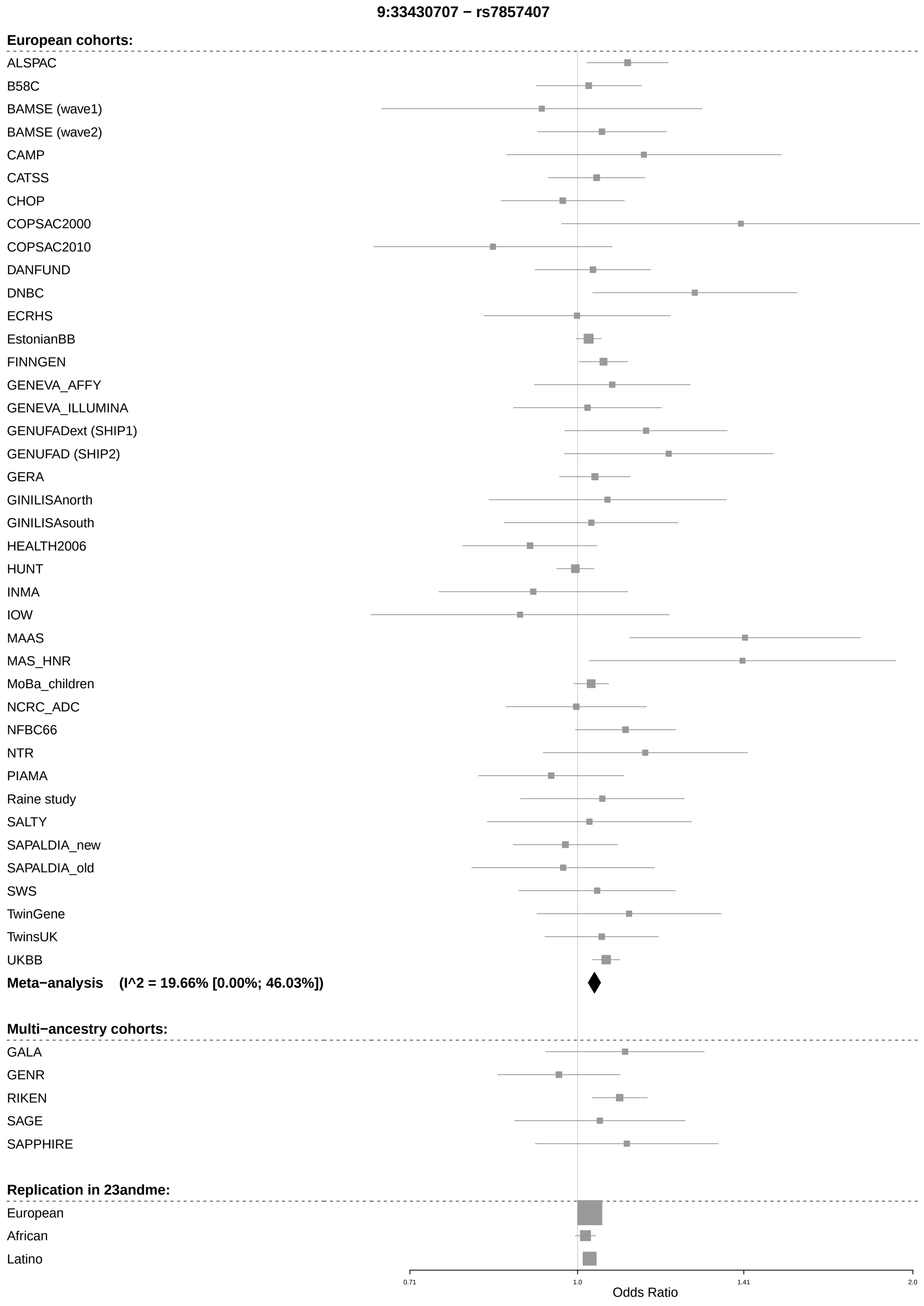

9:33430707 − rs7857407
European cohorts:
ALSPAC
B58C
BAMSE (wave1)
BAMSE (wave2)
CAMP
CATSS
CHOP
COPSAC2000
COPSAC2010
DANFUND
DNBC
ECRHS
EstonianBB
FINNGEN
GENEVA_AFFY
GENEVA_ILLUMINA
GENUFADext (SHIP1)
GENUFAD (SHIP2)
GERA
GINILISAnorth
GINILISAsouth
HEALTH2006
HUNT
INMA
IOW
MAAS
MAS_HNR
MoBa_children
NCRC_ADC
NFBC66
NTR
PIAMA
Raine study
SALTY
SAPALDIA_new
SAPALDIA_old
SWS
TwinGene
TwinsUK
UKBB
Meta−analysis (I^2 = 19.66% [0.00%; 46.03%])
Multi−ancestry cohorts:
GALA
GENR
RIKEN
SAGE
SAPPHIRE
Replication in 23andme:
European
African
Latino
0.71
1.0
1.41
2.0
Odds Ratio

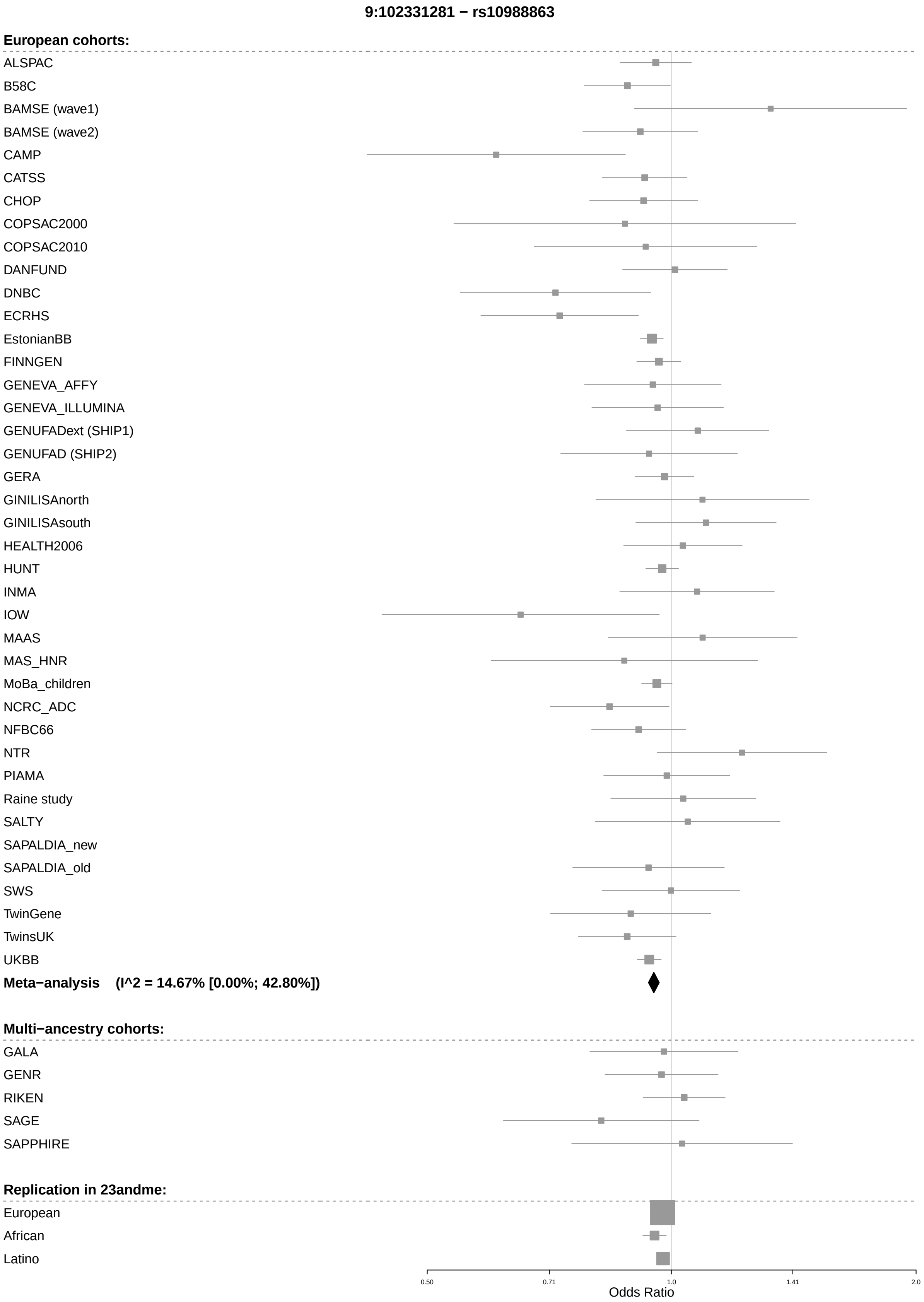
9:102331281 − rs10988863
European cohorts:
ALSPAC
B58C
BAMSE (wave1)
BAMSE (wave2)
CAMP
CATSS
CHOP
COPSAC2000
COPSAC2010
DANFUND
DNBC
ECRHS
EstonianBB
FINNGEN
GENEVA_AFFY
GENEVA_ILLUMINA
GENUFADext (SHIP1)
GENUFAD (SHIP2)
GERA
GINILISAnorth
GINILISAsouth
HEALTH2006
HUNT
INMA
IOW
MAAS
MAS_HNR
MoBa_children
NCRC_ADC
NFBC66
NTR
PIAMA
Raine study
SALTY
SAPALDIA_new
SAPALDIA_old
SWS
TwinGene
TwinsUK
UKBB
Meta−analysis (I^2 = 14.67% [0.00%; 42.80%])
Multi−ancestry cohorts:
GALA
GENR
RIKEN
SAGE
SAPPHIRE
Replication in 23andme:
European
African
Latino
0.50
0.71
1.0
1.41
2.0
Odds Ratio

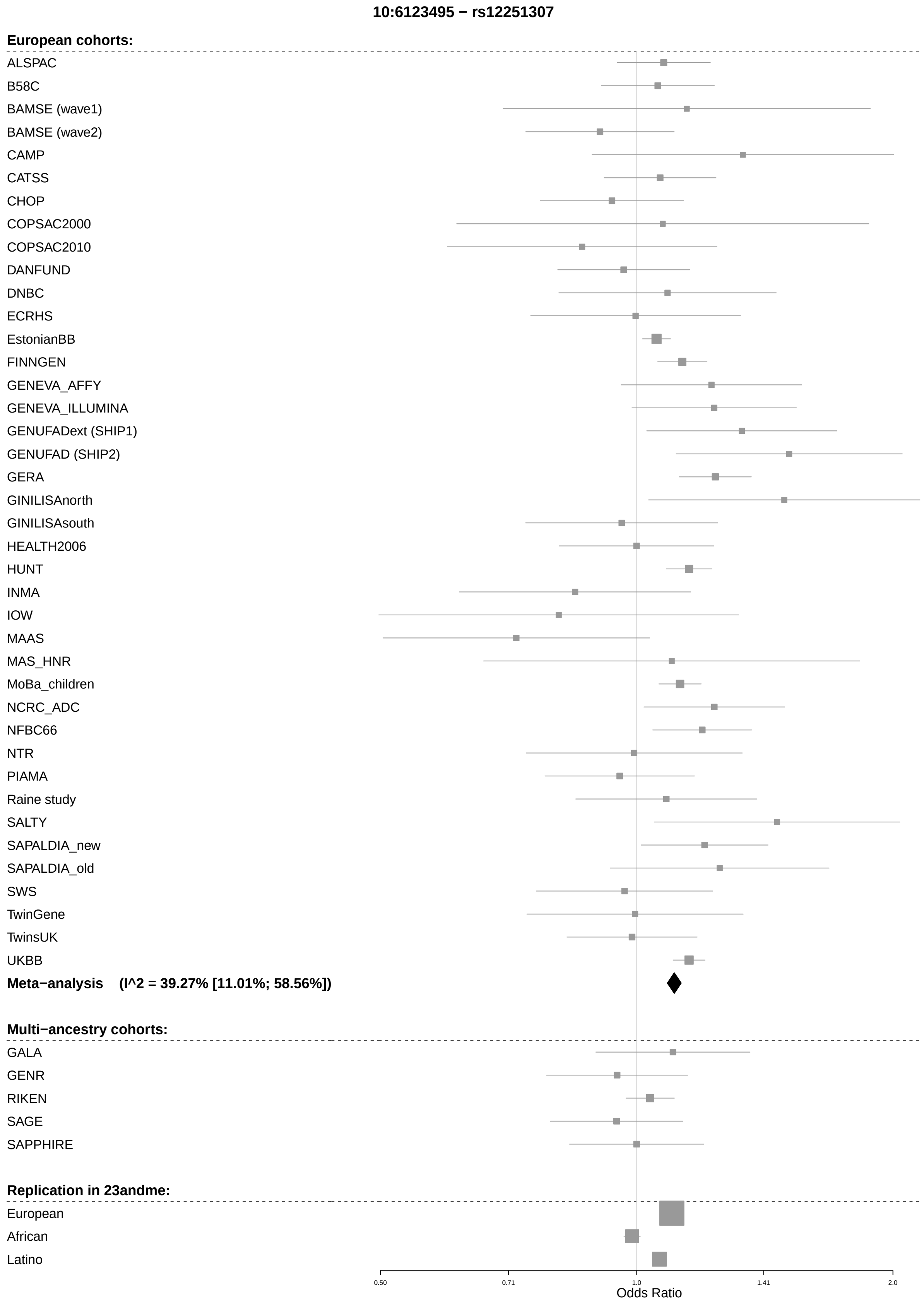
10:6123495 − rs12251307
European cohorts:
ALSPAC
B58C
BAMSE (wave1)
BAMSE (wave2)
CAMP
CATSS
CHOP
COPSAC2000
COPSAC2010
DANFUND
DNBC
ECRHS
EstonianBB
FINNGEN
GENEVA_AFFY
GENEVA_ILLUMINA
GENUFADext (SHIP1)
GENUFAD (SHIP2)
GERA
GINILISAnorth
GINILISAsouth
HEALTH2006
HUNT
INMA
IOW
MAAS
MAS_HNR
MoBa_children
NCRC_ADC
NFBC66
NTR
PIAMA
Raine study
SALTY
SAPALDIA_new
SAPALDIA_old
SWS
TwinGene
TwinsUK
UKBB
Meta−analysis (I^2 = 39.27% [11.01%; 58.56%])
Multi−ancestry cohorts:
GALA
GENR
RIKEN
SAGE
SAPPHIRE
Replication in 23andme:
European
African
Latino
0.50
0.71
1.0
1.41
2.0
Odds Ratio

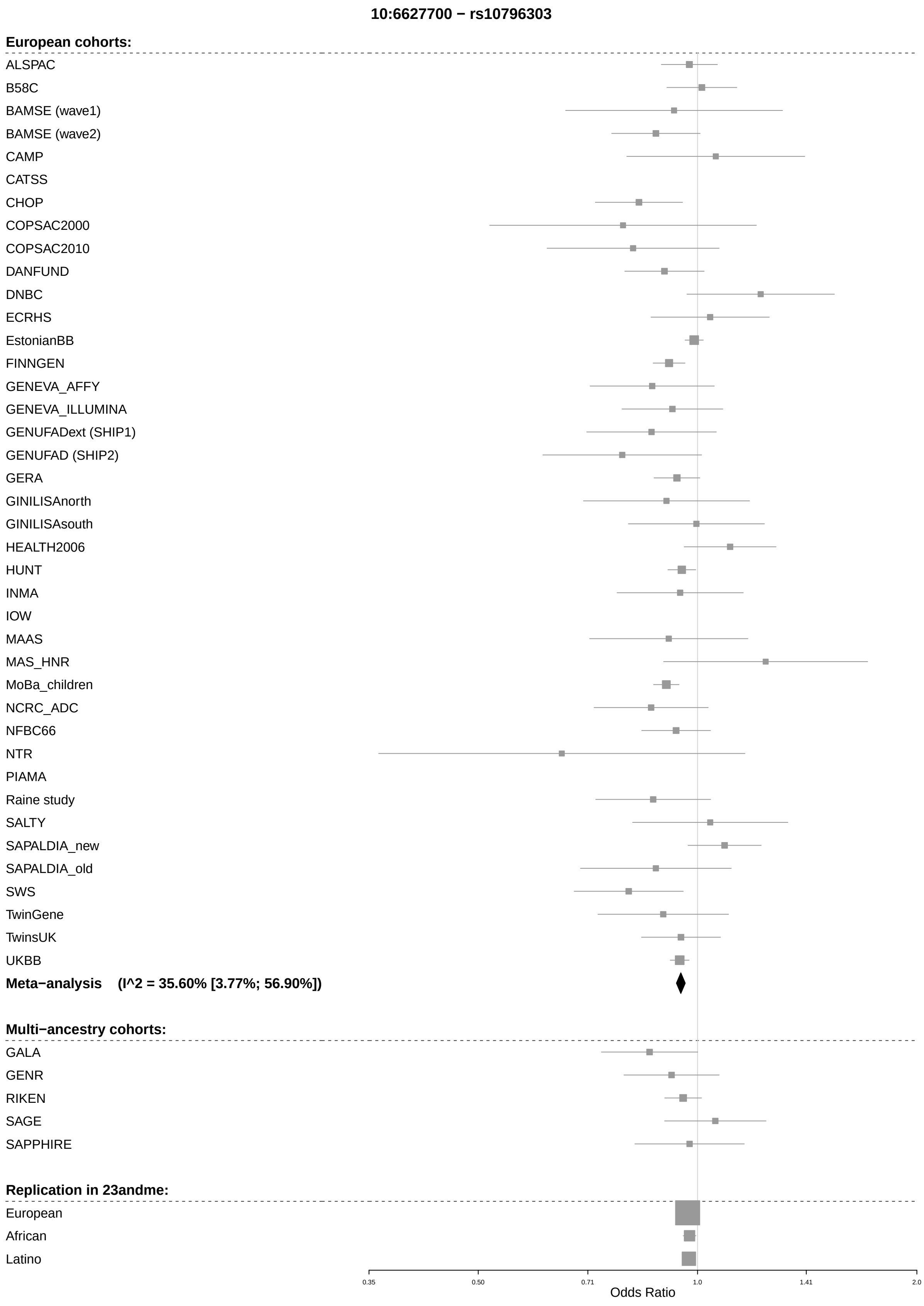
10:6627700 − rs10796303
European cohorts:
ALSPAC
B58C
BAMSE (wave1)
BAMSE (wave2)
CAMP
CATSS
CHOP
COPSAC2000
COPSAC2010
DANFUND
DNBC
ECRHS
EstonianBB
FINNGEN
GENEVA_AFFY
GENEVA_ILLUMINA
GENUFADext (SHIP1)
GENUFAD (SHIP2)
GERA
GINILISAnorth
GINILISAsouth
HEALTH2006
HUNT
INMA
IOW
MAAS
MAS_HNR
MoBa_children
NCRC_ADC
NFBC66
NTR
PIAMA
Raine study
SALTY
SAPALDIA_new
SAPALDIA_old
SWS
TwinGene
TwinsUK
UKBB
Meta−analysis (I^2 = 35.60% [3.77%; 56.90%])
Multi−ancestry cohorts:
GALA
GENR
RIKEN
SAGE
SAPPHIRE
Replication in 23andme:
European
African
Latino
0.35
0.50
0.71
1.0
1.41
2.0
Odds Ratio

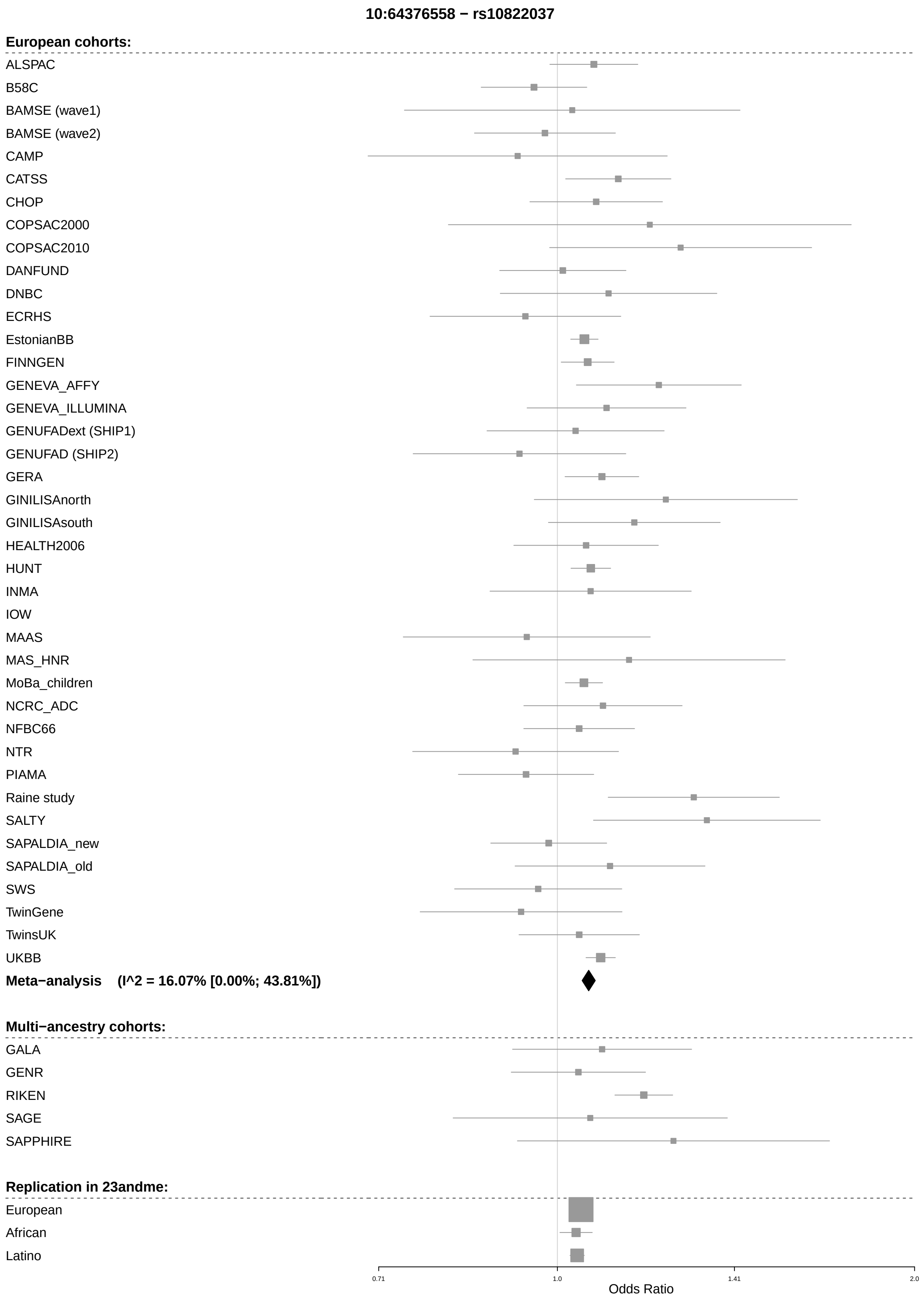
10:64376558 − rs10822037
European cohorts:
ALSPAC
B58C
BAMSE (wave1)
BAMSE (wave2)
CAMP
CATSS
CHOP
COPSAC2000
COPSAC2010
DANFUND
DNBC
ECRHS
EstonianBB
FINNGEN
GENEVA_AFFY
GENEVA_ILLUMINA
GENUFADext (SHIP1)
GENUFAD (SHIP2)
GERA
GINILISAnorth
GINILISAsouth
HEALTH2006
HUNT
INMA
IOW
MAAS
MAS_HNR
MoBa_children
NCRC_ADC
NFBC66
NTR
PIAMA
Raine study
SALTY
SAPALDIA_new
SAPALDIA_old
SWS
TwinGene
TwinsUK
UKBB
Meta−analysis (I^2 = 16.07% [0.00%; 43.81%])
Multi−ancestry cohorts:
GALA
GENR
RIKEN
SAGE
SAPPHIRE
Replication in 23andme:
European
African
Latino
0.71
1.0
1.41
2.0
Odds Ratio

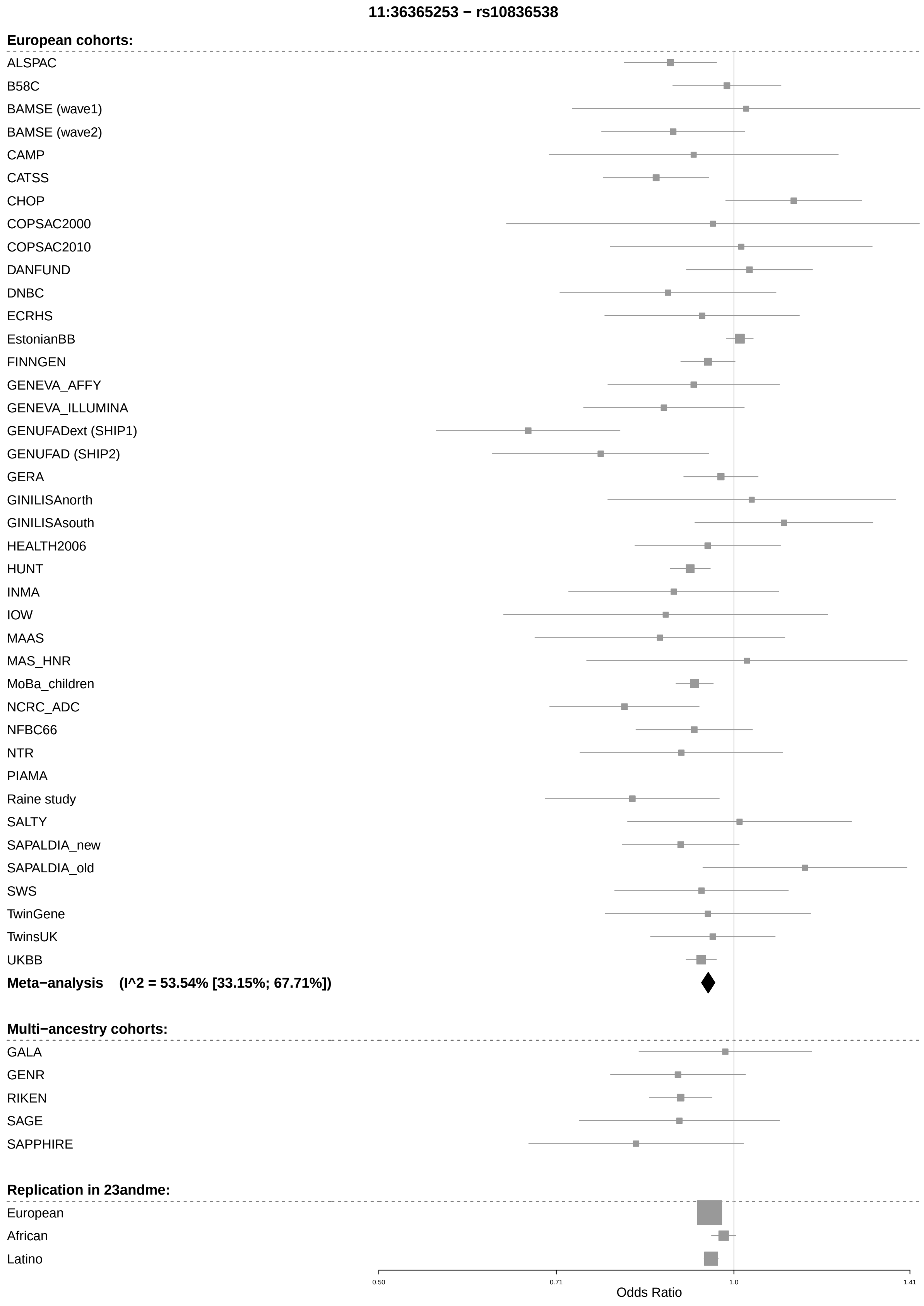
11:36365253 − rs10836538
European cohorts:
ALSPAC
B58C
BAMSE (wave1)
BAMSE (wave2)
CAMP
CATSS
CHOP
COPSAC2000
COPSAC2010
DANFUND
DNBC
ECRHS
EstonianBB
FINNGEN
GENEVA_AFFY
GENEVA_ILLUMINA
GENUFADext (SHIP1)
GENUFAD (SHIP2)
GERA
GINILISAnorth
GINILISAsouth
HEALTH2006
HUNT
INMA
IOW
MAAS
MAS_HNR
MoBa_children
NCRC_ADC
NFBC66
NTR
PIAMA
Raine study
SALTY
SAPALDIA_new
SAPALDIA_old
SWS
TwinGene
TwinsUK
UKBB
Meta−analysis (I^2 = 53.54% [33.15%; 67.71%])
Multi−ancestry cohorts:
GALA
GENR
RIKEN
SAGE
SAPPHIRE
Replication in 23andme:
European
African
Latino
0.50
0.71
1.0
1.41
Odds Ratio

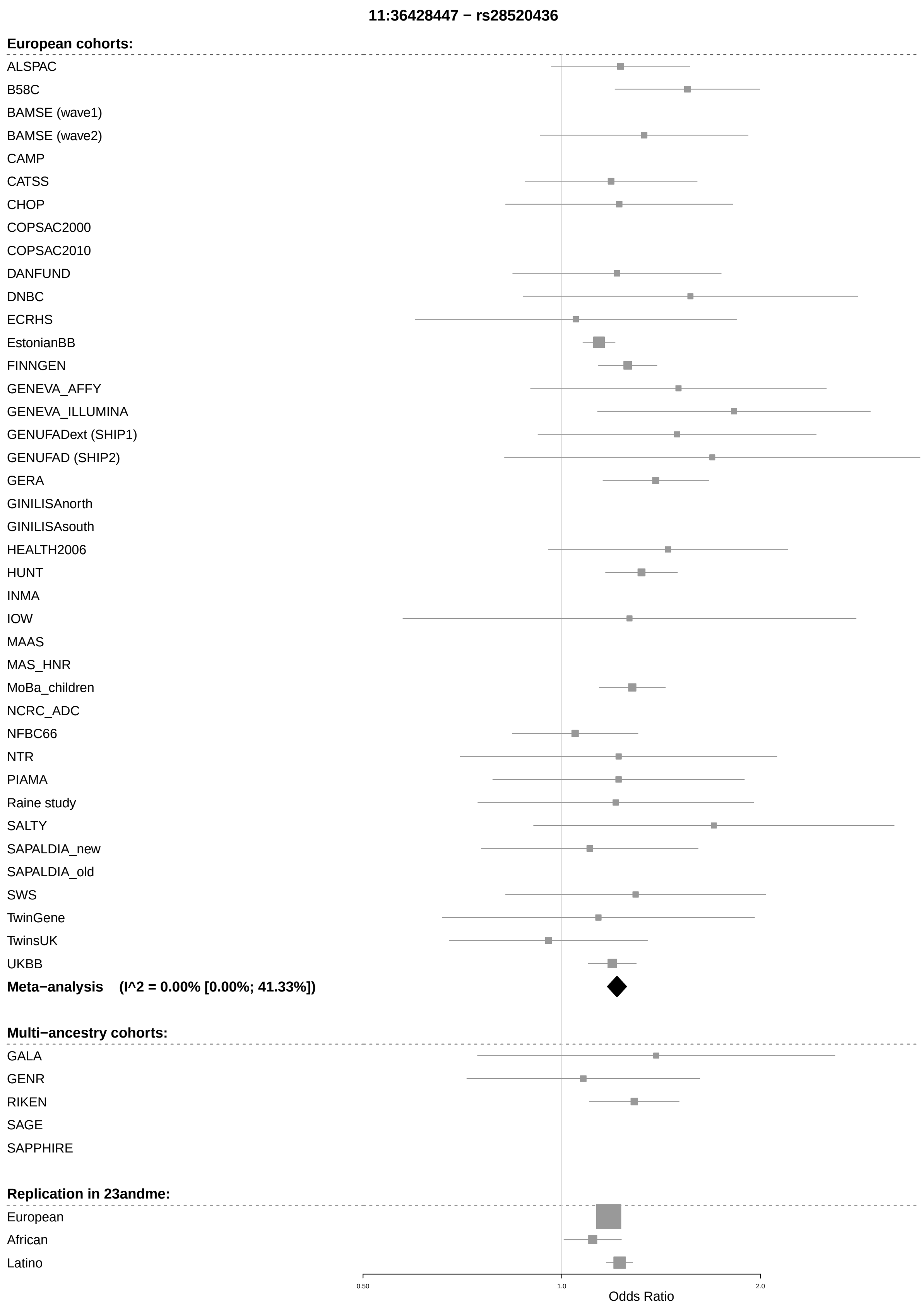
11:36428447 − rs28520436
European cohorts:
ALSPAC
B58C
BAMSE (wave1)
BAMSE (wave2)
CAMP
CATSS
CHOP
COPSAC2000
COPSAC2010
DANFUND
DNBC
ECRHS
EstonianBB
FINNGEN
GENEVA_AFFY
GENEVA_ILLUMINA
GENUFADext (SHIP1)
GENUFAD (SHIP2)
GERA
GINILISAnorth
GINILISAsouth
HEALTH2006
HUNT
INMA
IOW
MAAS
MAS_HNR
MoBa_children
NCRC_ADC
NFBC66
NTR
PIAMA
Raine study
SALTY
SAPALDIA_new
SAPALDIA_old
SWS
TwinGene
TwinsUK
UKBB
Meta−analysis (I^2 = 0.00% [0.00%; 41.33%])
Multi−ancestry cohorts:
GALA
GENR
RIKEN
SAGE
SAPPHIRE
Replication in 23andme:
European
African
Latino
0.50
1.0
2.0
Odds Ratio

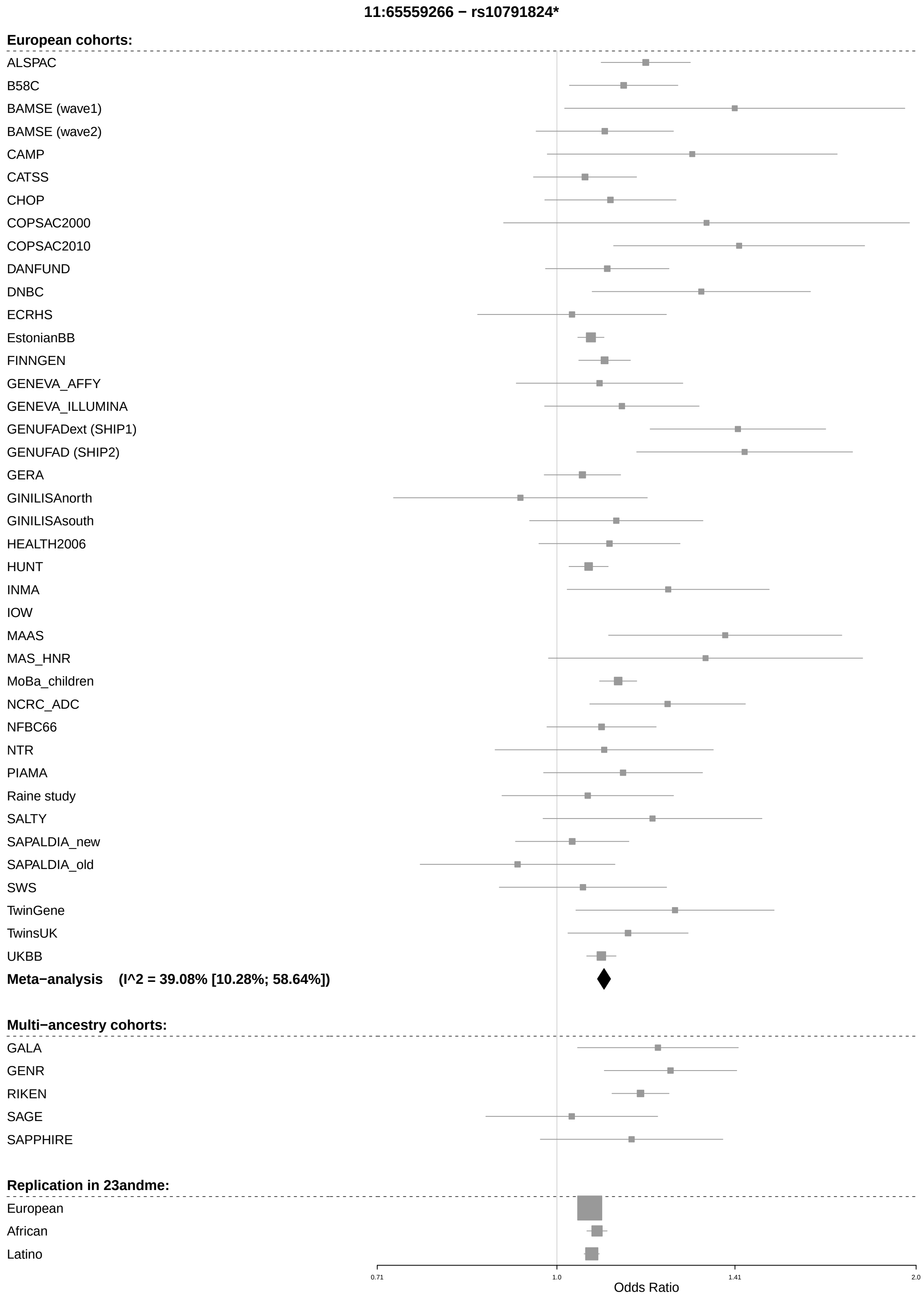

11:65559266 − rs10791824*
European cohorts:
ALSPAC
B58C
BAMSE (wave1)
BAMSE (wave2)
CAMP
CATSS
CHOP
COPSAC2000
COPSAC2010
DANFUND
DNBC
ECRHS
EstonianBB
FINNGEN
GENEVA_AFFY
GENEVA_ILLUMINA
GENUFADext (SHIP1)
GENUFAD (SHIP2)
GERA
GINILISAnorth
GINILISAsouth
HEALTH2006
HUNT
INMA
IOW
MAAS
MAS_HNR
MoBa_children
NCRC_ADC
NFBC66
NTR
PIAMA
Raine study
SALTY
SAPALDIA_new
SAPALDIA_old
SWS
TwinGene
TwinsUK
UKBB
Meta−analysis (I^2 = 39.08% [10.28%; 58.64%])
Multi−ancestry cohorts:
GALA
GENR
RIKEN
SAGE
SAPPHIRE
Replication in 23andme:
European
African
Latino
0.71
1.0
1.41
2.0
Odds Ratio

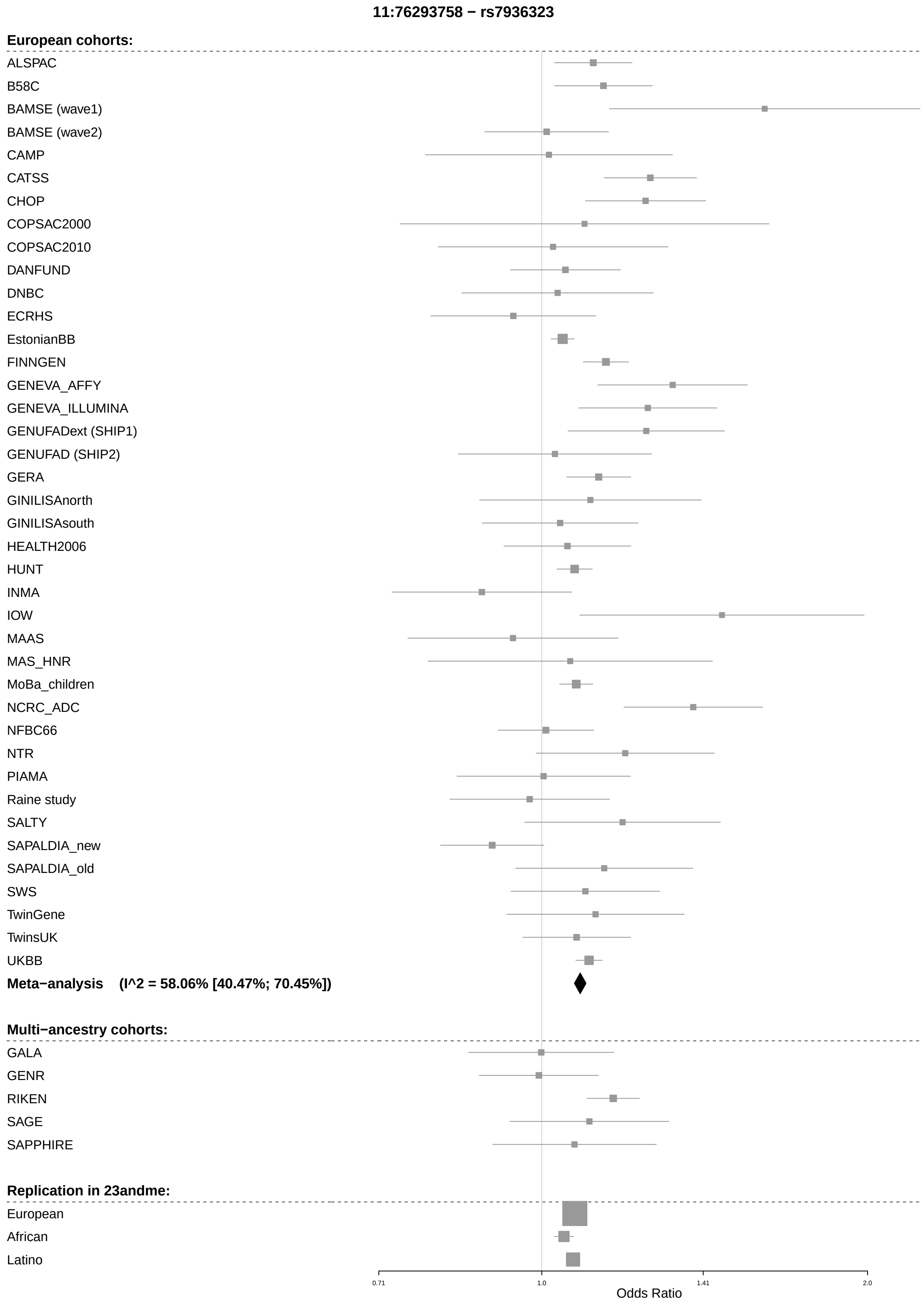

11:76293758 − rs7936323
European cohorts:
ALSPAC
B58C
BAMSE (wave1)
BAMSE (wave2)
CAMP
CATSS
CHOP
COPSAC2000
COPSAC2010
DANFUND
DNBC
ECRHS
EstonianBB
FINNGEN
GENEVA_AFFY
GENEVA_ILLUMINA
GENUFADext (SHIP1)
GENUFAD (SHIP2)
GERA
GINILISAnorth
GINILISAsouth
HEALTH2006
HUNT
INMA
IOW
MAAS
MAS_HNR
MoBa_children
NCRC_ADC
NFBC66
NTR
PIAMA
Raine study
SALTY
SAPALDIA_new
SAPALDIA_old
SWS
TwinGene
TwinsUK
UKBB
Meta−analysis (I^2 = 58.06% [40.47%; 70.45%])
Multi−ancestry cohorts:
GALA
GENR
RIKEN
SAGE
SAPPHIRE
Replication in 23andme:
European
African
Latino
0.71
1.0
1.41
2.0
Odds Ratio

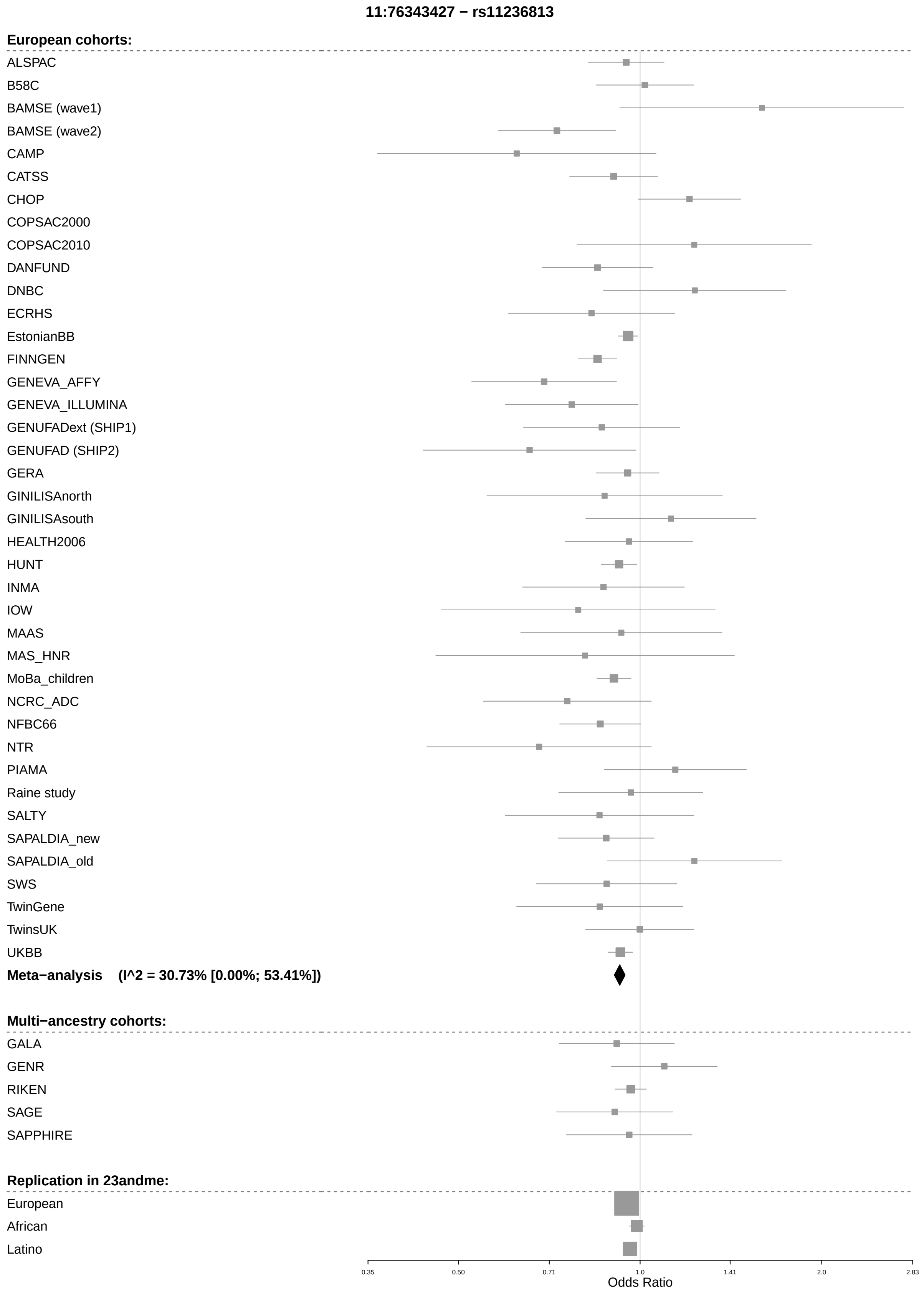
11:76343427 − rs11236813
European cohorts:
ALSPAC
B58C
BAMSE (wave1)
BAMSE (wave2)
CAMP
CATSS
CHOP
COPSAC2000
COPSAC2010
DANFUND
DNBC
ECRHS
EstonianBB
FINNGEN
GENEVA_AFFY
GENEVA_ILLUMINA
GENUFADext (SHIP1)
GENUFAD (SHIP2)
GERA
GINILISAnorth
GINILISAsouth
HEALTH2006
HUNT
INMA
IOW
MAAS
MAS_HNR
MoBa_children
NCRC_ADC
NFBC66
NTR
PIAMA
Raine study
SALTY
SAPALDIA_new
SAPALDIA_old
SWS
TwinGene
TwinsUK
UKBB
Meta−analysis (I^2 = 30.73% [0.00%; 53.41%])
Multi−ancestry cohorts:
GALA
GENR
RIKEN
SAGE
SAPPHIRE
Replication in 23andme:
European
African
Latino
0.35
0.50
0.71
1.0
1.41
2.0
2.83
Odds Ratio

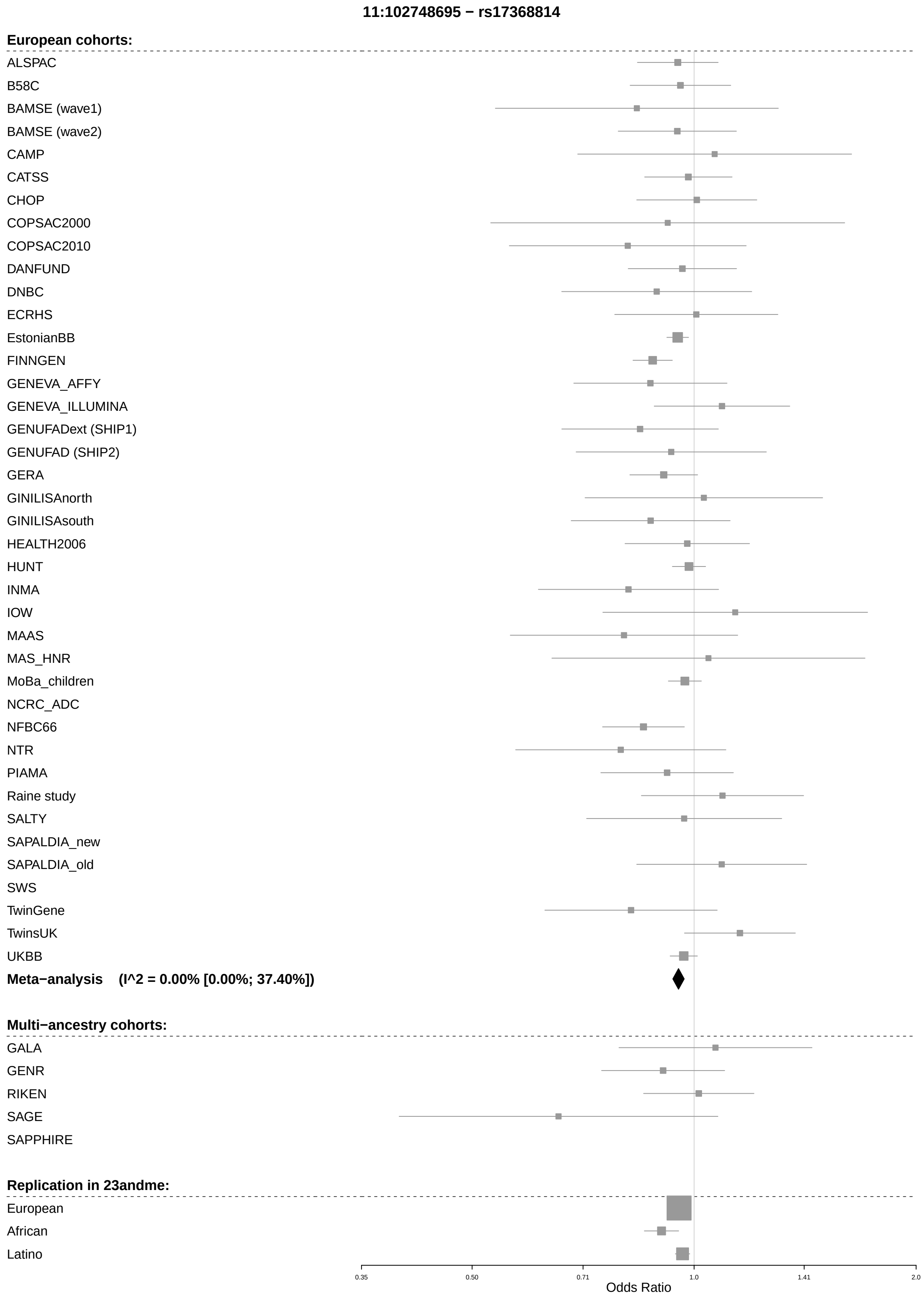
11:102748695 − rs17368814
European cohorts:
ALSPAC
B58C
BAMSE (wave1)
BAMSE (wave2)
CAMP
CATSS
CHOP
COPSAC2000
COPSAC2010
DANFUND
DNBC
ECRHS
EstonianBB
FINNGEN
GENEVA_AFFY
GENEVA_ILLUMINA
GENUFADext (SHIP1)
GENUFAD (SHIP2)
GERA
GINILISAnorth
GINILISAsouth
HEALTH2006
HUNT
INMA
IOW
MAAS
MAS_HNR
MoBa_children
NCRC_ADC
NFBC66
NTR
PIAMA
Raine study
SALTY
SAPALDIA_new
SAPALDIA_old
SWS
TwinGene
TwinsUK
UKBB
Meta−analysis (I^2 = 0.00% [0.00%; 37.40%])
Multi−ancestry cohorts:
GALA
GENR
RIKEN
SAGE
SAPPHIRE
Replication in 23andme:
European
African
Latino
0.35
0.50
0.71
1.0
1.41
2.0
Odds Ratio

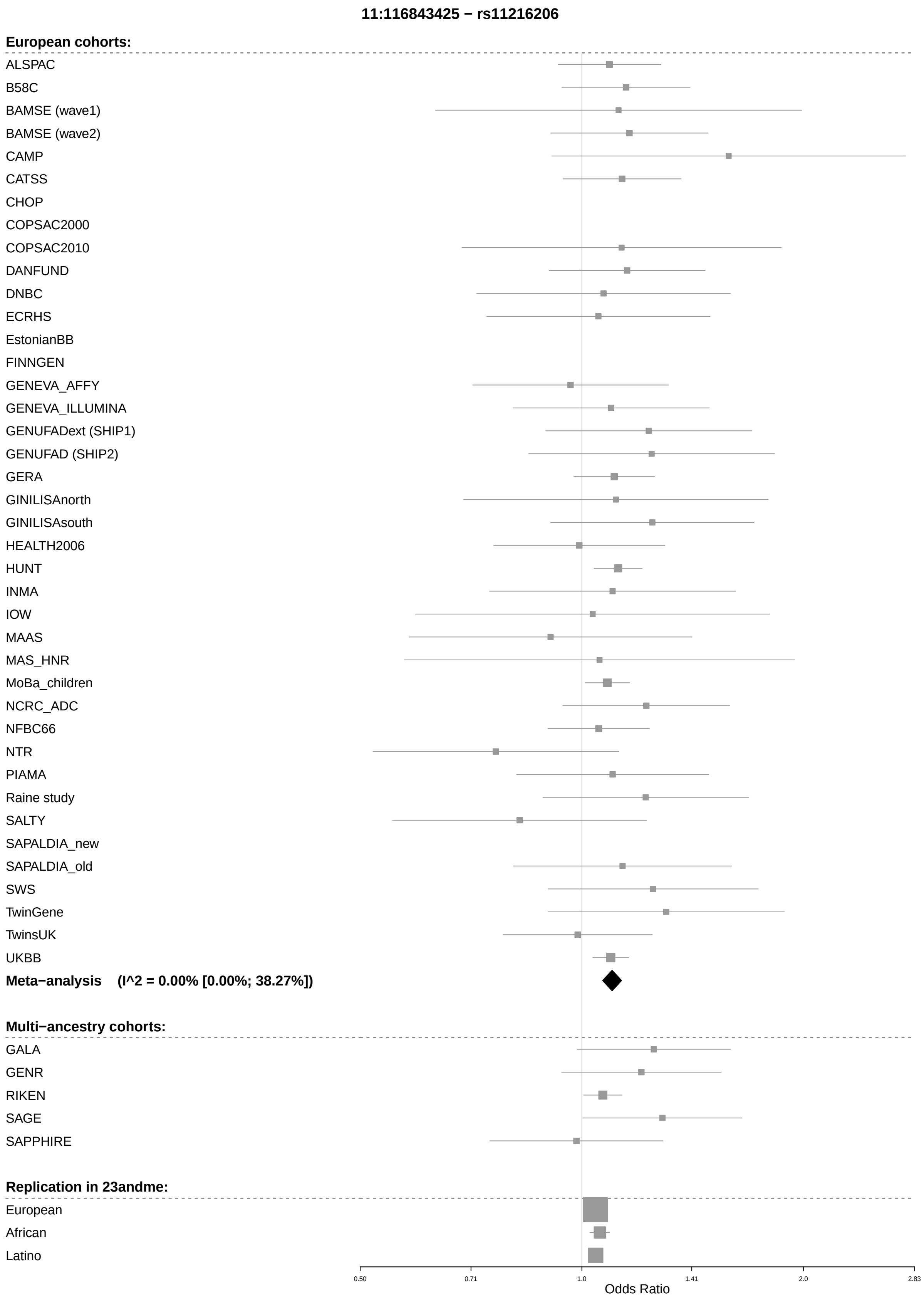
11:116843425 − rs11216206
European cohorts:
ALSPAC
B58C
BAMSE (wave1)
BAMSE (wave2)
CAMP
CATSS
CHOP
COPSAC2000
COPSAC2010
DANFUND
DNBC
ECRHS
EstonianBB
FINNGEN
GENEVA_AFFY
GENEVA_ILLUMINA
GENUFADext (SHIP1)
GENUFAD (SHIP2)
GERA
GINILISAnorth
GINILISAsouth
HEALTH2006
HUNT
INMA
IOW
MAAS
MAS_HNR
MoBa_children
NCRC_ADC
NFBC66
NTR
PIAMA
Raine study
SALTY
SAPALDIA_new
SAPALDIA_old
SWS
TwinGene
TwinsUK
UKBB
Meta−analysis (I^2 = 0.00% [0.00%; 38.27%])
Multi−ancestry cohorts:
GALA
GENR
RIKEN
SAGE
SAPPHIRE
Replication in 23andme:
European
African
Latino
0.50
0.71
1.0
1.41
2.0
2.83
Odds Ratio

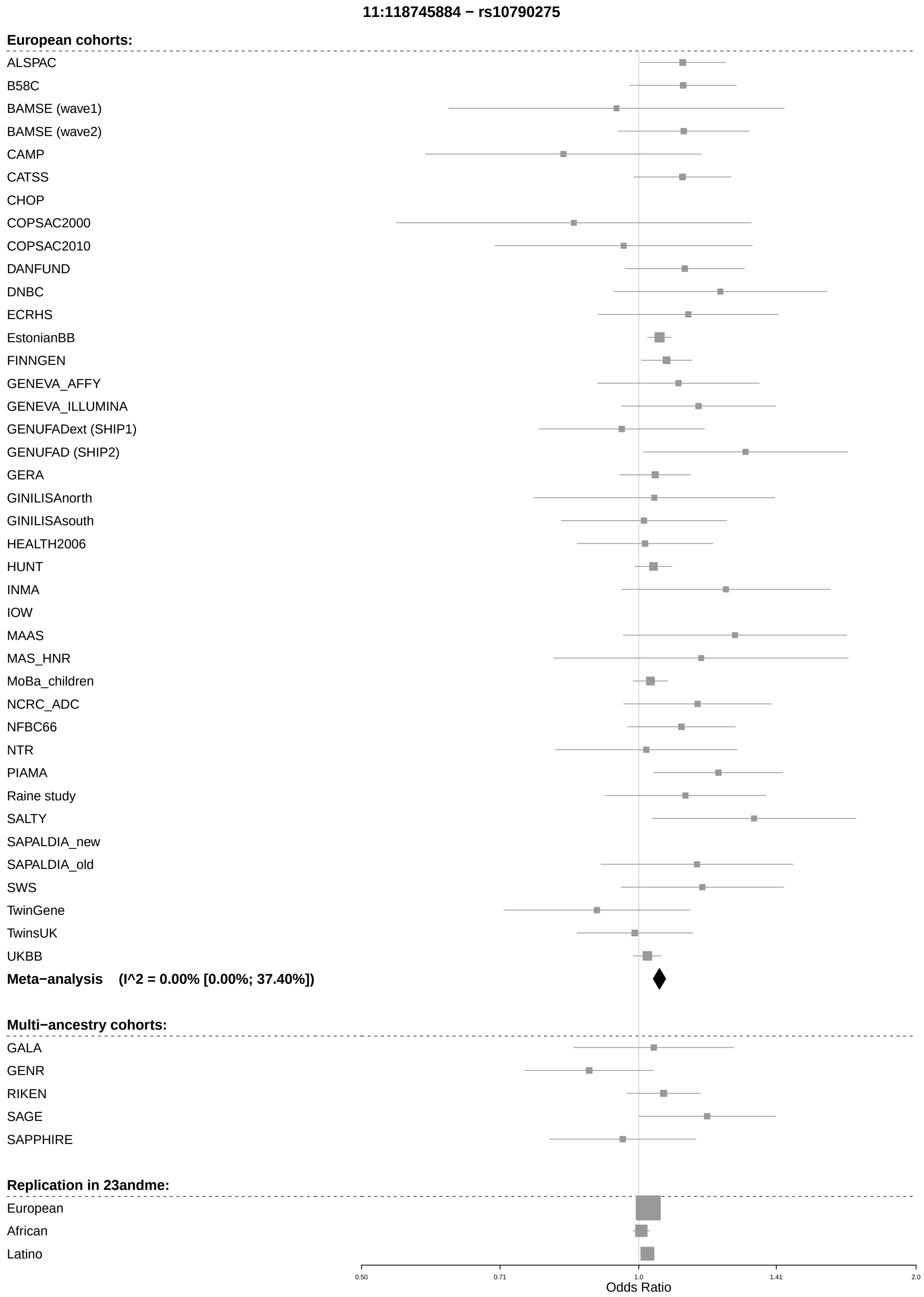
11:118745884 − rs10790275
European cohorts:
ALSPAC
B58C
BAMSE (wave1)
BAMSE (wave2)
CAMP
CATSS
CHOP
COPSAC2000
COPSAC2010
DANFUND
DNBC
ECRHS
EstonianBB
FINNGEN
GENEVA_AFFY
GENEVA_ILLUMINA
GENUFADext (SHIP1)
GENUFAD (SHIP2)
GERA
GINILISAnorth
GINILISAsouth
HEALTH2006
HUNT
INMA
IOW
MAAS
MAS_HNR
MoBa_children
NCRC_ADC
NFBC66
NTR
PIAMA
Raine study
SALTY
SAPALDIA_new
SAPALDIA_old
SWS
TwinGene
TwinsUK
UKBB
Meta−analysis (I^2 = 0.00% [0.00%; 37.40%])
Multi−ancestry cohorts:
GALA
GENR
RIKEN
SAGE
SAPPHIRE
Replication in 23andme:
European
African
Latino
0.50
0.71
1.0
1.41
2.0
Odds Ratio

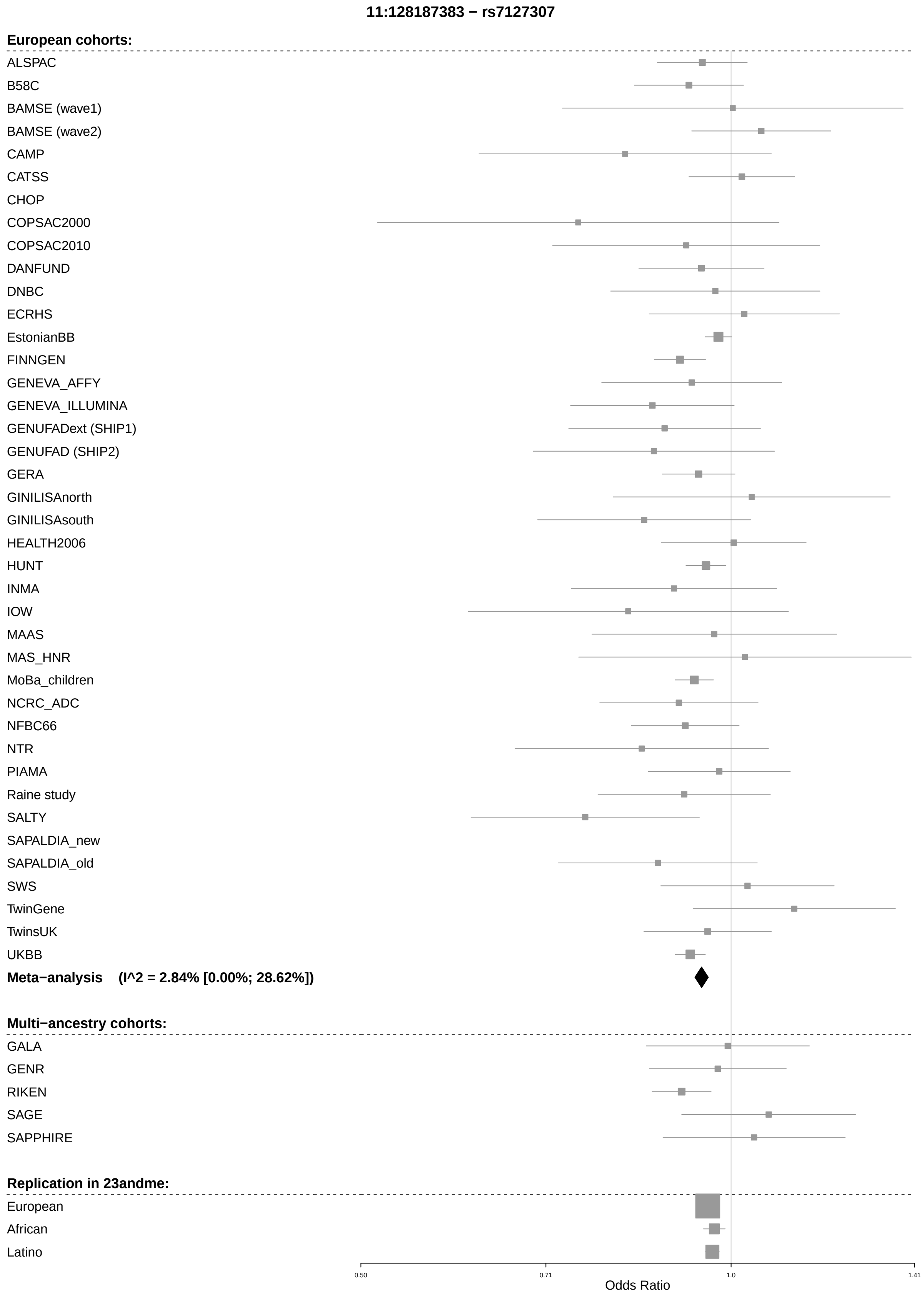
11:128187383 − rs7127307
European cohorts:
ALSPAC
B58C
BAMSE (wave1)
BAMSE (wave2)
CAMP
CATSS
CHOP
COPSAC2000
COPSAC2010
DANFUND
DNBC
ECRHS
EstonianBB
FINNGEN
GENEVA_AFFY
GENEVA_ILLUMINA
GENUFADext (SHIP1)
GENUFAD (SHIP2)
GERA
GINILISAnorth
GINILISAsouth
HEALTH2006
HUNT
INMA
IOW
MAAS
MAS_HNR
MoBa_children
NCRC_ADC
NFBC66
NTR
PIAMA
Raine study
SALTY
SAPALDIA_new
SAPALDIA_old
SWS
TwinGene
TwinsUK
UKBB
Meta−analysis (I^2 = 2.84% [0.00%; 28.62%])
Multi−ancestry cohorts:
GALA
GENR
RIKEN
SAGE
SAPPHIRE
Replication in 23andme:
European
African
Latino
0.50
0.71
1.0
1.41
Odds Ratio

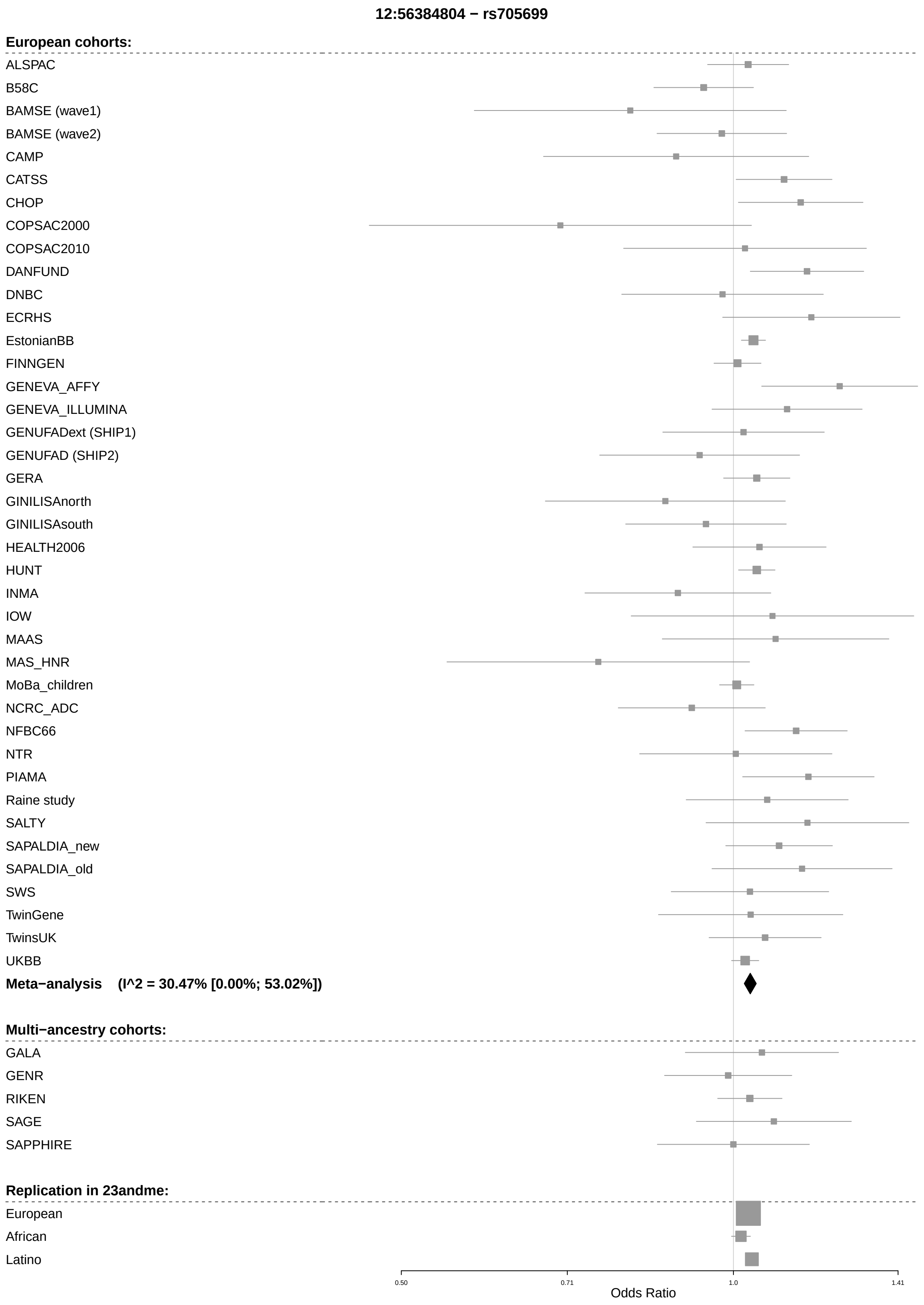
12:56384804 − rs705699
European cohorts:
ALSPAC
B58C
BAMSE (wave1)
BAMSE (wave2)
CAMP
CATSS
CHOP
COPSAC2000
COPSAC2010
DANFUND
DNBC
ECRHS
EstonianBB
FINNGEN
GENEVA_AFFY
GENEVA_ILLUMINA
GENUFADext (SHIP1)
GENUFAD (SHIP2)
GERA
GINILISAnorth
GINILISAsouth
HEALTH2006
HUNT
INMA
IOW
MAAS
MAS_HNR
MoBa_children
NCRC_ADC
NFBC66
NTR
PIAMA
Raine study
SALTY
SAPALDIA_new
SAPALDIA_old
SWS
TwinGene
TwinsUK
UKBB
Meta−analysis (I^2 = 30.47% [0.00%; 53.02%])
Multi−ancestry cohorts:
GALA
GENR
RIKEN
SAGE
SAPPHIRE
Replication in 23andme:
European
African
Latino
0.50
0.71
1.0
1.41
Odds Ratio

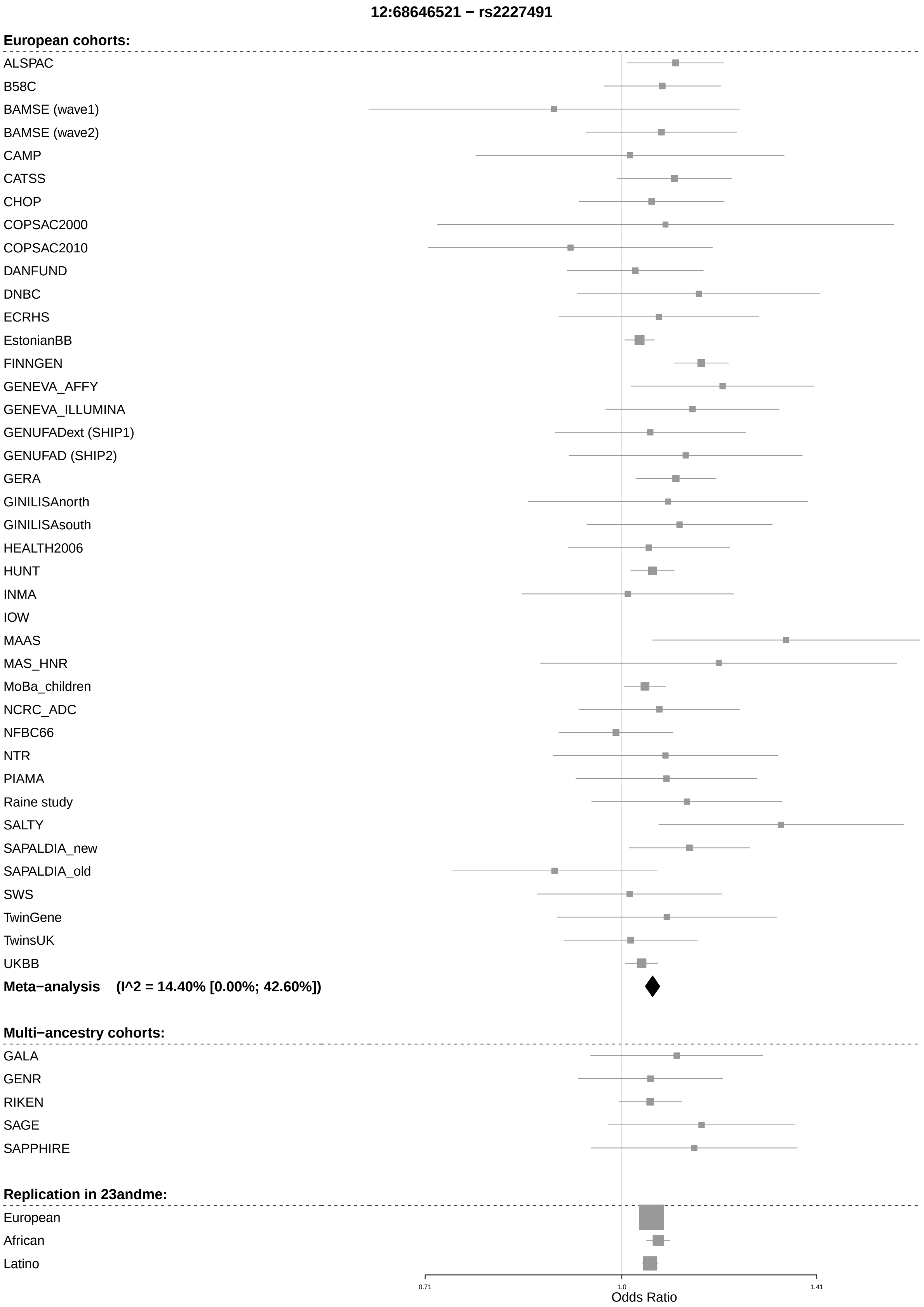
12:68646521 − rs2227491
European cohorts:
ALSPAC
B58C
BAMSE (wave1)
BAMSE (wave2)
CAMP
CATSS
CHOP
COPSAC2000
COPSAC2010
DANFUND
DNBC
ECRHS
EstonianBB
FINNGEN
GENEVA_AFFY
GENEVA_ILLUMINA
GENUFADext (SHIP1)
GENUFAD (SHIP2)
GERA
GINILISAnorth
GINILISAsouth
HEALTH2006
HUNT
INMA
IOW
MAAS
MAS_HNR
MoBa_children
NCRC_ADC
NFBC66
NTR
PIAMA
Raine study
SALTY
SAPALDIA_new
SAPALDIA_old
SWS
TwinGene
TwinsUK
UKBB
Meta−analysis (I^2 = 14.40% [0.00%; 42.60%])
Multi−ancestry cohorts:
GALA
GENR
RIKEN
SAGE
SAPPHIRE
Replication in 23andme:
European
African
Latino
0.71
1.0
1.41
Odds Ratio

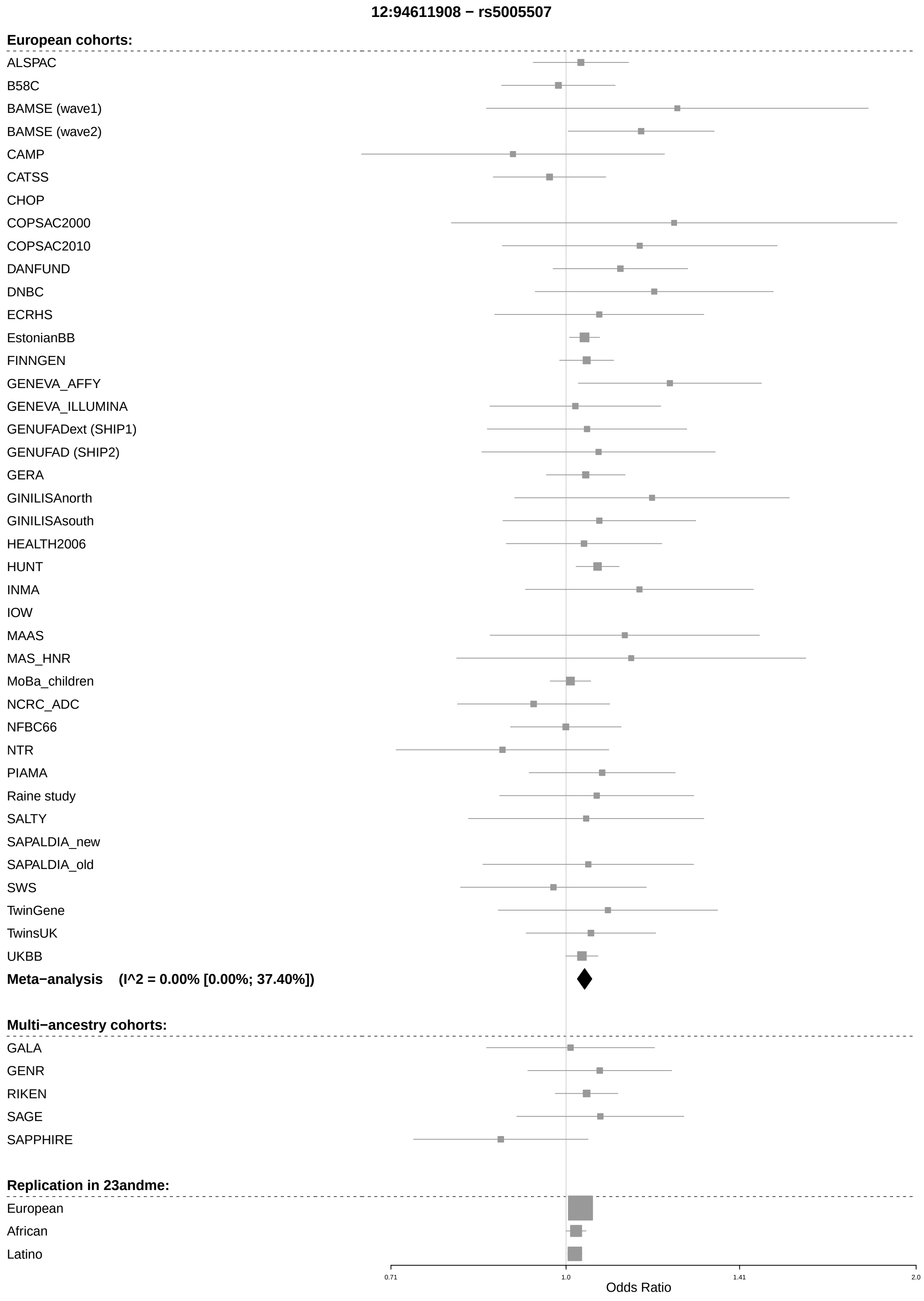

12:94611908 − rs5005507
European cohorts:
ALSPAC
B58C
BAMSE (wave1)
BAMSE (wave2)
CAMP
CATSS
CHOP
COPSAC2000
COPSAC2010
DANFUND
DNBC
ECRHS
EstonianBB
FINNGEN
GENEVA_AFFY
GENEVA_ILLUMINA
GENUFADext (SHIP1)
GENUFAD (SHIP2)
GERA
GINILISAnorth
GINILISAsouth
HEALTH2006
HUNT
INMA
IOW
MAAS
MAS_HNR
MoBa_children
NCRC_ADC
NFBC66
NTR
PIAMA
Raine study
SALTY
SAPALDIA_new
SAPALDIA_old
SWS
TwinGene
TwinsUK
UKBB
Meta−analysis (I^2 = 0.00% [0.00%; 37.40%])
Multi−ancestry cohorts:
GALA
GENR
RIKEN
SAGE
SAPPHIRE
Replication in 23andme:
European
African
Latino
0.71
1.0
1.41
2.0
Odds Ratio

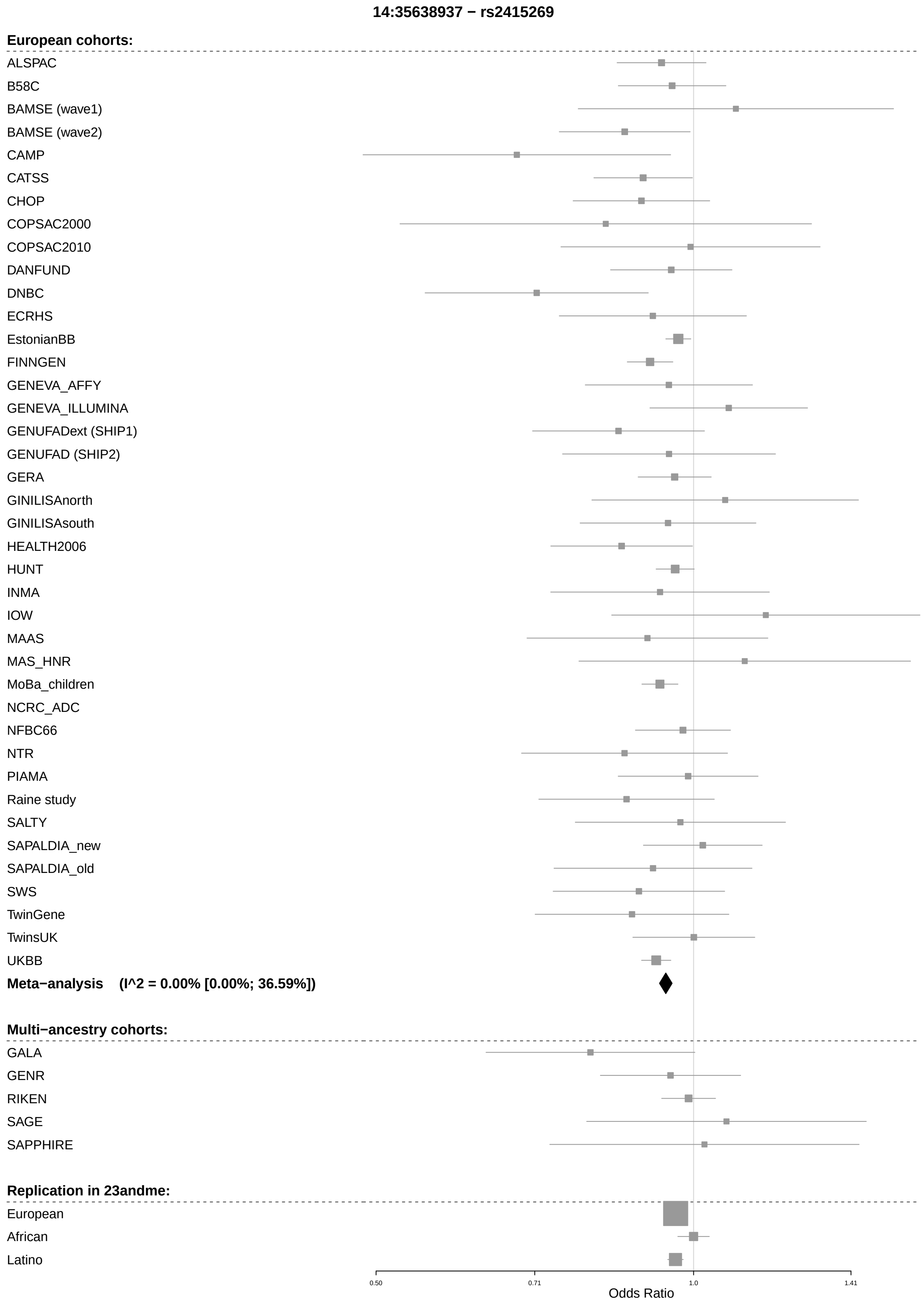
14:35638937 − rs2415269
European cohorts:
ALSPAC
B58C
BAMSE (wave1)
BAMSE (wave2)
CAMP
CATSS
CHOP
COPSAC2000
COPSAC2010
DANFUND
DNBC
ECRHS
EstonianBB
FINNGEN
GENEVA_AFFY
GENEVA_ILLUMINA
GENUFADext (SHIP1)
GENUFAD (SHIP2)
GERA
GINILISAnorth
GINILISAsouth
HEALTH2006
HUNT
INMA
IOW
MAAS
MAS_HNR
MoBa_children
NCRC_ADC
NFBC66
NTR
PIAMA
Raine study
SALTY
SAPALDIA_new
SAPALDIA_old
SWS
TwinGene
TwinsUK
UKBB
Meta−analysis (I^2 = 0.00% [0.00%; 36.59%])
Multi−ancestry cohorts:
GALA
GENR
RIKEN
SAGE
SAPPHIRE
Replication in 23andme:
European
African
Latino
0.50
0.71
1.0
1.41
Odds Ratio

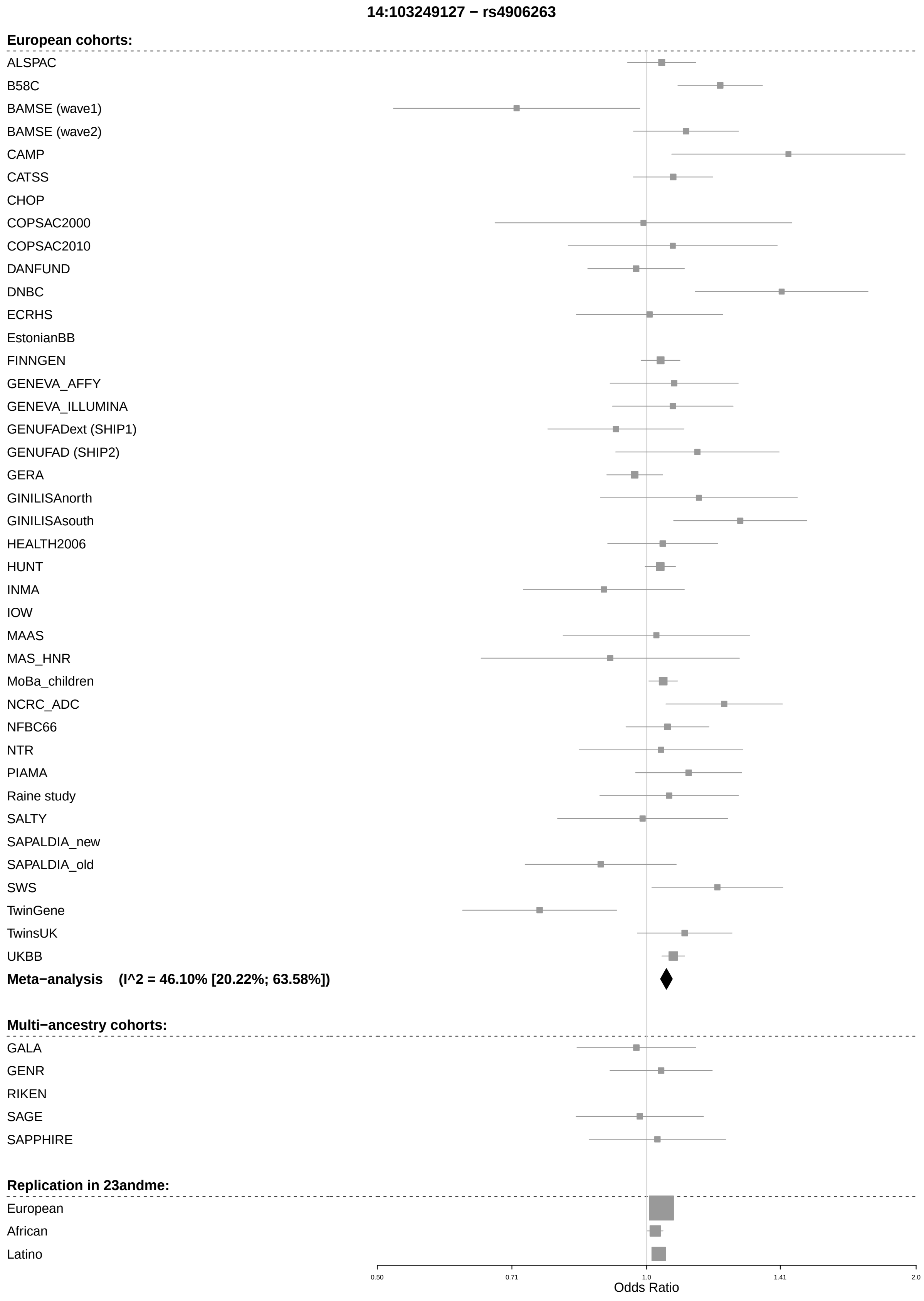
14:103249127 − rs4906263
European cohorts:
ALSPAC
B58C
BAMSE (wave1)
BAMSE (wave2)
CAMP
CATSS
CHOP
COPSAC2000
COPSAC2010
DANFUND
DNBC
ECRHS
EstonianBB
FINNGEN
GENEVA_AFFY
GENEVA_ILLUMINA
GENUFADext (SHIP1)
GENUFAD (SHIP2)
GERA
GINILISAnorth
GINILISAsouth
HEALTH2006
HUNT
INMA
IOW
MAAS
MAS_HNR
MoBa_children
NCRC_ADC
NFBC66
NTR
PIAMA
Raine study
SALTY
SAPALDIA_new
SAPALDIA_old
SWS
TwinGene
TwinsUK
UKBB
Meta−analysis (I^2 = 46.10% [20.22%; 63.58%])
Multi−ancestry cohorts:
GALA
GENR
RIKEN
SAGE
SAPPHIRE
Replication in 23andme:
European
African
Latino
0.50
0.71
1.0
1.41
2.0
Odds Ratio

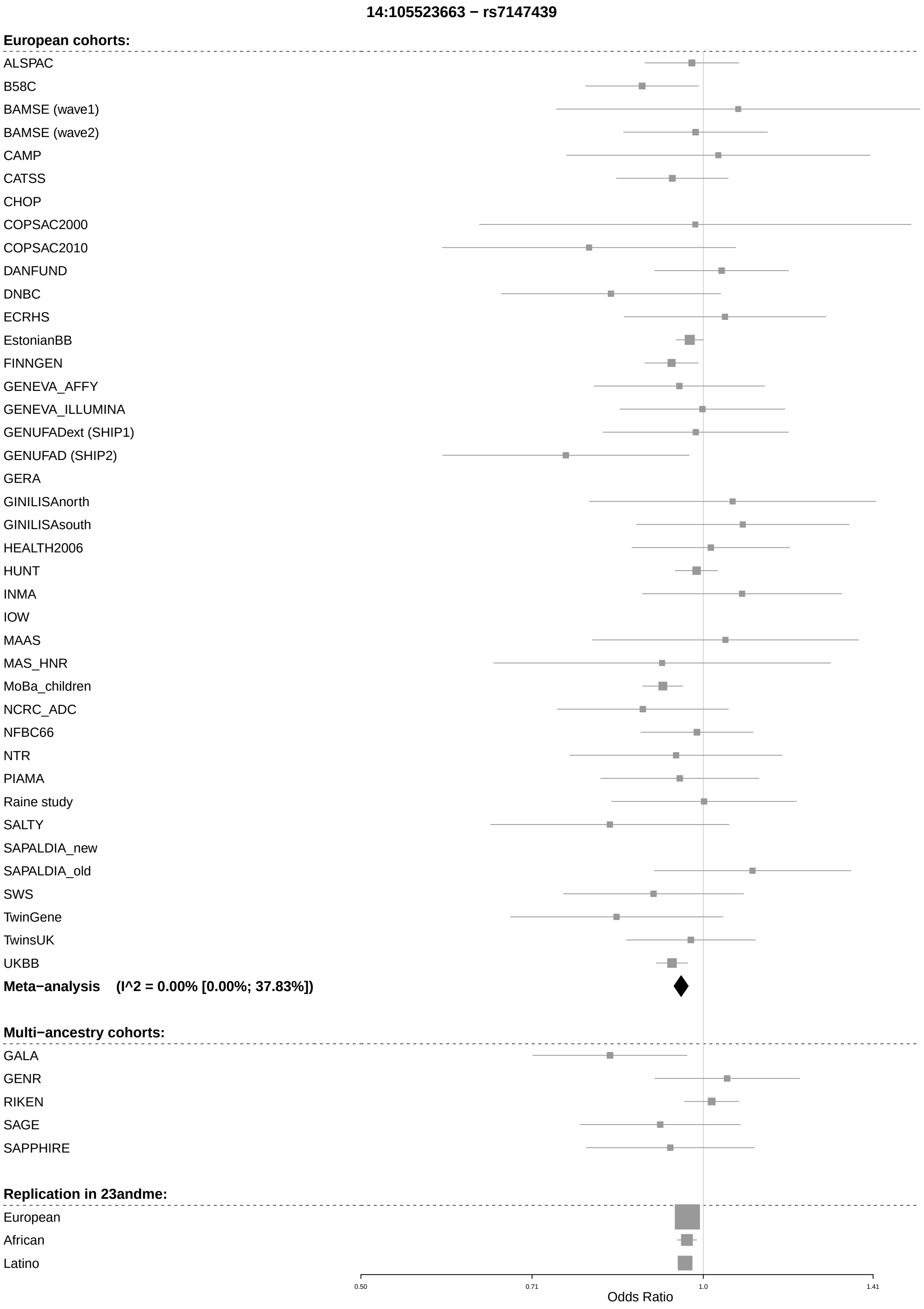
14:105523663 − rs7147439
European cohorts:
ALSPAC
B58C
BAMSE (wave1)
BAMSE (wave2)
CAMP
CATSS
CHOP
COPSAC2000
COPSAC2010
DANFUND
DNBC
ECRHS
EstonianBB
FINNGEN
GENEVA_AFFY
GENEVA_ILLUMINA
GENUFADext (SHIP1)
GENUFAD (SHIP2)
GERA
GINILISAnorth
GINILISAsouth
HEALTH2006
HUNT
INMA
IOW
MAAS
MAS_HNR
MoBa_children
NCRC_ADC
NFBC66
NTR
PIAMA
Raine study
SALTY
SAPALDIA_new
SAPALDIA_old
SWS
TwinGene
TwinsUK
UKBB
Meta−analysis (I^2 = 0.00% [0.00%; 37.83%])
Multi−ancestry cohorts:
GALA
GENR
RIKEN
SAGE
SAPPHIRE
Replication in 23andme:
European
African
Latino
0.50
0.71
1.0
1.41
Odds Ratio

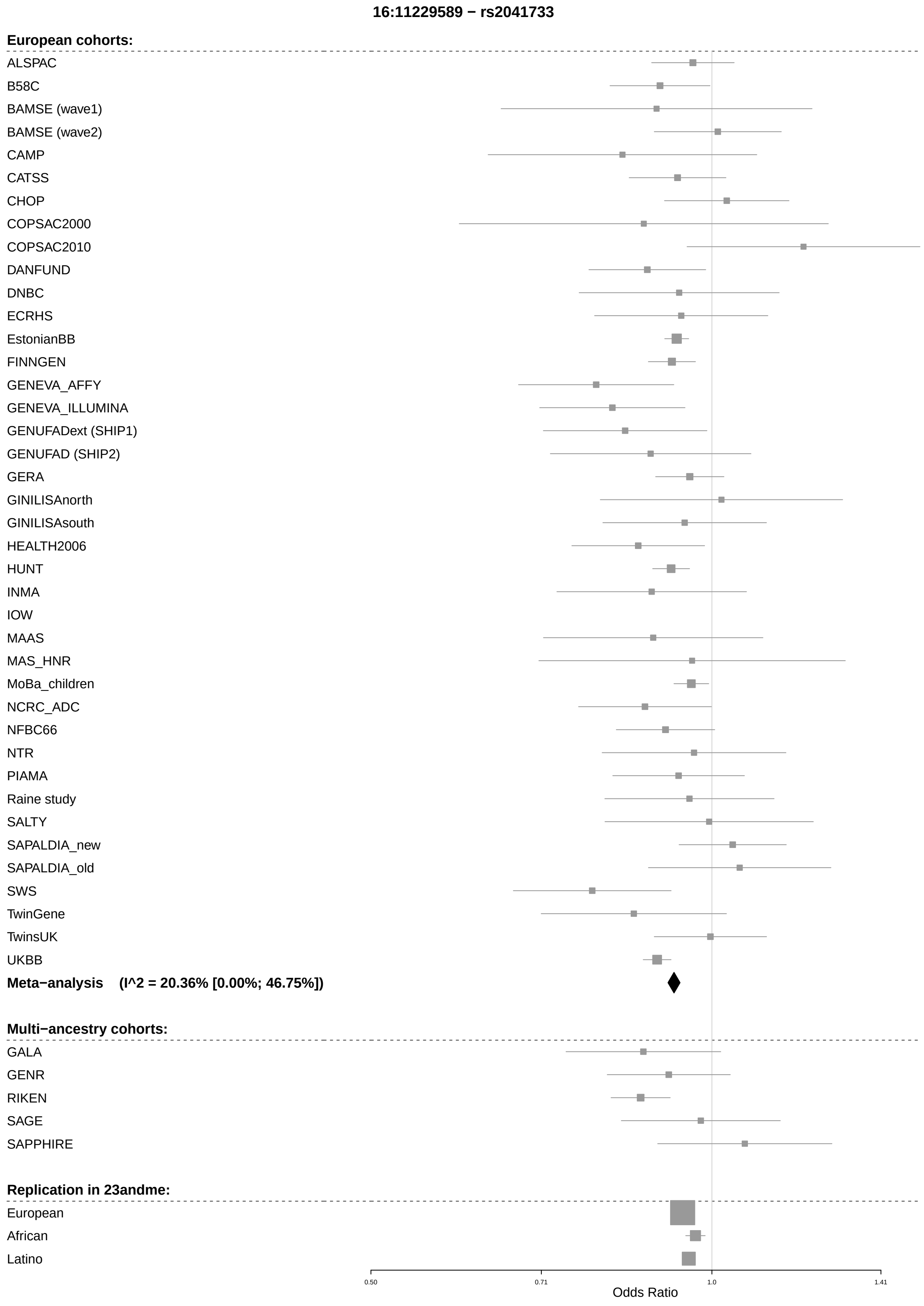
16:11229589 − rs2041733
European cohorts:
ALSPAC
B58C
BAMSE (wave1)
BAMSE (wave2)
CAMP
CATSS
CHOP
COPSAC2000
COPSAC2010
DANFUND
DNBC
ECRHS
EstonianBB
FINNGEN
GENEVA_AFFY
GENEVA_ILLUMINA
GENUFADext (SHIP1)
GENUFAD (SHIP2)
GERA
GINILISAnorth
GINILISAsouth
HEALTH2006
HUNT
INMA
IOW
MAAS
MAS_HNR
MoBa_children
NCRC_ADC
NFBC66
NTR
PIAMA
Raine study
SALTY
SAPALDIA_new
SAPALDIA_old
SWS
TwinGene
TwinsUK
UKBB
Meta−analysis (I^2 = 20.36% [0.00%; 46.75%])
Multi−ancestry cohorts:
GALA
GENR
RIKEN
SAGE
SAPPHIRE
Replication in 23andme:
European
African
Latino
0.50
0.71
1.0
1.41
Odds Ratio

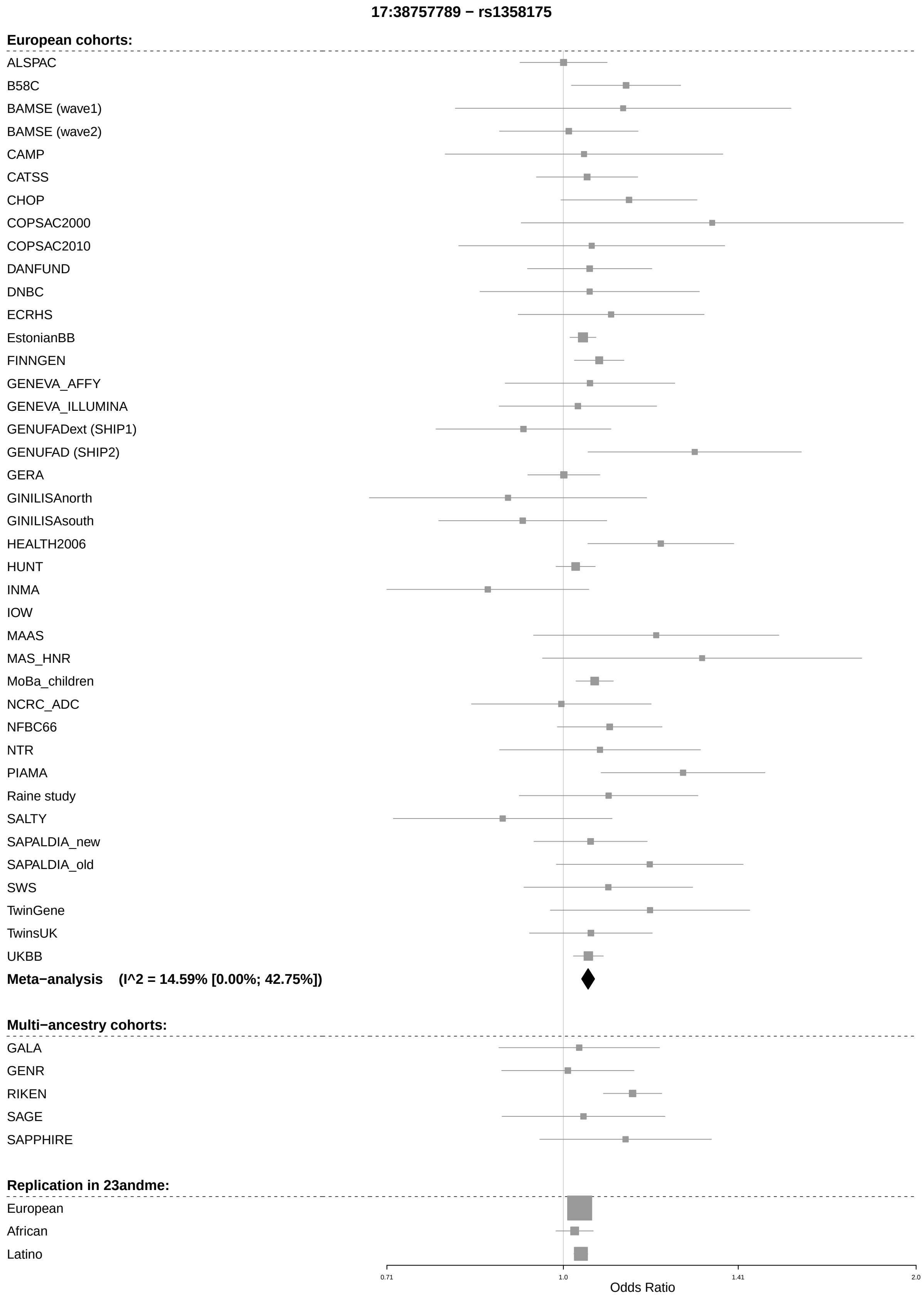
17:38757789 − rs1358175
European cohorts:
ALSPAC
B58C
BAMSE (wave1)
BAMSE (wave2)
CAMP
CATSS
CHOP
COPSAC2000
COPSAC2010
DANFUND
DNBC
ECRHS
EstonianBB
FINNGEN
GENEVA_AFFY
GENEVA_ILLUMINA
GENUFADext (SHIP1)
GENUFAD (SHIP2)
GERA
GINILISAnorth
GINILISAsouth
HEALTH2006
HUNT
INMA
IOW
MAAS
MAS_HNR
MoBa_children
NCRC_ADC
NFBC66
NTR
PIAMA
Raine study
SALTY
SAPALDIA_new
SAPALDIA_old
SWS
TwinGene
TwinsUK
UKBB
Meta−analysis (I^2 = 14.59% [0.00%; 42.75%])
Multi−ancestry cohorts:
GALA
GENR
RIKEN
SAGE
SAPPHIRE
Replication in 23andme:
European
African
Latino
0.71
1.0
1.41
2.0
Odds Ratio

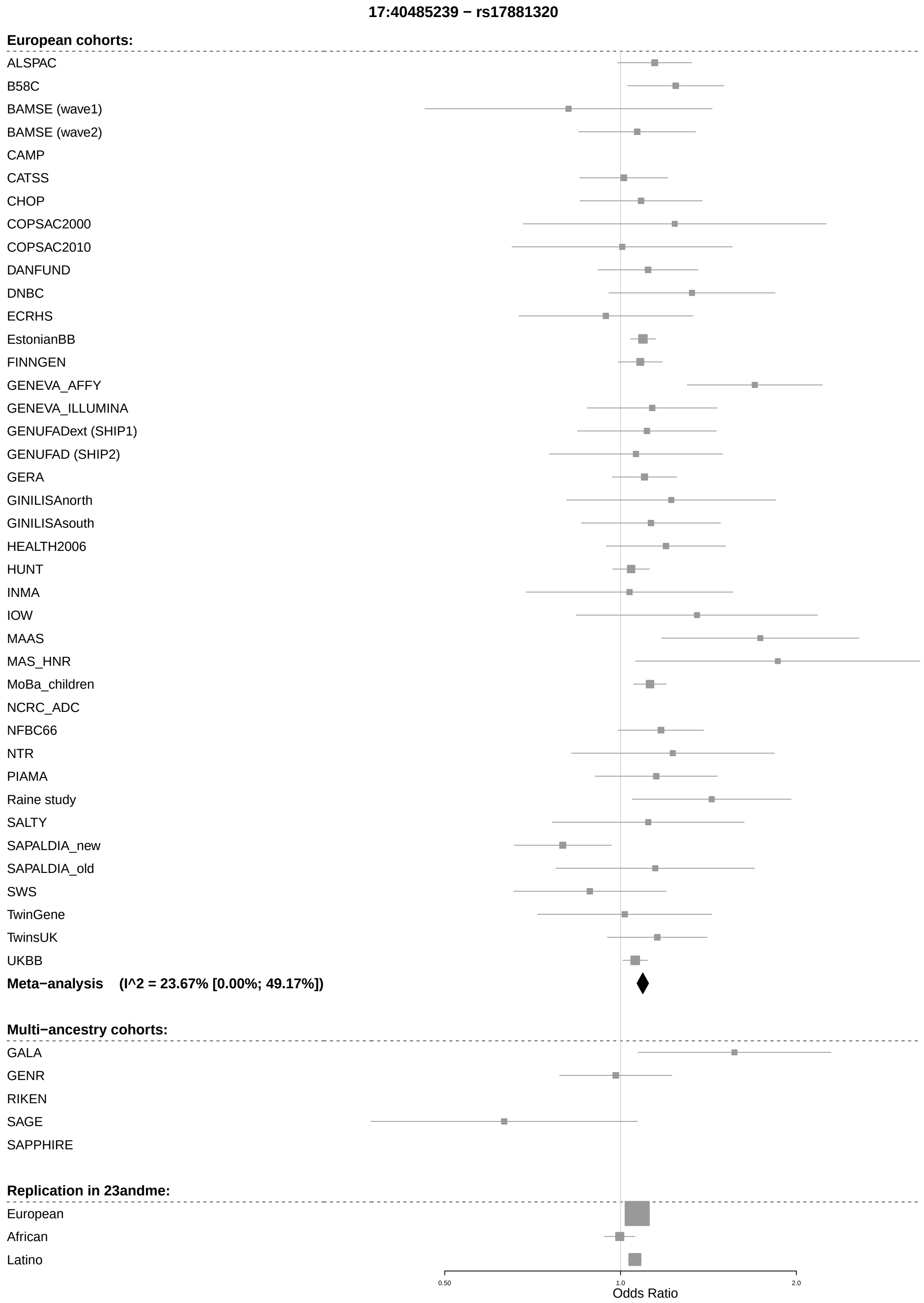

17:40485239 − rs17881320
European cohorts:
ALSPAC
B58C
BAMSE (wave1)
BAMSE (wave2)
CAMP
CATSS
CHOP
COPSAC2000
COPSAC2010
DANFUND
DNBC
ECRHS
EstonianBB
FINNGEN
GENEVA_AFFY
GENEVA_ILLUMINA
GENUFADext (SHIP1)
GENUFAD (SHIP2)
GERA
GINILISAnorth
GINILISAsouth
HEALTH2006
HUNT
INMA
IOW
MAAS
MAS_HNR
MoBa_children
NCRC_ADC
NFBC66
NTR
PIAMA
Raine study
SALTY
SAPALDIA_new
SAPALDIA_old
SWS
TwinGene
TwinsUK
UKBB
Meta−analysis (I^2 = 23.67% [0.00%; 49.17%])
Multi−ancestry cohorts:
GALA
GENR
RIKEN
SAGE
SAPPHIRE
Replication in 23andme:
European
African
Latino
0.50
1.0
2.0
Odds Ratio

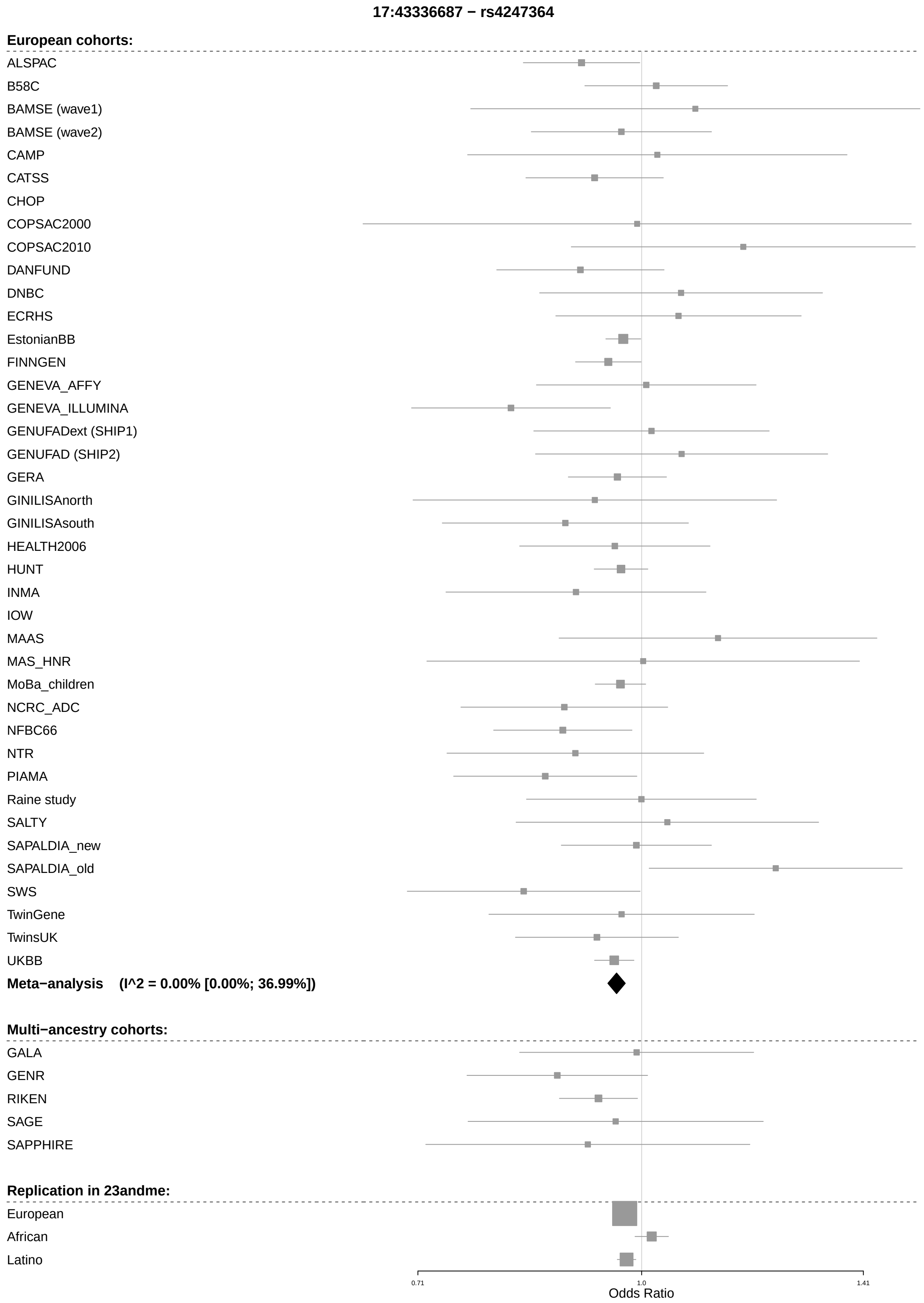

17:43336687 − rs4247364
European cohorts:
ALSPAC
B58C
BAMSE (wave1)
BAMSE (wave2)
CAMP
CATSS
CHOP
COPSAC2000
COPSAC2010
DANFUND
DNBC
ECRHS
EstonianBB
FINNGEN
GENEVA_AFFY
GENEVA_ILLUMINA
GENUFADext (SHIP1)
GENUFAD (SHIP2)
GERA
GINILISAnorth
GINILISAsouth
HEALTH2006
HUNT
INMA
IOW
MAAS
MAS_HNR
MoBa_children
NCRC_ADC
NFBC66
NTR
PIAMA
Raine study
SALTY
SAPALDIA_new
SAPALDIA_old
SWS
TwinGene
TwinsUK
UKBB
Meta−analysis (I^2 = 0.00% [0.00%; 36.99%])
Multi−ancestry cohorts:
GALA
GENR
RIKEN
SAGE
SAPPHIRE
Replication in 23andme:
European
African
Latino
0.71
1.0
1.41
Odds Ratio

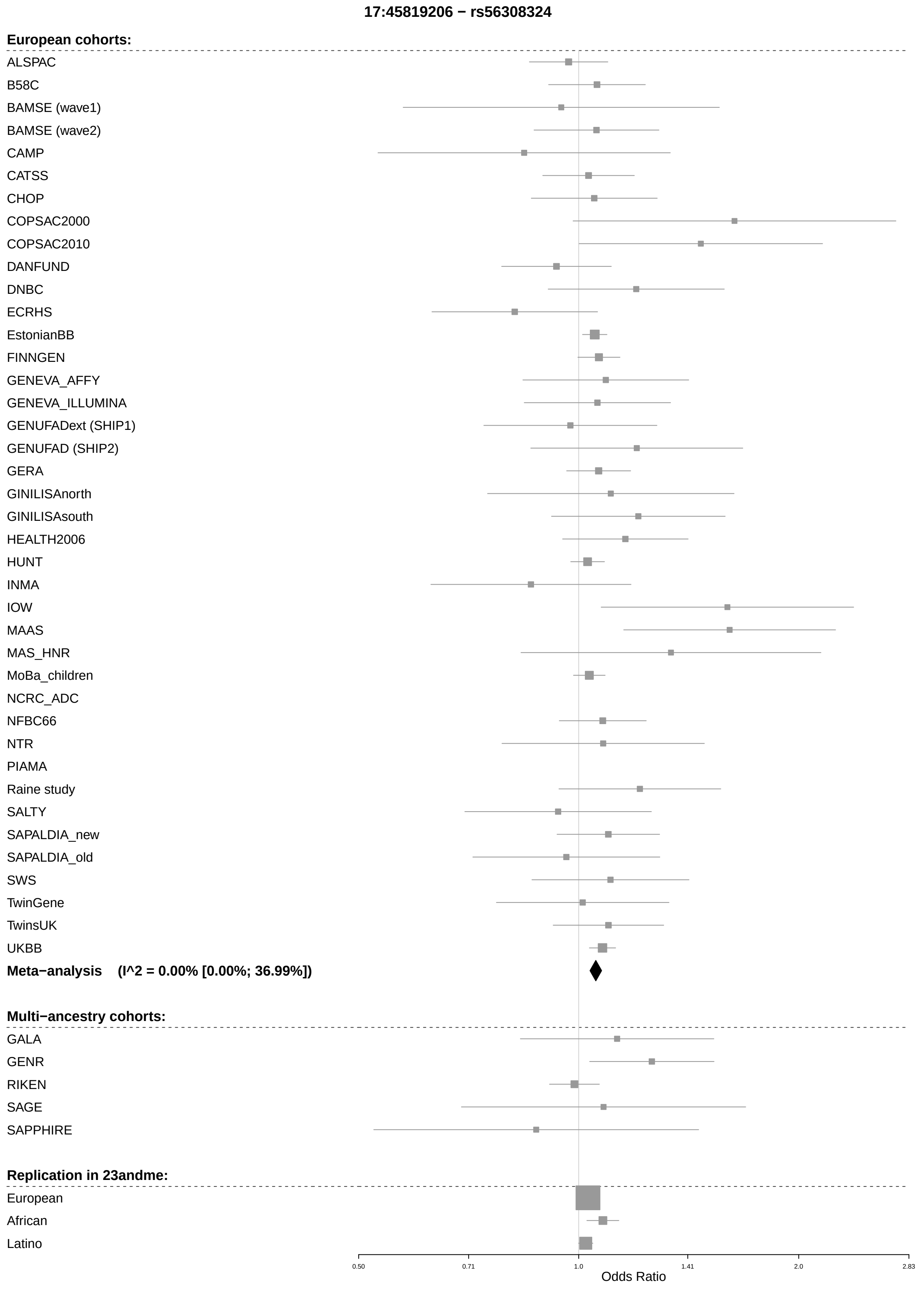
17:45819206 − rs56308324
European cohorts:
ALSPAC
B58C
BAMSE (wave1)
BAMSE (wave2)
CAMP
CATSS
CHOP
COPSAC2000
COPSAC2010
DANFUND
DNBC
ECRHS
EstonianBB
FINNGEN
GENEVA_AFFY
GENEVA_ILLUMINA
GENUFADext (SHIP1)
GENUFAD (SHIP2)
GERA
GINILISAnorth
GINILISAsouth
HEALTH2006
HUNT
INMA
IOW
MAAS
MAS_HNR
MoBa_children
NCRC_ADC
NFBC66
NTR
PIAMA
Raine study
SALTY
SAPALDIA_new
SAPALDIA_old
SWS
TwinGene
TwinsUK
UKBB
Meta−analysis (I^2 = 0.00% [0.00%; 36.99%])
Multi−ancestry cohorts:
GALA
GENR
RIKEN
SAGE
SAPPHIRE
Replication in 23andme:
European
African
Latino
0.50
0.71
1.0
1.41
2.0
2.83
Odds Ratio

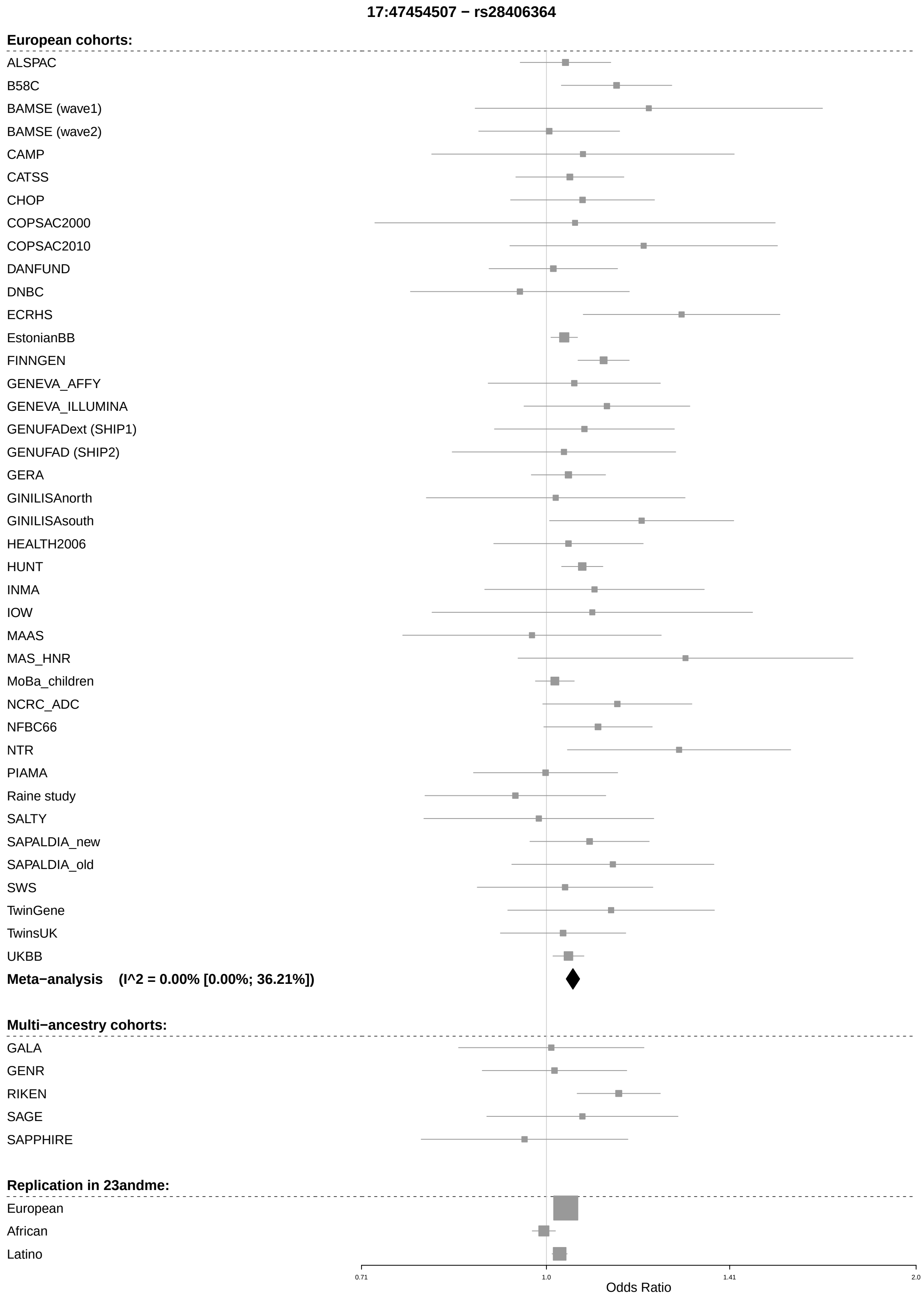
17:47454507 − rs28406364
European cohorts:
ALSPAC
B58C
BAMSE (wave1)
BAMSE (wave2)
CAMP
CATSS
CHOP
COPSAC2000
COPSAC2010
DANFUND
DNBC
ECRHS
EstonianBB
FINNGEN
GENEVA_AFFY
GENEVA_ILLUMINA
GENUFADext (SHIP1)
GENUFAD (SHIP2)
GERA
GINILISAnorth
GINILISAsouth
HEALTH2006
HUNT
INMA
IOW
MAAS
MAS_HNR
MoBa_children
NCRC_ADC
NFBC66
NTR
PIAMA
Raine study
SALTY
SAPALDIA_new
SAPALDIA_old
SWS
TwinGene
TwinsUK
UKBB
Meta−analysis (I^2 = 0.00% [0.00%; 36.21%])
Multi−ancestry cohorts:
GALA
GENR
RIKEN
SAGE
SAPPHIRE
Replication in 23andme:
European
African
Latino
0.71
1.0
1.41
2.0
Odds Ratio

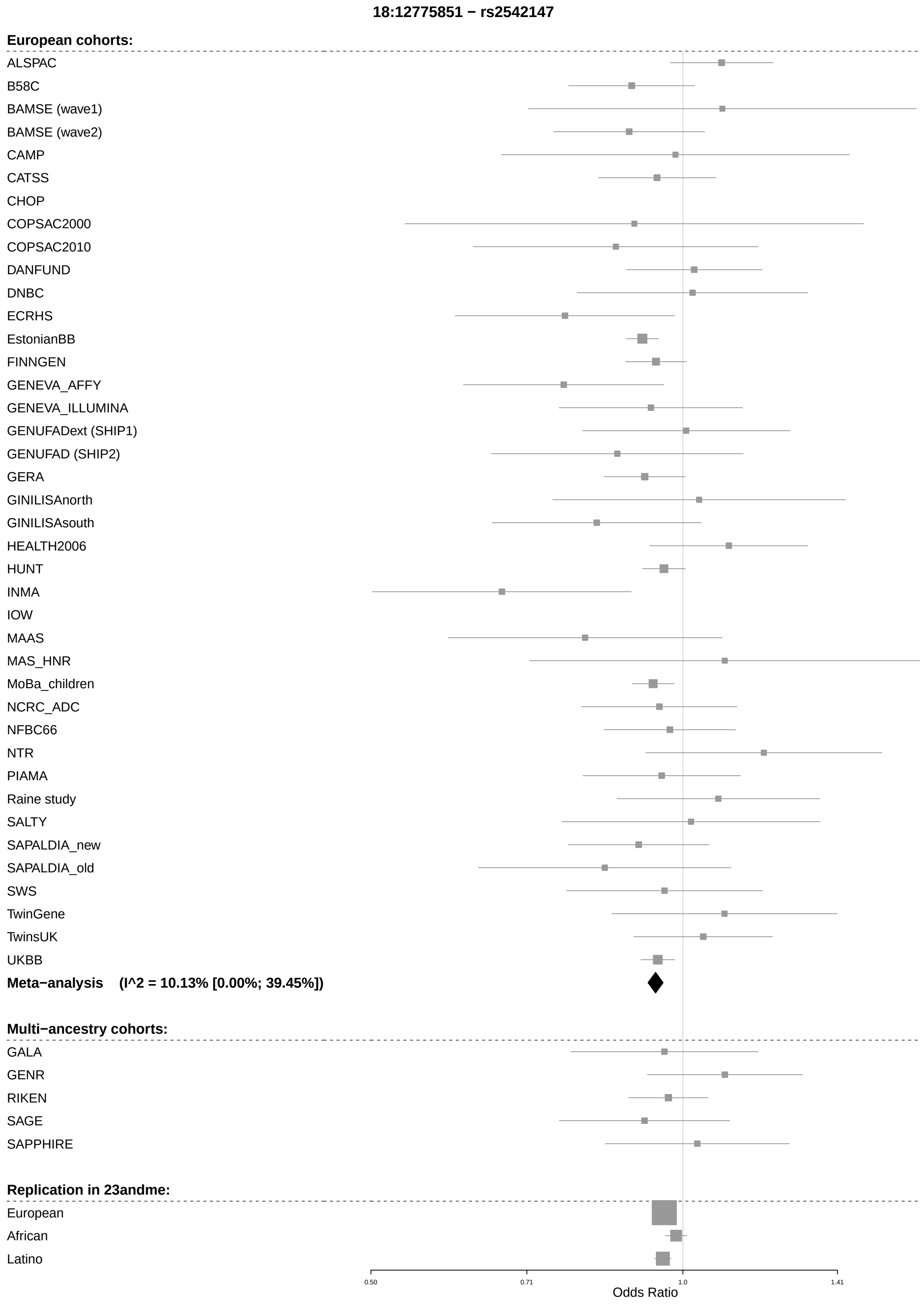
18:12775851 − rs2542147
European cohorts:
ALSPAC
B58C
BAMSE (wave1)
BAMSE (wave2)
CAMP
CATSS
CHOP
COPSAC2000
COPSAC2010
DANFUND
DNBC
ECRHS
EstonianBB
FINNGEN
GENEVA_AFFY
GENEVA_ILLUMINA
GENUFADext (SHIP1)
GENUFAD (SHIP2)
GERA
GINILISAnorth
GINILISAsouth
HEALTH2006
HUNT
INMA
IOW
MAAS
MAS_HNR
MoBa_children
NCRC_ADC
NFBC66
NTR
PIAMA
Raine study
SALTY
SAPALDIA_new
SAPALDIA_old
SWS
TwinGene
TwinsUK
UKBB
Meta−analysis (I^2 = 10.13% [0.00%; 39.45%])
Multi−ancestry cohorts:
GALA
GENR
RIKEN
SAGE
SAPPHIRE
Replication in 23andme:
European
African
Latino
0.50
0.71
1.0
1.41
Odds Ratio

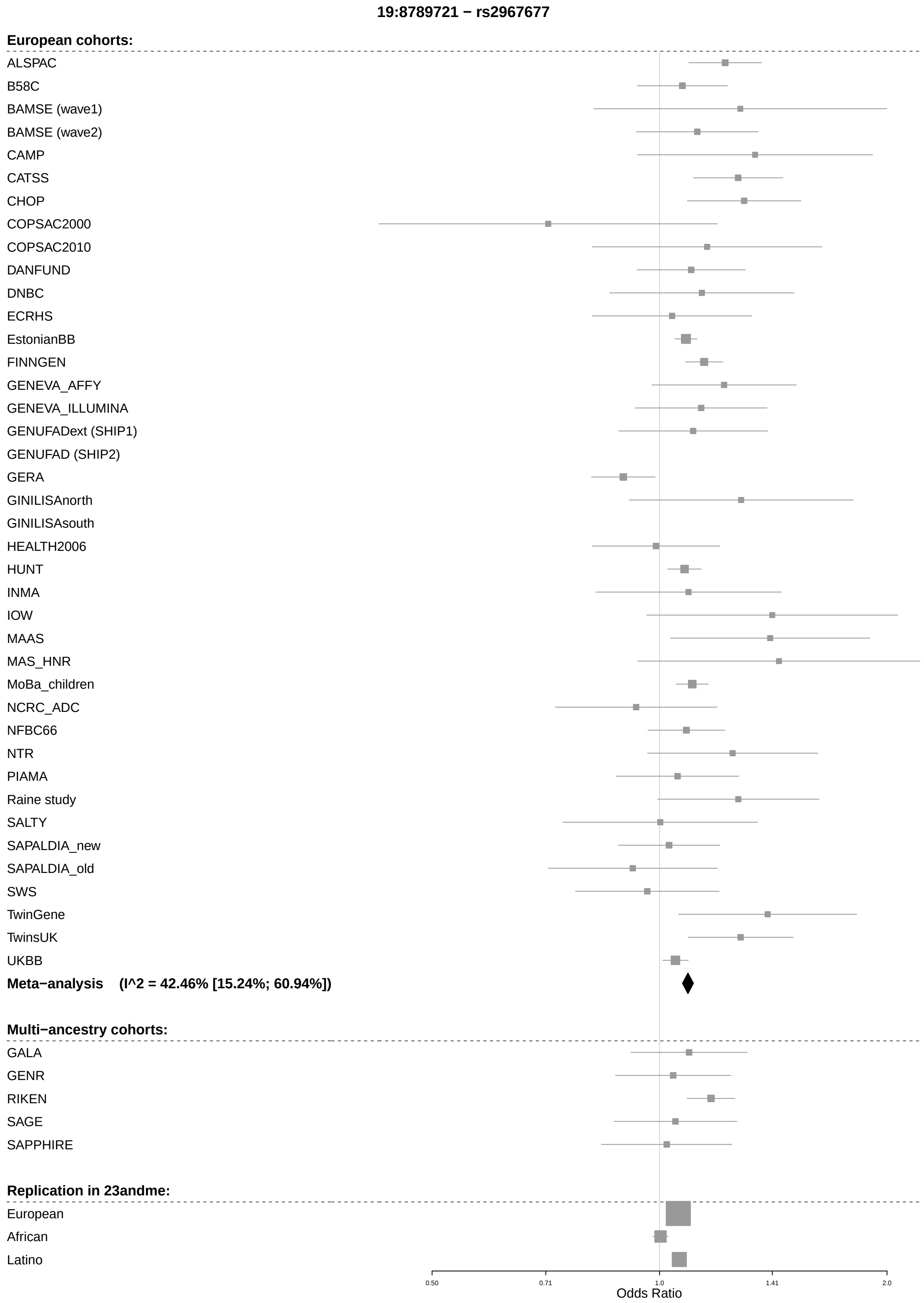
19:8789721 − rs2967677
European cohorts:
ALSPAC
B58C
BAMSE (wave1)
BAMSE (wave2)
CAMP
CATSS
CHOP
COPSAC2000
COPSAC2010
DANFUND
DNBC
ECRHS
EstonianBB
FINNGEN
GENEVA_AFFY
GENEVA_ILLUMINA
GENUFADext (SHIP1)
GENUFAD (SHIP2)
GERA
GINILISAnorth
GINILISAsouth
HEALTH2006
HUNT
INMA
IOW
MAAS
MAS_HNR
MoBa_children
NCRC_ADC
NFBC66
NTR
PIAMA
Raine study
SALTY
SAPALDIA_new
SAPALDIA_old
SWS
TwinGene
TwinsUK
UKBB
Meta−analysis (I^2 = 42.46% [15.24%; 60.94%])
Multi−ancestry cohorts:
GALA
GENR
RIKEN
SAGE
SAPPHIRE
Replication in 23andme:
European
African
Latino
0.50
0.71
1.0
1.41
2.0
Odds Ratio

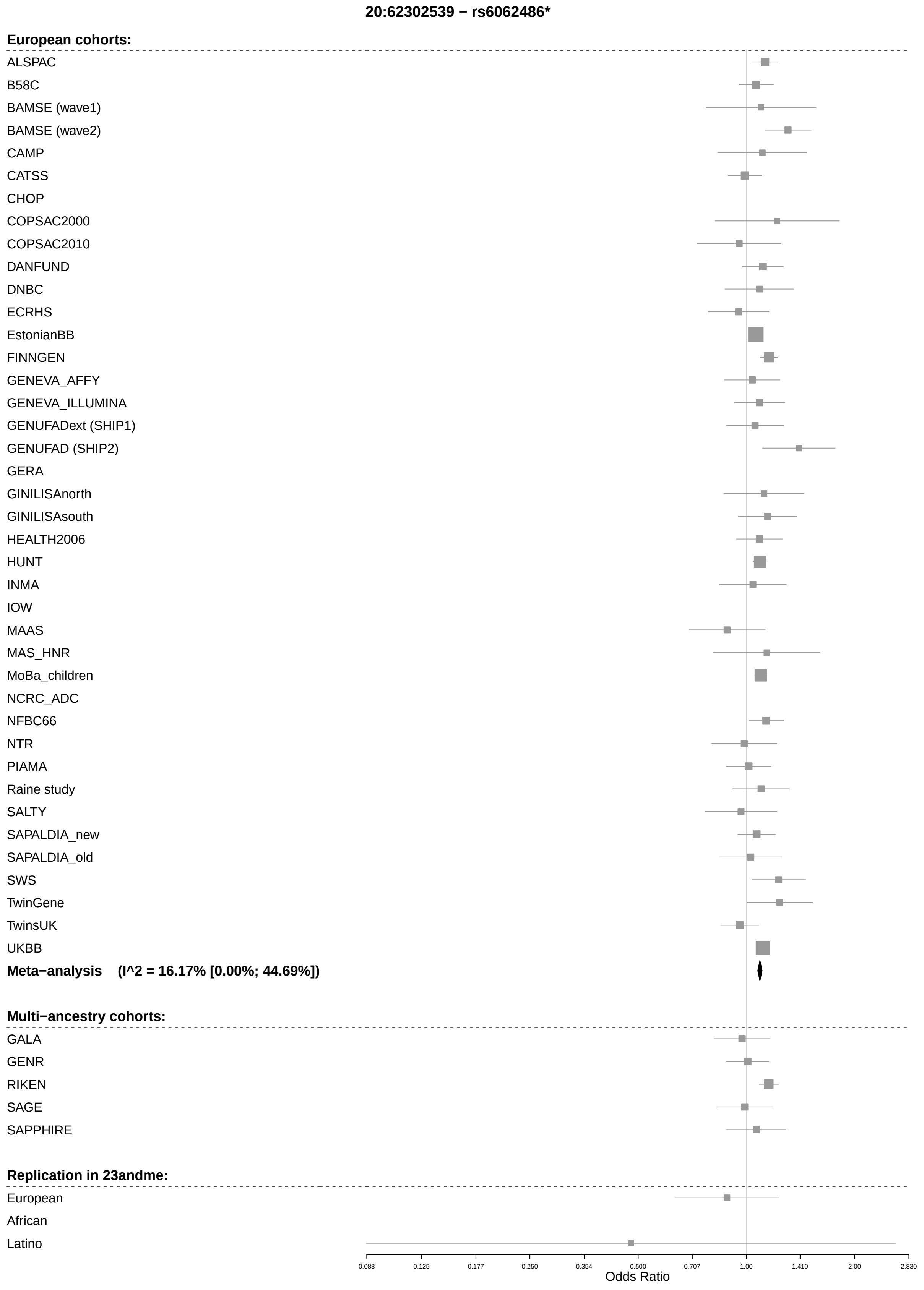
20:62302539 − rs6062486*
European cohorts:
ALSPAC
B58C
BAMSE (wave1)
BAMSE (wave2)
CAMP
CATSS
CHOP
COPSAC2000
COPSAC2010
DANFUND
DNBC
ECRHS
EstonianBB
FINNGEN
GENEVA_AFFY
GENEVA_ILLUMINA
GENUFADext (SHIP1)
GENUFAD (SHIP2)
GERA
GINILISAnorth
GINILISAsouth
HEALTH2006
HUNT
INMA
IOW
MAAS
MAS_HNR
MoBa_children
NCRC_ADC
NFBC66
NTR
PIAMA
Raine study
SALTY
SAPALDIA_new
SAPALDIA_old
SWS
TwinGene
TwinsUK
UKBB
Meta−analysis (I^2 = 16.17% [0.00%; 44.69%])
Multi−ancestry cohorts:
GALA
GENR
RIKEN
SAGE
SAPPHIRE
Replication in 23andme:
European
African
Latino
0.088
0.125
0.177
0.250
0.354
0.500
0.707
1.00
1.410
2.00
2.830
Odds Ratio

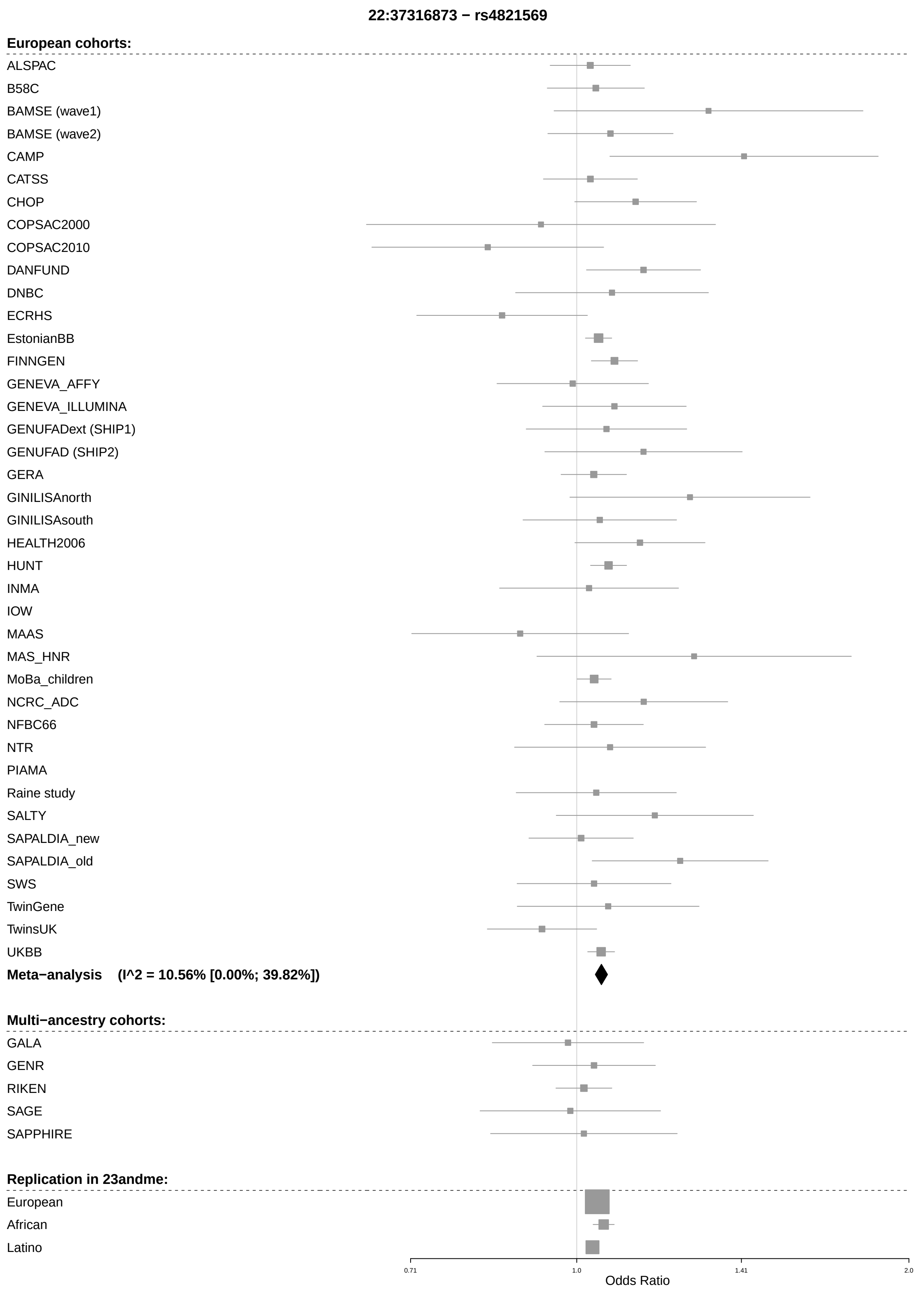
22:37316873 − rs4821569
European cohorts:
ALSPAC
B58C
BAMSE (wave1)
BAMSE (wave2)
CAMP
CATSS
CHOP
COPSAC2000
COPSAC2010
DANFUND
DNBC
ECRHS
EstonianBB
FINNGEN
GENEVA_AFFY
GENEVA_ILLUMINA
GENUFADext (SHIP1)
GENUFAD (SHIP2)
GERA
GINILISAnorth
GINILISAsouth
HEALTH2006
HUNT
INMA
IOW
MAAS
MAS_HNR
MoBa_children
NCRC_ADC
NFBC66
NTR
PIAMA
Raine study
SALTY
SAPALDIA_new
SAPALDIA_old
SWS
TwinGene
TwinsUK
UKBB
Meta−analysis (I^2 = 10.56% [0.00%; 39.82%])
Multi−ancestry cohorts:
GALA
GENR
RIKEN
SAGE
SAPPHIRE
Replication in 23andme:
European
African
Latino
0.71
1.0
1.41
2.0
Odds Ratio

#### Additional SNPs from Multi-ancestry analysis

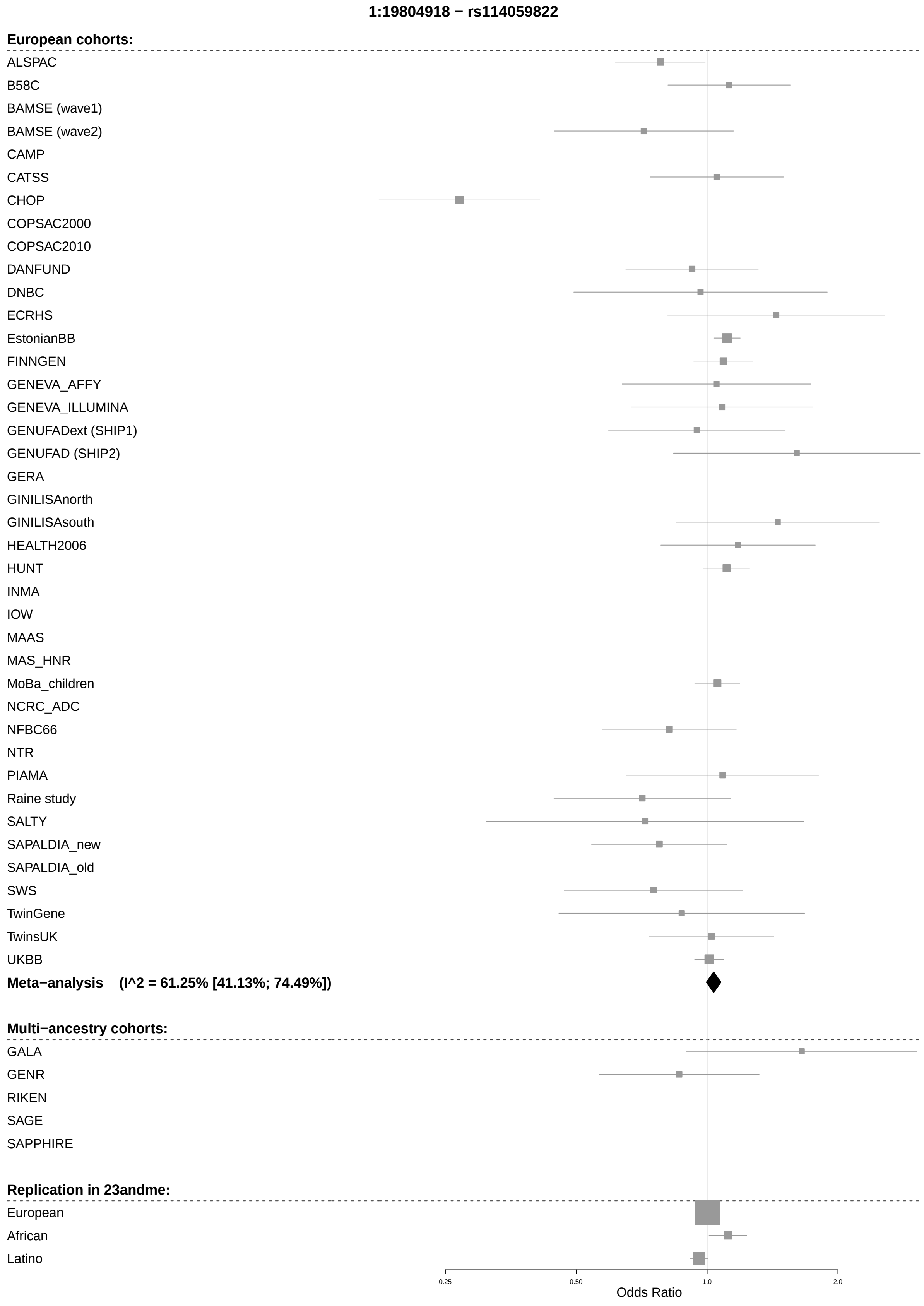

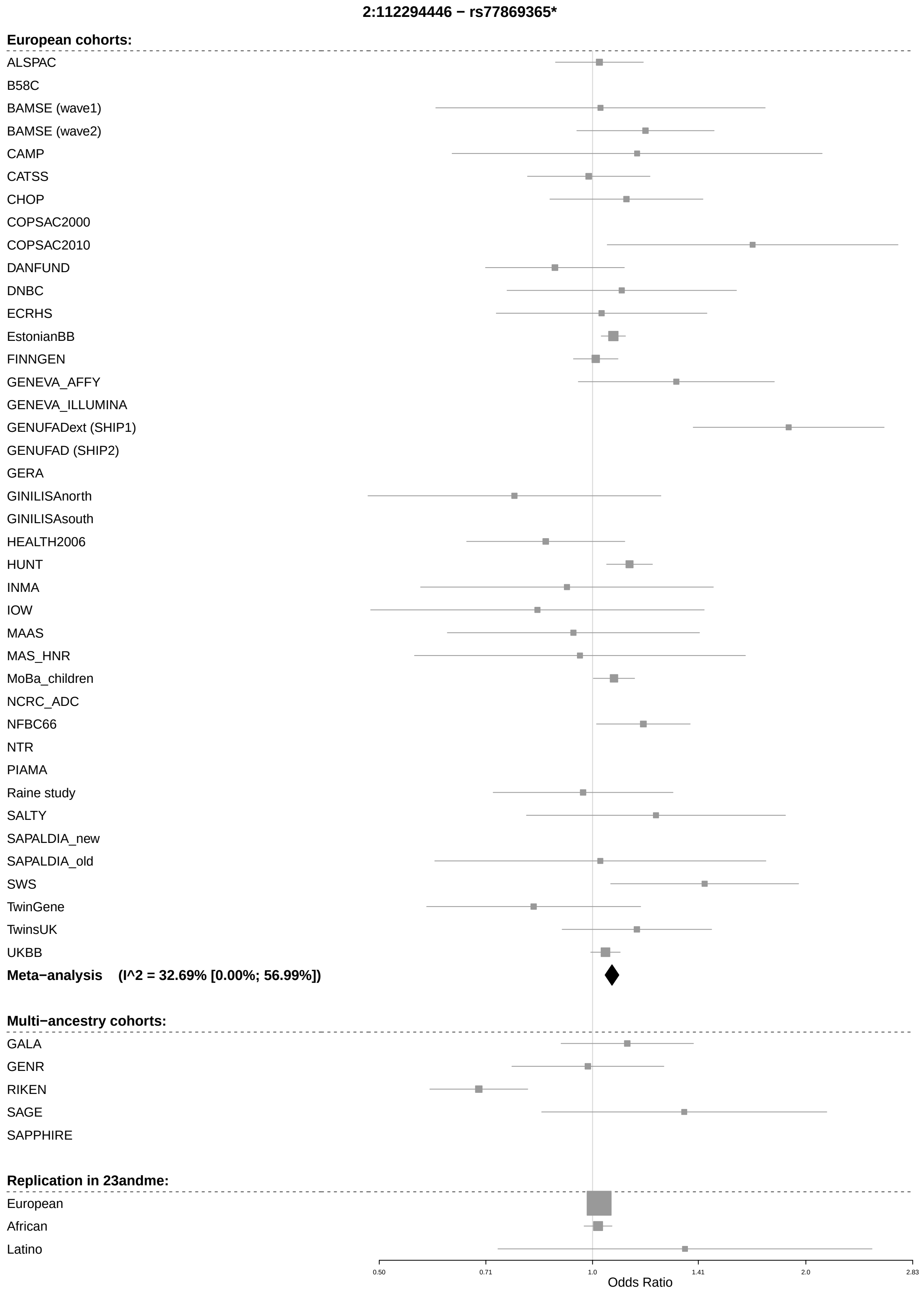
2:112294446 − rs77869365*
European cohorts:
ALSPAC
B58C
BAMSE (wave1)
BAMSE (wave2)
CAMP
CATSS
CHOP
COPSAC2000
COPSAC2010
DANFUND
DNBC
ECRHS
EstonianBB
FINNGEN
GENEVA_AFFY
GENEVA_ILLUMINA
GENUFADext (SHIP1)
GENUFAD (SHIP2)
GERA
GINILISAnorth
GINILISAsouth
HEALTH2006
HUNT
INMA
IOW
MAAS
MAS_HNR
MoBa_children
NCRC_ADC
NFBC66
NTR
PIAMA
Raine study
SALTY
SAPALDIA_new
SAPALDIA_old
SWS
TwinGene
TwinsUK
UKBB
Meta−analysis (I^2 = 32.69% [0.00%; 56.99%])
Multi−ancestry cohorts:
GALA
GENR
RIKEN
SAGE
SAPPHIRE
Replication in 23andme:
European
African
Latino
0.50
0.71
1.0
1.41
2.0
2.83
Odds Ratio

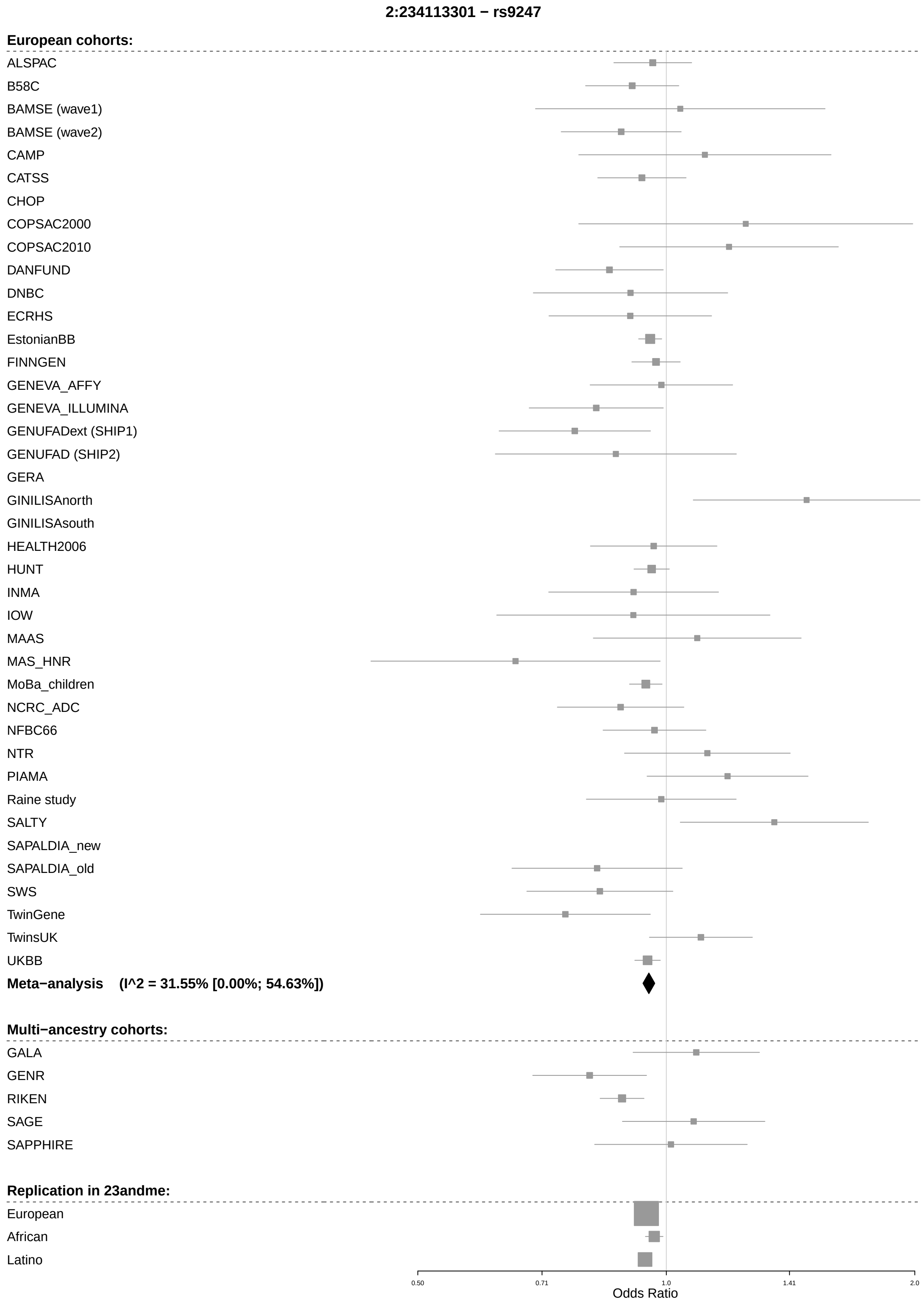
2:234113301 − rs9247
European cohorts:
ALSPAC
B58C
BAMSE (wave1)
BAMSE (wave2)
CAMP
CATSS
CHOP
COPSAC2000
COPSAC2010
DANFUND
DNBC
ECRHS
EstonianBB
FINNGEN
GENEVA_AFFY
GENEVA_ILLUMINA
GENUFADext (SHIP1)
GENUFAD (SHIP2)
GERA
GINILISAnorth
GINILISAsouth
HEALTH2006
HUNT
INMA
IOW
MAAS
MAS_HNR
MoBa_children
NCRC_ADC
NFBC66
NTR
PIAMA
Raine study
SALTY
SAPALDIA_new
SAPALDIA_old
SWS
TwinGene
TwinsUK
UKBB
Meta−analysis (I^2 = 31.55% [0.00%; 54.63%])
Multi−ancestry cohorts:
GALA
GENR
RIKEN
SAGE
SAPPHIRE
Replication in 23andme:
European
African
Latino
0.50
0.71
1.0
1.41
2.0
Odds Ratio

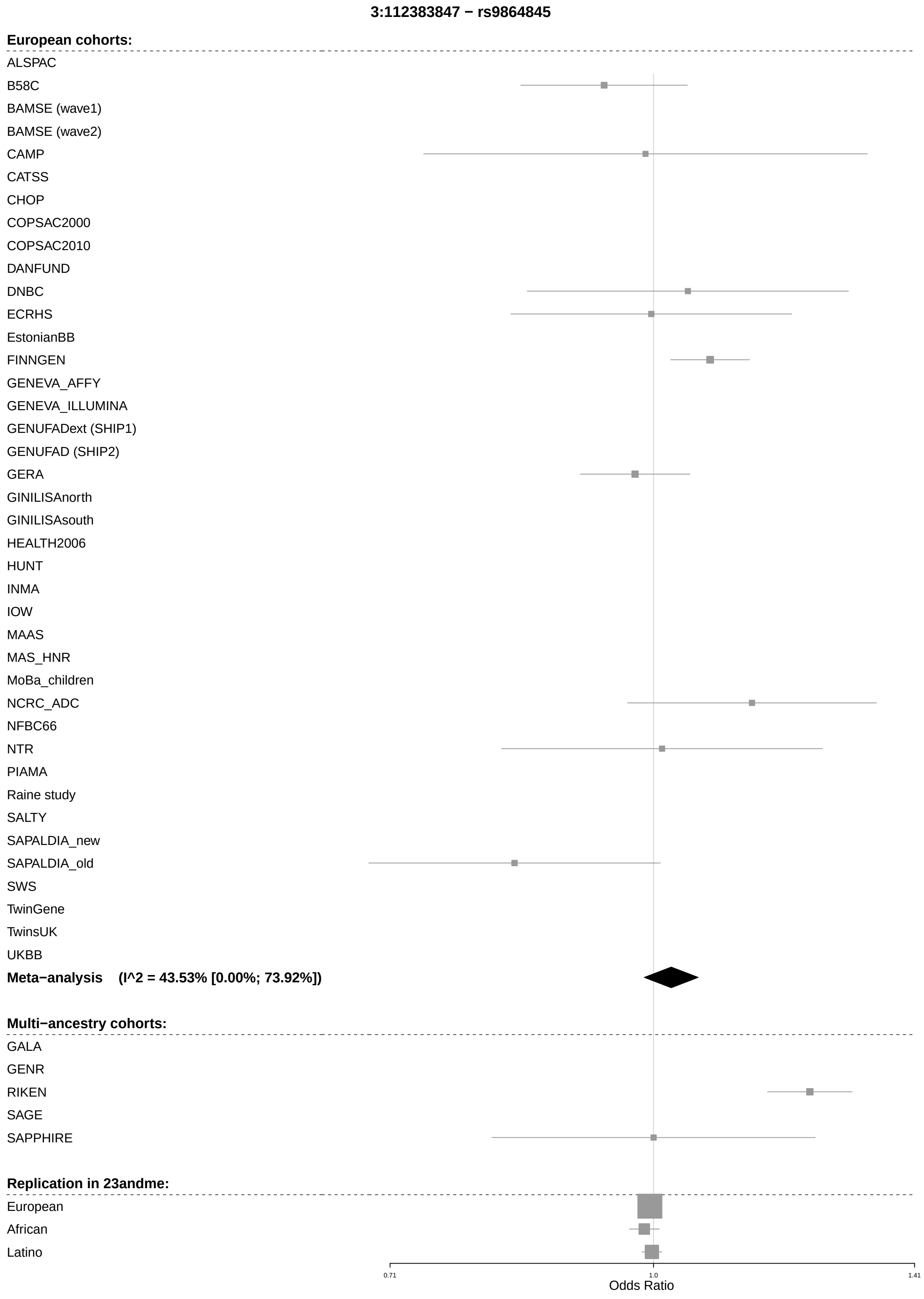
3:112383847 − rs9864845
European cohorts:
ALSPAC
B58C
BAMSE (wave1)
BAMSE (wave2)
CAMP
CATSS
CHOP
COPSAC2000
COPSAC2010
DANFUND
DNBC
ECRHS
EstonianBB
FINNGEN
GENEVA_AFFY
GENEVA_ILLUMINA
GENUFADext (SHIP1)
GENUFAD (SHIP2)
GERA
GINILISAnorth
GINILISAsouth
HEALTH2006
HUNT
INMA
IOW
MAAS
MAS_HNR
MoBa_children
NCRC_ADC
NFBC66
NTR
PIAMA
Raine study
SALTY
SAPALDIA_new
SAPALDIA_old
SWS
TwinGene
TwinsUK
UKBB
Meta−analysis (I^2 = 43.53% [0.00%; 73.92%])
Multi−ancestry cohorts:
GALA
GENR
RIKEN
SAGE
SAPPHIRE
Replication in 23andme:
European
African
Latino
0.71
1.0
1.41
Odds Ratio

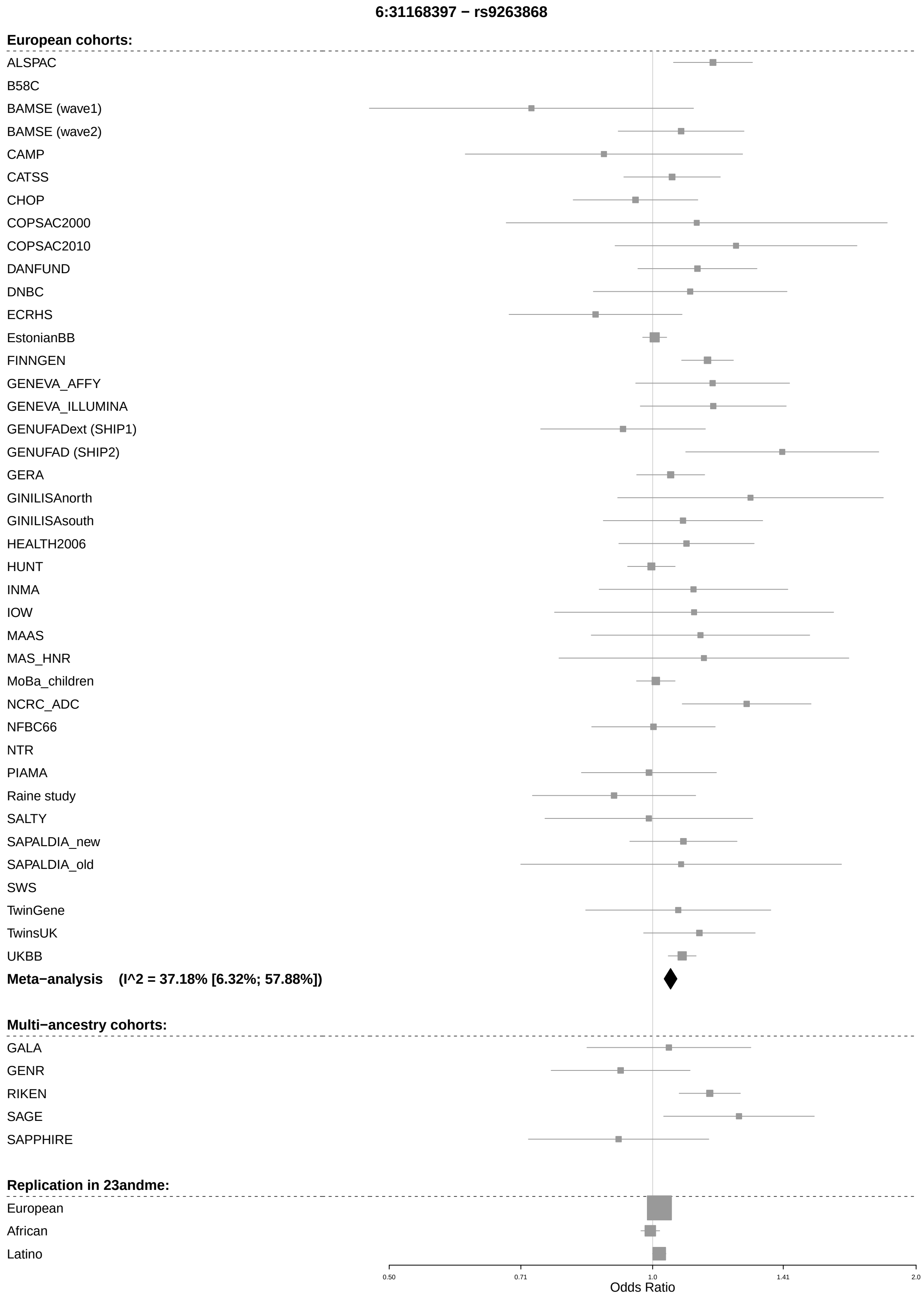
6:31168397 − rs9263868
European cohorts:
ALSPAC
B58C
BAMSE (wave1)
BAMSE (wave2)
CAMP
CATSS
CHOP
COPSAC2000
COPSAC2010
DANFUND
DNBC
ECRHS
EstonianBB
FINNGEN
GENEVA_AFFY
GENEVA_ILLUMINA
GENUFADext (SHIP1)
GENUFAD (SHIP2)
GERA
GINILISAnorth
GINILISAsouth
HEALTH2006
HUNT
INMA
IOW
MAAS
MAS_HNR
MoBa_children
NCRC_ADC
NFBC66
NTR
PIAMA
Raine study
SALTY
SAPALDIA_new
SAPALDIA_old
SWS
TwinGene
TwinsUK
UKBB
Meta−analysis (I^2 = 37.18% [6.32%; 57.88%])
Multi−ancestry cohorts:
GALA
GENR
RIKEN
SAGE
SAPPHIRE
Replication in 23andme:
European
African
Latino
0.50
0.71
1.0
1.41
2.0
Odds Ratio

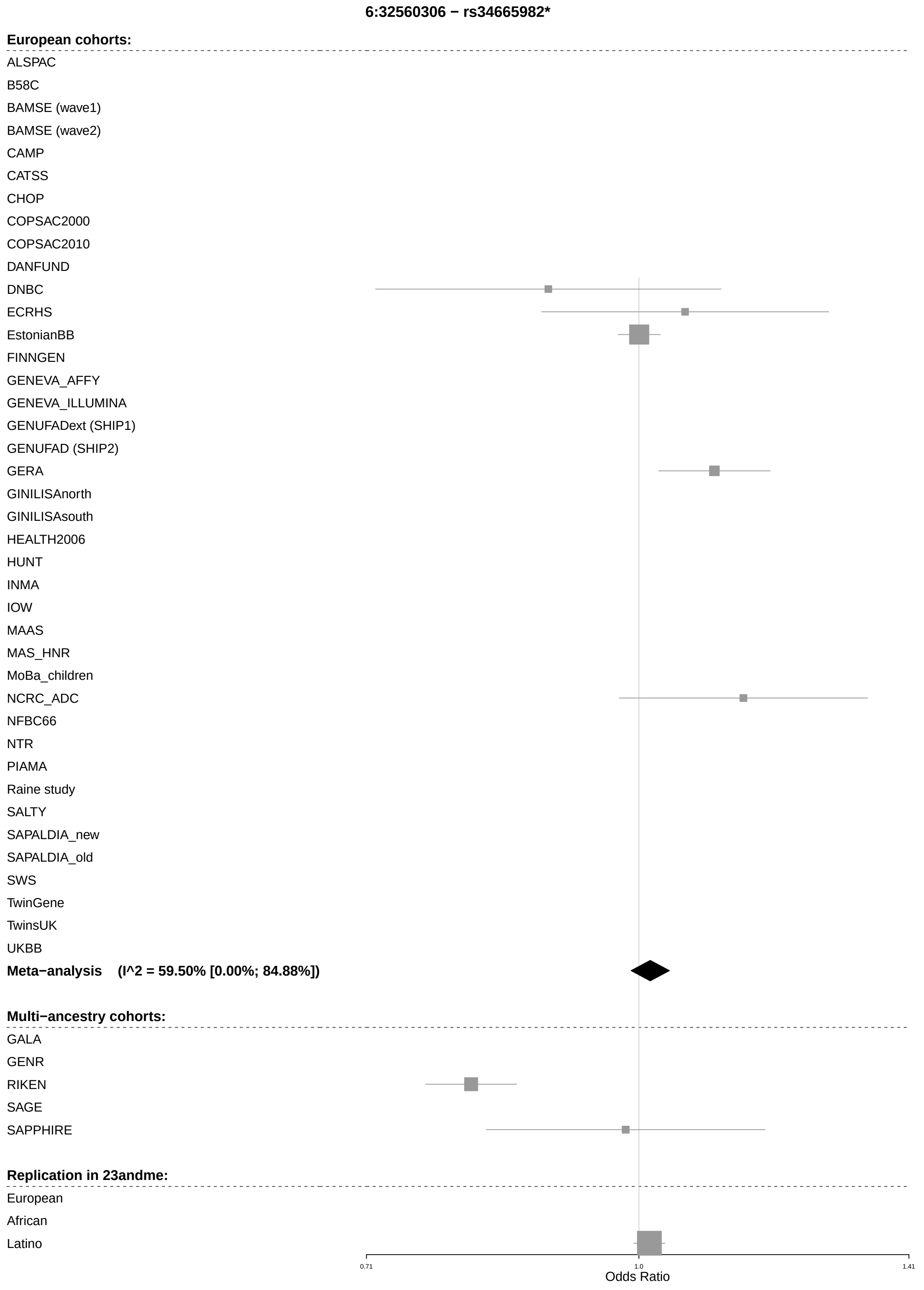

6:32560306 − rs34665982*
European cohorts:
ALSPAC
B58C
BAMSE (wave1)
BAMSE (wave2)
CAMP
CATSS
CHOP
COPSAC2000
COPSAC2010
DANFUND
DNBC
ECRHS
EstonianBB
FINNGEN
GENEVA_AFFY
GENEVA_ILLUMINA
GENUFADext (SHIP1)
GENUFAD (SHIP2)
GERA
GINILISAnorth
GINILISAsouth
HEALTH2006
HUNT
INMA
IOW
MAAS
MAS_HNR
MoBa_children
NCRC_ADC
NFBC66
NTR
PIAMA
Raine study
SALTY
SAPALDIA_new
SAPALDIA_old
SWS
TwinGene
TwinsUK
UKBB
Meta−analysis (I^2 = 59.50% [0.00%; 84.88%])
Multi−ancestry cohorts:
GALA
GENR
RIKEN
SAGE
SAPPHIRE
Replication in 23andme:
European
African
Latino
0.71
1.0
1.41
Odds Ratio

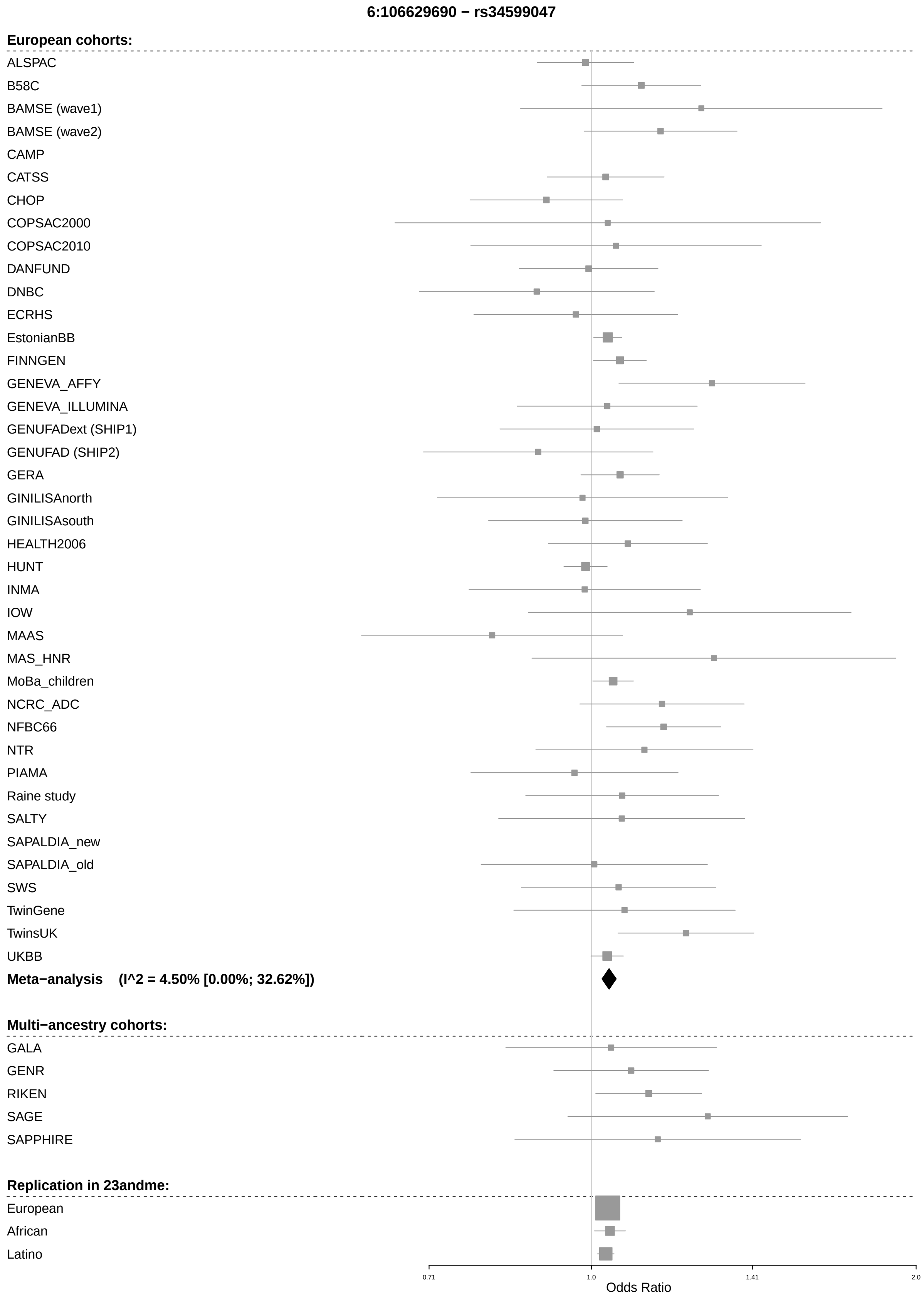

6:106629690 − rs34599047
European cohorts:
ALSPAC
B58C
BAMSE (wave1)
BAMSE (wave2)
CAMP
CATSS
CHOP
COPSAC2000
COPSAC2010
DANFUND
DNBC
ECRHS
EstonianBB
FINNGEN
GENEVA_AFFY
GENEVA_ILLUMINA
GENUFADext (SHIP1)
GENUFAD (SHIP2)
GERA
GINILISAnorth
GINILISAsouth
HEALTH2006
HUNT
INMA
IOW
MAAS
MAS_HNR
MoBa_children
NCRC_ADC
NFBC66
NTR
PIAMA
Raine study
SALTY
SAPALDIA_new
SAPALDIA_old
SWS
TwinGene
TwinsUK
UKBB
Meta−analysis (I^2 = 4.50% [0.00%; 32.62%])
Multi−ancestry cohorts:
GALA
GENR
RIKEN
SAGE
SAPPHIRE
Replication in 23andme:
European
African
Latino
0.71
1.0
1.41
2.0
Odds Ratio

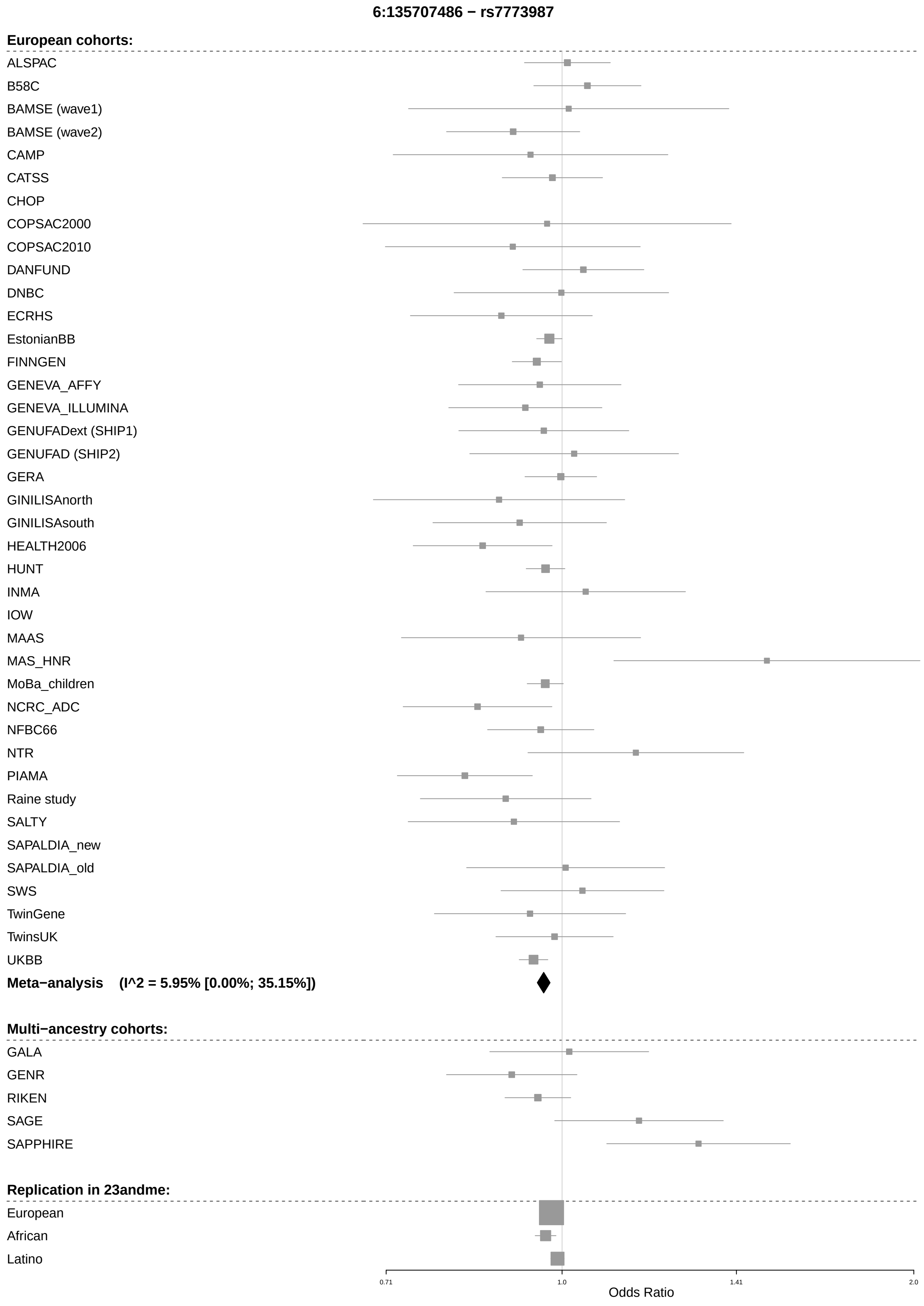
6:135707486 − rs7773987
European cohorts:
ALSPAC
B58C
BAMSE (wave1)
BAMSE (wave2)
CAMP
CATSS
CHOP
COPSAC2000
COPSAC2010
DANFUND
DNBC
ECRHS
EstonianBB
FINNGEN
GENEVA_AFFY
GENEVA_ILLUMINA
GENUFADext (SHIP1)
GENUFAD (SHIP2)
GERA
GINILISAnorth
GINILISAsouth
HEALTH2006
HUNT
INMA
IOW
MAAS
MAS_HNR
MoBa_children
NCRC_ADC
NFBC66
NTR
PIAMA
Raine study
SALTY
SAPALDIA_new
SAPALDIA_old
SWS
TwinGene
TwinsUK
UKBB
Meta−analysis (I^2 = 5.95% [0.00%; 35.15%])
Multi−ancestry cohorts:
GALA
GENR
RIKEN
SAGE
SAPPHIRE
Replication in 23andme:
European
African
Latino
0.71
1.0
1.41
2.0
Odds Ratio

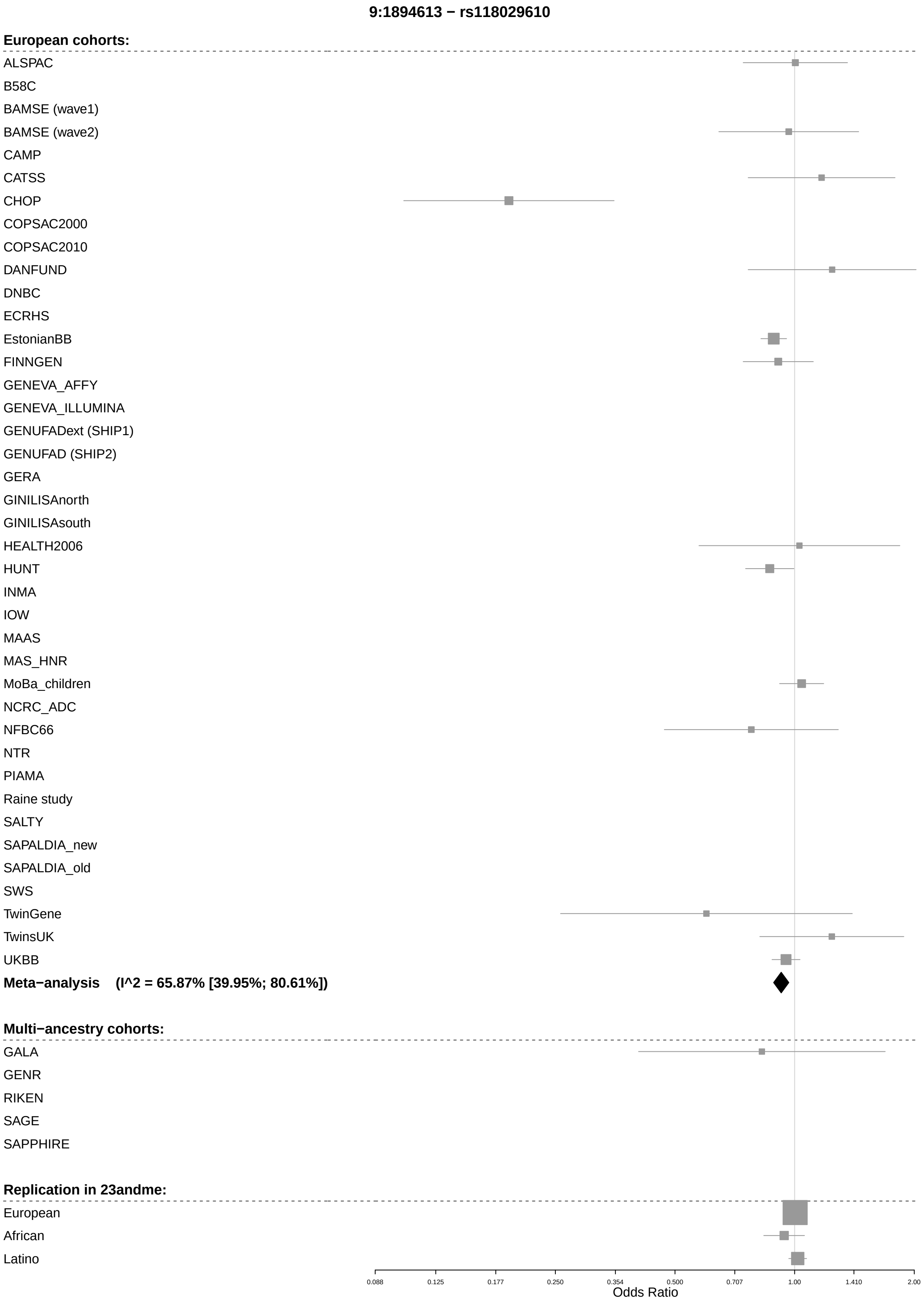
9:1894613 − rs118029610
European cohorts:
ALSPAC
B58C
BAMSE (wave1)
BAMSE (wave2)
CAMP
CATSS
CHOP
COPSAC2000
COPSAC2010
DANFUND
DNBC
ECRHS
EstonianBB
FINNGEN
GENEVA_AFFY
GENEVA_ILLUMINA
GENUFADext (SHIP1)
GENUFAD (SHIP2)
GERA
GINILISAnorth
GINILISAsouth
HEALTH2006
HUNT
INMA
IOW
MAAS
MAS_HNR
MoBa_children
NCRC_ADC
NFBC66
NTR
PIAMA
Raine study
SALTY
SAPALDIA_new
SAPALDIA_old
SWS
TwinGene
TwinsUK
UKBB
Meta−analysis (I^2 = 65.87% [39.95%; 80.61%])
Multi−ancestry cohorts:
GALA
GENR
RIKEN
SAGE
SAPPHIRE
Replication in 23andme:
European
African
Latino
0.088
0.125
0.177
0.250
0.354
0.500
0.707
1.00
1.410
2.00
Odds Ratio

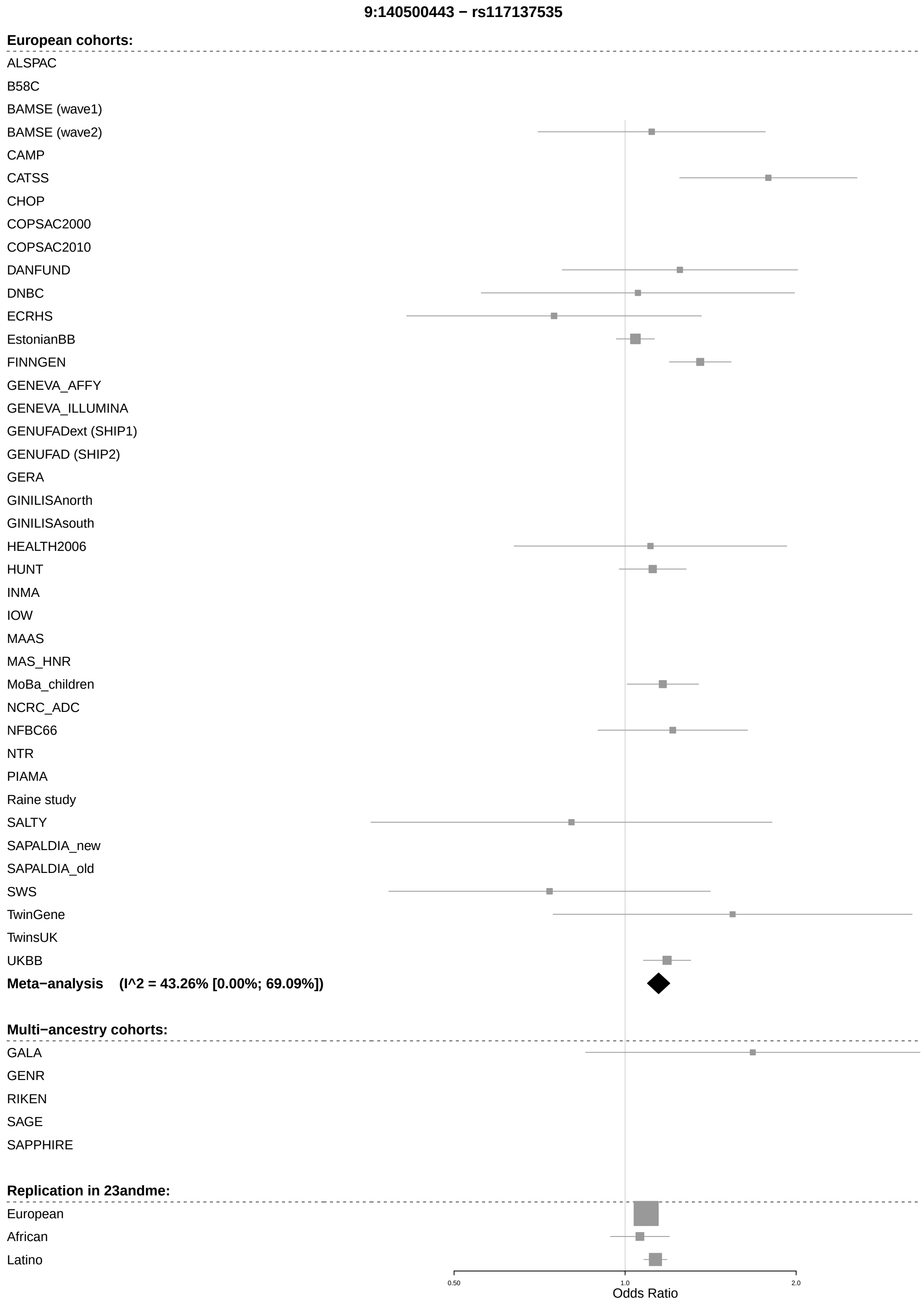
9:140500443 − rs117137535
European cohorts:
ALSPAC
B58C
BAMSE (wave1)
BAMSE (wave2)
CAMP
CATSS
CHOP
COPSAC2000
COPSAC2010
DANFUND
DNBC
ECRHS
EstonianBB
FINNGEN
GENEVA_AFFY
GENEVA_ILLUMINA
GENUFADext (SHIP1)
GENUFAD (SHIP2)
GERA
GINILISAnorth
GINILISAsouth
HEALTH2006
HUNT
INMA
IOW
MAAS
MAS_HNR
MoBa_children
NCRC_ADC
NFBC66
NTR
PIAMA
Raine study
SALTY
SAPALDIA_new
SAPALDIA_old
SWS
TwinGene
TwinsUK
UKBB
Meta−analysis (I^2 = 43.26% [0.00%; 69.09%])
Multi−ancestry cohorts:
GALA
GENR
RIKEN
SAGE
SAPPHIRE
Replication in 23andme:
European
African
Latino
0.50
1.0
2.0
Odds Ratio

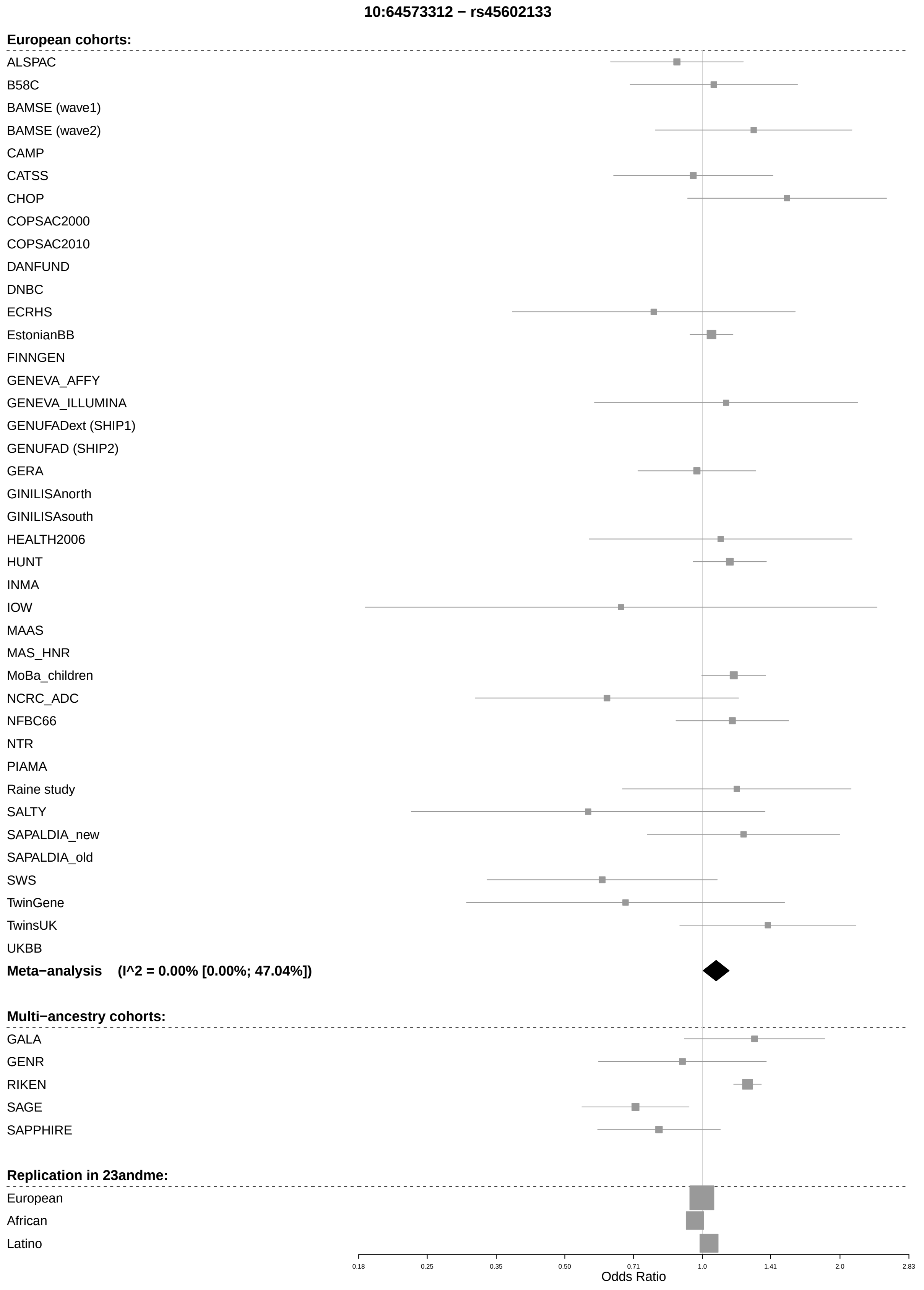
10:64573312 − rs45602133
European cohorts:
ALSPAC
B58C
BAMSE (wave1)
BAMSE (wave2)
CAMP
CATSS
CHOP
COPSAC2000
COPSAC2010
DANFUND
DNBC
ECRHS
EstonianBB
FINNGEN
GENEVA_AFFY
GENEVA_ILLUMINA
GENUFADext (SHIP1)
GENUFAD (SHIP2)
GERA
GINILISAnorth
GINILISAsouth
HEALTH2006
HUNT
INMA
IOW
MAAS
MAS_HNR
MoBa_children
NCRC_ADC
NFBC66
NTR
PIAMA
Raine study
SALTY
SAPALDIA_new
SAPALDIA_old
SWS
TwinGene
TwinsUK
UKBB
Meta−analysis (I^2 = 0.00% [0.00%; 47.04%])
Multi−ancestry cohorts:
GALA
GENR
RIKEN
SAGE
SAPPHIRE
Replication in 23andme:
European
African
Latino
0.18
0.25
0.35
0.50
0.71
1.0
1.41
2.0
2.83
Odds Ratio

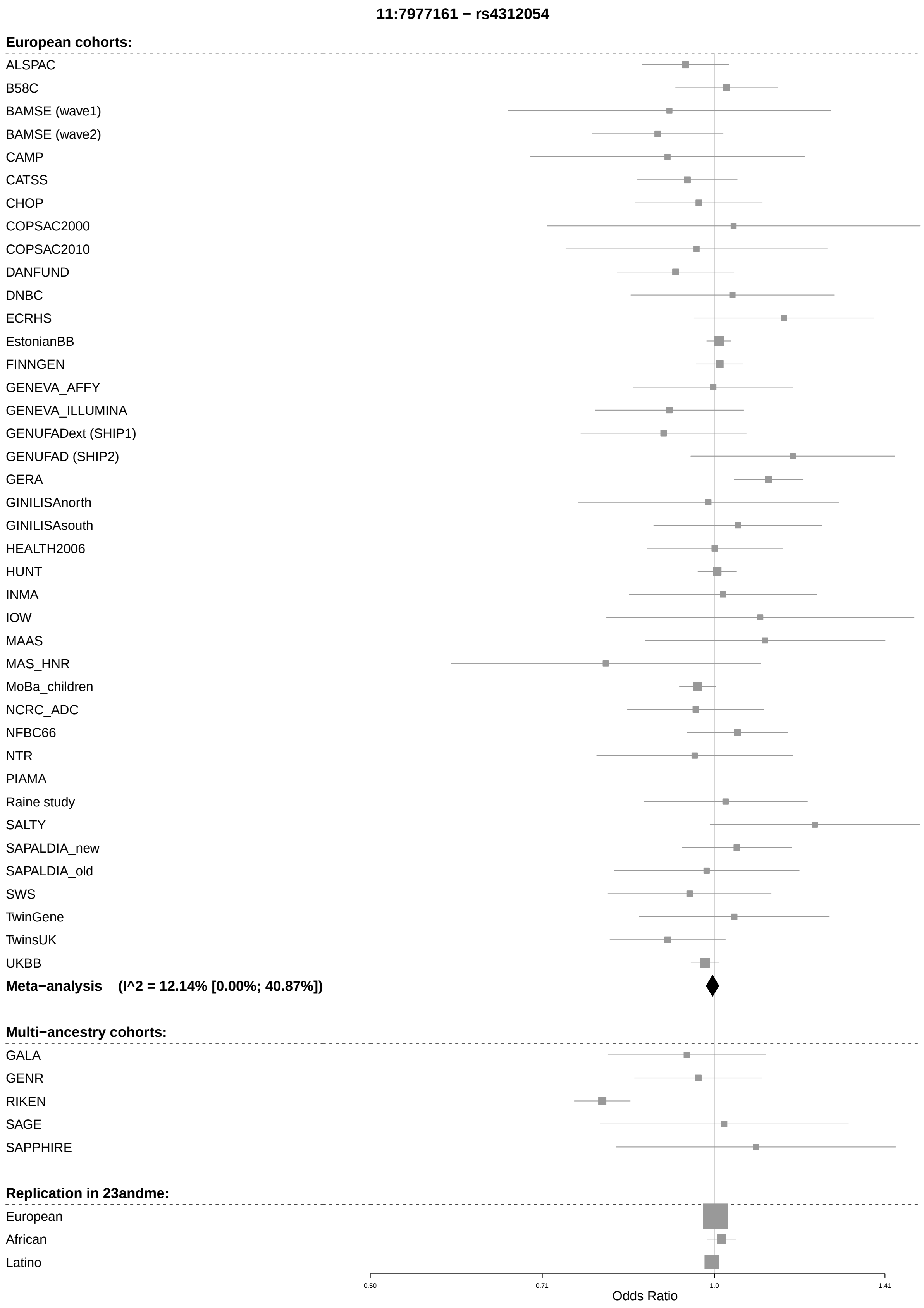
11:7977161 − rs4312054
European cohorts:
ALSPAC
B58C
BAMSE (wave1)
BAMSE (wave2)
CAMP
CATSS
CHOP
COPSAC2000
COPSAC2010
DANFUND
DNBC
ECRHS
EstonianBB
FINNGEN
GENEVA_AFFY
GENEVA_ILLUMINA
GENUFADext (SHIP1)
GENUFAD (SHIP2)
GERA
GINILISAnorth
GINILISAsouth
HEALTH2006
HUNT
INMA
IOW
MAAS
MAS_HNR
MoBa_children
NCRC_ADC
NFBC66
NTR
PIAMA
Raine study
SALTY
SAPALDIA_new
SAPALDIA_old
SWS
TwinGene
TwinsUK
UKBB
Meta−analysis (I^2 = 12.14% [0.00%; 40.87%])
Multi−ancestry cohorts:
GALA
GENR
RIKEN
SAGE
SAPPHIRE
Replication in 23andme:
European
African
Latino
0.50
0.71
1.0
1.41
Odds Ratio

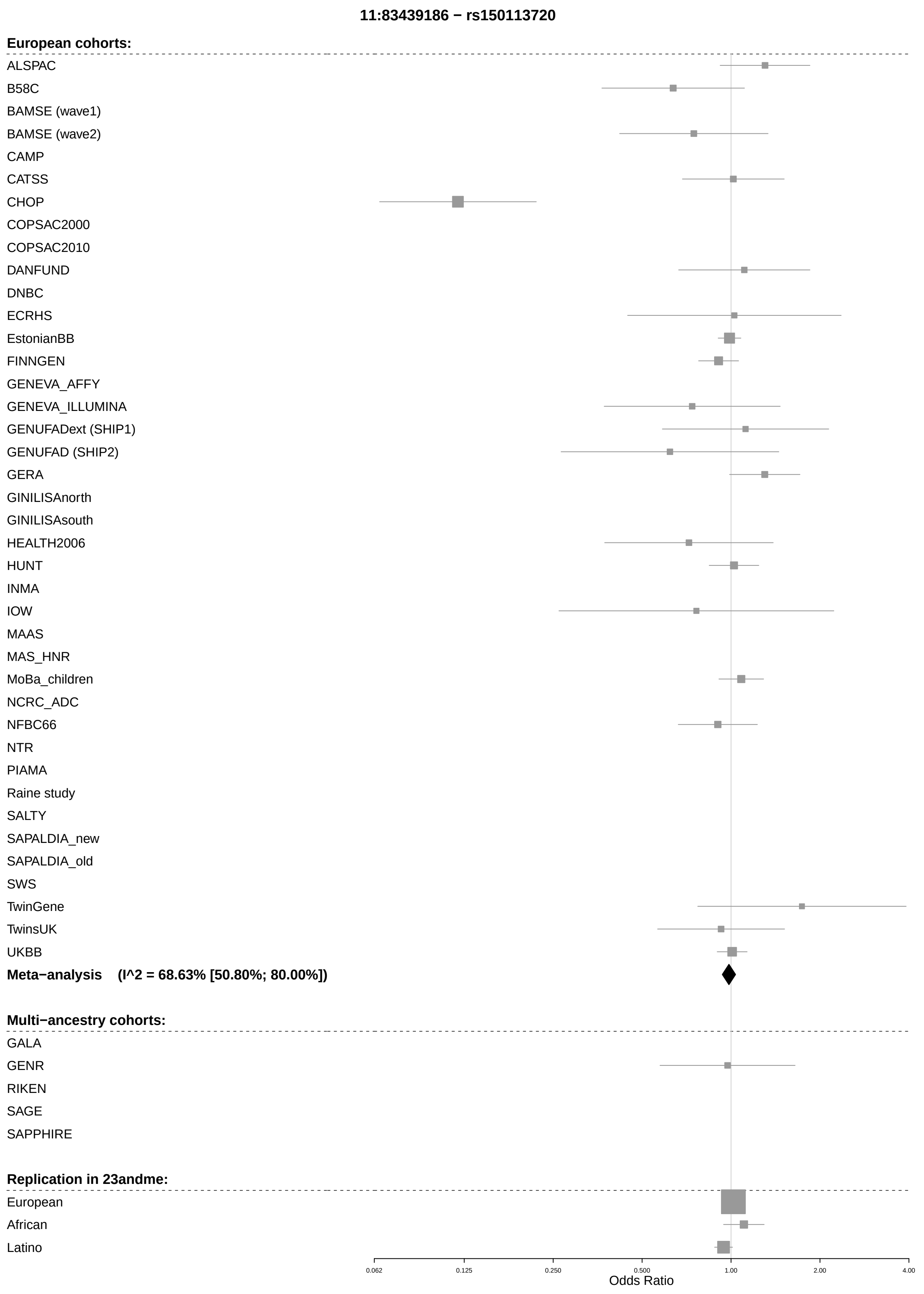
11:83439186 − rs150113720
European cohorts:
ALSPAC
B58C
BAMSE (wave1)
BAMSE (wave2)
CAMP
CATSS
CHOP
COPSAC2000
COPSAC2010
DANFUND
DNBC
ECRHS
EstonianBB
FINNGEN
GENEVA_AFFY
GENEVA_ILLUMINA
GENUFADext (SHIP1)
GENUFAD (SHIP2)
GERA
GINILISAnorth
GINILISAsouth
HEALTH2006
HUNT
INMA
IOW
MAAS
MAS_HNR
MoBa_children
NCRC_ADC
NFBC66
NTR
PIAMA
Raine study
SALTY
SAPALDIA_new
SAPALDIA_old
SWS
TwinGene
TwinsUK
UKBB
Meta−analysis (I^2 = 68.63% [50.80%; 80.00%])
Multi−ancestry cohorts:
GALA
GENR
RIKEN
SAGE
SAPPHIRE
Replication in 23andme:
European
African
Latino
0.062
0.125
0.250
0.500
1.00
2.00
4.00
Odds Ratio

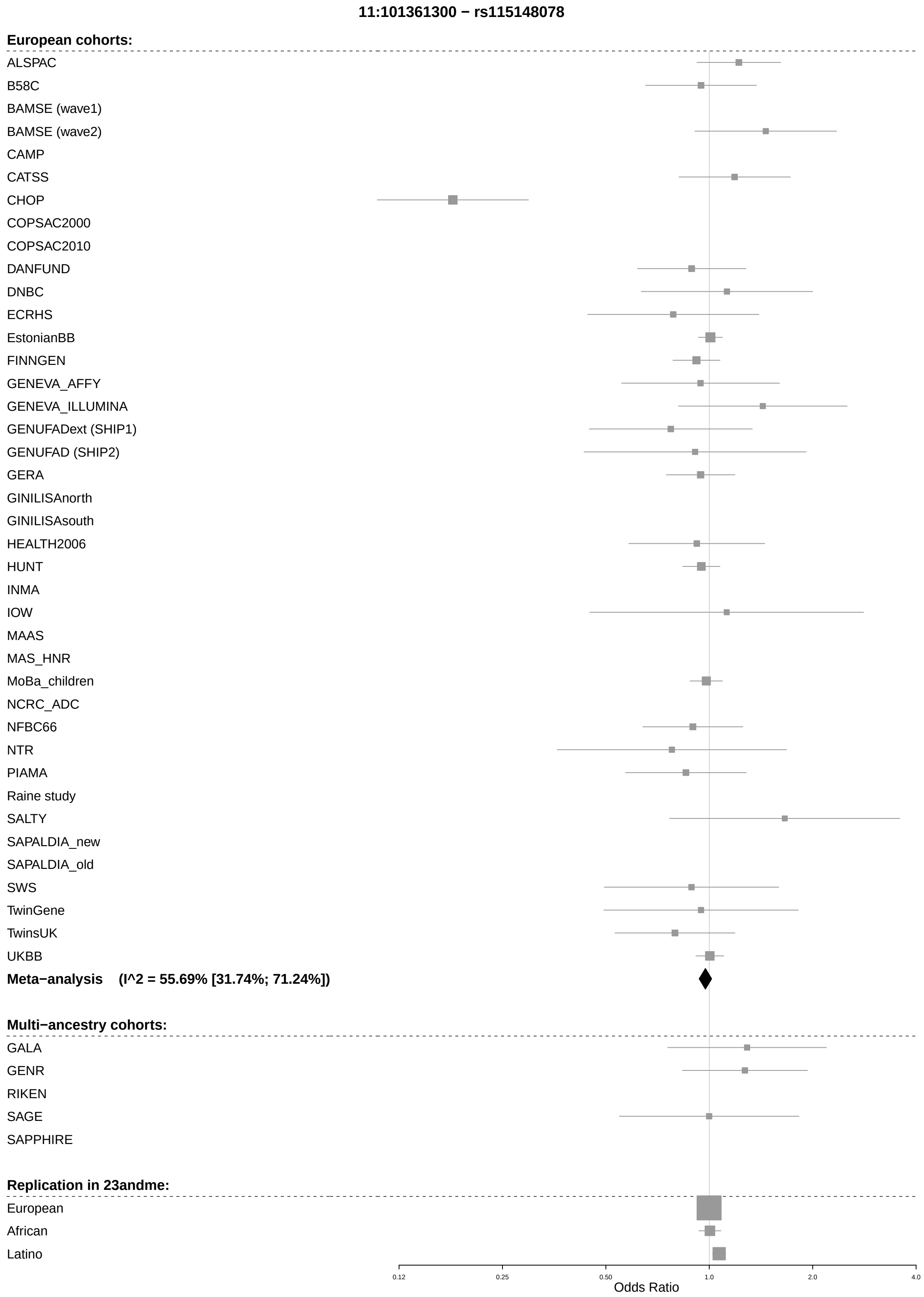
11:101361300 − rs115148078
European cohorts:
ALSPAC
B58C
BAMSE (wave1)
BAMSE (wave2)
CAMP
CATSS
CHOP
COPSAC2000
COPSAC2010
DANFUND
DNBC
ECRHS
EstonianBB
FINNGEN
GENEVA_AFFY
GENEVA_ILLUMINA
GENUFADext (SHIP1)
GENUFAD (SHIP2)
GERA
GINILISAnorth
GINILISAsouth
HEALTH2006
HUNT
INMA
IOW
MAAS
MAS_HNR
MoBa_children
NCRC_ADC
NFBC66
NTR
PIAMA
Raine study
SALTY
SAPALDIA_new
SAPALDIA_old
SWS
TwinGene
TwinsUK
UKBB
Meta−analysis (I^2 = 55.69% [31.74%; 71.24%])
Multi−ancestry cohorts:
GALA
GENR
RIKEN
SAGE
SAPPHIRE
Replication in 23andme:
European
African
Latino
0.12
0.25
0.50
1.0
2.0
4.0
Odds Ratio

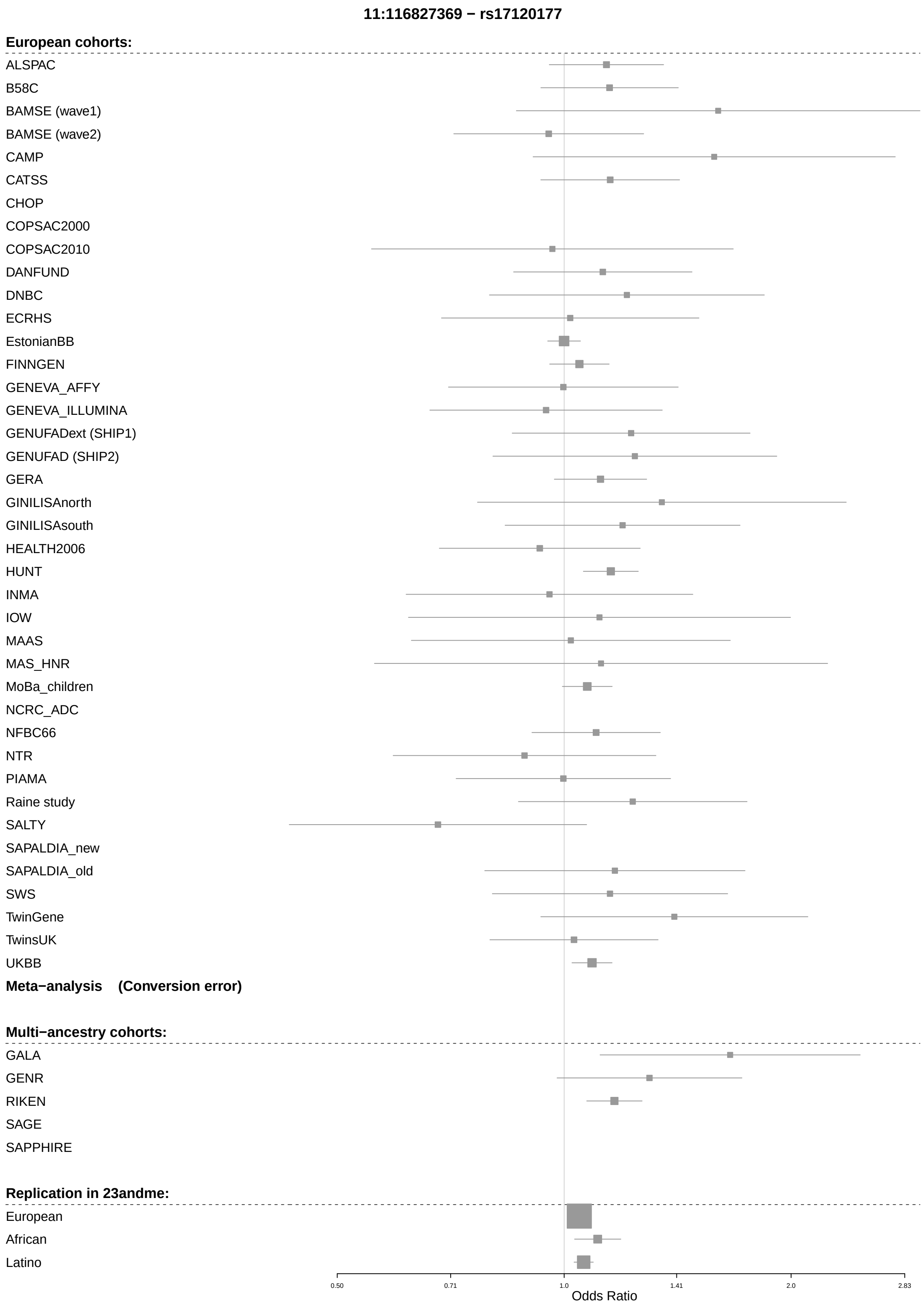
11:116827369 − rs17120177
European cohorts:
ALSPAC
B58C
BAMSE (wave1)
BAMSE (wave2)
CAMP
CATSS
CHOP
COPSAC2000
COPSAC2010
DANFUND
DNBC
ECRHS
EstonianBB
FINNGEN
GENEVA_AFFY
GENEVA_ILLUMINA
GENUFADext (SHIP1)
GENUFAD (SHIP2)
GERA
GINILISAnorth
GINILISAsouth
HEALTH2006
HUNT
INMA
IOW
MAAS
MAS_HNR
MoBa_children
NCRC_ADC
NFBC66
NTR
PIAMA
Raine study
SALTY
SAPALDIA_new
SAPALDIA_old
SWS
TwinGene
TwinsUK
UKBB
Meta−analysis (Conversion error)
Multi−ancestry cohorts:
GALA
GENR
RIKEN
SAGE
SAPPHIRE
Replication in 23andme:
European
African
Latino
0.50
0.71
1.0
1.41
2.0
2.83
Odds Ratio

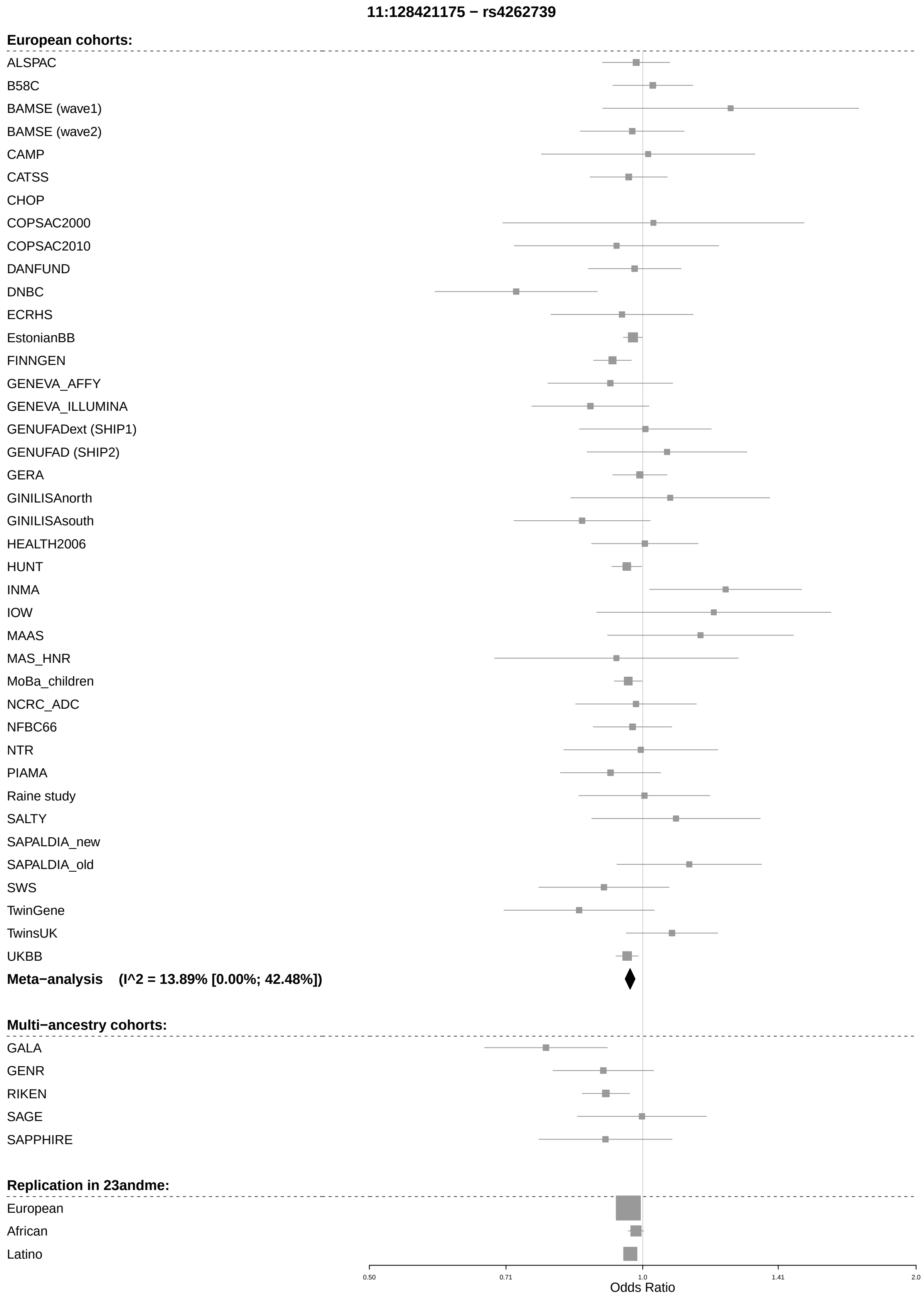
11:128421175 − rs4262739
European cohorts:
ALSPAC
B58C
BAMSE (wave1)
BAMSE (wave2)
CAMP
CATSS
CHOP
COPSAC2000
COPSAC2010
DANFUND
DNBC
ECRHS
EstonianBB
FINNGEN
GENEVA_AFFY
GENEVA_ILLUMINA
GENUFADext (SHIP1)
GENUFAD (SHIP2)
GERA
GINILISAnorth
GINILISAsouth
HEALTH2006
HUNT
INMA
IOW
MAAS
MAS_HNR
MoBa_children
NCRC_ADC
NFBC66
NTR
PIAMA
Raine study
SALTY
SAPALDIA_new
SAPALDIA_old
SWS
TwinGene
TwinsUK
UKBB
Meta−analysis (I^2 = 13.89% [0.00%; 42.48%])
Multi−ancestry cohorts:
GALA
GENR
RIKEN
SAGE
SAPPHIRE
Replication in 23andme:
European
African
Latino
0.50
0.71
1.0
1.41
2.0
Odds Ratio

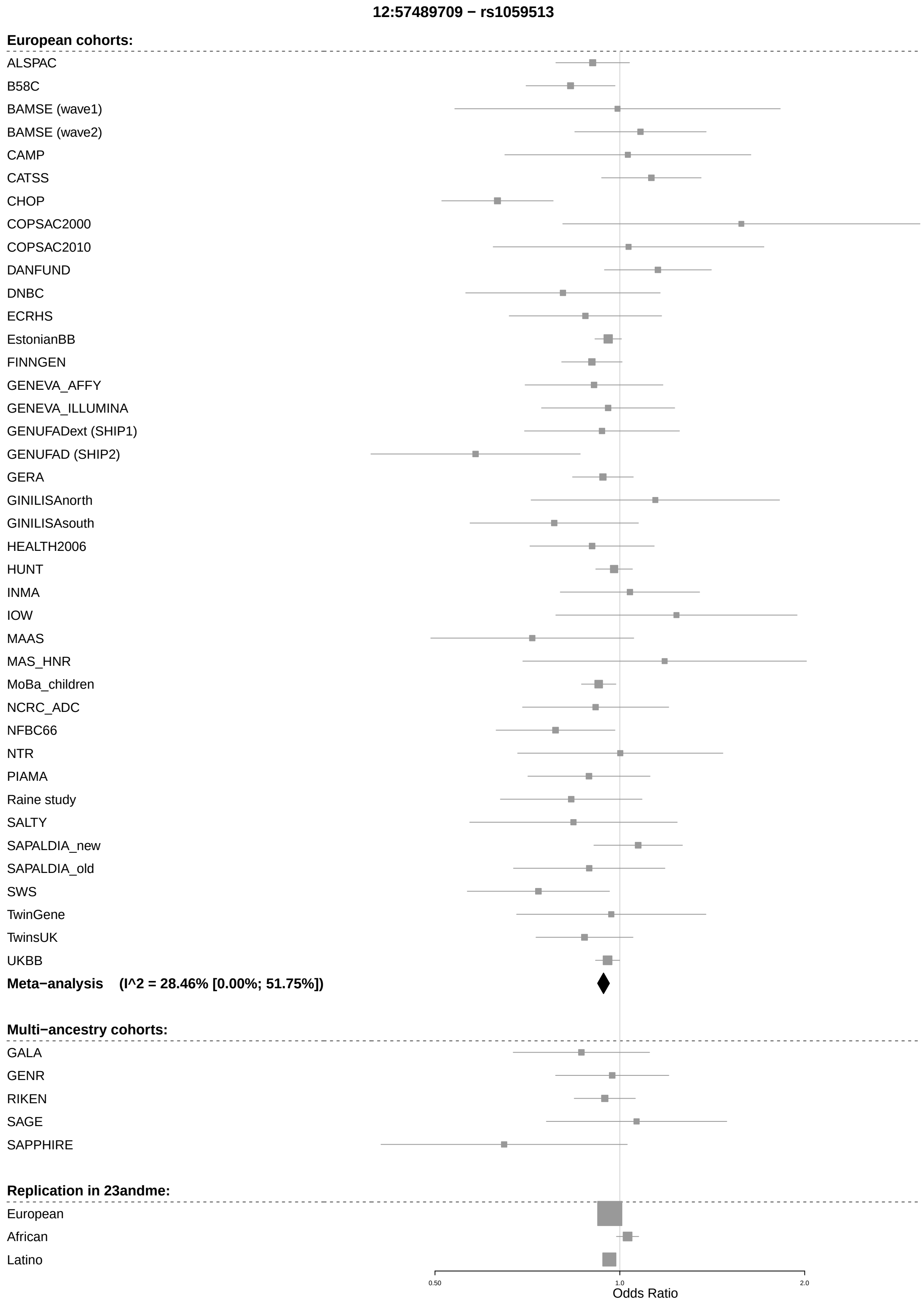
12:57489709 − rs1059513
European cohorts:
ALSPAC
B58C
BAMSE (wave1)
BAMSE (wave2)
CAMP
CATSS
CHOP
COPSAC2000
COPSAC2010
DANFUND
DNBC
ECRHS
EstonianBB
FINNGEN
GENEVA_AFFY
GENEVA_ILLUMINA
GENUFADext (SHIP1)
GENUFAD (SHIP2)
GERA
GINILISAnorth
GINILISAsouth
HEALTH2006
HUNT
INMA
IOW
MAAS
MAS_HNR
MoBa_children
NCRC_ADC
NFBC66
NTR
PIAMA
Raine study
SALTY
SAPALDIA_new
SAPALDIA_old
SWS
TwinGene
TwinsUK
UKBB
Meta−analysis (I^2 = 28.46% [0.00%; 51.75%])
Multi−ancestry cohorts:
GALA
GENR
RIKEN
SAGE
SAPPHIRE
Replication in 23andme:
European
African
Latino
0.50
1.0
2.0
Odds Ratio

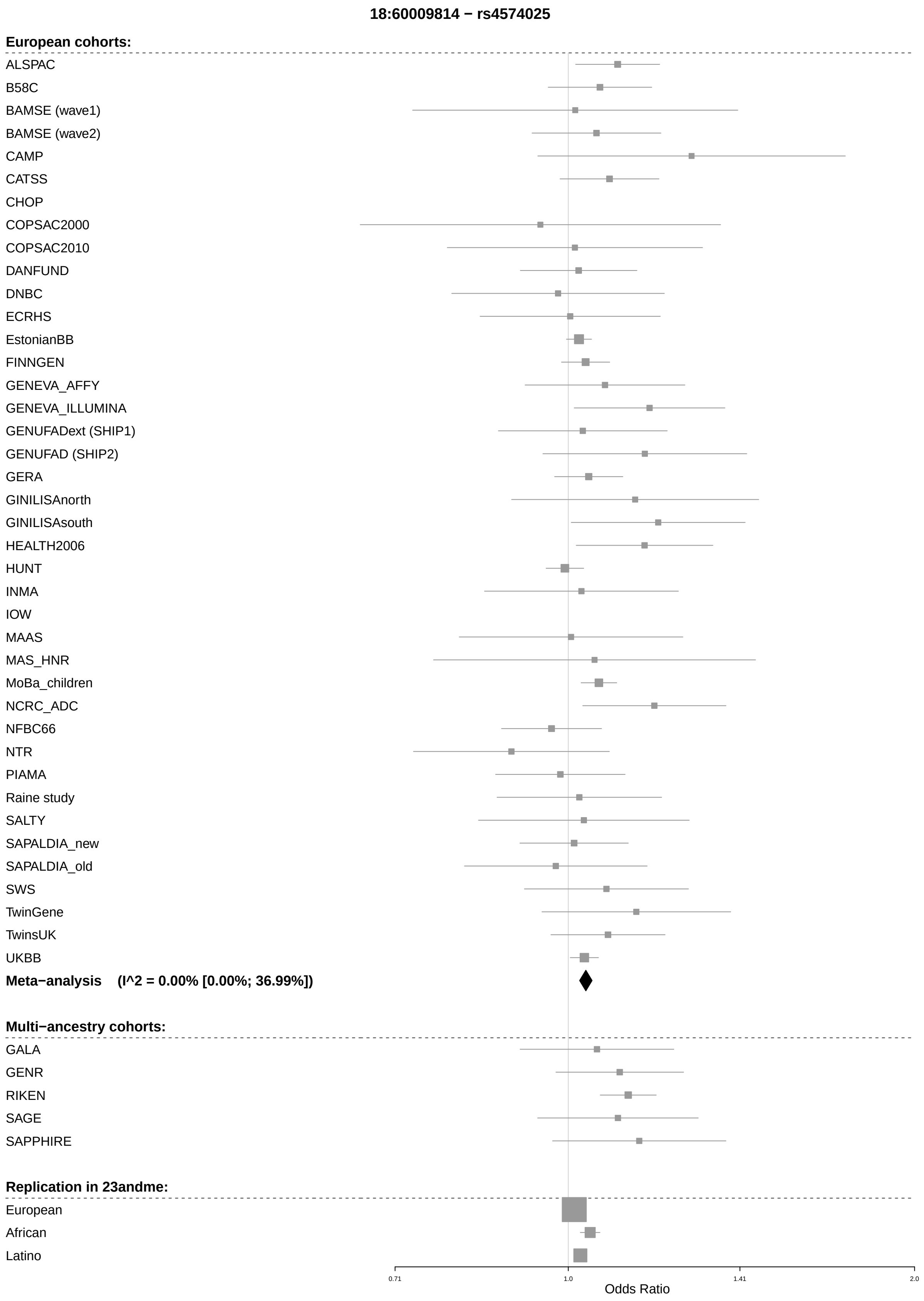
18:60009814 − rs4574025
European cohorts:
ALSPAC
B58C
BAMSE (wave1)
BAMSE (wave2)
CAMP
CATSS
CHOP
COPSAC2000
COPSAC2010
DANFUND
DNBC
ECRHS
EstonianBB
FINNGEN
GENEVA_AFFY
GENEVA_ILLUMINA
GENUFADext (SHIP1)
GENUFAD (SHIP2)
GERA
GINILISAnorth
GINILISAsouth
HEALTH2006
HUNT
INMA
IOW
MAAS
MAS_HNR
MoBa_children
NCRC_ADC
NFBC66
NTR
PIAMA
Raine study
SALTY
SAPALDIA_new
SAPALDIA_old
SWS
TwinGene
TwinsUK
UKBB
Meta−analysis (I^2 = 0.00% [0.00%; 36.99%])
Multi−ancestry cohorts:
GALA
GENR
RIKEN
SAGE
SAPPHIRE
Replication in 23andme:
European
African
Latino
0.71
1.0
1.41
2.0
Odds Ratio

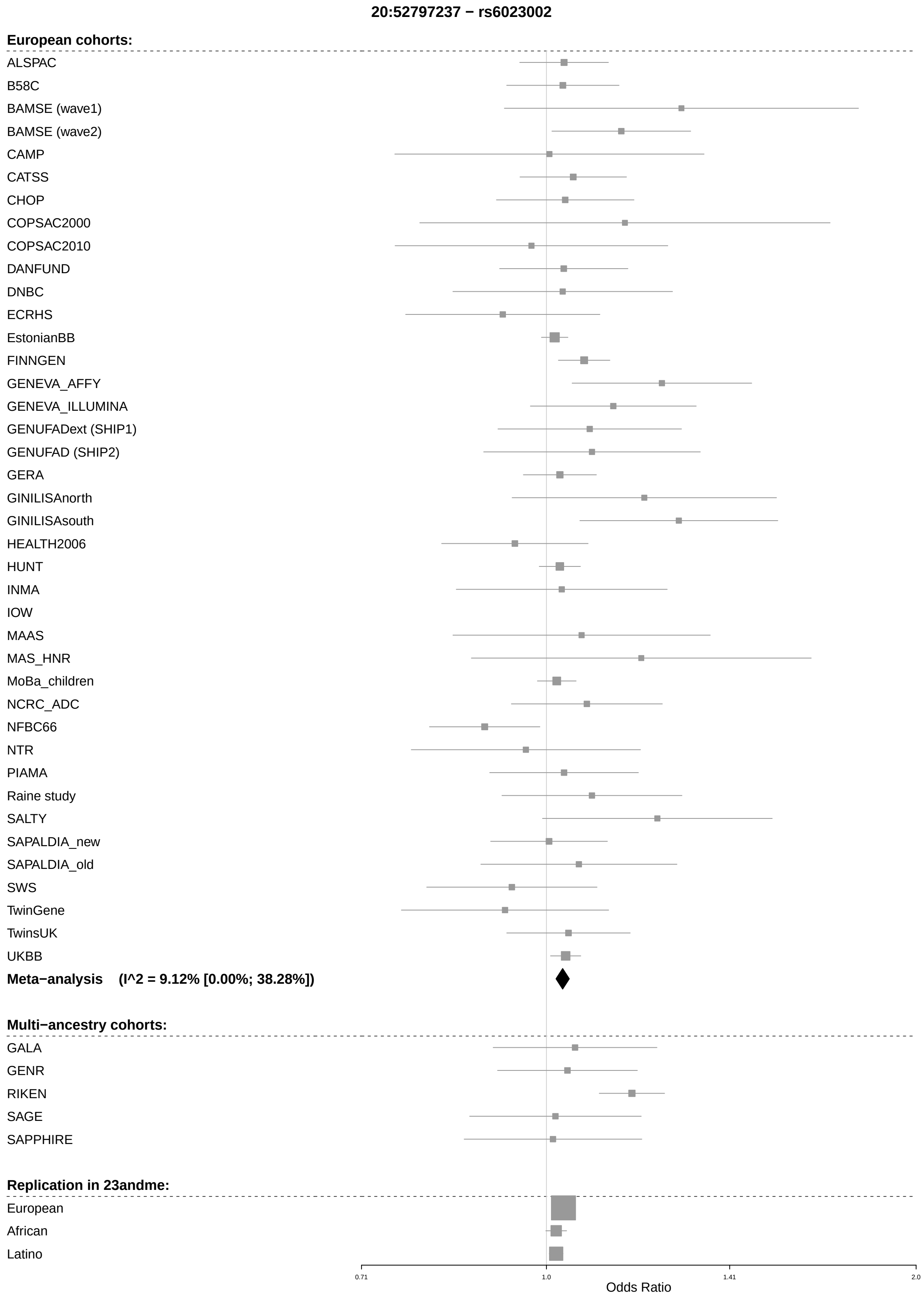
20:52797237 − rs6023002
European cohorts:
ALSPAC
B58C
BAMSE (wave1)
BAMSE (wave2)
CAMP
CATSS
CHOP
COPSAC2000
COPSAC2010
DANFUND
DNBC
ECRHS
EstonianBB
FINNGEN
GENEVA_AFFY
GENEVA_ILLUMINA
GENUFADext (SHIP1)
GENUFAD (SHIP2)
GERA
GINILISAnorth
GINILISAsouth
HEALTH2006
HUNT
INMA
IOW
MAAS
MAS_HNR
MoBa_children
NCRC_ADC
NFBC66
NTR
PIAMA
Raine study
SALTY
SAPALDIA_new
SAPALDIA_old
SWS
TwinGene
TwinsUK
UKBB
Meta−analysis (I^2 = 9.12% [0.00%; 38.28%])
Multi−ancestry cohorts:
GALA
GENR
RIKEN
SAGE
SAPPHIRE
Replication in 23andme:
European
African
Latino
0.71
1.0
1.41
2.0
Odds Ratio

(*) Marked SNPs have failed quality control in 23andme latinos, even though they are still found.
