## Supplementary figures and images for "European and multi-ancestry genome-wide association meta-analysis of atopic dermatitis highlights importance of systemic immune regulation"

### Supplementary Figure 3

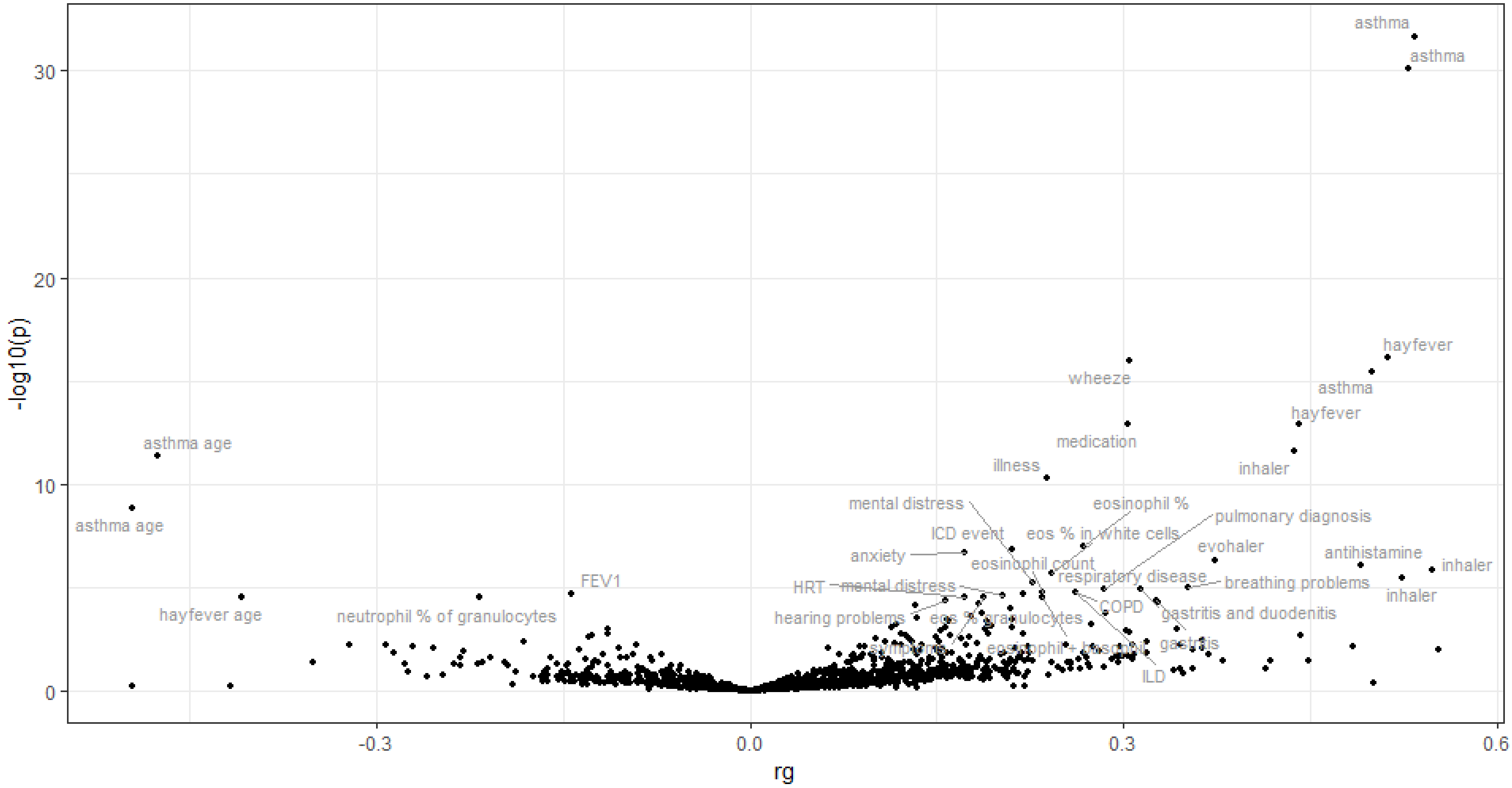

### Supplementary Figure 4

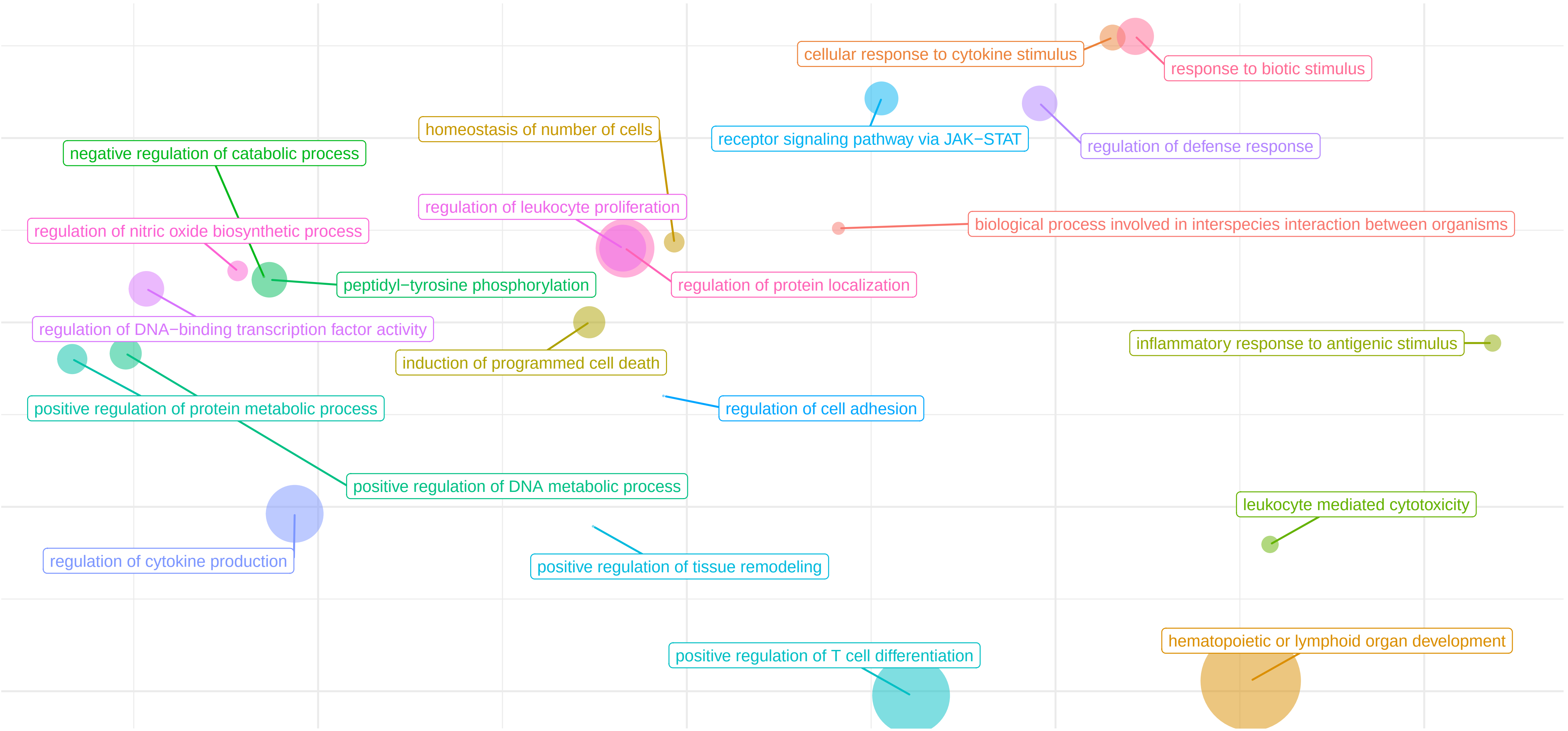

### Supplementary Figure 5

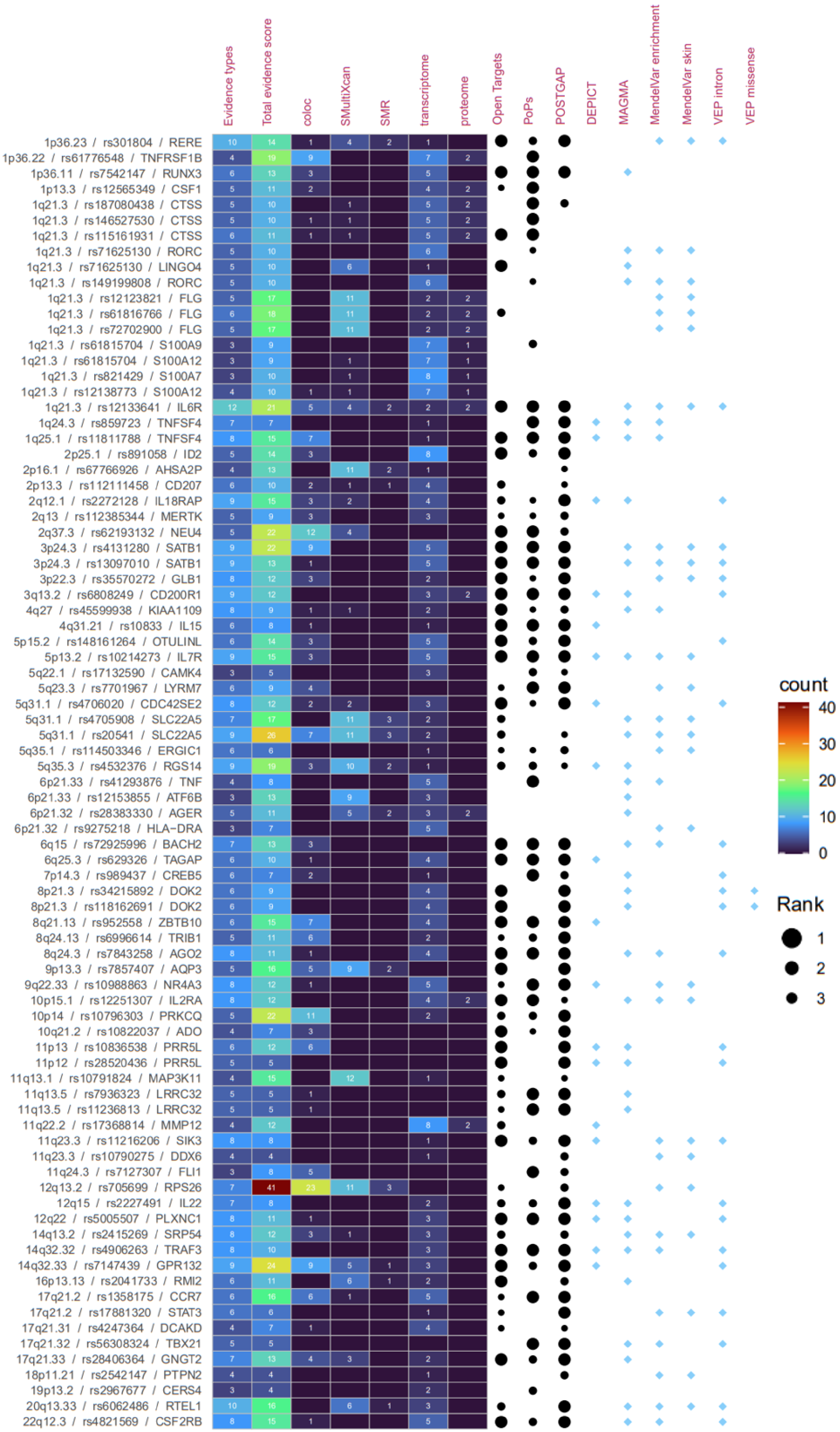
