## Supplementary Methods for "European and multi-ancestry genome-wide association meta-analysis of atopic dermatitis highlights importance of systemic immune regulation"

**Budu-Aggrey, A et al**

**Supplementary Methods**

**Individual cohort details** ... 3
ALSPAC ... 3
1958 Birth Cohort (B58C) ... 5
BAMSE ... 6
Biobank Japan (BBJ) ... 7
CAMP ... 8
Child and Adolescent Twin Study in Sweden (CATSS) ... 10
Children's Hospital of Philadelphia (CHOP) ... 10
Copenhagen Prospective Studies on Asthma in Childhood 2000 (COPSAC2000) ... 11
Copenhagen Prospective Studies on Asthma in Childhood 2010 (COPSAC2010) ... 12
DanFunD ... 12
Danish National Birth Cohort (DNBC) ... 13
ECHRS ... 14
Estonian Biobank ... 16
FinnGen (Release 3) ... 16
The Genes-environments & Admixture in Latino Americans (GALA II) study ... 17
Generation R ... 19
GENEVA ... 21
GENUFADext (SHIP1) ... 22
GENUFAD (SHIP2) ... 23
GERA ... 24
GINIplus/LISA north ... 26
GINI / LISAsouth ... 28
Health Cohorts ... 30
HUNT ... 31
INfancia y Medio Ambiente Project (INMA) ... 32
The Isle of Wight birth cohort ... 33
Manchester Asthma and Allergy Study (MAAS) ... 34

Multicentre Allergy Study / Heinz Nixdorf RECALL (MAS / HNR) ... 35
MoBa ... 35
National Children's Research Centre - Atopic Dermatitis Collection (NCRC - ADC) ... 37
Northern Finland Birth Cohort 1966 (NFBC66) ... 38
Netherlands Twin Register (NTR) ... 39
PIAMA ... 41
The Raine Study ... 42
The Study of African Americans, Asthma, Genes, and Environments (SAGE) ... 43
SALTY ... 44
SAPALDIA ... 45
SAPPHIRE ... 47
Southampton Women's Survey (SWS) ... 48
TwinGene ... 49
TwinsUK ... 50
UK Biobank ... 50
**Replication Studies** ... 53
23andMe, Inc. ... 53
**Cohort ethical approval** ... 55

### Individual cohort details

#### ALSPAC

Recruitment

Pregnant women resident in Avon, UK with expected dates of delivery 1st April 1991 to 31st December 1992 were invited to take part in the study. The initial number of pregnancies enrolled is 14,541. Of these initial pregnancies, there was a total of 14,676 foetuses, resulting in 14,062 live births and 13,988 children who were alive at 1 year of age. When the oldest children were approximately 7 years of age, an attempt was made to bolster the initial sample with eligible cases who had failed to join the study originally. Therefore, the total sample size for analyses using any data collected after the age of seven is therefore 15,454 pregnancies, resulting in 15,589 foetuses. Further details of enrolment have been described previously[1,2]. Please note that the study website contains details of all the data that is available through a fully searchable data dictionary and variable search tool: http://www.bristol.ac.uk/alspac/researchers/our-data/. Ethical approval for the study was obtained from the ALSPAC Ethics and Law Committee and the Local Research Ethics Committees. Informed consent for the use of data collected via questionnaires and clinics was obtained from participants following the recommendations of the ALSPAC Ethics and Law Committee at the time. Consent for biological samples has been collected in accordance with the Human Tissue Act (2004).

Case/Control definition

The children have been followed up with regular questionnaires and clinic visits. Data collected from the questionnaires was used to classify children as AD cases or controls. When the children were approximately 81, 91, 103 months, 10, 13, 14 years, parents were asked the following questions [possible answers]:

1. Has your child in the past 12 months had eczema? [Yes and saw a Dr; Yes, but did not see a Dr; No]
2. Has a doctor ever actually said that your child has eczema? [yes; no] (10 & 14 years only)

We defined cases as the children whose parents answered “Yes and saw a Dr” to Q1 or “yes” to Q2. We defined controls as the children who were not a case and whose parents answered “No” to Q2 at 14 years.

Genotyping and imputation

ALSPAC children were genotyped using the Illumina HumanHap550 quad chip genotyping platforms by 23andme subcontracting the Wellcome Trust Sanger Institute, Cambridge, UK and the Laboratory Corporation of America, Burlington, NC, US. The resulting raw genome-wide data were subjected to standard quality control methods. Individuals were excluded on the basis of gender mismatches; minimal or excessive heterozygosity; disproportionate levels of individual missingness (>3%) and insufficient sample replication (IBD < 0.8). Population stratification was assessed by multidimensional scaling analysis and compared with Hapmap II (release 22) European descent (CEU), Han Chinese, Japanese and Yoruba reference populations; all individuals with non-European ancestry were removed. SNPs with a minor allele frequency of < 1%, a call rate of < 95% or evidence for violations of Hardy-Weinberg equilibrium ($P < 5E-7$) were removed. Cryptic relatedness was measured as proportion of identity by descent (IBD > 0.1). Related subjects that passed all other quality control thresholds were retained during subsequent phasing and imputation. 9,115 subjects and 500,527 SNPs passed these quality control filters.

ALSPAC mothers were genotyped using the Illumina human660W-quad array at Centre National de Génotypage (CNG) and genotypes were called with Illumina GenomeStudio. PLINK (v1.07) was used to carry out quality control measures on an initial set of 10,015 subjects and 557,124 directly genotyped SNPs. SNPs were removed if they displayed more than 5% missingness or a Hardy-Weinberg equilibrium P value of less than 1.0E-06. Additionally, SNPs with a minor allele frequency of less than 1% were removed. Samples were excluded if they displayed more than 5% missingness, had indeterminate X chromosome heterozygosity or extreme autosomal heterozygosity. Samples showing evidence of population stratification were identified by multidimensional scaling of genome-wide identity by state pairwise distances using the four HapMap populations as a reference, and then excluded. Cryptic relatedness was assessed using a IBD estimate of more than 0.125 which is expected to correspond to roughly 12.5% alleles shared IBD or a relatedness at the first cousin level. Related subjects that passed all other quality control thresholds were retained during subsequent phasing and imputation. 9,048 subjects and 526,688 SNPs passed these quality control filters.

We combined 477,482 SNP genotypes in common between the sample of mothers and sample of children. We removed SNPs with genotype missingness above 1% due to poor quality (11,396 SNPs removed) and removed a further 321 subjects due to potential ID mismatches. This resulted in a dataset of 17,842 subjects containing 6,305 duos and 465,740 SNPs (112 were removed during liftover and 234 were out of HWE after combination). We estimated haplotypes using ShapeIT (v2.r644) which utilises relatedness during phasing. The phased haplotypes were then imputed to the Haplotype Reference Consortium (HRCr1.1, 2016) panel of approximately 31,000 phased whole genomes. The HRC panel was phased using ShapeIt v2, and the imputation was performed using the Michigan imputation server.

This gave 8,237 eligible children and 8,196 eligible mothers with available genotype data after exclusion of related subjects using cryptic relatedness measures described previously.

### Statistical analysis

Genome-wide association analysis was performed using SNPTEST (version 2.5.2) for 1,620 cases and 3,538 controls with genetic and phenotypic data while controlling for sex and 10 principal components. Summary data was available for 7,662,269 variants that were successfully analysed.

### Acknowledgements and Funding

We are extremely grateful to all the families who took part in this study, the midwives for their help in recruiting them, and the whole ALSPAC team, which includes interviewers, computer and laboratory technicians, clerical workers, research scientists, volunteers, managers, receptionists and nurses.

The UK Medical Research Council and Wellcome (Grant ref: 217065/Z/19/Z) and the University of Bristol provide core support for ALSPAC. GWAS data was generated by Sample Logistics and Genotyping Facilities at Wellcome Sanger Institute and LabCorp (Laboratory Corporation of America) using support from 23andMe. A comprehensive list of grants funding is available on the ALSPAC website (http://www.bristol.ac.uk/alspac/external/documents/grant-acknowledgements.pdf).

**1958 Birth Cohort (B58C)**

Recruitment

The British 1958 birth cohort is an ongoing follow-up of all persons born in England, Scotland and Wales during one week in 1958. At ages 7, 11 and 16 years, a history of eczematous rashes was obtained by interview with a parent, and the presence of visible eczema on skin examination was recorded by a school medical officer[3]. At the age of 44-45 years, the cohort were followed up with a biomedical examination and blood sampling[4], from which a DNA collection was established as a nationally representative reference panel.

Case/Control definition

For the purpose of this meta-analysis, cases were defined by a positive parental or self-report of eczema up to age 16, or eczema recorded on skin examination at ages 7, 11 and/or 16. The cases included 305 ascertained by medical examination, of which 245 were also reported by parents; 467 additional cases reported by parents, but without eczema when examined at school; plus 146 cohort members who self-reported a history of eczema starting before age 17 years, when interviewed as adults. Controls (N=4560) were defined as cohort members with no parentally reported or self-reported history of eczema by age 16, and no record of eczema on skin examination at ages 7, 11 or 16 years.

Genotyping and imputation

Three non-overlapping subsets of the DNA collection from cohort members of white European ethnicity contributed to the genome-wide dataset:

*i.* 3027 specimens selected as nationally representative controls for use by the Wellcome Trust Case-Control Consortium (WTCCC)[5] – 1430 from WTCCC1 and 1597 from WTCCC2;

*ii.* 2592 specimens selected as nationally representative controls for the Type 1 Diabetes Genetics Consortium (T1DGC)[6]; and

*iii.* 872 specimens selected by the GABRIEL consortium[7], including equal numbers of asthmatics and non-asthmatics.

Genotyping was performed using the Illumina 550K array (WTCCC1 and T1DGC), the Illumina 610K array (GABRIEL) or the Illumina 1M array (WTCCC2). A set of SNPs common to these arrays were used for imputation against the March 2012 (phase 1, version 3) release of the 1000-genomes reference haplotypes for all ancestries.

Pre-imputation phasing was performed using MACH v1.0.18 and imputation was performed using Minimac (version dated 16 November 2012). SNPs were excluded from inputs to the imputation step if one or more of the following applied:

- SNP-wise call-rate <95% (>=5% missing genotypes)
- Minor allele frequency <1%
- Inconsistency of allele frequencies across the deposits (p<0.0001 for any of the pairwise comparisons)
- Departure from Hardy-Weinberg equilibrium (p<0.0001). (HWE on chrX was tested in females only)

- Inconsistency of allele frequencies in males and females ($p<0.0001$) for chrX SNPs.
- Inconsistency of allele frequencies with 1000-genomes European ancestry haplotypes.

Additionally, for this eczema analysis, four filaggrin mutations (R501X, 2282del4, R2447X and S3247X) were genotyped directly by LGC Genomics using KASP genotyping technology, as described for the ALSPAC samples.

Statistical Analysis

Within-cohort logistic regression analyses for eczema were performed using ProbAbel v0.1-9e using the imputed allele dosage for each SNP as the explanatory variable. Imputed allele dosages were not replaced with original genotypes for the directly genotyped SNPs. Thus, there were no missing data for any of the imputed variants. Analyses of the four filaggrin mutations were restricted to those with valid genotypes: 809 cases (155 of whom had one or more null mutations) and 3918 controls (383 of whom had a null mutation).

Acknowledgements and Funding

We acknowledge use of phenotype and genotype data from the British 1958 Birth Cohort DNA collection, funded by the Medical Research Council grant G0000934 and the Wellcome Trust grant 068545/Z/02. Genotyping for the B58C-WTCCC subset was funded by the Wellcome Trust grant 076113/B/04/Z. The B58C-T1DGC genotyping utilized resources provided by the Type 1 Diabetes Genetics Consortium, a collaborative clinical study sponsored by the National Institute of Diabetes and Digestive and Kidney Diseases (NIDDK), National Institute of Allergy and Infectious Diseases (NIAID), National Human Genome Research Institute (NHGRI), National Institute of Child Health and Human Development (NICHD), and Juvenile Diabetes Research Foundation International (JDRF) and supported by U01 DK062418. B58C-T1DGC GWAS data were deposited by the Diabetes and Inflammation Laboratory, Cambridge Institute for Medical Research (CIMR), University of Cambridge, which is funded by Juvenile Diabetes Research Foundation International, the Wellcome Trust and the National Institute for Health Research Cambridge Biomedical Research Centre; the CIMR is in receipt of a Wellcome Trust Strategic Award (079895). The B58C-GABRIEL genotyping was supported by a contract from the European Commission Framework Programme 6 (018996) and grants from the French Ministry of Research.

**BAMSE**

Study design

The Children Allergy Milieu Stockholm an Epidemiological Study (BAMSE) is a prospective population-based birth cohort in which newborn infants between 1994 and 1996 from the north and central area of Stockholm were recruited. The baseline questionnaires were obtained when the children were about 2 months old and follow-up have been conducted at 1, 2, 4, 8, 16, and 24 years (Ref: PMID: 32489587). BAMSE was approved by the Regional ethical committee in Stockholm (Stockholm, Sweden) (ethics approval numbers: 02-420 and 2010/1474-31/3).

Genotyping

Within the BAMSE project, genotyping was done in two waves. Wave1 was done on the Illumina Human 610-quad array (Illumina, Inc., San Diego, CA). A total of 505 samples were genotyped (a subset of the study consisting of asthma cases and controls, out of which 485 samples were of good genotype quality. Wave2 was done on the Illumina Infinium Global Screening Array-24 v1.0 (GSA) BeadChip, where a total of 2387 samples were genotyped, out of which 2367 samples were of good genotype quality. Samples were further excluded if the 10 genetic principal components indicated a non-EU ethnic outlier after projection of the study samples on the 1000 Genomes reference sample. For imputation, samples were also excluded if their genotyping success rate was lower than 95% and SNPs were excluded for - Call rate < 95%; HWE p< 1e-6; MAF < 0.01, determined using Plink 2.0. These QCed SNPs were imputed using the Sanger imputation server (EAGLE2+PBWT) based upon the HRC (Version r1.1 2016).

After further quality control a total of 464 participants within wave1 and 2194 participants within wave2 remained. All data was imputed using the HRC version 1.1 on the Sanger imputation server (EAGLE2+PBWT) using HRC reference panel (Version r1.1 2016).

Case/control definitions

AD was defined as doctor's diagnosis of AD ever up to age 16 years. Controls: no diagnosis of AD up to age 16 years.

Acknowledgements and Funding

The BAMSE study was supported by grants from the Swedish Research Council (grant agreements 2016-03086; 2018-02524; 2020-02170), the Swedish Heart-Lung Foundation, the Swedish Asthma and Allergy research foundation and Region Stockholm (ALF projects, and for cohort and database maintenance).

**Biobank Japan (BBJ)**

Recruitment

The Biobank Japan (BBJ) project consisted of approximately 200,000 individuals diagnosed with one or more of 47 common diseases. Participants were enrolled at hospitals all over Japan from June 2003 to March 2008. Informed consent was obtained from all participants before enrolment. Clinical information was collected annually via interviews and medical record reviews until 2013. DNA samples at baseline were collected from all participants. A full description of the study design, targeting diseases, participants and data collecting methods have been described in detail previously[8,9]. The BBJ Project was approved by the research ethics committees at the Institute of Medical Science, the University of Tokyo, the RIKEN Center for Integrative Medical Sciences, and the cooperating hospitals.

Case/Control definition

All atopic dermatitis (AD) cases were diagnosed by physicians according to the criteria of Hanifin and Rajka[10]. Samples without AD-related diseases, mainly other allergic diseases (such as asthma), and autoimmune diseases (such as rheumatoid arthritis and Graves disease) as controls. The detailed list of related diseases was described previously[11].

Genotyping and imputation

BBJ samples were genotyped using Illumina Human OmniExpress Exome BeadChip or a combination of Illumina HumanOmniExpress and HumanExome BeadChips (Illumina Inc, San Diego, Calif). For quality control of genotypes, variants with call rate <99%, P value for Hardy-Weinberg equilibrium <1.0E$^{-6}$, difference of allele frequency between imputation panel >6.0%, and minor allele frequency (MAF) <0.005 were excluded. For quality control of subjects, we excluded subjects showing a call rate less than 0.98. The proportion of identical-by-descent was estimated by PI-HAT index using the PLINK software (version 1.9), and we excluded samples showing a high degree of relatedness (PI-HAT >0.25) or outliers from the East Asian (EAS) cluster by principal component analysis performed by smartpca (version 13050). 118,287 subjects (2,639 cases and 115,648 controls) passed these quality control filters. Genotype data were phased by Eagle (version 2.3.5). Phased genotype data were imputed by Minimac4 (version 1.0.0) using the reference panel conducted 1000 Genome Project (1KG) phase 3 (version5) and Japanese whole-genome sequence data (Jack Flanagan Thomas et al, manuscript in preparation, May 2021). After imputation, variants with low imputation accuracy ($R^2$ < 0.3) or MAF <0.005 among control samples were removed.

Statistical analysis

For the genome-wide association analysis, a logistic regression model with 10 principal components and sex as covariates was performed using PLINK (version 2.0).

Acknowledgements and Funding

We are extremely grateful to the individuals who participated and the staff in the BBJ project. We also thank the technical support from all members in Laboratory for Statistical and Translational Genetics, RIKEN Center for Integrative Medical Sciences. The BioBank Japan Project was funded by the Ministry of Education, Culture, Sports, Sciences and Technology of the Japanese Government and the Japan Agency for Medical Research and Development (grants 17km0305002 and 18km0605001).

CT was supported by Japan Agency for Medical Research and Development (AMED) grants JP21kk0305013, JP21tm0424220, JP21ck0106642, JP22ek0410079 and Japan Society for the Promotion of Science (JSPS) KAKENHI grant JP20H00462. NT was supported by the RIKEN Junior Research Associate Program.

**CAMP**

Recruitment

The Childhood Asthma Management Program (CAMP) population is composed of non-Hispanic white subjects from a multicenter clinical trial that followed 1,041 children with asthma for four years and 84% of the original participants for 12 years[12]. Stringent inclusion criteria ensured that participants had mild to moderate asthma, which was assessed as having asthma symptoms at least twice per week, using asthma medication daily, or using an inhaled bronchodilator twice per week for six or more months of the year prior to recruitment. CAMP subjects had increased airway responsiveness, as established by a bronchoprovocation test of up to 12.5mg/dl of methacholine resulting in 20% or greater forced expiratory volume in one second (FEV1) reduction.

Case/Control definition

581 subjects with genome-wide SNP genotype data were used for this analysis. Subjects were considered to have atopic dermatitis if they self-reported having seen a physician for eczema or atopic dermatitis at the baseline visit. A total of 143 subjects had atopic dermatitis and 438 did not have atopic dermatitis.

Genotyping and imputation

Genome-wide SNP genotyping of 581 CAMP subjects was performed on Illumina's HumanHap550v3 and 610 Genotyping BeadChips (Illumina, Inc., San Diego, CA). We included SNPs that satisfy minor allele frequency 2"5%, a minimum of 95% genotyping rate, and the Hardy Weinberg equilibrium (HWE) test at the 0.001 threshold. Details of the quality control (QC) criteria used to screen the genome-wide SNP data have been provided previously[13].

SNPs passing quality control criteria were used for imputation of variants from the 1000 Genomes project. This imputation was done using MaCH[14], version 1.0.16, and was imputed against "1000Genomes 0908" ( http://www.sph.umich.edu/csg/abecasis/MACH/download/1000G-Sanger-0908.html). Results were lifted over to hg19 format for the analysis.

Statistical Analysis

We tested for the association of SNPs with atopic dermatitis using an additive logistic regression model, where SNP was coded as allele dose. Models were implemented in PLINK[15] and controlled for age.

Acknowledgements and Funding

We thank all CAMP subjects for their ongoing participation in this study. We acknowledge the CAMP investigators and research team, supported by NHLBI, for collection of CAMP Genetic Ancillary Study data. All work on data collected from the CAMP Genetic Ancillary Study was conducted at the Channing Laboratory of the Brigham and Women's Hospital under appropriate CAMP policies and human subject's protections. The CAMP Genetics Ancillary Study is supported by U01 HL075419, U01 HL65899, P01 HL083069 R01 HL 086601, from the National Heart, Lung and Blood Institute, National Institutes of Health.

**Child and Adolescent Twin Study in Sweden (CATSS)**

Recruitment and phenotype definition

A detailed summary of the collection of biosamples for the Child and Adolescent Twin Study in Sweden (CATSS) is available in previous publications[16–18]. Eczema was defined if the study participant had answered YES to "Has the child been diagnosed with atopic dermatitis or atopic eczema by a doctor?". Exclusion for the controls included an affirmative response to the above criteria; if they had any dispense of R03AC, R03AK, R03BA or R03DC; if they had answered that the child had food allergies like celiac disease or lactose intolerant; or if they answered yes to any of the following: "Has or has the child had asthma, hay fever or eczema". Further details have been explained elsewhere [19].

Genotyping and imputation

Genotyping was performed in 18 batches at the SNP&SEQ Technology Platform in Uppsala, Sweden, using the Illumina PsychArray bead chip. QC filtering was performed to exclude SNPs with missingness > 2%, SNPs with more than 10% discordant genotypes across replicates or MZ pairs, SNPs out of Hardy-Weinberg equilibrium (exact test P value < $10^{-6}$), SNPs with clear batch effects or absolute MAF difference from 1000 Genomes European samples > 10%, Y-chromosome and mitochondrial SNPs, and SNPs with minor allele count < = 1. Haplotype phasing and imputation in the HRC panel version1.1 was performed using the Sanger server at the Wellcome Sanger Institute[20]. Variants with an imputation quality score of info < 0.5 or out of Hardy-Weinberg-Equilibrium P < $10^{-12}$ were excluded. Further details have been explained elsewhere[19].

Statistical analysis

RVTESTS was used to perform association testing between SNP allelic dosage and eczema status. Sex and the first four principle components were included as covariates. A kinship matrix was used to adjust for relatedness among the samples.

Acknowledgment and funding

We acknowledge The Swedish Twin Registry for access to data. The Swedish Twin Registry is managed by Karolinska Institutet and receives funding through the Swedish Research Council under the grant no 2017-00641. Financial support was also provided from the Swedish Research Council (grant no 2018-02640) and the Swedish Heart-Lung Foundation (grant no 20180512). We wish to thank the Biobank at Karolinska Institutet for professional biobank service.

**Children's Hospital of Philadelphia (CHOP)**

Recruitment and phenotype definition

The Center for Applied Genomics (CAG) has recruited ~80K pediatric patients from CHOP. Cases were defined from electronic medical records where 1) individuals were 60 days old or older with relevant ICD9 code for Atopic Dermatitis (691.8) in two or more in person visits to the hospital, on separate calendar days. Plus, two or more prescriptions for Atopic Dermatitis-related medications; or 2) individuals 60 days old or older with relevant ICD9 code for Atopic Dermatitis (691.8) in three or more in person visits, on separate calendar days. Individuals with ICD9 codes for Scabies, Wiskott-Aldrich Syndrome, Allergic Purpura or Ichtyosis congenita were excluded from the study. Controls were defined as individuals 60 days or older with two or more in person visits to the hospital over the preceding 5 years, no diagnosis codes for atopic dermatitis (691.8), no history of relevant medications and no exclusionary ICD 9 codes. Further details have been explained elsewhere[19].

Genotyping and Imputation

Genotyping was performed with either the Illumina HumanHap550 or the HH610. SNPs with minor allele frequencies of < 1%; genotyping failure rates of greater that 2% or Hardy-Weinberg P Values less that $1x10^{-6}$ were excluded from further analysis. Haplotype phasing and imputation in the HRC panel version1.1 was performed using the University of Michigan server.[20] Variants with an imputation quality score of $r^2 < 0.5$ or out of Hardy-Weinberg-Equilibrium P < 10-12 were excluded.

Statistical analysis

RVTESTS was used to perform association testing between SNP allelic dosage and eczema status. Sex, age and the first 2 principle components were included as covariates.

Acknowledgements and funding

Genotyping was funded by an Institute Development Fund and an Adele S. and Daniel S. Kubert Estate gift to the Center for Applied Genomics. Additional funding for phenotyping and analysis was provided by a U01 award from the National Institute of Health (HG006830-02).

**Copenhagen Prospective Studies on Asthma in Childhood 2000 (COPSAC2000)**

Recruitment and phenotype definition

The COPSAC2000 birth cohort study is a prospective clinical study of a birth cohort of 411 infants born to mothers with a history of asthma. The study was approved by the Ethics Committee for Copenhagen (KF 01- 289/96) and The Danish Data Protection Agency (2008-41-1754) and informed consent was obtained from both parents. AD was defined based on the Hanifin-Rajka criteria[10]. Further details have been explained elsewhere[19].

Gneotyping, imputation and statistical analysis

High throughput genome-wide SNP genotyping were performed using the Illumina infinium™ II HumanHap550 v1 and v3 platform (Illumina, San Diego). Haplotype phasing and imputation in the HRC panel version1.1 was performed using the University of Michigan server[20]. Variants with an imputation quality score of $r^2 < 0.5$ or out of Hardy-Weinberg-Equilibrium $P < 10^{-12}$ were excluded. RVTESTS was used to perform association testing, where sex, age and the first 4 principal components were included was covariates.

Acknowledgements and funding

COPSAC is funded by private and public research funds, all of which are listed at www.copsac.com. Aase and Ejnar Danielsens Fond, the Lundbeck Foundation, the Danish State Budget, the Danish Council for Strategic Research, the Danish Council for independent Research, and the Capital Region Research Foundation has provided core support for COPSAC. No pharmaceutical company was involved in the study. The funding agencies did not have any influence on design and conduct of the study; collection, management, and interpretation of the data; or preparation, review, or approval of the manuscript.

**Copenhagen Prospective Studies on Asthma in Childhood 2010 (COPSAC2010)**

Recruitment and phenotype definition

The COPSAC2010 birth cohort is a population based longitudinal clinical study of 800 pregnant women and their offspring. AD was defined based on the Hanifin-Rajka criteria[10]. Further details have been explained elsewhere[19].

Genotyping, imputation and statistical analysis

Genotyping was performed with the Illumina infinium HumanOmniExpressExome Bead chip. QC filters were applied on all individuals, where individuals with Hardy-Weinberg equilibria p values $>10^{-6}$, minor allele frequency (MAF>0.01), individual genotyping call rate > 0.95, and SNP genotyping call rate > 0.95 were retained. Haplotype phasing and imputation in the HRC panel version1.1 was performed using the University of Michigan server[20]. Variants with an imputation quality score of $r^2 < 0.5$ or out of Hardy-Weinberg-Equilibrium $P < 10^{-12}$ were excluded. Further details have been explained elsewhere[19]. RVTESTS was used to perform association testing, where sex, age and the first 4 principal components were included was covariates.

Acknowledgements and funding

COPSAC is funded by private and public research funds, all of which are listed at www.copsac.com. Aase and Ejnar Danielsens Fond, the Lundbeck Foundation, the Danish State Budget, the Danish Council for Strategic Research, the Danish Council for independent Research, and the Capital Region Research Foundation has provided core support for COPSAC. No pharmaceutical company was involved in the study. The funding agencies did not have any influence on design and conduct of the study; collection, management, and interpretation of the data; or preparation, review, or approval of the manuscript.

**DanFunD**

Recruitment

The Danish study of Functional Disorders (DanFunD) is a population-based epidemiological study of general health and fitness of individuals aged 18-69 years conducted at the Research Centre for Prevention and Health in Glostrup, Denmark. The study was approved by the Ethics Committee of Capital Region of Denmark.

Case/Control definition

Case-control status was defined by self-report from a health and lifestyle questionnaire filled in by the participants. Frequency of eczema was 5.4% among 7,298 participants.

Genotyping and imputation

DanFunD cohort individuals were genotyped using the Illumina HumanOmniExpress-24v1-0_A and HumanOmniExpress-24v1-1_A chip. We used the GenCall software application to automatically cluster, call genotypes, and assign confidence scores. The resulting raw genotype data were subjected to standard quality control methods. The inclusion criteria included the exclusion of monomorphic SNPs with a MAF ≥ 0 %, call rate ≥ 98 %, p-value for Hardy-Weinberg equilibrium $> 10^{-5}$. There were

510806 SNPs that met the previous QC criteria. All individuals with non-European ancestry were removed.

Remaining subjects were phased and imputed. The phasing was performed using Eagle autosomes and Shapit x-chromosome, and the phased haplotypes were then imputed to the Haplotype Reference Consortium (HRCr1.1, 2016) panel. The imputation was performed using the Michigan imputation server.

During the analysis, related individuals were removed leading to a percentage of individuals according to biological sex of 46.24 % males and 53.76 % females. Relatedness was studied using Identical By Descent method, filtering out pairs of individuals with a pi_hat coefficient larger than 0.1875.

Statistical analysis

Genome-wide association analysis was performed using RVtest (version 2.0.6) for 349 cases and 6213 controls with genetic and phenotypic data while including sex and 10 principal components as covariates.

Acknowledgements and Funding

This study was supported by TrygFonden (7-11-0213), the Lundbeck Foundation (R155-2013-14070), Novo Nordisk Foundation (NNF15OC0015896).

**Danish National Birth Cohort (DNBC)**

Recruitment

The Danish National Birth Cohort (DNBC) is a population-based cohort of more than 100,000 pregnancies, recruited in the years 1996-2002[21]. Extensive phenotype information was collected by computer-assisted telephone interviews twice during pregnancy as well as 6 and 18 months after delivery. Additional questionnaire-based follow-up surveys are conducted at regular intervals. The DNBC mothers provided written informed consent on behalf of themselves and their children. The study protocol was approved by the Danish Scientific Ethical Committee and the Danish Data Protection Agency.

Case/Control definition

Cases with early onset eczema were identified from the 18 months telephone interview data using an algorithm specifically developed for this purpose[22]. In addition, children with a positive response to both of the following two questions from the 7 year survey were included in the case group: 1) "Has a doctor ever said that your child had eczema, also known as allergic rash?" and 2) "Has your child ever had an itchy *rash which was coming and going for at least 6 months?*". Finally, children with ICD10 diagnosis code L20 in the Danish Hospital Discharge Register were also included in the case group. Controls were required not to have any eczema or eczema symptoms recorded in interview, questionnaire, or register data.

Genotyping and imputation

GWAS data were generated for 3,840 individuals from the DNBC (mothers and their children) in a study of prematurity and its complications within the Gene Environment Association Studies (GENEVA) consortium. Genotyping was performed using the Illumina Human660W-Quad BeadChip. Prior to imputation, we required participants to have a genotype call rate >97%, and we excluded SNPs based on a missing rate >2%, deviation from Hardy-Weinberg equilibrium in controls ($P<10^{-3}$), minor allele frequency <0.5%. We also converted the genotype data from NCBI build 36 to NCBI build 37 and aligned all genotypes to the forward strand. Finally, we excluded SNPs that did not match known variant positions in the 1000 Genomes project reference data. The remaining 529128 SNPs were used for imputation.

We used a two-step procedure to impute unobserved genotypes using phased haplotypes from the integrated Phase I release of the 1000 Genomes Project[23] (v3.20101123, ALL populations, no monomorphic/singletons). In a first prephasing step, we used SHAPEIT[24] to estimate the haplotypes for our study samples. In a second step, we imputed the missing alleles for additional SNPs directly onto these phased haplotypes using IMPUTE2[25]. Eczema information and genome-wide genotype and imputed data were available for 1,631 children.

Statistical Analysis

Genome-wide association analysis was carried out using SNPTEST[26] for the 224 cases and 1407 controls with genetic and phenotypic data. Summary statistics were available for 30,071,690 variants that were successfully analysed.

Acknowledgements and Funding

We are very grateful to the women and children taking part in the DNBC. The DNBC was established with the support of a major grant from the Danish National Research Foundation. Additional support for the DNBC has been obtained from the Danish Pharmacists' Fund, the Egmont Foundation, the March of Dimes Birth Defects Foundation, the Augustinus Foundation and the Health Fund of the Danish Health Insurance Societies. The generation of GWAS genotype data for the DNBC samples was carried out within the GENEVA consortium, with funding provided through the NIH Genes, Environment and Health Initiative (GEI) (U01HG004423, U01HG004438, U01HG004446).

**ECHRS**

Recruitment

Details of the methods of ECRHS I and ECRHS II, a multicenter international cohort study, have been published elsewhere[27]. Participants within the ECRHS were eligible for inclusion in this analysis if they were identified by random sampling of those who fulfilled the following criteria: 1) lived in centers that took part in genome-wide genotyping initiative under the auspices of GABRIEL[7] AND 2) were initially selected to take part in the ECRHS clinical measurements as part of the random sample (i.e. not specifically selected for inclusion because of any pre-existing disease). Each participating center obtained ethical permission from the appropriate local committee.

Case/Control definition

Cases were those answering positively to the question ‘Have you ever had an itchy rash that was coming and going for at least 6 months?’ AND yes to ‘Have you had this itchy rash in the last 12 months?’ during ECHRS II (aged 27-58). Further information on the distribution of eczema within the cohort is available[28].

Genotyping and imputation

Subjects were genotyped with the Human 610 Quad Chip (ILLUMINA). Genotypes were called using BeadStudio. Criteria for exclusion of variants were: a call rate below 95%, a MAF < 0.01 and a HWE p-value < 0.0001. Other QC criteria applied were: exclusion of males with high X heterozygosity, sex discrepancies, IBS analysis for relatedness and ancestry analysis using PCA. Imputation was done using IMPUTE2[29] considering the haplotypes from the 1000 Genomes Project Phase I version v3.20101123 as a reference.

Statistical analysis

Genome-wide association analysis of atopic eczema was carried out in SNPTEST V2.4.1[26] regressing expected allelic dosage on case-control status, adjusted for center, sex and the first four principal components informative of European ancestry.

Acknowledgements and Funding

The co-ordination of ECHRS II was supported by the European Commission, as part of their Quality of Life programme, The genotyping was funded through the EU funded GABRIEL initiative – GRANT Number 018996.

Financial support for ECHRS II: Albecete: Fondo de Investigaciones Santarias (FIS) (grant code: 97/0035-01, 99/0034-01 and 99/0034-02), Hospital Universitario de Albacete, Consejeria de Sanidad. Barcelona: SEPAR, Public health Service (grant code: R01 HL62633-01), Fondo de Investigaciones Sanatrias (FIS) (grant code: 97/0035-01, 99-0034-01 and 99/0034-02), CIRIT (grant code: 1999SGR 002541), Red Respira ISCII; CIBER Epidemiology y Salud Pública (CIBERESP), Spain. Basel: Swiss National Science Foundation, Swiss Federal Office for Education & Science, Swiss National Accident Insurance Fund (SUVA), USC NIEHS Center grant 5P30 ES07048. Bergen: Norwegian Research Council, Norwegian Asthma & Allergy Association (NAAF), Glaxo Wellcome AS, Norway Research Fund. Erfurt: Helmholtz Center Munich – National Research Center for Environment & Health, Deutsche Forschungsgemeinschaft (DFG) (grant code FR 1526/1-1). Galdakao: basque health Dept. Grenoble: Programme Hopitalier de Recherche Clinique-DRC de Grenoble 2000 no. 2610, ministry of Health, Direction de la Recherche Clinique, CHU de Grenoble, Ministere de l'Emploi et de la Solidarite, Direction Generale de la Sante, Comite des Maladies Respiratoires de l'Isere. Hamburg: Helmholtz Center Munich – National Research Center for Environment & Health, Deutsche Forschungsgemeinschaft (DFG) (grant code MA 711/4-1). Ipswich and Norwich: Asthma UK (formerly known as national Asthma Campaign). Huelva: Fondo de Investigaciones Santarias (FIS) (grant code: 97/0035-01, 99/0034-01 and 99/0035-02). Olviedo: Fondo de Investigaciones Santarias (FIS) (grant code: 97/0035-01, 99/0034-01 and 99/0035-02). Paris: Ministere de l'Emplou et de la Solidarite, Direction Generale de la Sante, UCB-Pharma (France), Aventis (France), Glaxo France, programme Hospitalier de Recherche Clinique-DRC de Grenoble 2000 no. 2610, Ministry of helath, Direction de la Recherche Clinique, CHU de Grenoble. Tartu: Estonian Science Foundation. Umeå: Swedish Heart Lung Foundation, Swedish Foundation for Health care Sciences & Allergy Research, Swedish Asthma & Allergy Foundation, Swedish Cancer & Allergy Foundation.

Financial support for ECHRS I: Ministerère de la Santé, Glaxo France, Institut Pneumologique d'Acuitaine, Contrat de Plan Etat-Région Languedoc-Rousillon, CNMATS, CNMRT (90MR/10, 91AF/6), Ministre delegué de la santé, RNSP, France; Helmholtz Center Munich, Bundesministerium für Forschung und Technologie, Bonn, Germany; Norwegian Research Council project no. 101422/310; Minstero Sanidad y Consumo FIS (grants #91/0016060/00E-05E and #93/0393), grants from Hospital General de Albacete, Hospital General Juan Ramón Jiménenz, Consejeria de Sanidad Principado de Asturias, Spain; The Swedish Medical research Council, the Swedish Heart Lung Foundation, the

Swedish Association against Asthma and Allergy; Swiss National Science Foundation grant 4026-28099; National Asthma Campaign, British Lung Foundation, Department of Health, South Thames Region Health Authority, UK.

**Estonian Biobank**

Case/control definition

The EstBB is a population-based biobank with around 200,000 participants. The 198 K data freeze was used for the analyses described here. All biobank participants have signed a broad informed consent form. Participants with eczema were identified using the ICD-10 code system (information on ICD codes is obtained via linking with the national Health Insurance Fund and other databases[30]. Individuals with eczema were identified using the ICD-10 codes including L20; controls excluding L20, L23.9, L24.9, L25.9, and L30.9, resulting in 16170 cases (74.1% female, 25.9% male; average age 43.7) and 150546 controls (63.7% female, 36.8% male; average age 49.4).

Genotyping and statistical analysis

All EstBB participants have been genotyped at the Core Genotyping Lab of the Institute of Genomics, University of Tartu, using Illumina Global Screening Array v1.0 and v2.0. Samples were genotyped and PLINK format files were created using Illumina GenomeStudio v2.0.4. Individuals were excluded from the analysis if their call-rate was <95% or if sex defined based on heterozygosity of X chromosome did not match sex in phenotype data. Before imputation, variants were filtered by call-rate <95%, HWE p value < 1e-4 (autosomal variants only), and minor allele frequency <1%. Variant positions were updated to b37 and all variants were changed to be from TOP strand using GSAMD-24v1-0_20011747_A1-b37.strand.RefAlt.zip files from https://www.well.ox.ac.uk/~wrayner/strand/ webpage. Prephasing was done using Eagle v2.3 software38 (number of conditioning haplotypes Eagle2 uses when phasing each sample was set to:–Kpbwt=20000) and imputation was done using Beagle v.28Sep18.79339 with effective population size ne = 20,000. Population specific imputation reference of 2297 WGS samples was used[31].

Association analysis was carried out using SAIGE (v0.43.1) software implementing mixed logistic regression model without LOCO option, using sex, age, age_sq and ten PCs as covariates in step I.

Acknowledgements and Funding

EstBB GWAS analysis is supported by Personal research funding: Team grant PRG1291. Computations were performed in the High Performance Computing Center, University of Tartu. Work in the Estonian Genome Center was partially supported by the EU through the ERDF, project no. 2014-2020.4.01.15-0012 “Gentransmed”.

**FinnGen (Release 3)**

Recruitment and Case/Control definition

FinnGen is a nation-wide study academic industrial collaboration launched in Finnland in 2017 which combines genotype data from Finnish biobanks and digital health record data from Finnish health registries (https://finngen.gitbook.io/documentation/). Phenotypes were derived from ICD codes in Finnish national hospital registries. We downloaded the summary statistics for the phenotype L20 Atopic dermatitis, totalling 2,921 cases and 132,591 controls, from the publicly available FinnGen data release 3.

Genotyping

FinnGen individuals were genotyped with Illumina and Affymetrix chip arrays (Illumina Inc., San Diego, and Thermo Fisher Scientific, Santa Clara, CA, USA). Genotype calls were made with GenCall and zCall algorithms for Illumina and AxiomGT1 algorithm for Affymetrix data. Chip genotyping data produced with previous chip platforms and reference genome builds were lifted over to build version 38 (GRCh38/hg38) following the protocol described here[32]. In sample-wise quality control, individuals with ambiguous gender, high genotype missingness (>5%), excess heterozygosity (+-4SD) and non-Finnish ancestry were excluded. In variant-wise quality control variants with high missingness (>2%), low HWE P-value (<1e-6) and minor allele count, MAC<3 were excluded. Prior to imputation, chip genotyped samples were pre-phased with Eagle 2.3.5 (https://data.broadinstitute.org/alkesgroup/Eagle/) with the default parameters, except the number of conditioning haplotypes was set to 20,000.

Imputation

Chip genotype data were imputed using the population-specific SISu v3 imputation reference panel of 3,775 whole genomes with Beagle 4.1 (version 08Jun17.d8b) as described in the following protocol[33]. Variant call set was produced with GATK HaplotypeCaller algorithm by following GATK best-practices for variant calling. Genotype-, sample- and variant-wise QC was applied in an iterative manner by using the Hail framework v0.1 and the resulting high-quality WGS data for 3,775 individuals were phased with Eagle 2.3.5 as described above. Post-imputation quality-control involved checking expected conformity of the imputation INFO-value distribution, MAF differences between the target dataset and the imputation reference panel and checking chromosomal continuity of the imputed genotype calls.

Statistical analysis

GWAS analysis was performed using SAIGE software, a mixed model logistic regression R/C++ packages which accounts for related samples. Sex, age, 10 principal components and genotyping batch were included as covariates in the model.

Acknowledgements and Funding

We want to acknowledge the participants and investigators of the FinnGen study.

**The Genes-environments & Admixture in Latino Americans (GALA II) study**

Recruitment

The Genes-environments & Admixture in Latino Americans (GALA II) study is a cross-sectional case-control study of asthma in Latino children and young adults from five urban centers across the mainland United States of America (Denver, CO; Chicago, IL; Bronx, NY; Houston, TX; San Francisco Bay Area, CA) and Puerto Rico. Participants were 8-21 years old at the time of recruitment and had no history of other lung or chronic illnesses aside from atopy and/or allergy-related diseases. Participants were required to identify themselves and their four grandparents as Hispanics/Latinos.

Asthma cases were included if i) had a physician-diagnose of asthma and ii) had experienced coughing, wheezing, or dyspnea and/or had been treated with asthma medication within the last two years. Non-asthmatic controls were eligible if they had no reported history of asthma, use of medication for allergies, or any asthma-related symptoms of coughing, wheezing, or dyspnea during their lifetime; active smokers were also excluded[34–36].

Detailed clinical measures (bronchodilator response testing, spirometry, exhaled nitric oxide, skin pigmentation, skin prick testing, complete blood counts, and IgE measurements), biologic specimens (whole blood, saliva, and nasal epithelium), geocoded air pollution measures, and questionnaire-based information regarding social and environmental risk factors were collected.

This study has been approved by the Human Research Protection Program Institutional Review Board of the University of California, San Francisco (San Francisco, USA, and each study site's institutional review board (IRB) approved the GALA II protocols (UCSF IRB approval No. 10-00889). All participants/parents provided written assent/consent, respectively.

### Case/Control definition

For the current study, case status was defined based on an affirmative response to the following question of the survey: "Has a doctor ever diagnosed the child with eczema or atopic dermatitis?". Controls were defined as individuals with absence of eczema based on a negative answer to the question. Cases were individuals that had an affirmative response. Eczema status was defined regardless of the asthma status. Specifically, cases were defined as asthma patients with atopic dermatitis, and controls were defined as asthma patients without atopic dermatitis plus individuals without asthma and atopic dermatitis.

### Genotyping and imputation

Genotyping and quality control have been described elsewhere[37]. Briefly, genome-wide genotyping was performed with the Affymetrix Axiom LAT 1 (World Array 4) and LAT plus HLA genome wide arrays, and genotype calling was carried out with the Affymetrix Power Tools software. Quality control was performed by removing SNPs with call rates <95% and/or deviating from the Hardy Weinberg equilibrium in the controls without asthma ($p < 10\text{-}6$). Samples with call rates <97%, discrepancy between genetic sex and reported sex, or cryptic relatedness (proportion of identity by descent > 0.3) were removed[37]. Data was imputed with the Michigan Imputation Server, using the reference panel of the Haplotype Reference Consortium 1.1[38]. Pre phasing was performed using SHAPEIT v2.r790[39], and imputation was conducted with Minimac3[40].

### Statistical analysis

Association between eczema and genetic variation was tested using RVTESTS software[41] through logistic regression models adjusted by sex and the two principal components of the genotype matrix to adjust for differences in genetic ancestry between cases and controls.

### Acknowledgements

The authors acknowledge the families and patients for their participation and thank the numerous researchers, health care providers, and community clinics for their support and participation in the GALA II study. In particular, the authors thank the study coordinator Sandra Salazar; the principal investigators involved in the recruitment: Kelley Meade, Harold J. Farber, Pedro C. Avila, Denise Serebrisky, Shannon M. Thyne, Emerita Brigino-Buenaventura, William Rodriguez-Cintron, Saunak Sen, Rajesh Kumar, Michael Lenoir, and Luisa N. Borrell; and the recruiters who obtained the data: Duanny Alva, Gaby Ayala-Rodriguez, Lisa Caine, Elizabeth Castellanos, Jaime Colon, Denise DeJesus, Blanca Lopez, Brenda Lopez, Louis Martos, Vivian Medina, Juana Olivo, Mario Peralta, Esther Pomares, Jihan Quraishi, Johanna Rodriguez, Shahdad Saeedi, Dean Soto, and Ana Taveras.

The Genes-environments and Admixture in Latino Americans (GALA II) study and the Study of African Americans, Asthma, Genes and Environments (SAGE) were supported by the Sandler Family Foundation, the American Asthma Foundation, the RWJF Amos Medical Faculty Development Program, Harry Wm. and Diana V. Hind Distinguished Professor in Pharmaceutical Sciences II, the National Heart, Lung, and Blood Institute of the National Institutes of Health (R01HL117004, R01HL128439, R01HL135156, X01HL134589, R01HL141992, and R01HL141845), National Institute of Health and Environmental Health Sciences (R01ES015794 and R21ES24844); the National Institute on Minority Health and Health Disparities (NIMHD) (P60MD006902, R01MD010443, and R56MD013312); the National Institute of General Medical Sciences (NIGMS) (RL5GM118984); the Tobacco-Related Disease Research Program (24RT-0025 and 27IR-0030); and the National Human Genome Research Institute (NHGRI) (U01HG009080) to EGB.

This work was also funded by the Spanish Ministry of Science and Innovation MCIN/AEI/10.13039/501100011033 (PID2020-116274RB-I00) and by Instituto de Salud Carlos III, Spain and the European Regional Development Fund "ERDF A way of making Europe" by the European Union (CB/06/06/1088). MP-Y and EH-L were funded by MCIN/AEI/10.13039/501100011033 and the European Social Fund "ESF Investing in your future" (Ramón y Cajal Program RYC-2015-17205 to MP-Y and PRE2018-083837 to EH-L). AE-O was funded by a fellowship from the Spanish Ministry of Science, Innovation, and Universities (MICIU) and Universidad de La Laguna (ULL), under de M-ULL agreement.

**Generation R**

Recruitment

The Generation R Study is a population-based prospective cohort study of pregnant women and their children from fetal life onwards in Rotterdam, The Netherlands[42,43]. All children were born between April 2002 and January 2006, and currently followed until young adulthood. Of all eligible children in the study area, 61% were participating in the study at birth. Cord blood samples including DNA have been collected at birth. The study protocol was approved by the Medical Ethical Committee of the Erasmus Medical Centre, Rotterdam (MEC 217.595/2002/20). Written informed consent was obtained from parents of all participants.

Case/Control definition

Information about ever physician diagnosed eczema (no; yes) and doctor-attendance for eczema in the past 12 months (no; yes) was collected by a questionnaire at age 6 years. Response rates for this questionnaire was 68%. Cases were defined as ever physician diagnosed eczema with doctor attendance in the last 12 months. We defined controls as the individuals who responded with "no" to either of these two questions.

Genotyping and imputation

Samples were genotyped using Illumina Infinium II HumanHap610 Quad Arrays following standard manufacturer's protocols. Intensity files were analysed using the Beadstudio Genotyping Module software v.3.2.32 and genotype calling based on default cluster files. Any sample displaying call rates below 97.5%, excess of autosomal heterozygosity (F<mean-4SD), and mismatch between called and phenotypic gender were excluded. In addition, individuals identified as genetic outliers by the IBS clustering analysis (>3 standard deviations away from the HapMap CEU population mean) were excluded from the analysis. Genotypes were imputed for all polymorphic SNPs from phased haplotypes in autosomal chromosomes using the 1000 Genomes GIANTv3 panel in minimac[44]. Twins were excluded from the analyses. Ethnicity was grouped into Caucasians and non-Caucasians, based on genetic ancestry. Ancestry determination analysis included the genomic data of all Generation R individuals merged with the three reference panels of the HapMap Project Phase II (YRI, CEU and CHB/JPT).

Statistical Analysis

The sample of 3,282 individuals from the Generation R study used for the analysis included a majority of individuals with Caucasian ancestry (63.4%). Caucasians and non-Caucasians were subsequently analysed separately. Numbers with genetic and phenotypic data were, Caucasians: 332cases and 1749 controls, non-Caucasians: 305 cases and 896 controls.

Association between atopic dermatitis phenotype and GWAS SNPs was performed using a regression framework adjusting for population stratification in the Generation R cohort using mach2dat[45] as implemented in GRIMP[46]. Since Generation R is a population-based study of unrelated individuals of different ethnic background, 4 principal components were used for the Caucasian subpopulation and 4 were used for the non-Caucasian subpopulation, both analyses had a Genomic Inflation Factor of 1.

Acknowledgements and Funding

The Generation R Study gratefully acknowledges the contributions of the children and their parents, the general practitioners, the hospitals and the midwives and pharmacies in Rotterdam. We would like to thank A. Abuseiris, K. Estrada, Dr. T. A. Knoch, and R. de Graaf as well as their institutions Biophysical Genomics, Erasmus MC Rotterdam, The Netherlands, and especially the national German MediGRID and Services@MediGRID part of the German D-Grid, both funded by the German Bundesministerium fuer Forschung und Technology under grants #01 AK 803 A-H and # 01 IG 07015 G, for access to their grid resources. We thank M. Jhamai, M. Ganesh, P. Arp, M. Verkerk, L. Herrera and M. Peters for their help in creating, managing and performing quality control for the genetic database. Also, we thank K. Estrada and C. Medina-Gomez for their support in the creation and analysis of imputed data. The Generation R Study is conducted by the Erasmus Medical Center in close collaboration with the School of Law and the Faculty of Social Sciences of Erasmus University Rotterdam, the Municipal Health Service, Rotterdam area, the Rotterdam Homecare Foundation and the Stichting Trombosedienst & Artsenlaboratorium Rijnmond (STAR-MDC; Rotterdam). The generation and management of genotype data for the Generation R Study were performed at the Genetic Laboratory of the Department of Internal Medicine at Erasmus Medical Center. The Generation R Study is made possible by financial support from the Erasmus Medical Center (Rotterdam), the Erasmus University Rotterdam and the Netherlands Organization for Health Research and Development (ZonMw; 21000074). Vincent Jaddoe and Liesbeth Duijts received funding for projects from the European Union's Horizon 2020 research and innovation programme (LIFECYCLE, grant agreement No 733206, 2016; EUCAN-Connect grant agreement No 824989; ATHLETE, grant agreement No 874583). Dr Vincent Jaddoe received an additional grant from the Netherlands Organization for Health Research and Development (VIDI 016.136.361) and Consolidator Grant from the European Research Council (ERC-2014-CoG-648916). The study sponsors had no role in the study design, data analysis, interpretation of data, or writing ofthis report.

**GENEVA**

Recruitment and phenotype definition

Atopic dermatitis patients were recruited from tertiary dermatology clinics based at three centers (Technische Universität Munich, as part of the GENEVA study, University of Kiel, University of Bonn). German controls were obtained from the population-representative PopGen biorepository[47] and the population-based KORA study[48,49]. Atopic dermatitis was diagnosed on the basis of a skin examination by experienced dermatologists according to standard criteria. Further details have been explained elsewhere[19].

Genotyping, imputation and statistical analysis

Prior to imputation SNPs with low genotyping rate (< 95%), low minor allele frequency (< 1%), strong deviation from HardyWeinberg equilibrium ($p< 10^{-8}$) and differential call rate between cases and controls were excluded. Further quality control measures applied have been previously described elsewhere[19]. Haplotype phasing and imputation in the HRC panel version1.1 was performed using the University of Michigan server[20] .Variants with an imputation quality score of $r^2 < 0.5$ or out of Hardy-WeinbergEquilibrium $P < 10^{-12}$ were excluded. RVTESTS was used to test the association between SNP allelic dosage and eczema status. Sex, age and the first 4 principle components were included as covariates.

Acknowledgements and funding

The project received infrastructure support through the DFG Clusters of Excellence "inflammation at interfaces" (grants EXC306 and EXC306/2), and was supported by the German Federal Ministry of Education and Research (BMBF) within the framework of the e:Med research and funding concept (sysINFLAME, grant # 01ZX1306A), and the PopGen 2.0 network (01EY1103). The KORA study was initiated and financed by the Helmholtz Zentrum München – German Research Center for Environmental Health, which is funded by the German Federal Ministry of Education and Research (BMBF) and by the State of Bavaria. Furthermore, KORA research was supported within the Munich Center of Health Sciences (MC-Health), Ludwig Maximilians-Universität, as part of LMUinnovativ.

**GENUFADext (SHIP1)**

Recruitment

Atopic dermatitis patients were recruited at Charite Universitatsmedizin Berlin, Germany for the extended GENUFAD study (GEnetic analysis of NUclear Families with Atopic Dermatitis) and have been described previously[50,51] (Berlin cases of set 1). Children of the extended GENUFAD study were unrelated individuals recruited based on moderate to severe atopic dermatitis and an age of onset below two years. A total of 417 unrelated atopic dermatitis patients were included in the present study. All controls originated from the population-based Study of Health in Pomerania (SHIP)[52], which included individuals in the North-Eastern part of Germany. The SHIP set was split for two case- control studies by a random function. 1667 unrelated individuals were included in SHIP-1

Case/Control definition

All cases had early onset eczema (< 2years) diagnosed by a physician according to standard criteria[10]. Controls were unrelated individuals from the population-based SHIP cohort.

Genotyping and imputation

All cases and controls were genotyped with Affymetrix Genome-Wide Human SNP Array 6.0. Individuals with a call rate < 0.95 were excluded from the study. In addition, samples were excluded when the gender estimated from X-chromosome heterozygosity did not match the clinical records. SNPs were filtered according to the following criteria: i) low call rate (< 0.95 in cases or controls), ii) low allele frequency (MAF < 0.01 in cases or controls), iii) genotypes out of Hardy-Weinberg equilibrium (p < 0.00001 in cases or p < 0.0005 in controls). Additionally, SNPs with a call rate < 0.99 were excluded if having MAF < 0.05 or if they were out of Hardy-Weinberg equilibrium (p < 0.001). Only SNPs fulfilling the above mentioned QC were used in subsequent steps. Genotypes of cases and controls were recoded to the "+" using the -flip command from PLINK[15]. In addition, markers were excluded if: i) 3 alleles were detected, ii) the allele frequency in the SHIP control population differed by more than 0.1 compared with the frequency in 379 Europeans available from the 1000 Genomes project. Haplotype phasing and imputation in the HRC panel version1.1 was performed using the University of Michigan server. Variants with an imputation quality score of $r^2 < 0.5$ or out of Hardy-Weinberg-Equilibrium $P < 10^{-12}$ were excluded. Principal component (PC) analysis was performed with EIGENSTRAT (SMARTPCA)[53].

Statistical Analysis

A score test implemented in RVTESTS was used to test the association between SNP allelic dosage and eczema status. Sex and the first 2 principle components were included as covariates.

Acknowledgements and Funding

We thank all individuals and families for their participation in this study. We thank all physicians and nurses involved in patient recruitment for their valuable contribution to the study. We are grateful to the laboratory technicians Christina Flachmeier and Theresa Thuß for their excellent technical assistance. The study was funded by the German Ministry of Education and Research (BMBF) through the Clinical Research Group for Allergy at Charité Berlin, the National Genome Research Network (NGFN). The SHIP authors are grateful to Mario Stanke for the opportunity to use his server cluster for

SNP imputation. We thank all staff members and participants of the SHIP studies, as well as all of the genotyping staff for generating the SHIP SNP data set. SHIP is part of the Community Medicine Research net of the University of Greifswald, Germany, which is funded by the Federal Ministry of Education and Research (grants no. 01ZZ9603, 01ZZ0103, and 01ZZ0403), the Ministry of Cultural Affairs as well as the Social Ministry of the Federal State of Mecklenburg-West Pomerania, and the network ‘Greifswald Approach to individualized Medicine (GANI_MED)’ funded by the Federal Ministry of Education and Research (grant 03IS2061A). Genome-wide data were supported by the Federal Ministry of Education and Research (grant 03ZIK012) and a joint grant from Siemens Healthcare, Erlangen, Germany, and the Federal State of Mecklenburg–West Pomerania. The University of Greifswald is a member of the 'Center of Knowledge interchange' program of the Siemens AG and the Caché Campus program of the interSystems GmbH.

**GENUFAD (SHIP2)**

Recruitment

All atopic dermatitis patients were recruited at Charité Universitätsmedizin Berlin for the GENUFAD study (GEnetic analysis of NUclear Families with Atopic Dermatitis) and have been previously described in a previous GWAS[50,51] (Set 2 in original study). 270 German families were recruited through two affected siblings with an age of onset below two years of age and moderate to severe disease expression. One affected child was selected from each family and 259 atopic dermatitis patients were included in the present study.

All controls originated from the population-based Study of Health in Pomerania (SHIP)[52], which recruited individuals in the North-Eastern part of Germany. The SHIP set was split for two case- control studies by a random function. 1792 unrelated individuals were included in SHIP-2.

Case/Control definition

All cases had an early onset (< 2years) physician's diagnosis of atopic dermatitis made according to standard criteria[10]. Controls were unrelated individuals from the population-based SHIP cohort.

Genotyping and imputation

All cases were genotyped with Affymetrix 500K arrays and only samples with high call rate (> 0.95) were used for the analysis. Controls were genotyped with Affymetrix Genome-Wide Human SNP Array 6.0 and were excluded when call rate < 0.96. For both case and control groups, samples were excluded when the gender estimated from X-chromosome heterozygosity did not match the clinical records. SNPs from the 500K array were filtered as previously described[50] according to the following criteria: i) low call rate (< 0.95), ii) low allele frequency (MAF < 0.01), iii) Mendelian errors in 5 or more families, iv) unlikely genotypes in more than 5 families (double recombinants as detected by Merlin[54]), v) founder genotypes out of Hardy-Weinberg equilibrium (p < 0.00001). Additionally, SNPs with a call rate < 0.99 were excluded if having MAF < 0.05 or if they were out of Hardy-Weinberg equilibrium (p < 0.001). SNPs on the Human SNP Array 6.0 were excluded if having: i) low call rate (< 0.97), ii) low allele frequency (MAF < 0.01), iii) founder genotypes out of Hardy-Weinberg equilibrium (p < 0.0005). Additionally SNPs with a call rate < 0.99 were excluded if having MAF < 0.05 or if they were out of Hardy-Weinberg equilibrium (p < 0.001). Only SNPs fulfilling the above mentioned QC on both arrays were used in subsequent steps, and the rest of non-overlapping SNPs were excluded.

Genotypes of cases and controls were recoded to the "+" using the -flip command from PLINK[15] and merged. Additionally, markers were excluded if: i) 3 alleles were detected, ii) the allele frequencies in the SHIP control population differed by more than 0.1 compared to the 379 Europeans available from the 1000 Genomes project. Haplotype phasing and imputation in the HRC panel version1.1 was performed using the University of Michigan server. Variants with an imputation quality score of $r^2 < 0.5$ or out of Hardy-Weinberg-Equilibrium $P < 10^{-12}$ were excluded. Principal component (PC) analysis was performed with EIGENSTRAT (SMARTPCA)[53].

Statistical Analysis

A score test implemented in RVTESTS was used to test the association between SNP allelic dosage and eczema status. Sex and the first 2 principle components were included as covariates.

Acknowledgements and Funding

We thank all individuals and families for their participation in this study. We thank all physicians and nurses involved in patient recruitment for their valuable contribution to the study. We are grateful to the laboratory technicians Christina Flachmeier and Theresa Thuß for their excellent technical assistance. The study was funded by the German Ministry of Education and Research (BMBF) through the Clinical Research Group for Allergy at Charité Berlin, the National Genome Research Network (NGFN). The SHIP authors are grateful to Mario Stanke for the opportunity to use his server cluster for SNP imputation. We thank all staff members and participants of the SHIP studies, as well as all of the genotyping staff for generating the SHIP SNP data set. SHIP is part of the Community Medicine Research net of the University of Greifswald, Germany, which is funded by the Federal Ministry of Education and Research (grants no. 01ZZ9603, 01ZZ0103, and 01ZZ0403), the Ministry of Cultural Affairs as well as the Social Ministry of the Federal State of Mecklenburg-West Pomerania, and the network 'Greifswald Approach to individualized Medicine (GANI_MED)' funded by the Federal Ministry of Education and Research (grant 03IS2061A). Genome-wide data were supported by the Federal Ministry of Education and Research (grant 03ZIK012) and a joint grant from Siemens Healthcare, Erlangen, Germany, and the Federal State of Mecklenburg–West Pomerania. The University of Greifswald is a member of the 'Center of Knowledge interchange' program of the Siemens AG and the Caché Campus program of the interSystems GmbH.

**GERA**

Recruitment

The Resource for Genetic Epidemiology Research on Aging (GERA) cohort was designed to facilitate research on the genetic and environmental factors that affect health and disease by linking together clinical data from electronic health records, survey data on demographic and behavioural factors, and environmental data from various sources, with genetic data from biospecimens collected from participants.

The GERA Cohort consists of more than 100,000 adults who are members of the Kaiser Permanente Medical Care Plan, Northern California Region (KPNC), and participants in its Research Program on

Genes, Environment and Health (RPGEH). KPNC is an integrated health care delivery system with a population of about 3.3 million people in northern California. The membership of KPNC is representative of the general population in the 14-county area in which facilities are located, although the membership is underrepresented for the extremes of income at both ends of the spectrum. The RPGEH utilizes the longitudinal electronic health records (EHR) of KPNC to obtain clinical, laboratory, imaging and pharmacy information on all cohort members, to which personal demographic, behavioural and health characteristics have been added through member surveys.

The GERA Cohort is a subsample of the longitudinal cohort enrolled in the Kaiser Permanente RPGEH. The RPGEH cohort includes about 400,000 survey participants of whom about 200,000 have provided broad consent and a sample of saliva or blood for use in studies of genetic and environmental factors in health and disease. The GERA Cohort was developed from a mailed survey sent to all adult members of KPNC who had been members for two years or more in 2007. All survey respondents were contacted and asked to complete a consent form; those who completed consent forms were asked to provide a saliva sample. Additional participants were added to the RPGEH through inclusion of the Northern California sample of the California Men's Health Study (CMHS) cohort of about 40,000 men from KPNC, ages 45-69 years old at the time of the CMHS survey in 2002-2003. The CMHS participants contributed about 15,400 saliva samples to the RPGEH and were eligible for inclusion in the GERA Cohort. CMHS participants were included according to the same sampling design as for the RPGEH cohort as a whole. Specifically, all minority participants were selected for inclusion in order to maximize representation of minorities in the GERA Cohort, and Non-Hispanic White participants were selected at random to complete the sample of 110,266 GERA Cohort participants.

Additional details can be found at https://www.ncbi.nlm.nih.gov/projects/gap/cgi-bin/study.cgi?study_id=phs000674.v3.p3

<u>Definition of cases and controls</u>

Cases were defined as those with at least 2 medical record ICD-9 codes for atopic dermatitis (691.8) on separate dates and controls were defined as those with no medical codes for atopic dermatitis. We restricted our analysis to non-Hispanic whites, which represented the majority of the cohort.

<u>Genotyping and imputation</u>

To maximize genome-wide coverage of common and less common variants, four custom Affymetrix Axiom arrays[55,56] were designed for individuals of non-Hispanic white (EUR), East Asian (EAS), African American (AFR), and Latino (LAT) race/ethnicity. The number of SNPs varied among arrays, ranging from 674,518 on the EUR array to 893,631 on the AFR array[55]. A total of 254,438 SNPs were common to all four arrays. Genotyping was performed at the University of California, San Francisco and is described in detail in Kvale et al[57].

High-quality genotype data for the GERA cohort was obtained by systematic examination and removal of SNP genotypes according to a specific protocol, as described in detail elsewhere[57]. For the genetic structure analyses, only SNPs that were common across all four arrays and that had a call rate of at least 99.5% were considered. This set also excluded SNPs that showed extreme deviation from Hardy–Weinberg equilibrium ($P < 1\times10^{-25}$). This resulted in a set of 144,799 high-performing SNPs used in further analyses of population structure and admixture.

High-density genotyping was conducted at UCSF using custom designed Affymetrix Axiom arrays, as described by Hoffmann et al[55,56]. To maximize genome-wide coverage of common and less common variants, four specific arrays were designed for individuals of Non-Hispanic White (EUR), East Asian (EAS), African-American (AFR), and Latino (LAT) race/ethnicity. There was broad overlap among the

SNPs on the arrays, which were designed using a hybrid greedy imputation algorithm[55] applied to genotype information validated by Affymetrix from the 1000 Genomes Project. However, in order to capture low frequency variants specific to particular race-ethnicity groups, SNP content varies between arrays. A more detailed description of the process of genotyping and results are found in Kvale et al., 2015[57]. Description of the analyses of population structure and development of principal components for adjustment of population structure is provided in Banda et al [58].

Imputation was performed on an array-wise basis. Genotypes were first pre-phased with SHAPEIT v2.r727[59], with cryptic relatives included to improve phasing. The 1000 Genomes Project (Phase I integrated release, August 2012; singletons removed; http://www.internationalgenome.org) was used as a cosmopolitan reference panel, and data were imputed with IMPUTE2 v2.3.0[25,44,60].

Statistical analysis and software (used to perform association testing, covariates included in regression model)

Using PLINK v1.90b3b[15] for each SNP, we conducted a logistic regression of eczema adjusting for the SNP (the coefficient of interest), sex, and the first 10 ancestry PCs.

**GINIplus/LISA north**

Recruitment

3,042 new born infants were recruited between September 1995 and June 1998 in 8 different maternity wards in Wesel for the prospective German Infant study on the influence of Nutrition Intervention plus environmental and genetic influences on allergy development (GINIplus) north study. The study region Wesel is a rural area in Western Germany. Data on parental allergic diseases, pet contact, detailed residential characteristics and socio-economic factors were collected at recruitment shortly after birth with a parental questionnaire. When the children were approximately 1, 2, 3, 4, 6, 10 and 15 years of age, similar questionnaires were sent to the parents, with a main focus on the children's symptoms related to allergic diseases and information on exposure factors. The study was approved by the Ethics Committee of the regional Medical Association (Nordrhein), Düsseldorf, Germany.

348 new-born infants were recruited between July 1994 and January 1999 in three different maternity awards in Wesel for the prospective study: The Influence of Life style factors on the development of the Immune System and Allergies in East and West Germany (LISA) north. The study population comprised 55% of all eligible children born in the three maternity wards. The study region Wesel is a rural area in Western Germany. Data on parental allergic diseases, pet contact, detailed residential characteristics and socio-economic factors were collected at recruitment shortly after birth with a parental questionnaire. When the children were approximately ½, 1, 1 ½, 2, 4, 6, 10 and 15 years of age, similar questionnaires were sent to the parents, with a main focus on the children's symptoms related to allergic diseases and information on exposure factors. The study was approved by the Ethics Committee of the regional Medical Association (Nordrhein), Düsseldorf, Germany.

For the GINIplus north and LISA north cohort, which have nearly identical study designs and outcome definitions, data were pooled.

Case/Control definition

The children have been followed up to collect information on atopic dermatitis using self-administered questionnaires as following. In GINIplus north, between 1- and 15-years parents were asked annually for physician-diagnosed eczema/atopic dermatitis within the last 12 months (one-year-prevalence, retrospective asked questions for ages 7 to 9 years and 11 to 14 years). In LISA north, between ½ and 2 years parents were asked half-annually for physician-diagnosed eczema/atopic dermatitis within the last 6 months. Between 4- and 15-years parents were asked annually for physician-diagnosed eczema/atopic dermatitis within the last 12 months (one-year-prevalence; retrospective asked questions for ages 5, 7 to 9 years and 11 to 14 years).

Cases of atopic dermatitis were defined as those who have ever had diagnosed with eczema by a doctor (at least one diagnosis at one observation time point, missing at other observation time points were allowed). Controls were defined as individuals who have never had atopic dermatitis (missing at any observation time point was not allowed).

### Genotyping and imputation

Genome-wide genotyping provided by the German Research Centre for Environmental Health at the Helmholtz Zentrum München in April 2019 was performed in 883 blood samples (collected in follow-up examination at age 10 years) using the Infinium Global Screening Array GSA v2 MD (GRCh37/hg19) with GenomeStudio Version 2.0 resulting in 712,189 variants.

In the pre-imputation quality control[61] variants on chromosome 0, insert/deletion variants, variants with low minor allele frequency (<0.01), and low call rates (<0.95) were excluded. After that, duplicated individuals, individuals with sex-mismatch, with low call rates (<0.95), with minimal heterozygosity (inbreeding coefficient 0.1), highly related individuals (identity-by-descent analysis with ld.tresh=0.2 and kin.tresh=0.1), as well as individuals belonging to non-European ancestry group (Tukey's rule based on the 1-10 eigenvectors from Principal Component Analysis), and violations of Hardy-Weinberg ($p<10^{-6}$) were removed. Finally, SNPs that deviate from Hardy-Weinberg equilibrium ($p<10^{-6}$) were removed. 792 individuals and 479,023 SNPs passed these quality control filters.

To find haplotype segments that are shared by study individuals and the HRC r1.1 2016 (GRCh37/hg19), we did a genotype imputation with minimac4 1.5.7 using the Michigan Imputation Server[62]. With the help of imputation, information for 792 individuals were calculated on 39,131,600 variants based on 469,304 SNPs that passed the quality control of Michigan Imputation Server.

In post-imputation processing multi-allelic markers and markers that deviate from Hardy-Weinberg equilibrium ($p<1x10^{-12}$ in controls) were excluded using vcftools.

### Statistical analysis

Genome-wide association analysis was performed with rvtests (version 2.1.0) for 219 AD cases and 363 controls while controlling for sex and 10 principal components.

### Acknowledgements and Funding

The authors thank all the families for their participation in the GINIplus study. Furthermore, we thank all members of the GINIplus Study Group for their excellent work. The GINIplus Study group consists of the following: Institute of Epidemiology, Helmholtz Zentrum München, German Research Center for Environmental Health, Neuherberg (Heinrich J, Brüske I, Schulz H, Flexeder C, Zeller C, Standl M, Schnappinger M, Ferland M, Thiering E, Tiesler C); Department of Pediatrics, Marien-Hospital, Wesel (Berdel D, von Berg A); Ludwig-Maximilians-University of Munich, Dr von Hauner Children's Hospital (Koletzko S); Child and Adolescent Medicine, University Hospital rechts der Isar of the Technical University Munich (Bauer CP, Hoffmann U); IUF- Environmental Health Research Institute, Düsseldorf (Schikowski T, Link E, Klümper C, Krämer U, Sugiri D).

The authors thank all the families for their participation in the LISA study. Furthermore, we thank all members of the LISA Study Group for their excellent work. The LISA Study group consists of the following: Helmholtz Zentrum München, German Research Center for Environmental Health, Institute of Epidemiology, Munich (Heinrich J, Schnappinger M, Brüske I, Ferland M, Schulz H, Zeller C, Standl M, Thiering E, Tiesler C, Flexeder C); Department of Pediatrics, Municipal Hospital "St. Georg", Leipzig (Borte M, Diez U, Dorn C, Braun E); Marien Hospital Wesel, Department of Pediatrics, Wesel (von Berg A, Berdel D, Stiers G, Maas B); Pediatric Practice, Bad Honnef (Schaaf B); Helmholtz Centre of Environmental Research – UFZ, Department of Environmental Immunology/Core Facility Studies, Leipzig (Lehmann I, Bauer M, Röder S, Schilde M, Nowak M, Herberth G , Müller J); Technical University Munich, Department of Pediatrics, Munich (Hoffmann U, Paschke M, Marra S); Clinical Research Group Molecular Dermatology, Department of Dermatology and Allergy, Technische Universität München (TUM), Munich (Ollert M, J. Grosch).

The GINIplus study was mainly supported for the first 3 years of the Federal Ministry for Education, Science, Research and Technology (interventional arm) and Helmholtz Zentrum Munich (former GSF) (observational arm). The 4 year, 6 year, 10 year and 15 year follow-up examinations of the GINIplus study were covered from the respective budgets of the 5 study centres (Helmholtz Zentrum Munich (former GSF), Research Institute at Marien-Hospital Wesel, LMU Munich, TU Munich and from 6 years onwards also from IUF - Leibniz Research-Institute for Environmental Medicine at the University of Düsseldorf) and a grant from the Federal Ministry for Environment (IUF Düsseldorf, FKZ 20462296). Further, the 15 year follow-up examination of the GINIplus study was supported by the Commission of the European Communities, the 7th Framework Program: MeDALL project, and as well by the companies Mead Johnson and Nestlé.

The LISA study was mainly supported by grants from the Federal Ministry for Education, Science, Research and Technology and in addition from Helmholtz Zentrum Munich (former GSF), Helmholtz Centre for Environmental Research - UFZ, Leipzig, Research Institute at Marien-Hospital Wesel, Pediatric Practice, Bad Honnef for the first 2 years. The 4 year, 6 year, 10 year and 15 year follow-up examinations of the LISA study were covered from the respective budgets of the involved partners (Helmholtz Zentrum Munich (former GSF), Helmholtz Centre for Environmental Research - UFZ, Leipzig, Research Institute at Marien-Hospital Wesel, Pediatric Practice, Bad Honnef, IUF – Leibniz-Research Institute for Environmental Medicine at the University of Düsseldorf) and in addition by a grant from the Federal Ministry for Environment (IUF Düsseldorf, FKZ 20462296). Further, the 15-year follow-up examination of the LISA study was supported by the Commission of the European Communities, the 7th Framework Program: MeDALL project. The research leading to the ESCAPE results has received funding from the European Community's Seventh Framework Program (FP7/2007-2011) under grant agreement number: 211250.

**GINI / LISAsouth**

<u>Recruitment and phenotype definition</u>

The influence of Life-style factors on the development of the Immune System and Allergies in East and West Germany (LISA) study is a population-based birth cohort study. A total of 3,094 healthy, full-term neonates were recruited between 1997 and 1999 in Munich, Leipzig, Wesel and Bad Honnef. The participants were not pre-selected based on family history of allergic diseases[63].

A total of 5,991 mothers and their newborns were recruited into the German infant study on the influence of Nutrition intervention PLUS environmental and genetic influences on allergy development (GINIplus) between September 1995 and June 1998 in Munich and Wesel. Infants with at least one allergic parent and/or sibling were allocated to the interventional study arm investigating the effect of different hydrolysed formulas for allergy prevention in the first year of life[64]. All children without a family history of allergic diseases and children whose parents did not give consent for the intervention were allocated to the non-interventional arm. Detailed descriptions of the LISA and GINIplus studies have been published elsewhere[63,64]. DNA was collected at the age 6 and 10 years. For both studies, approval by the local Ethics Committees and written consent from participant's families were obtained.

Case/Control definition

Information on ever having physician-diagnosed atopic eczema was collected using self-administered questionnaires completed by the parents. The questionnaires were completed at 6, 12, 18 and 24 months and 4, 5, 6, 10 years of age in the LISA study and 1, 2, 3, 4, 6 and 10 years in the GINIplus study, asking for each year of age since the previous follow-up. Cases were defined as subjects who reported having a diagnosis at any time point and controls were defined as those reporting no diagnosis at every time point, leading to 442 cases and 865 controls.

Genotyping and imputation

1,511 children from Munich from both studies were included (835 (55%) children from the GINIplus study and 676 (45%) children from the LISA study). 1423 individuals (835 from the GINIplus study and 588 from the LISA study) were analysed using the Affymetrix Human SNP Array 5.0 and 88 individuals from the LISA study were analysed using Affymetrix Human SNP Array 6.0. Genotypes were called using BRLMM-P algorithm (5.0), respectively BIRDSEED V2 algorithm (6.0). In each of the two data sets, criteria for exclusion of individuals were: a call rate below 95%, heterozygosity outside mean +/- 4sd, a failure of the sex check or a failure of the similarity quality control using MDS analysis based on IBS. Criteria for exclusion of variants were: a call rate below 95%, a MAF < 0.01 and a HWE p-value < 0.00001. Haplotype phasing and imputation in the HRC panel version1.1 was performed using the University of Michigan server[65]. Variants with an imputation quality score of $r^2 < 0.5$ or out of Hardy-Weinberg-Equilibrium p < 10-12 were excluded.

Statistical analysis

A score test implemented in RVTEST[41] (Version 20190205) was used to test the association between SNP allelic dosage and eczema status. Sex was included as a covariate.

Acknowledgements and funding

The authors thank all families for participation in the studies and the LISA and GINIplus study teams for their excellent work.

**Health Cohorts**

Recruitment

The Health cohorts are three consecutively sampled cohorts (Health 2006, Health 2008 and Health 2010). Health 2006 is a population-based epidemiological study of general health, diabetes and cardiovascular disease of individuals aged 18-74 years. The Health 2008 is an extension of the Health 2006 study, where a random sample of the general population aged 30 to 60 years from the same regional areas in Copenhagen County was drawn from the Civil Registration System. The Health 2010 study is a population-based epidemiologic study with primary focus on cardiovascular diseases, diabetes, asthma, and allergy based on a random sample of men and women aged between 18 and 69 years. All three studies were conducted at the Research Centre for Prevention and Health in Glostrup, Denmark. The studies were approved by the Ethics Committee of Capital Region of Denmark. For this study, the three cohorts were combined to one Health cohort.

Case/Control definition

Case-control status was defined by self-report from a health and lifestyle questionnaire filled in by the participants. Frequency of eczema was 5.6% among 5,667 participants.

Genotyping and imputation

Health cohort individuals were genotyped using the Illumina HumanOmniExpress-24v1-0_A and HumanOmniExpress-24v1-1_A chip. We used the GenCall software application to automatically cluster, call genotypes, and assign confidence scores. The resulting raw genotype data were subjected to standard quality control methods. The inclusion criteria included the exclusion of monomorphic SNPs with a MAF ≥ 0 %, call rate ≥ 98 %, p-value for Hardy-Weinberg equilibrium > 10^(-5). There were 632630 SNPs that met the previous QC criteria. All individuals with non-European ancestry were removed.

Remaining subjects were phased and imputed. The phasing was performed using Eagle autosomes and Shapit x-chromosome, and the phased haplotypes were then imputed to the Haplotype Reference Consortium (HRCr1.1, 2016) panel. The imputation was performed using the Michigan imputation server.

During the analysis, related individuals were removed leading to a percentage of individuals according to biological sex of 44.66 % males and 55.34 % females. Relatedness was studied using Identical By Descent method, filtering out pairs of individuals with a pi_hat coefficient larger than 0.1875.

Statistical analysis

Genome-wide association analysis was performed using RVtest (version 2.0.6) for 265 cases and 4524 controls with genetic and phenotypic data while including sex and 10 principal components as covariates.

Acknowledgements and Funding

Health 2006 was financially supported by grants from the Velux Foundation; The Danish Medical Research Council, Danish Agency for Science, Technology and Innovation; The Aase and Ejner Danielsens Foundation; ALK-Abello A/S, Hørsholm, Denmark, and Research Centre for Prevention and Health, the Capital Region of Denmark. Health 2008 was supported by the Timber Merchant Vilhelm Bang's Foundation, the Danish Heart Foundation (Grant number 07-10-R61-A1754-B838-22392F), and the Health Insurance Foundation (Helsefonden) (Grant number 2012B233). Further funding came from the Novo Nordisk Foundation (NNF15OC0015896).

**HUNT**

Recruitment

The Trøndelag Health Study (HUNT) is a population-based cohort study with information from >229,000 individuals collected during four time points over approximately 40 years (HUNT1 [1984-1986], HUNT2 [1995-1997] and HUNT3 [2006-2008] and HUNT4 [2017-2019])[66]. Every citizen aged 20 years and over of Trøndelag County in Norway was invited to take surveys documenting lifestyle, prevalence and incidence of somatic and mental illness and disease and health determinants. Participants are also linked to regional- and national health registers through the unique national identification number which enabled individuals with eczema to be identified by searching for eczema ICD codes. Participation in the HUNT Study is based on informed consent, and the study has been approved by the Data Inspectorate and the Regional Ethics Committee for Medical Research in Norway (REK 2014/144 and 2015/586).

Case/control definition

The eczema cases were either reporting yes on the question "Have you, or have you had, eczema on your hands (eczema)" in HUNT3. In addition, the following ICD-9, ICD-10 and ICPC2 codes were used to define eczema cases: 691.8, L20 and S87. Those who had not reported to have hand eczema (answered no) and did not have any of the listed codes above served as control.

Genotyping and imputation

Participants from HUNT2 and HUNT3 were genotyped using one of three different Illumina HumanCoreExome arrays (HumanCoreExome12 v1.0, HumanCoreExome12 v1.1 and UM HUNT Biobank v1.0). Genotype calling was performed with GenTrain v.2.0 in GenomeStudio v.2011.1 (Illumina). Samples with <99% genotype calls, with large chromosomal copy number variants, contamination >2.5% as estimated with BAF Regress (2), with genotypic and phenotypic sex discordance, and not of European ancestry were excluded, leaving 69,422 genotyped subjects. Genetic variants out of Hardy-Weinberg equilibrium (p-value <0.0001) were excluded. Imputation was performed on samples of recent European ancestry using Minimac3 (v2.0.1, http://genome.sph.umich.edu/wiki/Minimac3) from a merged reference panel constructed from the Haplotype Reference Consortium (HRC) panel (release version 1.1) and a local reference panel based on 2,202 whole-genome sequenced HUNT participants, resulting in 24.9 million SNPs (R2>0.3).

Statistical analysis

GWAS was run in SAIGE [67] version 0.35.8.3, using sex, birth year, genotyping batch and 4 ancestry principal components as covariates. Summary data was available for 22.3 million variants that were successfully analyzed.

Acknowledgements and funding

The Trøndelag Health Study (the HUNT Study) is a collaboration between HUNT Research Centre (Faculty of Medicine and Health Sciences, Norwegian University of Science and Technology NTNU), Trøndelag County Council, Central Norway Regional Health Authority, and the Norwegian Institute of Public Health. The genetic investigations of the HUNT Study, is a collaboration between researchers from the K.G. Jebsen center for genetic epidemiology and University of Michigan Medical School and the University of Michigan School of Public Health. The K.G. Jebsen Center for Genetic Epidemiology is financed by Stiftelsen Kristian Gerhard Jebsen; Faculty of Medicine and Health Sciences, NTNU, Norway. BMB, LT, ML and KH work in a research unit funded by Stiftelsen Kristian Gerhard Jebsen; Faculty of Medicine and Health Sciences, NTNU, Norwegian University of Science and Technology; The Liaison Committee for education, research and innovation in Central Norway; the Joint Research Committee between St. Olavs Hospital (Trondheim, Norway) and the Faculty of Medicine and Health Sciences, NTNU, Norwegian University of Science and Technology. ML is supported by grants from the Liaison Committee for education, research and innovation in Central Norway and by the Joint Research Committee between St. Olavs Hospital and the Faculty of Medicine and Health Sciences.

**INfancia y Medio Ambiente Project (INMA)**

Recruitment and phenotype definition

Population-based birth cohorts were established as part of the INMA – INfancia y Medio Ambiente [Environment and Childhood] Project in several regions of Spain following a common protocol. Further details have been previously described elsewhere[19]. Children from the subcohorts of INMA Sabadell, Valencia and Menorca were included in the present study. Atopic eczema (Sabadell and Valencia) and doctor atopic eczema (Menorca) was assessed by questionnaire at the ages of 1, 2 and 4y. Atopic eczema cases were those children that had had eczema at least in one of the three visits. Control children were those that had never had eczema.

Genotyping, imputation and statistical analysis

Genome-wide genotyping was performed using the HumanOmni1-Quad Beadchip (Illumina) at CEGEN. Genetic variants were filtered for SNP call rate>95%, MAF>1% and HWE $P$ value>$1.1x10^{-6}$. Haplotype phasing and imputation in the HRC panel version1.1 was performed using the University of Michigan server[20]. Variants with an imputation quality score of $r^2 < 0.5$ or out of Hardy-Weinberg-Equilibrium $P < 10^{-12}$ were excluded. A score test implemented in RVTESTS was used to test the association between SNP allelic dosage and eczema status. Sex and the first two principle components were included as covariates. Further details have been described elsewhere[19].

Acknowledgements and funding

This study was funded by grants from instituto de Salud Carlos III (CB06/02/0041, G03/176, FIS PI041436, PI081151, PI041705, PI061756, PI091958, and PS09/00432, FIS-FEDER 03/1615, 04/1509, 04/1112, 04/1931 , 05/1079, 05/1052, 06/1213, 07/0314, 09/02647, 11/01007, 11/02591, 11/02038, 13/1944, 13/2032 and CP11/0178), Spanish Ministry of Science and innovation (SAF2008-00357), European Commission (ENGAGE project and grant agreement HEALTH-F4-2007-201413, HEALTH.2010.2.4.5-1, FP7-ENV-2011 cod 282957), Fundació La Marató de TV3, Generalitat de Catalunya-CIRIT 1999SGR 00241 and Conselleria de Sanitat Generalitat Valenciana. Part of the DNA extractions and genotyping was performed at the Spanish National Genotyping Centre (CEGEN-Barcelona). The authors are grateful to Silvia Fochs, Anna Sànchez, Maribel López, Nuria Pey, Muriel Ferrer, Amparo Quiles, Sandra Pérez, Gemma León, Elena Romero, Maria Andreu, Nati Galiana, Maria Dolores Climent, Amparo Cases and Cristina Capo for their assistance in contacting the families and administering the questionnaires. A full roster of the INMA Project investigators can be found at http://www.proyectoinma.org/presentacion-inma/listado-investigadores/en_listado investigadores.html. The authors would particularly like to thank all the participants for their generous collaboration.

**The Isle of Wight birth cohort**

Recruitment

The Isle of Wight birth cohort (IOWBC) is an unselected birth cohort composed of mainly Caucasian descent (98%). After exclusion of adoptions and prenatal deaths, 1456 children born between January 1, 1989 and February 28, 1990 on the Isle of Wight, UK, were consented and included in the cohort. They were followed up at 1/2, 4, 10, and 18 years of age. Details about the birth cohort have been described in detail elsewhere[68]. At each follow-up, participants were evaluated for manifestations of allergic disease and administered detailed questionnaires including study specific questions. Blood samples were collected at ages 10 and 18 years of age. Atopic dermatitis (AD) was defined as chronic or chronically relapsing itchy dermatitis lasting more than 6 weeks with characteristic morphology and distribution[69,70], following Hanifin and Rajka criteria[10]. Ethical approval was obtained from the local Research Ethics Committee (06/Q1701/34). The informed consent was written for in-person visits. For participants joined by phone, the consent was documented on the face-to-face consent form. The name of the person giving consent and the name and signature of the person taking the form were documented.

Case/Control definition

Detailed questionnaires, resembling the questionnaire of the International Study of Asthma and Allergy in Childhood (ISAAC), were given at age of 4 years (before the ISAAC questionnaire was published). Eczema status (yes/no) included in the study was at this age based on n=979 subjects including n=110 cases.

Genotyping, imputation, and quality control

DNA from the blood samples were genotyped at Oxford Genomics using the Illumina InfiniumOmni2.5-8v1.3b37-based manifest. Using the Sanger Imputation Service (https://imputation.sanger.ac.uk/), genotypes were pre-phased using EAGLE2 and imputed using PBWT with the Haplotype Reference Consortium reference panel. We directly genotyped 2,372,784 SNPs. After quality control was applied, 1,641,983 SNPs of n=1067 subjects was directly genotyped. Imputation using the Sanger server generated in total 16,870,314 SNPs. After excluding SNPs with minor allele frequency (MAF) <0.01, SNPs violating Hardy-Weinberg equilibrium with p-value $>1x10^{-6}$, and duplicated SNPs, in total, 7,236,417 SNPs were concluded. After excluding subjects with high genetic heterozygosity, subjects who failed sex check, and subjects showing a third-degree relation, n=979 subjects were concluded. After all these processes, 7,236,417 SNPs of n=979 subjects with INFO >0.8 were retained for further analysis.
A principle component analysis (PCA) was performed to explore ethnicity in the IOWBC against the HapMap reference. Three major populations were considered, European, Asian, and African. Only SNPs common in HapMap and IOW were used for the PCA.

Statistical analysis

Logistic regressions in PLINK 2.0 were applied to analyze the data with sex and 20 principal components included in the analyses. The command to perform the analysis in plink is the format of plink2 --bfile PhenotypeData --covar CovariateData --covar-variance-standardize --logistic no-x-sex --chr 1 --adjust --out IoW_EAGLE_GWAS3_2021-01-25_chr1_Eczema.

Acknowledgements and Funding

The study was supported by NIAID/NIH with funding number R01AI121226 and by Isle of Wight Health Authority, United Kingdom. Genotype data was supported by the Medical Research Council UK as part of UNICORN (Unified Cohorts Research Network): Disaggregating asthma project (MR/S025340/1) and the Wellcome Trust strategic award, Breathing Together (108818/15/Z). We would like to acknowledge the contribution of all the staff at The David Hide Asthma and Allergy Research Centre in the assessments of 1989/90 Isle of Wight birth cohort. We are greatly thankful to the help of the study subjects and their families who participated in the study. We are also very thankful to the High Performance Computing facility at the University of Memphis to help us efficiently carry out the data analyses.

**Manchester Asthma and Allergy Study (MAAS)**

Recruitment and phenotype definition

The Manchester Asthma and Allergy Study is an unselected (i.e. population-based), birth cohort study. The study was approved by the Local Research Ethics Committee. Informed consent was obtained from all parents. Further details of study screening and recruitment have been described elsewhere[19].

Genotyping, imputation and statistical analysis

DNA samples were genotyping on an Illumina 610 quad chip. Quality control criteria for SNPs included a 95% call rate, HWE > $5.9x10^{-7}$, minor allele frequency > 0.005. Further detailed have been described elsewhere[19]. Haplotype phasing and imputation in the HRC panel version1.1 was performed using the University of Michigan server[20]. Variants with an imputation quality score of $r^2 < 0.5$ or out of Hardy-Weinberg-Equilibrium P < $10^{-12}$ were excluded. A score test implemented in RVTESTS was used to test the association between SNP allelic dosage and eczema status. Sex and the first two principle components were included as covariates.

Acknowledgements and funding

We greatly appreciate the commitment they have given to the project. We would also like to acknowledge the hard work and dedication of the study team (post-doctoral scientists, research fellows, nurses, physiologists, technicians and clerical staff). This report includes independent research supported by National Institute for Health Research Respiratory Clinical Research Facility at Manchester University NHS Foundation Trust (Wythenshawe). The views expressed in this publication are those of the author(s) and not necessarily those of the NHS, the National Institute for Health Research or the Department of Health. MAAS was supported by the Asthma UK Grants No 301 (1995-1998), No 362 (1998-2001), No 01/012 (2001-2004), No 04/014 (2004-2007), the BMA James Trust and Medical Research Council, UK (G0601361) and The Moulton Charitable Foundation (2004-current); the Medical Research Council (MRC) Grants G0601361, MR/K002449/1 and MR/L012693/1, and Angela Simpson is supported by the NIHR Manchester Biomedical Research Centre. The authors would like to acknowledge the North West Lung Centre Charity for supporting this project.

**Multicentre Allergy Study / Heinz Nixdorf RECALL (MAS / HNR)**

Recruitment and phenotype definition

The Multicentre Allergy Study (MAS) is a German birth cohort which has been described in detail previously[71,72]. Eczema was defined based on a parental report of a doctor's diagnosis of eczema up to the age of 13 years. Controls were unrelated individuals from the population-based Heinz Nixdorf RECALL (HNR) study[73]. Further details have been described elsewhere[19].

Genotyping, imputation, and statistical analysis

MAS samples were genotyped with the Illumina Human610 array, HNR samples with the Illumina Human550v3 array. Further details of quality control measures applied have been described elsewhere[19]. Haplotype phasing and imputation in the HRC panel version1.1 was performed using the University of Michigan server[20].7 Variants with an imputation quality score of $r^2 < 0.5$ or out of Hardy Weinberg-Equilibrium $P < 10^{-12}$ were excluded. A score test implemented in RVTESTS was used to test the association between SNP allelic dosage and eczema status. Sex and the first two principle components were included as covariates.

Acknowledgements and funding

We are grateful to all children and parents who participated in this study. Collaborators of the MAS group: R. Bergmann (Berlin, Germany) V. Wahn, M. Groeger (Dusseldorf, Germany); J. Forster, U. Tacke (Freiburg, Germany); C-P. Bauer (Gaisach, Germany); F. Zepp, I. Bieber (Mainz, Germany). This study was supported by the German Ministry of Education and Research (BMBF) through grants number 01 EE9406 and 01 GC9702/0. Genotyping of the MAS cases was supported through the EU Framework 6 integrated Project "GABRIEL ". The collection of probands in the Heinz Nixdorf RECALL Study (HNR) was supported by the Heinz Nixdorf Foundation. The genotyping of HNR probands was financed through a grant of the German Ministry of Education and Science (BMBF). MMN is member of the Excellence Cluster ImmunoSensation2 which is funded by the DFG under Germany's Excellence Strategy –EXC2151 (project number 390873048).

**MoBa**

Study population

The Norwegian Mother, Father and Child Cohort Study (MoBa) is an open-ended cohort study that recruited pregnant women in Norway from 1999 to 2008. Approximately 114,500 children, 95,200 mothers, and 75,000 fathers of predominantly Norwegian ancestry were enrolled in the study from 50 hospitals all across Norway[74]. The project Better Health by Harvesting Biobanks (HARVEST) randomly selected 11,490 umbilical cord blood DNA samples from the biobank of this study for family triad genotyping, excluding samples matching any of the following criteria: (1) stillborn, (2) deceased, (3) twins, (4) non-existing data at the Norwegian Medical Birth Registry, (5) missing anthropometric measurements at birth in Medical Birth Registry, (6) pregnancies where the mother did not answer the first questionnaire (as a proxy for higher dropout rate), and (7) missing parental DNA samples. In 2016, HARVEST randomly selected a second set of 8,900 triads using the same criteria. The same year NORMENT selected 5,910 triads with the same selection criteria as HARVEST, and extended this with 3,209 triads in 2018. Additionally, further 1,062 ADHD cases among the children were genotyped, and a study genotyped 5,834 randomly selected parents. Given the study design (parent-offspring study), mothers and fathers were analysed jointly, while children were analysed on their own. In this current study, only data from children was included in the analysis.

Genotyping, phasing and imputation

Genotyping of the samples was performed in seven different batches on different Illumina platforms (HumanCoreExome-12 v.1.1 and HumanCoreExome-24 v.1.0, Illumina's Global Screening Array v.1.0, InfiniumOmniExpress-24v1.2 and HumanOmniExpress-24-v1.0). The Genome Reference Consortium Human Build 37 (GRCh37) reference genome was used for all annotations.

Genotypes were called in Illumina GenomeStudio (v.2011.1 and v.2.0.3). Cluster positions were identified from samples with call rate ≥ 0.98 and GenCall score ≥ 0.15. We excluded variants with low call rates, signal intensity, quality scores, and deviation from Hardy-Weinberg equilibrium (HWE) based on the following QC parameters: call rate < 98 %, cluster separation < 0.4, 10% GC-score < 0.3, AA T Dev > 0.025, HWE p-value < $1 \times 10{-}6$. Samples were excluded based on call rate < 98 % and heterozygosity excess > 4 SD. Study participants with recent white Nordic ancestry were included after merging with ancestry reference samples from the HapMap project (ver. 3).

Pre-phasing was conducted locally using Shapeit v2.790[75]. Imputation was performed at the Sanger Imputation Server with positional Burrows-Wheeler transform and HRC version 1.1 as reference panel[38].

Phenotype

The data used is v12 of the quality-assured data files released by MoBa. Children with atopic dermatitis were proxy identified; a legal representative answered a questionnaire when the child was 18 months or 3 years of age. Cases were defined as children whose legal representative reported atopic eczema in any of the two questionnaires, and controls as children whose legal representative answered at least one questionnaire and reported no atopic dermatitis. The response was based on the following questions: "Does your child have or has he/she had any of the following health problems?" and "Has your child suffered any long-term illness or health problems since the age of 18 months?". A positive answer to these questions classified the child as a case. We identified 5,963 child cases and 16,750 child controls.

Association analysis

Prior to analysis, we used a greedy algorithm to exclude one individual from each set of related subjects (within children groups separately), where sets were defined as two individuals with a kinship coefficient >0.125. Imputed genetic variants with minor allele frequency < 0.001, Hardy-Weinberg equilibrium *P*-value< $10^{-12}$ and imputation INFO score> 0.5. All analyses were performed using rvtest (version: 20190205), using genotype dosage as input and including sex and the 10 first principal components as covariates.

Ethics statement

Informed consent was obtained from all study participants. The administrative board of the Norwegian Mother, Father and Child Cohort Study led by the Norwegian Institute of Public Health approved the study protocol. The MoBa cohort is currently regulated by the Norwegian Health Registry Act. The study was approved by The Regional Committee for Medical Research Ethics (#2015/2425).

Funding statement

This work was supported by grants (B.J.) from The Swedish Research Council, Stockholm, Sweden (2015-02559), The Research Council of Norway, Oslo, Norway (FRIMEDBIO #547711, #273291), March of Dimes (#21-FY16-121) and (to P.R.N.) from the European Research Council (AdG SELECTionPREDISPOSED #293574), the Bergen Research Foundation ("Utilizing the Mother and Child Cohort and the Medical Birth Registry for Better Health"), Stiftelsen Kristian Gerhard Jebsen (Translational Medical Center), the University of Bergen, the Research Council of Norway (FRIPRO grant #240413), the Western Norway Regional Health Authority (Strategic Fund "Personalized Medicine for Children and Adults"), the Novo Nordisk Foundation (grant #54741), and the Norwegian Diabetes Association. This work was partly supported by the Research Council of Norway through its Centres of Excellence funding scheme (#262700, #223273), Better Health by Harvesting Biobanks (#229624) and The Norwegian Mother, Father and Child Cohort Study is supported by the Norwegian Ministry of Health and Care Services and the Ministry of Education and Research, NIH/NIEHS (contract no N01-ES-75558), NIH/NINDS (grant no.1 UO1 NS 047537-01 and grant no.2 UO1 NS 047537-06A1). We are grateful to all the families in Norway who are taking part in this ongoing cohort study. All analyses were performed using digital labs in HUNT Cloud at the Norwegian University of Science and Technology, Trondheim, Norway.

Acknowledgements

The Norwegian Mother, Father and Child Cohort Study is supported by the Norwegian Ministry of Health and Care Services and the Ministry of Education and Research. We are grateful to all the participating families in Norway who take part in this on-going cohort study.

We thank the Norwegian Institute of Public Health (NIPH) for generating high-quality genomic data. This research is part of the HARVEST collaboration, supported by the Research Council of Norway (#229624). We also thank the NORMENT Centre for providing genotype data, funded by the Research Council of Norway (#223273), South East Norway Health Authority and KG Jebsen Stiftelsen. We further thank the Center for Diabetes Research, the University of Bergen for providing genotype data and performing quality control and imputation of the data funded by the ERC AdG project SELECTionPREDISPOSED, Stiftelsen Kristian Gerhard Jebsen, Trond Mohn Foundation, the Research Council of Norway, the Novo Nordisk Foundation, the University of Bergen, and the Western Norway health Authorities (Helse Vest). All analyses were performed using digital labs in HUNT Cloud at the Norwegian University of Science and Technology, Trondheim, Norway.

**National Children's Research Centre - Atopic Dermatitis Collection (NCRC - ADC)**

Recruitment

631 atopic dermatitis cases were recruited from secondary/tertiary clinics in Our Lady's Children's Hospital Crumlin, Dublin and clinics in Glasgow, Dundee and Edinburgh.

Case/Control definition

All patients all had early onset disease (<2 years; mean age at recruitment 2.8 years) with a mean atopic dermatitis severity score (Nottingham Eczema Severity Score; NESS) of 10.23 (SD 3.11). The Irish controls comprised 996 subjects with no history of atopic dermatitis from a collection of 1237 blood donor volunteers of the Trinity Biobank.

Genotyping and imputation

Genome-wide genotyping was performed on bar-coded LIMS (Laboratory Information Management System) tracked samples using the Illumina Human 610-Quad BeadChip (Illumina, San Diego) for AD cases and Affymetrix genome-wide SNP array 6.0 (Affymetrix, Santa Clara) for Trinity controls.

BeadChips were processed within an automated BeadLab as per the respective manufacturer's instructions. Samples were subject to strict quality control criteria including assessment of concentration, fragmentation and response to PCR. A total of 20 µl of DNA aliquoted to a concentration of 50 ng/µl was used for each array. Replication phase genotyping was performed using matrix-assisted laser desorption/ionization time-of flight (MALDI-TOF) mass spectrometry (http://www.sequenom.com), or Applied Biosystems TaqMan probes (http://www.appliedbiosystems.com/).

Prior to imputation we excluded samples with extensive missing data rate (>5%), excess of heterozygosity or homozygosity and ambiguous sex. We examined IBS and excluded close related samples with PI_HAT>0.1875 (halfway between expected IBD for third- and second degree relatives) as well as outliers of unusual ancestry by MDS analysis. SNPs with low genotyping rate (<95%), low minor allele frequency (<1%), strong deviation from Hardy-Weinberg equilibrium (p<10-8) and differential call rate between cases and controls were excluded. Cases and controls with high quality SNPs of the overlapping SNP-set were matched and carried forward to imputation. Pre-phasing was carried out with SHAPEIT[24] and imputation with IMPUTE[76] using phase I 1000 Genomes reference panel (integrated variant set of all populations, release March 2012). Post imputation SNPs with low imputation quality (info score<0.4), call rate<95%, deviation from Hardy-Weinberg equilibrium (p<10-8) or minor allele frequency<5% were excluded. 572 cases and 1797 controls with 5,262,635 overlapping SNPs were carried forward to analysis.

Statistical Analysis

Genome-wide association analysis was carried out using SNPTEST[26] using a frequentist approach with allele dosages (option -method expected) to account for imputation uncertainty.

Acknowledgements and Funding

The NCRC case Collection is supported by the National Children's Research Centre, Dublin and by the Wellcome Trust [reference: 090066/B/09/Z, 092530/Z/10/Z]

**Northern Finland Birth Cohort 1966 (NFBC66)**

Recruitment and phenotype definition

The Northern Finland Birth Cohort 1966 is a prospective follow-up study of children from the two northernmost provinces of Finland[77].The study was approved by the ethics committees in Oulu (Finland) and Oxford (UK) universities in accordance with the Declaration of Helsinki. Further recruitment details have been described elsewhere[19]. For the purpose of this meta-analysis, we included data from the following questions:

1. Have you had eczema (infantile, atopic or allergic)?

2. If yes, have you ever been treated by a doctor

Individuals who answered yes to both questions were defined as cases (1,200). Individuals that answered no to the first question were defined as controls (2,270).

Genotyping, imputation and statistical analysis

Genotyping was completed at the Broad Institute Biological Sample Repository in participants with available DNA using Illumina HumanCNV370DUO Analysis BeadChip array. Haplotype phasing and imputation in the HRC panel version1.1 was performed using the University of Michigan server[20]. Variants with an imputation quality score of $r^2 < 0.5$ or out of Hardy-Weinberg Equilibrium $P < 10^{-12}$ were excluded. A score test implemented in RVTESTS was used to test the association between SNP allelic dosage and eczema status. Sex and the first three principle components were included as covariates.

Acknowledgements and funding

We thank the late Professor Paula Rantakallio (launch of NFBC1966), and Ms Outi Tornwall and Ms Minttu Jussila (DNA biobanking). The authors would like to acknowledge the contribution of the late Academian of Science Leena Peltonen. NFBC1966 received financial support from the Academy of Finland (project grants 104781, 120315, 129269, 1114194, 24300796, Center of Excellence in Complex Disease Genetics and SALVE), Oulu University Hospital, Finland, Biocenter, University of Oulu, Finland 75617, 24002054, University of Oulu, Finland (Grant no. 24000692 and 24500283:Well-being and health: Research in the Northern Finland Birth Cohorts 1966 and 1986, Phenotypic and Genomic analyses). NIH/NHLBI NHLBI grant 5R01HL087679-02 through the STAMPEED program (1RL1MH083268-01), NHLBI Consortium for Neuropsychiatric Phenomics Co-ordinating Center (1-R01-HL087679-01) and. NIH/NIMH (5R01MH63706:02), USA. ENGAGE project and grant agreement HEALTH-F4-2007-201413. Medical Research Council (grant no. G1002319).The DNA extractions, sample quality controls, biobank up-keeping and aliquotting was supported financially by the Academy of Finland and Biocentrum Helsinki.

**Netherlands Twin Register (NTR)**

Recruitment

In the Young Netherlands Twin Register (YNTR), twins are followed from birth onwards. Around every two to three years, a survey is sent out inquiring about physical and mental health. At ages 1, 2, 3, 5, 7, 10 and 12 years, the surveys are completed by the parents and/or teachers, from age 14 onward, twins and their non-twin siblings are asked to complete the surveys by themselves.

Case/Control definition

Data on eczema were available from an YNTR survey that is sent out to the parents of twins at age 5 years[78]. Parents were asked to indicate for each child separately whether a doctor had ever diagnosed the child with eczema. A similar question concerned doctor diagnosed baby eczema.

Children were considered cases if their parents answered yes to any of the two questions and controls if they answered no to both questions. For MZ pairs, if both twins were cases or controls, this defined the phenotype. Discordant pairs received a case phenotype. MZ pairs were included as one case or control. For dizygotic twin pairs, both twins were included in the analysis (taking their relatedness into account in the analyses). After exclusion of subjects from non-Western European ancestry, data from 1,466 individuals were available for analysis.

In recent years, blood and buccal DNA samples were collected for various projects within the NTR [79,80]. From these samples, high molecular weight genomic DNA was extracted using the QIAamp DNA Blood Maxi kit (QIAGEN; Dusseldorf, Germany) following the manufacturers' protocol; DNA from buccal epithelial cells was extracted following a previous protocol[81]. Genotyping for the 1466 subjects was done on the Affymetrix 6.0 array and genotype calls were made with the Affymetrix GCT 4.0 software[82].

Genotypes were aligned to the GIANT 1000 Phase I Integrated release version 3 All panel. SNPs that were not mapped at all, SNPs that had ambiguous locations, and SNPs that did not have matching - or strand opposite alleles were removed. SNPs were also removed if they still had mismatching alleles with this imputation reference set, if the allele frequencies differed more than 0.20 with the reference set, if the MAF was < 1%, if the HWE p-value was < 0.00001 or if the call rate was <95%.

Lastly, SNPs with C/G and A/T allele combinations were removed if the MAF was between 0.35 and

0.50 to avoid wrong strand alignment for these SNPs. Samples were excluded from the data if their expected sex did not match their genotyped sex, if the genotype missing rate was above 10%, if the sample did not match the expected IBD sharing between relatives, or if the Plink F inbreeding value was either > 0.10 or < -0.10 (heterozygosity). Phasing was done in chromosomal chunks with the Mach program[14]. Then imputation of the reference panel haplotypes was performed with the minimac program[25] following the GIANT imputation protocols (http://genome.sph.umich.edu/wiki/Minimac:_1000_Genomes_Imputation_Cookbook ).

Statistical Analysis

A logistic regression was performed in Plink[15] in which eczema status was regressed on genome-wide SNP data, sex of the child and 5 additional covariates (3 Dutch PCs, one covariate for array effects and one for blood/ buccal) The analysis was corrected for familial clustering with the --family option in Plink.

Acknowledgements and Funding

Genetic influences on stability and change in psychopathology from childhood to young adulthood (ZonMW 91210020); Biobanking and Biomolecular Resources Research Infrastructure (BBMRI -NL, 184.021.007); VU University's Institute for Health and Care Research (EMGO+ ); Genetics of Mental Illness (European Research Council ERC-230374); Community's Seventh Framework Program (FP7/2007-2013) ENGAGE (HEALTH-F4-2007-201413); the Avera Institute, Sioux Falls, South Dakota (USA) and Grand Opportunity grants 1RC2 MH089951 and 1RC2 MH089995).

**PIAMA**

Recruitment

This study was performed within the PIAMA (Prevention and Incidence of Asthma and Mite Allergy) birth cohort study. Details of the cohort have been published previously[83]. The recruitment took place in 1996-1997 through prenatal clinics. In total, 10,232 pregnant women completed a validated screening questionnaire at their prenatal health care clinic (n=52 clinics). Based on this screening, 7,862 women were invited to participate, of whom 4,146 women agreed and gave informed consent. The study started with 3,963 newborns. Questionnaire based follow-up of the children took place at 3 months of age, annually from 1 to 8 years of age, and at 12, 14, and 16 or 17 years of age. The Medical Ethical Committees of the participating institutes approved the study, and all participants gave written informed consent.

Phenotype definition

Questionnaire information on eczema was obtained at ages 3m, 1y, 2y, 3y, 4y, 5y, 6y, 7y, 8y, 12y, 14y, 16/17y. For each time point case was defined as a positive response to one or more of these three questions: 1. has your child ever had atopic dermatitis? 2. Did a doctor ever diagnose atopic dermatitis in your child? And did you child have atopic dermatitis during the past 12 months? Eczema ever case was defined as any eczema from 3m to 16/17y. Since there were many missing phenotypes at some time points, the control was defined as 1) all time points from 2m to 16/17y were no eczema, or 2) 80% time points were no eczema when there were missing values from 2m to 16/17y. Control didn't include 3m and 1yr because at 3 m and 1 y the eczema definition does not include doctors' diagnosis and also not if the child had eczema in the last 12 months.

Genotyping, quality control and imputation

Genome-wide genotyping was performed in four phases. Quality control of each phase was performed and then the data were merged together. We removed individuals that were 1) sex-mismatch, 2) heterozygous outliers (deviate ± 3SD from the sample heterozygosity mean), 3) duplicated and highly related (IBD score <0.1875), 4) ethnic outliers from PCA plot. The first phase was performed within the framework of the GABRIEL Consortium using an Illumina Human 610k quad array37. Genotypes were available from 363 children after quality control. A second group of children were genotyped with an Illumina HumanOmniExpress array, 272 individuals were available after quality control. A third group of children was genotyped with the Illumina Human Omni Express Exome Array, 1333 individuals were available after quality control. A final group of children was genotyped with Illumina Infinium Global Screening Arrary, 107 individuals were available after quality control. This resulted in 2075 individuals that passed the quality control.

SNPs were harmonized by base pair position annotated to genome build 37. In total 2075 individuals remained after quality control, and data from four platforms were merged together. Before imputation we removed SNPs with call rate<0.95, MAF<1% and HWE<0.000001, which resulted in 83558 SNPs in total. Then, imputation was performed using Michigan server with a reference panel of HRC.r1. SNPs of high quality (Imputation quality score Rsq >0.5), MAF>0.001 and HWE <1E-12 were used for further analysis (N=7153756). The analysis was restricted to Caucasian individuals with genotype data and phenotype information (n=1919).

Statistical analysis

The SNP association analysis was carried out use rvtests (version 2.1.0) for 980 cases and 939 controls with genetic and phenotype data. Logistic regression was performed using dosages, with the correction of sex and first 3 principal components.

Acknowledgements

The PIAMA birth cohort is a collaboration of the Institute for Risk Assessment Sciences, University of Utrecht; Centre for Prevention and Health Services Research, National Institute for Public Health and the Environment, Bilthoven; and the Departments of Epidemiology and Pediatric Pulmonology and Pediatric Allergology, University Medical Center Groningen. The study team gratefully acknowledges all participants of the PIAMA birth cohort study, and coworkers who helped to conduct the medical examinations, field work and data management. For the generation of the genotype dataset, we especially acknowledge Dr F.N. Dijk and M. van Breugel.

Funding

The PIAMA study was funded by grants from the Netherlands Lung Foundation, Zon-MW Netherlands Organization for Health Research and Development, the Stichting Astma Bestrijding and the Ministry of the Environment. Genotyping was funded in part by grants from the European Commission (Gabriel, contract number 018996) and a grant from BBMRI-NL (CP29).

**The Raine Study**

Recruitment and phenotype definition

The Raine Study[84] is a prospective pregnancy cohort where 2900 were recruited from King Edward Memorial Hospital between 1989 and 1991. Further details have been described elsewhere[19]. Parents were asked the following questions at The Raine Study Gen2-5 year, Gen2-14 year and Gen2-17 year follow-ups:

1. Has anyone ever told you that your child has eczema? If yes, who told you your child has eczema?

We defined cases as the children of parents who answered ‘Yes and was diagnosed by a doctor or paediatrician at any one of the follow-ups. Controls were defined as children of parents who answered no at all 3 follow-ups.

Genotyping, imputation and statistical analysis

Individuals were genotyped using the e Illumina Human660W Quad Array at the Centre for Applied Genomics (Toronto, Ontario, Canada). Further details have been described elsewhere[19]. Haplotype phasing and imputation in the HRC panel version1.1 was performed using the University of Michigan server[20]. Variants with an imputation quality score of $r^2 < 0.5$ or out of Hardy-Weinberg Equilibrium $P < 10^{-12}$ were excluded. A score test implemented in RVTESTS was used to test the association between

SNP allelic dosage and eczema status. Sex and the first two principle components were included as covariates.

Acknowledgements and funding

This study was supported by the National Health and Medical Research Council of Australia [grant numbers 403981 and 003209] and the Canadian Institutes of Health Research [grant number MOP-82893]. The authors are grateful to the Raine Study participants and their families, and to the Raine Study research staff for cohort coordination and data collection. The authors gratefully acknowledge the NH&MRC for their long-term contribution to funding the study over the last 30 years and also the following institutions for providing funding for Core Management of the Raine Study: The University of Western Australia (UWA), Curtin University, Raine Medical Research Foundation, Telethon Kids Institute, Women and infants Research Foundation (King Edward Memorial Hospital), Murdoch University, The University of Notre Dame (Australia) and Edith Cowan University. The authors gratefully acknowledge the assistance of the Western Australian DNA Bank (National Health and Medical Research Council of Australia National Enabling Facility). This work was supported by resources provided by the Pawsey Supercomputing Centre with funding from the Australian Government and the Government of Western Australia.

**The Study of African Americans, Asthma, Genes, and Environments (SAGE)**

Recruitment details

The Study of African Americans, Asthma, Genes, and Environments (SAGE) is a cross sectional case control study of asthma in African American children and young adults from the San Francisco Bay Area, CA. Participants were eligible if they were 8 to 21 years-old, self-identified as African Americans, and had four African American grandparents.

Asthma was diagnosed by the attending physician and reports of symptoms or use of asthma controller or rescue medication in the two years preceding enrolment. Healthy controls were eligible if they had no reported history of asthma, other lung diseases or chronic illnesses as well as asthma symptoms of coughing, wheezing or shortness of breath[34–36] (1,2,3). Active smokers were excluded[34–36] (1,2,3).

The study has been approved by the Human Research Protection Program Institutional Review Board of the University of California, San Francisco (San Francisco, USA). The University of California, San Francisco (UCSF) institutional review board (IRB) approved the SAGE II protocols (UCSF IRB No. 10-02877). All participants/parents provided written assent/consent, respectively.

Case/ control definition

For the current study, case status was defined based on an affirmative response to the following question of the survey: "Has a doctor ever diagnosed the child with eczema or atopic dermatitis?". Controls were defined as individuals with absence of eczema based on a negative answer to the question. Cases were individuals that had an affirmative response. Eczema status was defined regardless of the asthma status.

Details of genotyping and imputation

Genotype data was obtained with the Affymetrix Axiom® LAT1 array (World Array 4, Affymetrix, Santa Clara, CA). Quality control was similar to the procedures described for GALA II and has been described elsewhere[37] (4). Imputation was performed at the Michigan Imputation Server using the reference panel of the Haplotype Reference Consortium 1.1[20] (5). Pre-phasing was performed using SHAPEIT v2.r790[39] (6) and imputation was conducted with Minimac3[40].

Details on statistical analysis

Association between eczema and genetic variants was tested through logistic regression models with covariate adjustment for sex and the two genotype principal components by using RVTESTS[41].

Acknowledgements

The authors acknowledge the families and patients for their participation and thank the numerous researchers, health care providers, and community clinics for their support and participation in the GALA II and SAGE studies. In particular, the authors thank the study coordinator Sandra Salazar; the principal investigators involved in the recruitment: Kelley Meade, Harold J. Farber, Pedro C. Avila, Denise Serebrisky, Shannon M. Thyne, Emerita Brigino-Buenaventura, William Rodriguez-Cintron, Saunak Sen, Rajesh Kumar, Michael Lenoir, and Luisa N. Borrell; and the recruiters who obtained the data: Duanny Alva, Gaby Ayala-Rodriguez, Lisa Caine, Elizabeth Castellanos, Jaime Colon, Denise DeJesus, Blanca Lopez, Brenda Lopez, Louis Martos, Vivian Medina, Juana Olivo, Mario Peralta, Esther Pomares, Jihan Quraishi, Johanna Rodriguez, Shahdad Saeedi, Dean Soto, and Ana Taveras.

The Genes-environments and Admixture in Latino Americans (GALA II) study and the Study of African Americans, Asthma, Genes and Environments (SAGE) were supported by the Sandler Family Foundation, the American Asthma Foundation, the RWJF Amos Medical Faculty Development Program, Harry Wm. and Diana V. Hind Distinguished Professor in Pharmaceutical Sciences II, the National Heart, Lung, and Blood Institute of the National Institutes of Health (R01HL117004, R01HL128439, R01HL135156, X01HL134589, R01HL141992, and R01HL141845), National Institute of Health and Environmental Health Sciences (R01ES015794 and R21ES24844); the National Institute on Minority Health and Health Disparities (NIMHD) (P60MD006902, R01MD010443, and R56MD013312); the National Institute of General Medical Sciences (NIGMS) (RL5GM118984); the Tobacco-Related Disease Research Program (24RT-0025 and 27IR-0030); and the National Human Genome Research Institute (NHGRI) (U01HG009080) to EGB.

This work was also funded by the Spanish Ministry of Science and Innovation MCIN/AEI/10.13039/501100011033 (PID2020-116274RB-I00) and by Instituto de Salud Carlos III, Spain and the European Regional Development Fund “ERDF A way of making Europe” by the European Union (CB/06/06/1088). MP-Y and EH-L were funded by MCIN/AEI/10.13039/501100011033 and the European Social Fund “ESF Investing in your future” (Ramón y Cajal Program RYC-2015-17205 to MP-Y and PRE2018-083837 to EH-L). AE-O was funded by a fellowship from the Spanish Ministry of Science, Innovation, and Universities (MICIU) and Universidad de La Laguna (ULL), under de M-ULL agreement.

**SALTY**

Recruitment and phenotype definition

The study participants had participated in a telephone interview called Screening Across the Lifespan Twin Study (SALT), conducted between 1998 and 2002. The data collection consisted of three parts: (1) an extensive self-report paper-questionnaire; (2) saliva collection for DNA extraction; and (3) a request to participate in an internet-based investigation. Some of the participants in SALTY were also prior participants of TwinGene – if they had already provided a blood sample they were not also asked to provide saliva. Case - control definition for eczema followed the same approach described for the CATSS study described above.

Genotyping, imputation and statistical analysis is as described above for CATSS.

Acknowledgement and funding is as described above for CATTS.

**SAPALDIA**

Recruitment details

The Swiss Cohort Study on Air Pollution And Lung And Heart Disease In Adults (SAPALDIA cohort) is a population-based multi-center study in eight geographic areas representing the range of environmental, meteorological and socio-demographic conditions in Switzerland. It was initiated in 1991 (SAPALDIA 1) with a follow-up assessment in 2002 (SAPALDIA 2), 2010 (SAPALDIA3), 2017 (SAPALDIA 4) and the fourth follow-up (SAPALDIA 5) currently ongoing. At baseline subjects aged 18 to 60 years from population registries in eight Swiss communities were recruited representing the three largest language groups (German, French, Italian) as well as different levels of air pollution, altitude and degrees of urbanization[85,86]. This cohort study has specifically been designed to investigate longitudinally lung function, respiratory and cardiovascular health; to study and identify the associations of these health indicators with individual long-term exposure to air pollution, other toxic inhalants, life style and molecular factors.

Case/Control definition

SAPALDIA data contributing to the current GWAS are derived from among 6,055 SAPALDIA cohort subjects who participated in both, the baseline (1991) and follow-up (2002) examinations and agreed to providing blood for genetic analysis. At follow-up, 8,047 of 9,651 baseline subjects re-participated in at least one part of the study and a formal biobank was established. At both baseline and follow-up examination subjects underwent spirometry as well as a detailed interview on respiratory health and allergies, smoking history, lifestyle factors and anthropometry. AD was defined as positive answer to the question “Have you ever had atopic dermatitis or any other kind of skin allergy?“ at either examination (SAPALDIA 1 or SAPALDIA 2).

Genotyping and imputation

Genotyping was done using two different genotyping platforms at two different genotyping centers. The first batch of samples was genotyped in the context of the GABRIEL genome-wide association study on asthma using the Illumina 610k quad platform, performed at the genotyping center Centre National de Génotypage (CNG) in Evry, France[87]. Only control subjects of this nested asthma case-control sample were included in the current analysis. The resulting raw genome-wide data were subjected to standard quality control methods. Individuals were excluded on the basis of gender mismatches; minimal or excessive heterozygosity; disproportionate levels of individual missingness (>5%). Population stratification was assessed by examining first two components (C1 and C2) from PCA on highly pruned SNPs using plink’s --mds-plot option; all individuals with non-European ancestry or first degree relatedness were removed. Samples that passed all thresholds were retained during subsequent phasing and imputation. The genotyped SNPs were subjected to the following quality control criteria: SNP call rate >95%, Hardy Weinberg *P*-value > $1x10^{-6}$ and minor allele frequency > 0.01. 545'131 SNPs remained after quality control. These were imputed using minimac release 2012-05-29 and the GIANT ALL reference panel, phase 1 v3.20101123 onto n=30'061'897 variants.

The second batch of samples was genotyped by subcontracting the Wellcome Trust Clinical Research Facility of the University of Edinburgh, United Kingdom, using the Illumina OmniExpressExome platform. As above this batch contains also an underrepresentation of asthmatics compared to the general population. The resulting raw genome-wide data were subjected to standard quality control methods. Samples and SNPs were subjected to the same threshold as above. 673'248 SNPs remained after quality control. These were imputed using Haplotype Reference Consortium panel (HRC r1.1 2016) onto n=39'127'678 variants.

Statistical analysis

Statistical analysis on the first SAPALDIA sample was done using routine 'palogist' in Probabel v. 0.3.0. software for 533 AD cases and 443 controls. For genome-wide analysis, logistic regression of atopic dermatitis on sex, study center and principal components capturing inner European ancestry was performed. The second SAPALDIA sample was statistically analysed using rvtests (version 2.1.0) for 1,589 AD cases and 1,134 controls while controlling for sex, study center and 10 principal components.

Acknowledgements & Funding

The study could not have been done without the help of the study participants, technical and administrative support and the medical teams and field workers at the local study sites.

Study directorate: NM Probst-Hensch (PI; e/g); T Rochat (p), C Schindler (s), N Künzli (e/exp), JM Gaspoz (c) Scientific team: JC Barthélémy (c), W Berger (g), R Bettschart (p), A Bircher (a), C Brombach (n), PO Bridevaux (p), L Burdet (p), Felber Dietrich D (e), M Frey (p), U Frey (pd), MW Gerbase (p), D Gold (e), E de Groot (c), W Karrer (p), F Kronenberg (g), B Martin (pa), A Mehta (e), D Miedinger (o), M Pons (p), F Roche (c), T Rothe (p), P Schmid-Grendelmeyer (a), D Stolz (p), A Schmidt-Trucksäss (pa), J Schwartz (e), A Turk (p), A von Eckardstein (cc), E Zemp Stutz (e). Scientific team at coordinating centers: M Adam (e), I Aguilera (exp), S Brunner (s), D Carballo (c), S Caviezel (pa), I Curjuric (e), A Di Pascale (s), J Dratva (e), R Ducret (s), E Dupuis Lozeron (s), M Eeftens (exp), I Eze (e), E Fischer (g), M Foraster (e), M Germond (s), L Grize (s), S Hansen (e), A Hensel (s), M Imboden (g), A Ineichen (exp), A Jeong (g), D Keidel (s), A Kumar (g), N Maire (s), A Mehta (e), R Meier (exp), E Schaffner (s), T Schikowski (e), M Tsai (exp)

(a) allergology, (c) cardiology, (cc) clinical chemistry, (e) epidemiology, (exp) exposure, (g) genetic and molecular biology, (m) meteorology, (n) nutrition, (o) occupational health, (p) pneumology, (pa) physical activity, (pd) pediatrics, (s) statistic

Local fieldworkers: Aarau: S Brun, G Giger, M Sperisen, M Stahel, Basel: C Bürli, C Dahler, N Oertli, I Harreh, F Karrer, G Novicic, N Wyttenbacher, Davos: A Saner, P Senn, R Winzeler, Geneva: F Bonfils, B Blicharz, C Landolt, J Rochat, Lugano: S Boccia, E Gehrig, MT Mandia, G Solari, B Viscardi, Montana: AP Bieri, C Darioly, M Maire, Payerne: F Ding, P Danieli A Vonnez, Wald: D Bodmer, E Hochstrasser, R Kunz, C Meier, J Rakic, U Schafroth, A Walder. Administrative staff: N Bauer Ott, C Gabriel, R Gutknecht.

The Swiss National Science Foundation (grants no 33CS30-177506/1, 33CS30-148470/1&2, 33CSCO-

134276/1, 33CSCO-108796, 324730_135673, 3247BO-104283, 3247BO-104288, 3247BO-104284, 3247-065896, 3100-059302, 3200-052720, 3200-042532, 4026-028099, PMPDP3_129021/1, PMPDP3_141671/1, 33CS30_177506 and the SNF-SiRENE (grant number CRSII3_147635)), the Federal Office for the Environment, the Federal Office of Public Health, the Federal Office of Roads and Transport, the canton's government of Aargau, Basel-Stadt, Basel-Land, Geneva, Luzern, Ticino, Valais, and Zürich, the Swiss Lung League, the canton's Lung League of Basel Stadt/ Basel Landschaft, Geneva, Ticino, Valais, Graubünden and Zurich, Stiftung ehemals Bündner Heilstätten, SUVA, Freiwillige Akademische Gesellschaft, UBS Wealth Foundation, Talecris Biotherapeutics GmbH, Abbott Diagnostics, Klinik Barmelweid, Hirslanden Klinik Aarau, European Commission 018996 (GABRIEL), Wellcome Trust WT 084703MA, Exposomics EC FP7 grant (Grant agreement No: 308610), ALEC Horizon2020 (Grant agreement No: 633212).

**SAPPHIRE**

Recruitment

The Study of Asthma Phenotypes and Pharmacogenomic Interactions by Race-ethnicity (SAPPHIRE) is an ongoing study that was approved by the Institutional Review Board of Henry Ford Health System. Study individuals included in the current study were members of a large health system, which serves southeast Michigan and all of the Detroit metropolitan statistical area. Individuals were 12-56 years of age and no prior diagnosis of congestive heart failure or chronic obstructive pulmonary disease.

Patient recruitment included patients with and without a clinical diagnosis of asthma. Written informed consent was required at the time of enrolment as a condition for study participation. The exam at the time of enrolment included both a staff-administered questionnaire and lung function testing.

Case/Control definition

For the current analysis we used patient responses to the staff-administered questionnaire to define cases and controls. Patients were considered to be a case if they answered in the affirmative to the question, "Have you ever had eczema or any kind of skin allergy?"

Genotyping and imputation

DNA was isolated from whole blood samples. Genome wide genotyping was performed using the Axiom® AFR array (Affymetrix Inc., Santa Clara, CA. To be included in the analysis, an individual's array results had to have a dish quality control measure 2"0.82, an overall call rate 2"97%, and no discordance between patient-reported sex and the measured X-heterozygosity. We removed SNPs from the analysis which had an overall SNP call rate <95%, Fisher's linear discriminant <3.6, Het strength offset <-0.1, minor allele frequency (MAF) <5%, and Hardy-Weinberg equilibrium p-value < $10^{-5}$. We imputed missing genotypes using the SNPs that passed quality control and data from the March 2012 release of the 1000 Genomes Project. The software program, IMPUTE2[25], performed the imputation.

Statistical Analysis

An additive logistic regression model was used to evaluate the association between individual SNP variant dosage and atopic dermatitis status. Models were adjusted for sex. Variants on the X chromosome were evaluated separately for females and males, since SNP dosage varied from 0-2 in the former and from 0-1 in the latter.

Acknowledgements and Funding

This work was supported by grants from the Fund for Henry Ford Hospital, the American Asthma Foundatin, and the following institutes of the National Institutes of Health: National Institute of Allergy and Infectious Diseases ( AI079139 and AI061774), the National Heart Lung and Blood Institute (HL118267 and HL079055), and the National Institute of Diabetes and Digestive and Kidney diseases (DK064695).

**Southampton Women's Survey (SWS)**

Recruitment

Between 1998 and 2002 the Southampton Women's Survey team interviewed 12,583 Southampton women aged 20 to 34 years. Those who became pregnant after interview were invited to take part in the pregnancy phase of the survey. Enrolment is described in more detail in the cohort profile paper[88]; and via the website http://www.mrc.soton.ac.uk/sws/. There were 3,158 babies born to women in the study between 1998 and 2007. The survey has followed up these children with home visits at six months, one year, 2, 3, 4 and 6-7 years. Biological samples including DNA have been collected for 1940 of the children from this cohort. The study was conducted according to the guidelines in the Declaration of Helsinki, and the Southampton and South West Hampshire Research Ethics Committee approved all procedures (276/97, 307/97, 340/97, 06/Q1702/104). Written informed consent was obtained from all participating women and by parents or guardians with parental responsibility on behalf of their children.

Case/Control definition

The children have been followed up with regular questionnaires and clinic visits. For the current study, data collected from the questionnaires and skin examination was used to classify children as eczema cases or controls. When the children were approximately 6 months, 12 months and 6-7 years, parents were asked about skin conditions in the child. Ascertainment of infantile eczema in the children has previously been reported[89] based on the UK Working Party case definition validated by Williams et al.[90], omitting a history of asthma or hay fever as a criterion given the young age of the infants at 6 and 12 months. Cases at each visit were thus defined as: (A) must have a history of an itchy skin condition plus (B) 3 or more of the following:

1. History of flexural involvement
2. History of asthma/hay fever
3. History of generalised dry skin
4. Onset of rash under the age of 2
5. Visible flexural dermatitis identified by trained research nurses.

Controls did not have eczema at 6 months, 12 months or 6-7 years (allowing one visit to be missing provided that the other 2 visits indicated that the infant/child did not have eczema).

Genotyping

SNP genotyping was carried out using Infinium OmniExpress-24 v1.2Array (713,599 SNPs). Raw IDAT files were loaded into Illumina Genome Studio 2.0.0 and analysed using genotyping Module 2.0.0. Data was processed using Illumina hard cut-off technical specifications. Individuals were additionally excluded on the basis of sex mismatches, relatedness (identity by descent (IBD)) using $\theta$=0.1875 (midpoint between 2nd and 3rd degree relatedness) cut-off, HWE<1x10$^{-6}$, and where possible genotypes were checked against mothers genotype for Mendelian errors. BCFtools and PLINK 1.9 beta were used to prepare the QC genotype data ready for imputation according to SANGER specifications (https://imputation.sanger.ac.uk/?instructions=1#prepareyourdata). Data were uploaded in VCF format to SANGER servers, pre-phased using EAGLE2 pipeline and imputed using Haplotype reference consortium (HRC) r1.1 reference panel[20]. Post-imputation, multi-allelic markers, as well as markers that deviate from HWE (p<1x10$^{-12}$ in the control group) were excluded from the dataset. All individuals with non-European ancestry were removed. The final dataset consisted of 35,7062,88 SNPs for 1773 children (387 cases and 1386 controls).

Statistical Analysis

Statistical analysis performed using rvtests (version v2.1.0, released Feb 2019) for 387 cases and 1386 controls. Logistic regression was performed assuming additive allelic effects while controlling for sex and 10 principal components as covariates.

Acknowledgements and Funding

We thank the mothers of the Southampton Women's Survey who gave us their time and the team of dedicated research nurses and ancillary staff for their assistance. Funding for the components of the Southampton Women's Survey came from the Medical Research Council, British Heart Foundation, Food Standards Agency and Arthritis Research UK. KMG is supported by the UK Medical Research Council (MC_UU_12011/4), the National Institute for Health Research (NIHR Senior Investigator (NF-SI-0515-10042) and NIHR Southampton Biomedical Research Centre (IS-BRC-1215-20004)), the European Union (Erasmus+ Programme ImpENSA 598488-EPP-1-2018-1-DE-EPPKA2-CBHE-JP) and the British Heart Foundation (RG/15/17/3174. SEH is supported through an NIHR Clinical Lectureship.

**TwinGene**

Recruitment and phenotype definition

TwinGene is a longitudinal follow-up study of Swedish twins born during 1911-1958 who have previously participated in SALT (see also description in SALTY) and both twin within each twin pair were still alive during follow-up between 2004 and 2008. Each twin pair was contacted by paper questionnaire asking information regarding their lifestyles and common diseases as well as by health check-up at local healthcare facilities. The study was approved by the Regional Ethical Review Board in Stockholm, Sweden and all participants included in the analysis gave informed consent.

Case-control definition for eczema followed the same approach described for the SALTY study described above.

Genotyping, imputation and statistical analysis

Blood samples and anthropometric measurements were provided at health check-up and genotyping was performed using the Illumina OmniExpress BeadChip. Imputation and statistical analysis is as described above for SALTY and CATSS.

Acknowledgements and funding is as described above for CATSS

**TwinsUK**

Recruitment and phenotype definition

The St Thomas's UK Adult Twin Registry includes 14,000 mainly female twins from throughout the United Kingdom and who are unselected for any diseases or traits. Further details have been described elsewhere[19].

Volunteers twins were asked "Have you ever had eczema?" or "Has a doctor ever told you that you have eczema" on multiple occasions at different time points. Cases were determined from those people who had consistently answered positively in one or more occasions. Only female twins participated to this study. The study was approved by the Local Research Ethics Committee of St Thomas's Hospital, and subjects gave full informed consent.

Genotyping, imputation and statistical analysis

Genotyping of the TwinsUK dataset was done with a combination of Illumina arrays (HumanHap300, HumanHap610Q, 1M-Duo and 1.2MDuo 1M). Further details have been described previously[19]. Haplotype phasing and imputation in the HRC panel version1.1 was performed using the University of Michigan server[20]. Variants with an imputation quality score of $r^2 < 0.5$ or out of Hardy-Weinberg Equilibrium $P < 10^{-12}$ were excluded. A score test implemented in RVTESTS was used to test the association between SNP allelic dosage and eczema status. The first four principle components were included as covariates.

Acknowledgements and funding

TwinsUK was funded by the Wellcome Trust and MRC. The study also receives support from the National Institute for Health Research (NIHR)- funded BioResource, Clinical Research Facility and Biomedical Research Centre based at Guy’s and St Thomas’ NHS Foundation Trust in partnership with King’s College London. We gratefully acknowledge support provided by the JPI HDHL funded DINAMIC consortium (administered by the MRC UK, MR/N030125/1).

**UK Biobank**

Recruitment

UK Biobank is a population-based health research resource consisting of approximately 500,000 people, aged between 38 years and 73 years, who were recruited between the years 2006 and 2010 from across the UK[91]. Particularly focused on identifying determinants of human diseases in middle-aged and older individuals, participants provided a range of information (such as demographics, health status, lifestyle measures, cognitive testing, personality self-report, and physical and mental health measures) via questionnaires and interviews; anthropometric measures, BP readings and samples of blood, urine and saliva were also taken (data available at www.ukbiobank.ac.uk). A full description of the study design, participants and quality control (QC) methods have been described in detail previously[92]. UK Biobank received ethical approval from the Research Ethics Committee (REC reference for UK Biobank is 11/NW/0382; UK Biobank application number 10074).

Case/control definition

AD cases were defined based on their response during a verbal interview with a trained staff member at the assessment centre. Participants were asked to tell the interviewer which serious illnesses or disabilities they had been diagnosed with by a doctor, as were defined as AD if this disease was mention. Disease information was also obtained from the Hospital Episode Statistics (HES) data extract service where health-related outcomes had been defined by International Classification of Diseases (ICD)- 10 code L20. Additionally, individuals were excluded from the AD controls if they had answered "yes" to "Has a doctor ever told you that you have hay fever, allergic rhinitis or eczema".

Genotyping and imputation

The full data release contains the cohort of successfully genotyped samples (n=488,377). 49,979 individuals were genotyped using the UK BiLEVE array and 438,398 using the UK Biobank axiom array. Pre-imputation QC, phasing and imputation are described elsewhere[93]. In brief, prior to phasing, multiallelic SNPs or those with MAF ≤1% were removed. Phasing of genotype data was performed using a modified version of the SHAPEIT2 algorithm[94]. Genotype imputation to a reference set combining the UK10K haplotype and HRC reference panels[95] was performed using IMPUTE2 algorithms[60]. The analyses presented here were restricted to autosomal variants using a graded filtering with varying imputation quality for different allele frequency ranges. Therefore, rarer genetic variants are required to have a higher imputation INFO score (Info>0.3 for MAF >3%; Info>0.6 for MAF 1-3%; Info>0.8 for MAF 0.5-1%; Info>0.9 for MAF 0.1-0.5%) with MAF and Info scores having been recalculated on an in-house derived 'European' subset.

Data quality control

Individuals with sex-mismatch (derived by comparing genetic sex and reported sex) or individuals with sex-chromosome aneuploidy were excluded from the analysis (n=814). We restricted the sample to individuals of white British ancestry who self-report as "White British" and who have very similar ancestral backgrounds according to the PCA (n=409,703), as described by Bycroft[93]. Estimated kinship coefficients using the KING toolset[96] identified 107,162 pairs of related individuals[93]. An inhouse algorithm was then applied to this list and preferentially removed the individuals related to the greatest number of other individuals until no related pairs remain. These individuals were excluded (n=79,448). Additionally 2 individuals were removed due to them relating to a very large number (>200) of individuals.

Statistical analysis

Genome-wide association analysis was performed with rvtests (version 2.1.0) for 10,135 AD cases and 326,899 controls while controlling for sex, genotyping chip and 10 principal components. Following association analysis summary data was available for 5,842,148 variants.

Acknowledgements

This research has been conducted using the UK Biobank Resource under Application Number 10074. Quality Control filtering of the UK Biobank data was conducted within the MRC-IEU by R.Mitchell, G.Hemani, T.Dudding, L.Corbin, S.Harrison, L.Paternoster as described in the published protocol (doi:

10.5523/bris.1ovaau5sxunp2cv8rcy88688v). The MRC IEU UK Biobank GWAS pipeline was developed by B.Elsworth, R.Mitchell, C.Raistrick, L.Paternoster, G.Hemani, T.Gaunt (doi: 10.5523/bris.pnoat8cxo0u52p6ynfaekeigi).

### Replication Studies

#### 23andMe, Inc.

Recruitment details

Individuals included in the analysis were users of 23andMe's personal genomics services that provides genotype and health-related information to customers. They provided informed consent and answered online surveys in accordance with a protocol approved by the external Association for the Accreditation of Human Research Protection Programs-accredited institutional review board, Ethical and Independent Review Services.

Participants provided informed consent and participated in the research online, under a protocol approved by the external AAHRPP-accredited IRB, Ethical & Independent Review Services (E&I Review). Participants were included in the analysis on the basis of consent status as checked at the time data analyses were initiated. The name of the IRB at the time of the approval was Ethical & Independent Review Services. Ethical & Independent Review Services was recently acquired, and its new name as of July 2022 is Salus IRB (https://www.versiticlinicaltrials.org/salusirb)

Case/Control definition

Cases self-identify as having eczema, controls self-identify as not having eczema. Customers responded to the following phrase: "Have you ever been diagnosed with, or treated for, eczema (atopic dermatitis)?"

Genotyping and imputation

Please see details in the following paper[97] under `Genotyping and SNP imputation`: https://www.nature.com/articles/s41588-021-00986-w

Association testing

We performed logistic regression assuming an additive model for allelic effects, using the model:

eczema ~ age + sex + pc.0 + pc.1 + pc.2 + pc.3 + pc.4 + v2_platform + v3_0_platform + v3_1_platform + v4_platform + genotype

Association testing was performed as a likelihood ratio test for each SNP.

This GWAS for the replication analysis includes data from 390,035 cases and 2,531,328 controls filtered to remove close relatives. We removed 16,699 samples (0.6%) based on consent as of 2022-02-25. The results in this report have been adjusted for a LDscore intercept λ=1.353, se=0.025. The LDscore estimated heritability is h^2=0.031, se=0.002.

Acknowledgements

We would like to thank the research participants and employees of 23andMe for making this work possible
The following members of the 23andMe Research Team contributed to this study:
Stella Aslibekyan, Adam Auton, Elizabeth Babalola, Robert K. Bell, Jessica Bielenberg, Katarzyna Bryc, Emily Bullis, Daniella Coker, Gabriel Cuellar Partida, Devika Dhamija, Sayantan Das, Sarah L. Elson, Nicholas Eriksson, Teresa Filshtein, Alison Fitch, Kipper Fletez-Brant, Pierre Fontanillas, Will Freyman, Julie M. Granka, Karl Heilbron, Alejandro Hernandez, Barry Hicks, David A. Hinds, Ethan M. Jewett, Yunxuan Jiang, Katelyn Kukar, Alan Kwong, Keng-Han Lin, Bianca A. Llamas, Maya Lowe, Jey C. McCreight, Matthew H. McIntyre, Steven J. Micheletti, Meghan E. Moreno, Priyanka Nandakumar, Dominique T. Nguyen, Elizabeth S. Noblin, Jared O'Connell, Aaron A. Petrakovitz, G. David Poznik, Alexandra Reynoso, Morgan Schumacher, Anjali J. Shastri, Janie F. Shelton, Jingchunzi Shi, Suyash Shringarpure, Qiaojuan Jane Su, Susana A. Tat, Christophe Toukam Tchakouté, Vinh Tran, Joyce Y. Tung, Xin Wang, Wei Wang, Catherine H. Weldon, Peter Wilton, Corinna D. Wong

Cohort ethical approval

| Cohort Name | Ethics Committee / Institutional Review Board (IRB) | Approved / Waived |
|---|---|---|
| **ALSPAC** | ALSPAC Ethics and Law Committee the Local Research Ethics Committees | Approved |
| **B58C** | NHS Research Ethics Committees | Approved |
| **BAMSE** | the Regional ethical committee in Stockholm | Approved |
| **CAMP** | institutional review boards (IRB) of the Brigham and Women's Hospital, and each of the participating CAMP study centers | Approved |
| **CATSS** | the Regional Ethical Review Board in Stockholm, Sweden | Approved |
| **CHOP** | CHOP institutional Review Board (IRB) | Approved |
| **COPSAC2000** | the Ethics Committee for Copenhagen and The Danish Data Protection Agency | Approved |
| **COPSAC2010** | the Ethics Committee for Copenhagen and The Danish Data Protection Agency | Approved |
| **DANFUND** | the Ethics Committee of Capital Region of Denmark | Approved |
| **DNBC** | the Danish Scientific Ethical Committee and the Danish Data Protection Agency | Approved |
| **ECRHS** | Obtained ethical approval from their local ethics committees and followed the rules for ethics and data protection from their country, which were in accordance with the Declaration of Helsinki | Approved |
| **Estonian Biobank** | Estonian Committee on Bioethics and Human Research. | Approved |
| **FINNGEN** | The Coordinating Ethics Committee of the Helsinki and Uusimaa Hospital District | Approved |
| **GENEVA_AFFY** | the Ethics Committees of the Bavarian Medical Chamber (No. 02014) and the Medical Faculty of the Christian-Albrechts-University Kiel (No. A100/12) | Approved |
| **GENEVA_ILLUMINA** | the Ethics Committees of the Bavarian Medical Chamber (No. 02014) and the Medical Faculty of the Christian-Albrechts-University Kiel (No. A100/12) | Approved |
| **GENUFADext (SHIP1)** | by the institutional review board of Charité - Universitätsmedizin Berlin, Berlin, Germany, and by the ethics committee of the University of Greifswald, Greifswald, Germany | Approved |
| **GENUFAD (SHIP2)** | by the institutional review board of Charité - Universitätsmedizin Berlin, Berlin, Germany, and by the ethics committee of the University of Greifswald, Greifswald, Germany | Approved |
| **GERA** | All study procedures were approved by the Institutional Review Board of the Kaiser Foundation Research Institute | Approved |
| **GINILISAsouth** | the local Ethics Committees | Approved |
| **GINILISAnorth** | the Ethics Committee of the regional Medical Association (Nordrhein), Düsseldorf, Germany | Approved |
| **HEALTH Cohorts** | Ethics Committee of Capital Region of Denmark | Approved |

| | | |
|---|---|---|
| **HUNT** | Data Inspectorate and the Regional Ethics Committee for Medical Research in Norway | Approved |
| **INMA** | Hospital Ethics Committees in each participating region | Approved |
| **IOW** | the local Research Ethics Committee (06/Q1701/34) | Approved |
| **MAAS** | the Local Research Ethics Committee | Approved |
| **MAS_HNR** | the institutional review board of Charité - Universitätsmedizin Berlin, Berlin, Germany | Approved |
| **MoBa_children** | The Regional Committee for Medical Research Ethics (#2015/2425). | Approved |
| **NCRC_ADC** | the Research Ethics Committee of Our Lady's Hospital for Sick Children, Crumlin, Dublin (ref SAC 68/06) | Approved |
| **NFBC66** | The ethics committees in Oulu (Finland) and Oxford (UK) universities | Approved |
| **NTR** | The Central Ethics Committee on Research Involving Human Subjects of the VU University Medical Center, Amsterdam | Approved |
| **PIAMA** | The Medical Ethical Committees of the participating institutes | Approved |
| **The Raine Study** | King Edward Memorial Hospital and Princess Margaret Hospital | Approved |
| **SALTY** | the Regional Ethical Review Board in Stockholm, Sweden | Approved |
| **SAPALDIA** | All procedures in this cohort study were conducted in accordance with the World Medical Association's Declaration of Helsinki and Declaration of Taipei. Written informed consent was obtained from the study participants prior to health examination and blood sample collection. The study protocols of this multi-centric long-term study with baseline and follow-up assessments were approved by Swiss national overarching ethics committees and by regional cantonal ethics committees for each time point of data collection. The SAPALDIA cohort study was approved by the ethics committee of the medical faculty of the University of Lausanne, Switzerland, for the baseline examination (SAPALDIA1) in 1989; and by the Supra-regional Ethics Committee for Clinical Research (UREK approval N° 123/00) of the Swiss Academy of Medical Sciences for the second examination (SAPALDIA2) in September 2001; and given the multi-centric design of the long-term cohort, by multiple cantonal ethics committees for the third examination (SAPALDIA3) in 2009 (ethics committee of the Department of Health and Social Affairs of Aargau, approval N° 2009/056, ethics committee of both Basel, approval N° 219/09, cantonal ethics committee of Zurich, approval N° 52/09, departmental ethics committee for Internal Medicine and Community Medicine of Geneva, approval N° 09-174, cantonal commission for Medical Ethics of Valais, approval N°033/09, cantonal Commission for Ethics in Human Research of Vaud, approval N° 200/09, cantonal Ethics Committee of Ticino, approval N° CE2276). | Approved |

| | | |
|---|---|---|
| **SWS** | The Southampton and South West Hampshire Research Ethics Committee approved all procedures (276/97, 307/97, 340/97, 06/Q1702/104) | Approved |
| **TwinGene** | the Regional Ethical Review Board in Stockholm, Sweden | Approved |
| **TwinsUK** | the Local Research Ethics Committee of St Thomas's Hospital | Approved |
| **UK Biobank** | the Research Ethics Committee REC reference for UK Biobank is 11/NW/0382 | Approved |
| **GALA** | the Human Research Protection Program Institutional Review Board of the University of California, San Francisco UCSF IRB approval No. 10-00889 | Approved |
| **Generation R** | the Medical Ethical Committee of the Erasmus Medical Centre, Rotterdam | Approved |
| **RIKEN** | the research ethics committees at the Institute of Medical Science, the University of Tokyo | Approved |
| **SAGE** | Human Research Protection Program Institutional Review Board of the University of California, San Francisco, UCSF IRB No. 10-02877 | Approved |
| **SAPPHIRE** | the Institutional Review Board of Henry Ford Health System | Approved |
| **23andMe** | the external AAHRPP-accredited IRB, Ethical & Independent Review Services (E&I Review) | Approved |

### References


1. Boyd, A. *et al.* Cohort Profile: The 'Children of the 90s'—the index offspring of the Avon Longitudinal Study of Parents and Children. *Int J Epidemiol* **42**, 111–127 (2013).

2. Fraser, A. *et al.* Cohort Profile: The Avon Longitudinal Study of Parents and Children: ALSPAC mothers cohort. *Int J Epidemiol* **42**, 97–110 (2013).

3. Williams HC, Stratchan DP & Hay RJ. Childhood eczema: disease of the advantaged? *Br Med J* **308**, 1132–1135 (1994).

4. Strachan, D. P. *et al.* Lifecourse influences on health among British adults: Effects of region of residence in childhood and adulthood. *Int J Epidemiol* **36**, 522–531 (2007).

5. Burton, P. R. *et al.* Genome-wide association study of 14,000 cases of seven common diseases and 3,000 shared controls. *Nature 2007 447:7145* **447**, 661–678 (2007).

6. Barrett, J. C. *et al.* Genome-wide association study and meta-analysis find that over 40 loci affect risk of type 1 diabetes. *Nature Genetics 2009 41:6* **41**, 703–707 (2009).

7. Moffatt, M. F. *et al.* A Large-Scale, Consortium-Based Genomewide Association Study of Asthma. *New England Journal of Medicine* **363**, 1211–1221 (2010).

8. Nagai, A. *et al.* Overview of the BioBank Japan Project: Study design and profile. *J Epidemiol* **27**, S2–S8 (2017).

9. Hirata, M. *et al.* Cross-sectional analysis of BioBank Japan clinical data: A large cohort of 200,000 patients with 47 common diseases. *J Epidemiol* **27**, S9–S21 (2017).

10. Hanifin MJ & Rajka G. Diagnostic features of atopic dermatitis. *Acta Dermatovener (Stockholm)* **92**, 44–47 (1980).

11. Tanaka, N. *et al.* Eight novel susceptibility loci and putative causal variants in atopic dermatitis. *Journal of Allergy and Clinical Immunology* **148**, 1293–1306 (2021).

12. The Childhood Asthma Management Program (CAMP): design, rationale, and methods. Childhood Asthma Management Program Research Group. *Control Clin Trials* **20**, 91–120 (1999).

13. Wu, K. *et al.* Genome-wide interrogation of longitudinal FEV1 in children with asthma. *Am J Respir Crit Care Med* **190**, 619–27 (2014).

14. Li, Y., Willer, C. J., Ding, J., Scheet, P. & Abecasis, G. R. MaCH: using sequence and genotype data to estimate haplotypes and unobserved genotypes. *Genet Epidemiol* **34**, 816–34 (2010).

15. Purcell, S. *et al.* PLINK: a tool set for whole-genome association and population-based linkage analyses. *Am J Hum Genet* **81**, 559–75 (2007).

16. Anckarsäter, H. *et al.* The Child and Adolescent Twin Study in Sweden (CATSS). *Twin Res Hum Genet* **14**, 495–508 (2011).

17. Lichtenstein, P. *et al.* The Swedish Twin Registry: a unique resource for clinical, epidemiological and genetic studies. *J Intern Med* **252**, 184–205 (2002).

18. Lichtenstein, P. *et al.* The Swedish Twin Registry in the third millennium: an update. *Twin Res Hum Genet* **9**, 875–82 (2006).

19. Grosche, S. *et al.* Rare variant analysis in eczema identifies exonic variants in DUSP1, NOTCH4 and SLC9A4. *Nature Communications 2021 12:1* **12**, 1–11 (2021).

20. McCarthy, S. *et al.* A reference panel of 64,976 haplotypes for genotype imputation. *Nat Genet* **48**, 1279–1283 (2016).

21. Olsen, J. *et al.* The Danish National Birth Cohort--its background, structure and aim. *Scand J Public Health* **29**, 300–7 (2001).

22. Benn, C. S. *et al.* Atopic dermatitis in young children: diagnostic criteria for use in epidemiological studies based on telephone interviews. *Acta Derm Venereol* **83**, 347–50 (2003).

23. The 1000 Genomes Project Consortium. A map of human genome variation from population-scale sequencing. *Nature* **467**, 1061–1073 (2010).

24. Delaneau, O., Marchini, J. & Zagury, J. F. A linear complexity phasing method for thousands of genomes. *Nature Methods 2011 9:2* **9**, 179–181 (2011).

25. Howie, B. N., Donnelly, P. & Marchini, J. A flexible and accurate genotype imputation method for the next generation of genome-wide association studies. *PLoS Genet* **5**, e1000529 (2009).

26. Marchini, J. & Howie, B. Genotype imputation for genome-wide association studies. *Nature Reviews Genetics 2010 11:7* **11**, 499–511 (2010).

27. Burney, P., Luczynska, C., Chinn, S. & Jarvis, D. The European Community Respiratory Health Survey. *European Respiratory Journal* **7**, (1994).

28. Committee, T. E. C. R. H. S. I. S. The European Community Respiratory Health Survey II. *European Respiratory Journal* **20**, 1071–1079 (2002).

29. Howie, B. N., Donnelly, P. & Marchini, J. A Flexible and Accurate Genotype Imputation Method for the Next Generation of Genome-Wide Association Studies. *PLoS Genet* **5**, 1000529 (2009).

30. Leitsalu, L. *et al.* Cohort Profile: Estonian Biobank of the Estonian Genome Center, University of Tartu. *Int J Epidemiol* **44**, 1137–1147 (2015).

31. Laisk, T. *et al.* Genome-wide association study identifies five risk loci for pernicious anemia. *Nature Communications 2021 12:1* **12**, 1–9 (2021).

32. Pärn, K. *et al.* Genotyping chip data lift-over to reference genome build GRCh38/hg38 V.1. *protocols.io* https://www.protocols.io/view/genotyping-chip-data-lift-over-to-reference-genome-n2bvjmbpvk5w/v1 (2018) doi:https://dx.doi.org/10.17504/protocols.io.nqtddwn.

33. Pärn, K. *et al.* Genotype imputation workflow v3.0 V.1. *protocols.io* (2018).

34. Borrell, L. N. *et al.* Childhood Obesity and Asthma Control in the GALA II and SAGE II Studies. *https://doi.org/10.1164/rccm.201211-2116OC* **187**, 697–702 (2013).

35. Nishimura, K. K. *et al.* Early-Life air pollution and asthma risk in minority children the GALA II and SAGE II studies. *Am J Respir Crit Care Med* **188**, 309–318 (2013).

36. Thakur, N. *et al.* Socioeconomic status and childhood asthma in urban minority youths: The GALA II and SAGE II studies. *Am J Respir Crit Care Med* **188**, 1202–1209 (2013).

37. Herrera-Luis, E. *et al.* Genome-wide association study reveals a novel locus for asthma with severe exacerbations in diverse populations. *Pediatric Allergy and Immunology* **32**, 106–115 (2021).

38. McCarthy, S. *et al.* A reference panel of 64,976 haplotypes for genotype imputation. *Nature Genetics 2016 48:10* **48**, 1279–1283 (2016).

39. Delaneau, O., Coulonges, C. & Zagury, J. F. Shape-IT: New rapid and accurate algorithm for haplotype inference. *BMC Bioinformatics* **9**, 1–14 (2008).

40. Das, S. *et al.* Next-generation genotype imputation service and methods. *Nature Genetics 2016 48:10* **48**, 1284–1287 (2016).

41. Zhan, X., Hu, Y., Li, B., Abecasis, G. R. & Liu, D. J. RVTESTS: an efficient and comprehensive tool for rare variant association analysis using sequence data. *Bioinformatics* **32**, 1423–1426 (2016).

42. Jaddoe, V. W. v *et al.* The Generation R Study: design and cohort update 2012. *Eur J Epidemiol* **27**, 739–56 (2012).

43. Kruithof, C. J. *et al.* The Generation R Study: Biobank update 2015. *Eur J Epidemiol* **29**, 911–27 (2014).

44. Howie, B., Fuchsberger, C., Stephens, M., Marchini, J. & Abecasis, G. R. Fast and accurate genotype imputation in genome-wide association studies through pre-phasing. *Nat Genet* **44**, 955–9 (2012).

45. Li, Y., Willer, C., Sanna, S. & Abecasis, G. Genotype imputation. *Annu Rev Genomics Hum Genet* **10**, 387–406 (2009).

46. Estrada, K. *et al.* GRIMP: a web- and grid-based tool for high-speed analysis of large-scale genome-wide association using imputed data. *Bioinformatics* **25**, 2750–2 (2009).

47. Krawczak, M. *et al.* PopGen: population-based recruitment of patients and controls for the analysis of complex genotype-phenotype relationships. *Community Genet* **9**, 55–61 (2006).

48. Wichmann, H.-E., Gieger, C., Illig, T. & MONICA/KORA Study Group. KORA-gen--resource for population genetics, controls and a broad spectrum of disease phenotypes. *Gesundheitswesen* **67 Suppl 1**, S26-30 (2005).

49. Holle, R., Happich, M., Löwel, H., Wichmann, H. E. & MONICA/KORA Study Group. KORA--a research platform for population based health research. *Gesundheitswesen* **67 Suppl 1**, S19-25 (2005).

50. Esparza-Gordillo, J. *et al.* A common variant on chromosome 11q13 is associated with atopic dermatitis. *Nat Genet* **41**, 596–601 (2009).

51. Marenholz, I. *et al.* The eczema risk variant on chromosome 11q13 (rs7927894) in the population-based ALSPAC cohort: a novel susceptibility factor for asthma and hay fever. *Hum Mol Genet* **20**, 2443–9 (2011).

52. Völzke, H. *et al.* Cohort profile: the study of health in Pomerania. *Int J Epidemiol* **40**, 294–307 (2011).

53. Price, A. L. *et al.* Principal components analysis corrects for stratification in genome-wide association studies. (2006) doi:10.1038/ng1847.

54. Abecasis, G. R., Cherny, S. S., Cookson, W. O. & Cardon, L. R. Merlin--rapid analysis of dense genetic maps using sparse gene flow trees. *Nat Genet* **30**, 97–101 (2002).

55. Hoffmann, T. J. *et al.* Design and coverage of high throughput genotyping arrays optimized for individuals of East Asian, African American, and Latino race/ethnicity using imputation and a novel hybrid SNP selection algorithm. *Genomics* **98**, 422–30 (2011).

56. Hoffmann, T. J. *et al.* Next generation genome-wide association tool: design and coverage of a high-throughput European-optimized SNP array. *Genomics* **98**, 79–89 (2011).

57. Kvale, M. N. *et al.* Genotyping Informatics and Quality Control for 100,000 Subjects in the Genetic Epidemiology Research on Adult Health and Aging (GERA) Cohort. *Genetics* **200**, 1051–60 (2015).

58. Banda, Y. *et al.* Characterizing Race/Ethnicity and Genetic Ancestry for 100,000 Subjects in the Genetic Epidemiology Research on Adult Health and Aging (GERA) Cohort. *Genetics* **200**, 1285–95 (2015).

59. Delaneau, O., Marchini, J. & Zagury, J.-F. A linear complexity phasing method for thousands of genomes. *Nat Methods* **9**, 179–81 (2011).

60. Howie, B., Marchini, J. & Stephens, M. Genotype Imputation with Thousands of Genomes. (2011) doi:10.1534/g3.111.001198.

61. Reed, E. *et al.* A guide to genome-wide association analysis and post-analytic interrogation. *Stat Med* **34**, 3769 (2015).

62. Das, S. *et al.* Next-generation genotype imputation service and methods. *Nature Genetics 2016 48:10* **48**, 1284–1287 (2016).

63. Heinrich, J. *et al.* Allergens and endotoxin on mothers' mattresses and total immunoglobulin E in cord blood of neonates. *European Respiratory Journal* **20**, 617–623 (2002).

64. Berg, A. v. *et al.* Impact of early feeding on childhood eczema: development after nutritional intervention compared with the natural course – the GINIplus study up to the age of 6 years. *Clinical & Experimental Allergy* **40**, 627–636 (2010).

65. McCarthy, S. *et al.* A reference panel of 64,976 haplotypes for genotype imputation. *Nature Genetics 2016 48:10* **48**, 1279–1283 (2016).

66. Krokstad, S. *et al.* Cohort profile: The HUNT study, Norway. *Int J Epidemiol* **42**, 968–977 (2013).

67. Zhou, W. *et al.* Efficiently controlling for case-control imbalance and sample relatedness in large-scale genetic association studies. *Nature Genetics 2018 50:9* **50**, 1335–1341 (2018).

68. Arshad, S. H. *et al.* Cohort Profile: The Isle Of Wight Whole Population Birth Cohort (IOWBC). *Int J Epidemiol* **47**, 1043–1044i (2018).

69. Arshad, S. H. *et al.* Polymorphisms in the interleukin 13 and GATA binding protein 3 genes and the development of eczema during childhood. *British Journal of Dermatology* **158**, 1315–1322 (2008).

70. Quraishi, B. M. *et al.* Identifying CpG sites associated with eczema via random forest screening of epigenome-scale DNA methylation. *Clin Epigenetics* **7**, 1–11 (2015).

71. Lau, S. *et al.* Early exposure to house-dust mite and cat allergens and development of childhood asthma: a cohort study. Multicentre Allergy Study Group. *Lancet* **356**, 1392–7 (2000).

72. Nickel, R. *et al.* Messages from the German Multicentre Allergy Study. *Pediatr Allergy Immunol* **13**, 7–10.

73. Schmermund, A. *et al.* Assessment of clinically silent atherosclerotic disease and established and novel risk factors for predicting myocardial infarction and cardiac death in healthy middle-aged subjects: Rationale and design of the Heinz Nixdorf RECALL Study. *Am Heart J* **144**, 212–218 (2002).

74. Magnus, P. *et al.* Cohort Profile Update: The Norwegian Mother and Child Cohort Study (MoBa). *Int J Epidemiol* **45**, 382–388 (2016).

75. Delaneau, O., Zagury, J. F. & Marchini, J. Improved whole-chromosome phasing for disease and population genetic studies. *Nature Methods 2013 10:1* **10**, 5–6 (2013).

76. Hoffmann, T. J. *et al.* Design and coverage of high throughput genotyping arrays optimized for individuals of East Asian, African American, and Latino race/ethnicity using imputation and a novel hybrid SNP selection algorithm. *Genomics* **98**, 422–430 (2011).

77. Rantakallio, P. The longitudinal study of the northern Finland birth cohort of 1966. *Paediatr Perinat Epidemiol* **2**, 59–88 (1988).

78. van Beijsterveldt, C. E. M. & Boomsma, D. I. Genetics of parentally reported asthma, eczema and rhinitis in 5-yr-old twins. *Eur Respir J* **29**, 516–21 (2007).

79. van Beijsterveldt, C. E. M. *et al.* The Young Netherlands Twin Register (YNTR): longitudinal twin and family studies in over 70,000 children. *Twin Res Hum Genet* **16**, 252–67 (2013).

80. Willemsen, G. *et al.* The Adult Netherlands Twin Register: twenty-five years of survey and biological data collection. *Twin Res Hum Genet* **16**, 271–81 (2013).

81. Meulenbelt, I., Droog, S., Trommelen, G. J., Boomsma, D. I. & Slagboom, P. E. High-yield noninvasive human genomic DNA isolation method for genetic studies in geographically dispersed families and populations. *Am J Hum Genet* **57**, 1252–4 (1995).

82. Scheet, P. *et al.* Twins, tissue, and time: an assessment of SNPs and CNVs. *Twin Res Hum Genet* **15**, 737–45 (2012).

83. Wijga, A. H. *et al.* Cohort profile: The Prevention and Incidence of Asthma and Mite Allergy (PIAMA) birth cohort. *Int J Epidemiol* **43**, 527–535 (2014).

84. Newnham, J. P., Evans, S. F., Michael, C. A., Stanley, F. J. & Landau, L. I. Effects of frequent ultrasound during pregnancy: a randomised controlled trial. *Lancet* **342**, 887–91 (1993).

85. Martin, B. W. *et al.* SAPALDIA: methods and participation in the cross-sectional part of the Swiss Study on Air Pollution and Lung Diseases in Adults. *Soz Praventivmed* **42**, 67–84 (1997).

86. Ackermann-Liebrich, U. *et al.* Follow-up of the Swiss Cohort Study on Air Pollution and Lung Diseases in Adults (SAPALDIA 2) 1991-2003: methods and characterization of participants. *Soz Praventivmed* **50**, 245–63 (2005).

87. Moffatt, M. F. *et al.* A large-scale, consortium-based genomewide association study of asthma. *N Engl J Med* **363**, 1211–1221 (2010).

88. Inskip, H. M. *et al.* Cohort profile: The Southampton Women's Survey. *Int J Epidemiol* **35**, 42–8 (2006).

89. El-Heis, S. *et al.* Higher maternal serum concentrations of nicotinamide and related metabolites in late pregnancy are associated with a lower risk of offspring atopic eczema at age 12 months. *Clinical & Experimental Allergy* **46**, 1337–1343 (2016).

90. Williams, H. C., Burney, P. G. J., Pembroke, A. C. & Hay, R. J. The U.K. Working Party's Diagnostic Criteria for Atopic Dermatitis. III. Independent hospital validation. *British Journal of Dermatology* **131**, 406–416 (1994).

91. Allen, N. E., Sudlow, C., Peakman, T. & Collins, R. UK Biobank Data: Come and Get It. *Sci Transl Med* **6**, (2014).

92. Collins, R. What makes UK Biobank special? *The Lancet* **379**, 1173–1174 (2012).

93. Bycroft, C. *et al.* The UK Biobank resource with deep phenotyping and genomic data. *Nature* (2018) doi:10.1038/s41586-018-0579-z.

94. Sharp, K. *et al.* Haplotype estimation for biobank-scale data sets. *Nature Publishing Group* **48**, (2016).

95. Huang, J. *et al.* Improved imputation of low-frequency and rare variants using the UK10K haplotype reference panel. *Nat Commun* (2015) doi:10.1038/ncomms9111.

96. Manichaikul, A. *et al.* Robust relationship inference in genome-wide association studies. **26**, 2867–2873 (2010).

97. Shelton, J. F. *et al.* The UGT2A1/UGT2A2 locus is associated with COVID-19-related loss of smell or taste. *Nat Genet* **54**, 121–124 (2022).
